# Immunogenicity and safety of a 4^th^ homologous booster dose of a SARS-CoV-2 recombinant spike protein vaccine (NVX-CoV2373): a phase 2, randomized, placebo-controlled trial

**DOI:** 10.1101/2022.11.18.22282414

**Authors:** Katia Alves, Joyce S Plested, Shirley Galbiati, Gordon Chau, Shane Cloney-Clark, Mingzhu Zhu, Raj Kalkeri, Nita Patel, Kathy Smith, Alex Marcheschi, Susan Pfeiffer, Heather McFall, Gale Smith, Gregory M. Glenn, Filip Dubovsky, Raburn M. Mallory, the Novavax 2019nCoV-101 Study Group

## Abstract

**Background:** The emergence of SARS-CoV-2 variants has significantly reduced the efficacy of some approved vaccines. A fourth dose of NVX-CoV2373 (5µg SARS-CoV-2 rS + 50µg Matrix-M™ adjuvant) was evaluated to determine induction of cross-reactive antibodies to variants of concern.

**Methods:** A phase 2 randomized study assessed a fourth dose of NVX-CoV2373 in adults 18–84 years of age (2-dose primary series followed by third and fourth doses at 6-month intervals). Local/systemic reactogenicity was assessed the day of vaccination and for 6 days thereafter. Unsolicited adverse events (AEs) were reported. Immunogenicity was measured before, and 14 days after, fourth dose administration using anti-spike neutralization assays against the ancestral SARS-CoV-2 strain and Omicron sublineages. Antigenic cartography assessed antigenic distances between ancestral and variant strains.

**Results:** Among 1283 enrolled participants, 258 were randomized to receive the 2-dose primary series, of whom 104 received a third dose, and 45 received a fourth dose of NVX-CoV2373. The incidence of local/systemic reactogenicity events increased after the first three doses of NVX-CoV2373, and leveled off after dose four. Unsolicited AEs were reported in 9% of participants after dose 4 (none severe or serious). Neutralization antibody titers increased following booster doses. Antigenic cartography demonstrated reductions in antigenic distance between ancestral and variant SARS-CoV-2 strains with increased number of NVX-CoV2373 doses.

**Conclusions:** A fourth dose of NVX-CoV2373 enhanced immunogenicity without increasing reactogenicity. Antigenic cartography demonstrated a more universal-like response against SARS-CoV-2 variants after a fourth dose of NVX-CoV2373, indicating that updates to the vaccine composition may not be warranted.

**Trial registration number:** NCT04368988

## Introduction

The emergence and rapid propagation of SARS-CoV-2 variants, in particular the Omicron sublineages, which have mutations that increase viral transmissibility and enhance the viruses’ ability to evade vaccine immunity,^1,2^ can significantly reduce the efficacy of approved vaccines. The FDA has recommended the development of variant-specific vaccines for the 2022-2023 Winter^3^, although some have countered that any additional protection provided by Omicron-specific vaccines may be minimal.^4,5^

The degree to which immunity induced by the SARS-CoV-2 ancestral strain-based vaccines is effective against the Omicron sublineages depends, to a large degree, on the extent that the vaccine is able to induce broadly cross-reactive antibodies. The NVX-CoV2373 vaccine may be able to preferentially induce these antibodies due to its composition. The vaccine consists of full-length, pre-fusion recombinant S protein trimers with epitopes conserved across variants. In addition, the vaccine is co-formulated with a saponin-based adjuvant, Matrix-M™. Similar saponin-based adjuvants have demonstrated the ability to enhance antibody avidity, affinity maturation and epitope spreading, which may drive recognition of conserved, more immunologically cryptic but broadly neutralizing epitopes.^6^ For example, the ISCOMATRIX™ adjuvant promotes epitope spreading and antibody affinity maturation of a pandemic influenza A H7N9 virus-like particle vaccine which correlates with virus neutralization in humans.^6^ In the context of seasonal influenza, a hemagglutinin (HA) nanoparticle Matrix-M adjuvanted vaccine was shown to induce polyclonal antibodies in humans that mimic broadly neutralizing monoclonal antibodies which recognize two conserved regions of the head domain, namely the receptor binding site and the vestigial esterase subdomain. The findings raised the potential for an adjuvanted HA subunit vaccine to induce broadly protective immunity.^7^ Clinical and ferret studies using a recombinant full-length HA in a nanoparticle, adjuvanted with Matrix-M, confirmed that the H3N2 component of the vaccine-induced broadly cross-reacting antibodies against the past, current, and forward drifted H3N2 strains covering multiple years.^8,9^ Humans are universally primed to influenza, and as this is gradually also becoming the case for SARS-CoV-2, we speculated that repetitive boosting with an ancestral rS nanoparticle vaccine, adjuvanted with Matrix-M, might similarly induce broadly neutralizing antibodies that recognize the drift variants.

In two phase 3, randomized, placebo-controlled clinical trials of healthy adult participants who received two doses of NVX-CoV2373, vaccine efficacy of 89·7% (95% CI 80·2–94·6) and 90·4% (95% CI 82·9– 94·6) was established in the UK (N=15 139) and the US and Mexico (N=29 582), respectively.^10,11^ Recently, however, as waning immunity both over time and with the emergence of SARS-CoV-2 variants has been noted with authorized and approved COVID-19 vaccines, the safety and immunogenicity of booster doses of NVX-CoV2373 were evaluated.

As part of an ongoing phase 2, randomized, placebo-controlled clinical trial of NVX-CoV2373 conducted in Australia and the US, two doses of NVX-CoV2373 were administered 21 days apart,^12^ followed initially by a single booster dose after approximately 6 months.^13^ Administration of a single booster dose resulted in an incremental increase in the frequency of reactogenicity events along with significantly enhanced immunogenicity, including an approximate 33.7-fold increase from pre-booster levels in anti-spike (S) serum immunoglobulin G (IgG) against the ancestral SARS-CoV-2 strain. In a comparison of SARS-CoV-2 variants, markedly higher anti-rS IgG activity, neutralization titers, and hACE2 receptor inhibition were noted against several variants, including Omicron BA.1 and BA.2 sublineages.^13^ In a continuation of this study, a second booster dose of NVX-CoV2373 was administered after another 6 months. The safety and immunogenicity after the fourth dose of NVX-CoV2373 are reported herein. Antigenic cartography^14–17^ was used to measure the distances between immune responses to the Wuhan (ancestral) strain and predominant circulating strains to help determine whether a vaccine update might be warranted.

## Methods

### Study design

As part of an ongoing phase 2, randomized, observer-blinded, placebo-controlled study of NVX-CoV2373 (methods previously reported^13^), a subset of healthy adult participants received a 2-dose primary series followed by third and fourth doses of NVX-CoV2373 administered at 6-month intervals. All active booster vaccinations were administered at the same dose level as the primary vaccination series (0.5 mL containing 5 µg SARS-CoV-2 rS with 50 µg Matrix-M adjuvant) via intramuscular injection. Only participants who received four doses of NVX-CoV2373 are included in this analysis.

Participants utilized an electronic diary to record solicited local and systemic reactogenicity on the day of vaccination and for an additional 6 days thereafter. Unsolicited AEs were reported through 28 days after vaccination. SAEs, MAAEs attributed to vaccine, and AESIs were reported through the end of the study.^18^ Blood samples for immunogenicity analysis were collected prior to each vaccination and after each vaccination on Days 0, 35, 189, 217, 357, and 371.

Immune response was assessed as previously described before and after each dose of NVX-CoV2373 via serum neutralizing antibodies using validated pseudovirus neutralization assays^19^ for ancestral, BA.1, and BA.4/BA.5 variants; via a validated anti-rS IgG assay,^20^ and via a fit-for-purpose SARS-CoV-2 hACE2 receptor binding inhibition assay.^13^ Antigenic cartography maps were constructed using Cartography software available at acmacs-web.antigenic-cartography.org. Serum neutralizing titers collected after 2 doses, three doses, and four doses were input into the software to construct a SARS-CoV-2 antigenic map. SARS-CoV-2 antigens are depicted as circles and sera are indicated as squares. Each grid square represents one antigenic unit representing a two-fold change in titers. The antigenic distances from the ancestral vaccine strain to Omicron variants were calculated after two, three, and four doses of vaccine, followed by conversion of antigenic distances to fold-differences, as previously described.^17^

Only participants who were SARS-CoV-2 negative prior to NVX-CoV2373 administration, who received all vaccine doses per-protocol, who did not have a positive anti-nucleocapsid protein result on or after Day 371, and had no significant protocol deviations that impacted immunogenicity response at the corresponding study visit (e.g., receipt of vaccine for COVID-19 or other prohibited medication, receipt of incorrect treatment) were included in the analysis. For this analysis, unblinding was not considered a protocol deviation that impacts immune response.

The trial protocol was approved by the Alfred Hospital Ethics Committee (Melbourne, Victoria, Australia) and Advarra Central Institutional Review Board (Colombia, Maryland, USA) and is registered on Clinicaltrials.gov (NCT04368988). This study was performed in accordance with the International Conference on Harmonization, Good Clinical Practice guidelines. Safety oversight for the study was provided by an independent Safety Monitoring Committee.

## Results

### Participants

Of 1610 participants screened from 24 August 2020 to 25 September 2020, 1288 were randomized and 1283 received at least 1 dose of study vaccine. Of the 258 participants who were randomized to receive 2-doses of NVX-CoV2373 (5 μg SARS-CoV-2 rS + 50 μg Matrix-M), 207 received both doses and were then re-randomized to receive a dose of placebo (n=102) or a third dose of NVX-CoV2373 (n=105) after 6 months. Of the participants who received a third dose of NVX-CoV2373 and chose to continue in the study, 45 participants received a fourth dose of NVX-CoV2373 after another 6 months from the third dose (**Figure 1**). A subset of participants who received the fourth dose and who met the immunogenicity analysis criteria (n=34) were included in the cartography analysis.

**Figure 1.**
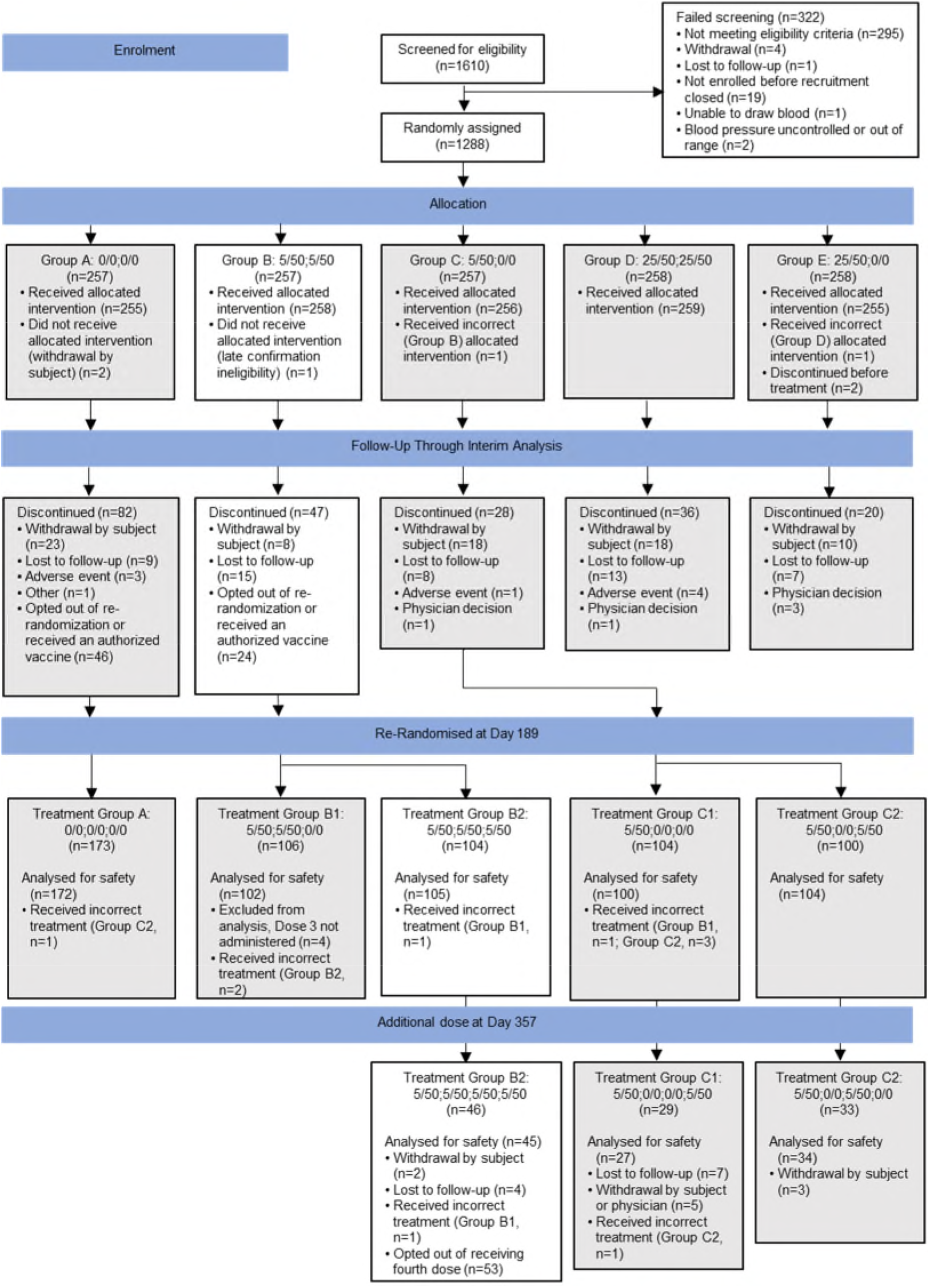
CONSORT diagram. White boxes demonstrate the flow of participants included in this analysis, including those that received 1, 2, 3, or 4 doses of 5 µg SARS-CoV-2 rS + 50 µg Matrix-M™ adjuvant (5/50) on Day 0, Day 21, Day 189, and Day 357, respectively. Gray boxes include participant groups not covered in this analysis. 0/0: 0 µg SARS-CoV-2 rS + 0 µg Matrix-M adjuvant; 5/50: 5 µg SARS-CoV-2 rS + 50 µg Matrix-M adjuvant; 25/50: 25 µg SARS-CoV-2 rS + 50 µg Matrix-M adjuvant.

The demographic characteristics of participants were similar when compared by number of doses received (**Table 1**). For the 4^th^ dose, the mean age of 55 years was slightly higher than the mean of 51 years after the primary series and after dose 3. This is reflected in the age group distribution, where 56% were ≥60 to ≤84 years of age compared with 46% for the primary series and dose 3. Most participants were White (89%) with a negative baseline SARS-CoV-2 serostatus (98%).

**Table 1:** Demographics characteristics for participants who received up to 4 doses of NVX-CoV2373 (safety analysis set^a^)

|  | n (%) |  |  |
| --- | --- | --- | --- |
| Parameter | Primary series<br>(Doses 1 &2)<br>N=258 | Dose 3<br>N=105 | Dose 4<br>N=45 |
| Age – years, mean (SD) | 51.3 (17.5) | 51.7 (17.1) | 55.3 (15.4) |
| 18–59, n (%) | 140 (54) | 57 (54) | 20 (44) |
| 60–84, n (%) | 118 (46) | 48 (46) | 25 (56) |
| Sex, n (%) |  |  |  |
| Male | 119 (46) | 58 (55) | 18(40) |
| Female | 139 (54) | 47 (45) | 27 (60) |
| Race or ethnic group, n (%) |  |  |  |
| White | 226 (87) | 93 (89) | 40 (89) |
| Black or African American | 7 (3) | 3 (3) | 2 (4) |
| Asian | 18 (7) | 7 (7) | 3 (7) |
| American Indian or Alaska Native | 2 (1) | 1 (1) | 0 |
| Multiracial | 3 (1) | 1 (1) | 0 |
| Not reported/missing | 3 (1) | 0 | 0 |
| Hispanic or Latino | 6 (2) | 1 (1) | 0 |
| Body mass index, kg/m<sup>2</sup>, mean (SD) | 27.10 (4.12) | 27.43 (4.04) | 28.37 (4.47) |
| Baseline SARS-CoV-2 seropositive, n (%) | 5 (2) | 3 (3) | 1 (2) |
| Country of participants, n (%) |  |  |  |
| Australia | 134 (52) | 57 (54) | 23 (51) |
| United States | 124 (48) | 48 (46) | 22 (49) |

<sup>a</sup> Participants in the safety analysis set are counted according to the treatment received to accommodate for treatment errors.

### Safety

Reporting of solicited local and systemic reactogenicity events of any grade generally increased after each of the first three doses of NVX-CoV2373 and leveled off or decreased after the fourth dose (**Figure 2**). Grade 3 or higher (Grade 3+) reactogenicity events generally followed a similar pattern. Following the fourth dose, 73% (n=30) of participants reported any solicited local reactions (tenderness, pain, swelling, erythema) of any Grade and 19% (n=8) for Grade 3+ compared with 83% (n=80) for any grade and 13% (n=13) for Grade 3+, following the third dose. Only erythema was reported more frequently after the fourth dose, with 20% (n=8) for any Grade and 15% (n=6) for Grade 3+ compared with 10% (n=10) any grade and 1% (n=1) Grade 3+ after the third dose. Following the fourth dose, local reactions were short-lived (median duration: pain, 2 days; tenderness and erythema, 3 days; swelling, 4 days).

**Figure 2.**
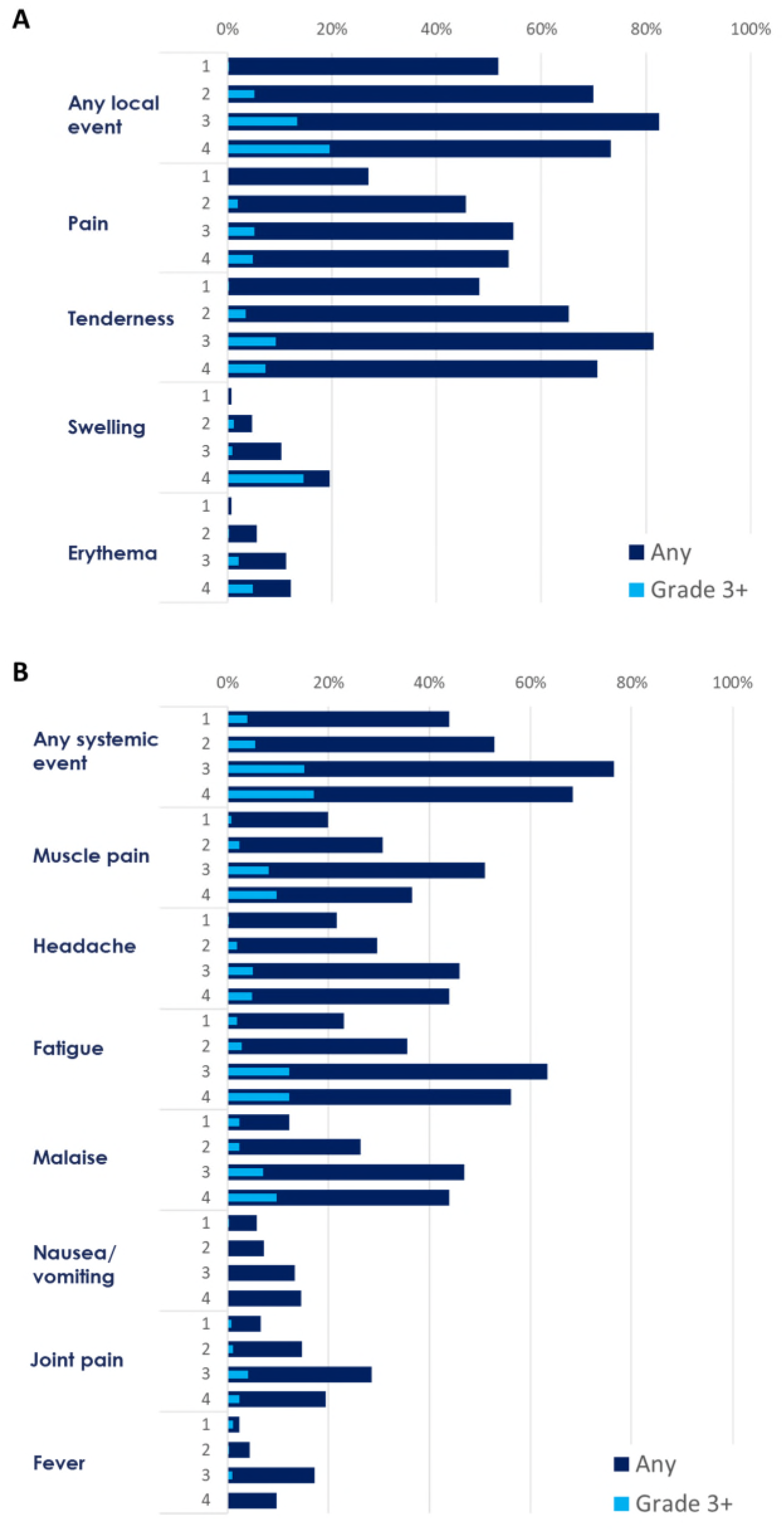
Solicited (A) local and (B) systemic reactogenicity events for participants who received NVX-CoV2373, by dose number and severity.

Solicited systemic reactions showed a similar pattern with reporting rates for any event (fatigue, headache, muscle pain, malaise, joint pain, nausea/vomiting, and fever) of 68% (n=28) for any grade (17% [n=7] Grade 3+) after the fourth dose compared with 77% (n=75) for any grade (15% [n=15] Grade 3+) after the third dose. Notably, the incidence of fever remained low at 10% with no Grade 3+ events reported. Following the fourth dose, solicited systemic reactions were also transient in nature (median duration: fever and headache, 1 day; fatigue, malaise, joint pain, nausea/vomiting, and muscle pain, 2 days).

After the fourth dose, unsolicited TEAEs occurred in 4/45 participants (9%), none of which were severe or serious (**Table 2**). The most common unsolicited TEAEs were injection site pain, injection site erythema, injection site pruritus, and pruritus, all of which occurred in 1 participant each. There were no serious adverse events, MAAEs, or PIMMCs after the fourth dose.

**Table 2:** Overall summary of unsolicited treatment-emergent adverse events for participants who received up to 4 doses of NVX-CoV2373 (safety analysis set^a^)

|  | n (%) |  |  |  |
| --- | --- | --- | --- | --- |
| Parameter | Dose 1<br>N=258 | Dose 2<br>N=255 | Dose 3<br>N=105 | Dose 4<br>N=45 |
| Any unsolicited TEAE | 31 (12) | 76 (30) | 14 (13) | 4 (9) |
| Treatment-related | 0 | 6 (2) | 4 (4) | 4 (9) |
| Severe | 2 (1) | 8 (3) | 2 (2) | 0 |
| Leading to vaccination discontinuation | 0 | 1 (<1) | 0 | 0 |
| Leading to study discontinuation | 0 | 0 | 0 | 0 |
| Any serious TEAEs | 0 | 8 (3) | 0 | 0 |
| Treatment-related | 0 | 0 | 0 | 0 |
| Treatment-emergent MAAEs | 11 (4) | 49 (19) | 7 (7) | 0 |
| Treatment-related | 0 | 1 (<1) | 1 (1) | 0 |
| PIMMC | 0 | 1 (<1) | 0 | 0 |
| Treatment-related | 0 | 0 | 0 | 0 |
| AESIs relevant to COVID-19 | 0 | 1 (<1) | 0 | 0 |

NVX-CoV2373 = 5 µg SARS-CoV-2 rS + 50 µg Matrix-M adjuvant on Day 0, Day 21, Day 189, and Day 357.
<sup>a</sup> Participants in the safety analysis set are counted according to the treatment received to accommodate for treatment errors.
AESI=adverse event of special interest. MAAE=medically attended adverse event. PIMMC=potentially immune-mediated medical condition. TEAE=treatment-emergent adverse event.

### Immunogenicity

After an initial decline following primary vaccination, a third dose of NVX-CoV2373 increased anti-rS IgG titers for the ancestral strain to a level approximately 4-fold higher than that observed after the primary series (**Figure 3A**). Compared with the decline after the primary series, a more gradual decrease in titers occurred after the third dose, with anti-rS IgG titers increasing again after the fourth dose to levels similar to those seen after the third dose. The immune response to Omicron BA.1 demonstrated a similar pattern, with a progressive narrowing of the gap between the magnitude of response to the ancestral strain compared with the Omicron BA.1 strain as the number of doses increased (**Figure 3A**).

**Figure 3.**
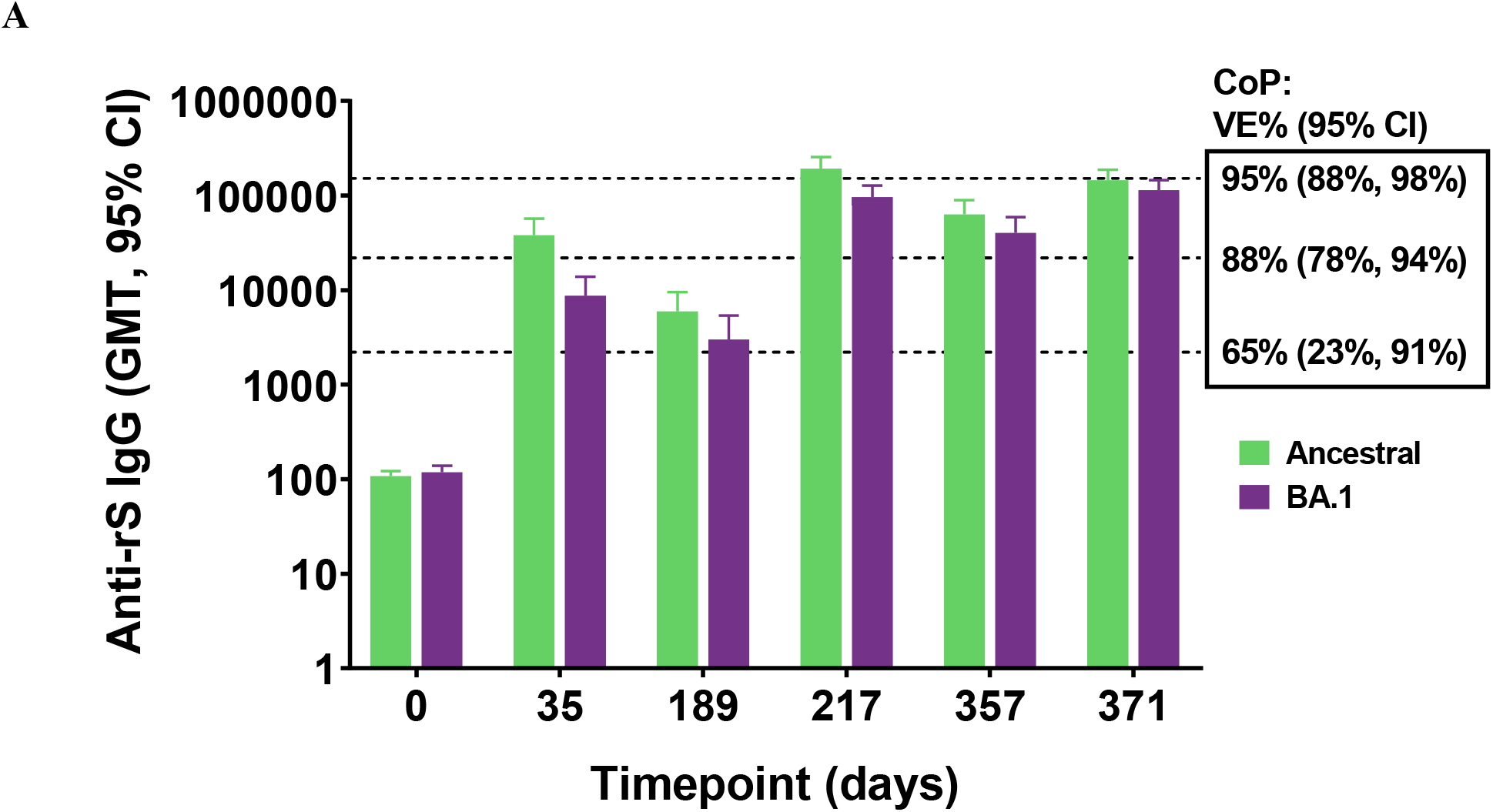
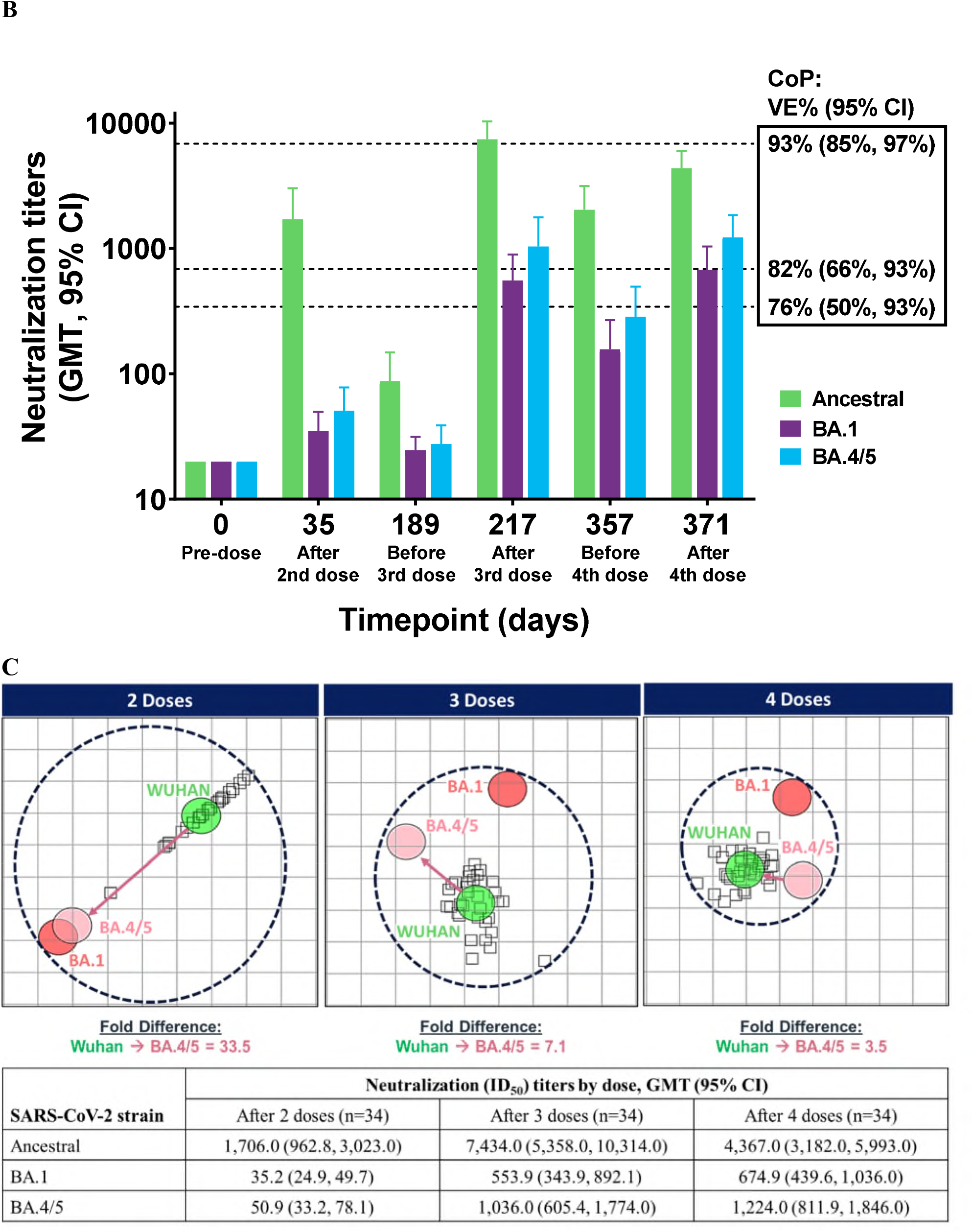
Immunogenicity of NVX-CoV2373 against ancestral and variant strains of SARS-CoV-2, by dose (n=34). (A) Anti-rS IgG titers by dose for the ancestral (n=34) and BA.1 variant (n=31). Data were not available for the BA.1 variant for 3 of 34 participants. Dotted line represents approximate correlates of protection titers as derived for the ancestral strain.^22^ (B) Neutralizing (ID_50_) antibody titers by dose for the ancestral, BA.1, and BA.4/BA.5 variants. Dotted line represents approximate correlates of protection titers as derived for the ancestral strain.^22^ (C) Antigenic cartography by dose for the ancestral and variants strains of SARS-CoV-2. SARS-CoV-2 antigens are depicted as circles and sera are depicted as squares. Each grid square represents one antigenic unit representing a two-fold change in neutralization titers from ancestral to variant. CoP, correlate of protection; VE, vaccine efficacy.

Neutralization titers followed a similar pattern to anti-rS IgG titers. A third dose of NVX-CoV2373 increased SARS-CoV-2 neutralization titers for the ancestral strain to a level approximately 4.7-fold higher than that observed after the primary series, and titers after the fourth dose were similar to those seen after the third dose (**Figure 3B**). The immune response to Omicron BA.1 and BA.4/BA.5 variants demonstrated a similar pattern in neutralization titers.

Antigenic distance mapping of the neutralization titers for multiple SARS-CoV-2 variants demonstrated a reduction in the antigenic distance and fold difference with increased number of doses of NVX-CoV2373 (**Figure 3C**). After the primary series of NVX-CoV2373, the antigenic distance between the Omicron subvariant BA.4/5 and the ancestral strain was 33.5-fold. After the third dose of NVX-CoV2373, the antigenic distance between BA.4/5 and the ancestral strain decreased to 7.1-fold. After the fourth dose, the antigenic distance further decreased to 3.5-fold.

A fit-for-purpose functional hACE2 receptor binding inhibition assay was used to compare hACE2 binding inhibition titers after each dose of NVX-CoV2373 (**Supplemental Figure**). Serum hACE2 inhibition titers increased 15.6-fold from baseline after the primary series (Day 35). From just before the third dose to Day 217, hACE2 inhibition titers increased 50.6-fold. From just before the fourth dose to Day 371, inhibition titers increased another 2.0-fold, for a total 16.6-fold increase over Day 189.

## Discussion

In this report, we describe the first available safety and immunogenicity data after a fourth dose of NVX-CoV2373 from an ongoing phase 2, randomized, observer-blinded, placebo-controlled study. With an increasing number of doses of NVX-CoV2373, there appears to be enhanced cross-reactive immunogenicity without any notable increases in either solicited local or systemic reactogenicity or in unsolicited TEAEs.

The incidence of both unsolicited local and systemic reactogenicity leveled off after the fourth dose of NVX-CoV2373 compared with the increases reported after each of the first three doses. Grade 3+ events remained steady compared with the third dose, except for erythema, which demonstrated a minor increase in the frequency of Grade 3+ events out of the total number of events (15%, n=6). The total incidence of unsolicited TEAEs remained consistent after each dose, with no reports of MAAEs, PIMMCs, or SAEs after the fourth dose. Reactogenicity reported in the present study was similar to that reported for the fourth dose of either BNT162b2 or mRNA-1273, with pain and fatigue the most reported solicited local and systemic reactogenicity events, respectively.^21^

As expected, prior to boosting with a fourth dose at Day 357, anti-SARS-CoV-2 neutralizing antibody titers (ID_50_) to the ancestral strain had gradually decreased after the first booster (third dose) on Day 189, from 7,992 at Day 217 to 2,036 at Day 357. Notably, the decrease in neutralizing antibody titers was more gradual than that seen after the primary series; and titers to the ancestral strain during this period remained similar to or higher than the titers reported in the pivotal Phase 3 efficacy studies that were associated with ∼90% efficacy.^10,11,22^ After the fourth dose of NVX-CoV2373, neutralizing antibody titers (ID_50_) to the ancestral strain increased to 4,367 by Day 371. While this titer was somewhat lower than that seen after the third booster dose, antibody titers were only assessed 14 days after the fourth booster as compared to 28 days after the third dose, and it is possible that further increases would have been seen over the next two weeks. Consistent with other approved vaccines^23^, lower antibody levels were noted for the BA.1 and BA.4/5 variants in comparison with the ancestral strain after the primary series, but significantly increased after the subsequent booster doses. In addition, as can be seen from both Figures 3a and 3b, the magnitude of difference between the BA.1 or BA.4/5 variants and ancestral strain diminished with each booster dose.

Based on a recent preprint on correlates of protection for the NVX-CoV2373,^22^ the antibody titers for the ancestral strain after the first booster remained well above the level associated with 92.8% efficacy (neutralizing antibody titer of 1,000) for the full 6-months prior to the second booster dose administration. If these correlates were to be applied to the BA.1 and BA.5 Omicron variants, they would suggest that efficacy of at least 81.7% (pseudoneutralization titer of 100) might be maintained for these variants during this same timeframe.

A similar immunogenicity pattern as the neutralization titers was seen with anti-S IgG titers. The anti-S IgG antibodies were similar to values reported for mRNA vaccines.^23^ After conversion to similar units, we observed anti-S IgG GMT titers of 1896, 9629, and 7258 BAU/mL after Doses 2, 3, and 4, respectively. Yochay et al noted IgG titers after vaccination with BNT162b2 of 950 BAU/mL 4 weeks after Dose 2, 2102 BAU/mL 4 weeks after Dose 3, and 2975 BAU/mL 2 weeks after Dose 4. A fourth dose of mRNA-1273 resulted in IgG titers of 3502 BAU/mL 2 weeks after Dose 4. These data support the use of NVX-CoV2373 and other SARS-CoV-2 vaccines as repeat boosters to restore immunogenicity against SARS-CoV-2.

Antigenic cartography, a method frequently employed to help evaluate the potential effectiveness of influenza vaccination against variant strains and to guide vaccine strain composition,^14^ provides a more direct visualization of the immunogenicity of vaccine antigen and dose. After additional booster doses following the two-dose primary vaccination series, cross-reactive immunity to SARS-CoV-2 variants was enhanced, the gap between immune recognition of the variants and the ancestral strain was significantly decreased, and a potentially more universal response against the SARS-CoV-2 variants was induced. We believe this phenomenon may be driven by the conserved epitopes found on the recombinant protein vaccine whereby expression of the full-length trimers of the spike protein present epitopes that are conserved across variants for recognition by the immune system. This process may be further enhanced by the saponin-based matrix adjuvant, through epitope spreading.

Our study was subject to certain limitations. As these results are from an ongoing phase 2 study conducted with a limited sample size, the clinical efficacy of the booster dose was not evaluated, and assumptions concerning the efficacy of the booster doses for the ancestral strain and the Omicron variants are based on the derived correlates for the vaccine. Because this study occurred during an ongoing pandemic, a number of initially randomized participants voluntarily discontinued the study between the primary series and the booster doses to receive already approved vaccines. As such, less than 50% of the original study population received the fourth dose, which could introduce population bias. Future studies assessing the efficacy of NVX-CoV2373 after booster doses will be conducted as part of agreed post-marketing commitments with regulatory authorities (e.g., FDA and EMA).

In conclusion, despite the call for variant-specific vaccines, an increase in number of vaccine booster doses with NVX-CoV2373 enhances immunogenicity, and decreases the antigenic gap between the ancestral SARS-CoV-2 strain and its variants without a notable increase in reactogenicity. Therefore, these data suggest that further boosting with the ancestral sequence used in NVX-CoV2373 should retain meaningful utility in preventing variant virus-associated disease.

## Supporting information

CONSORT Checklist

Study Protocol

Addendum

## Data Availability

The trial protocol was a part of the peer-review process and will be included with the published manuscript; more information is available at https://clinicaltrials.gov/ct2/show/NCT04368988.

## Acknowledgments

We thank all of the study participants who volunteered for this study. This study was funded by Novavax, Inc. and initially by the Coalition for Epidemic Preparedness Innovations (CEPI^®^).

The authors would like to thank Lou Fries for his contributions to the development of the manuscript, and Andreana Robertson for biostatistical support of the study. Medical writing support was provided by Kelly Cameron, PhD, CMPP, of Ashfield MedComms, and Inizio company, supported by Novavax, Inc.

## Contributors

RMM, KA, JSP, NP, GMG, and FD were involved in the study design, data collection and interpretation. SG and GC performed the statistical analyses. All authors reviewed, commented on, and approved this manuscript prior to submission for publication. The Authors were not precluded from accessing data in the study, and they accept responsibility to submit the manuscript for publication.

---

**Supplementa1 figure.**
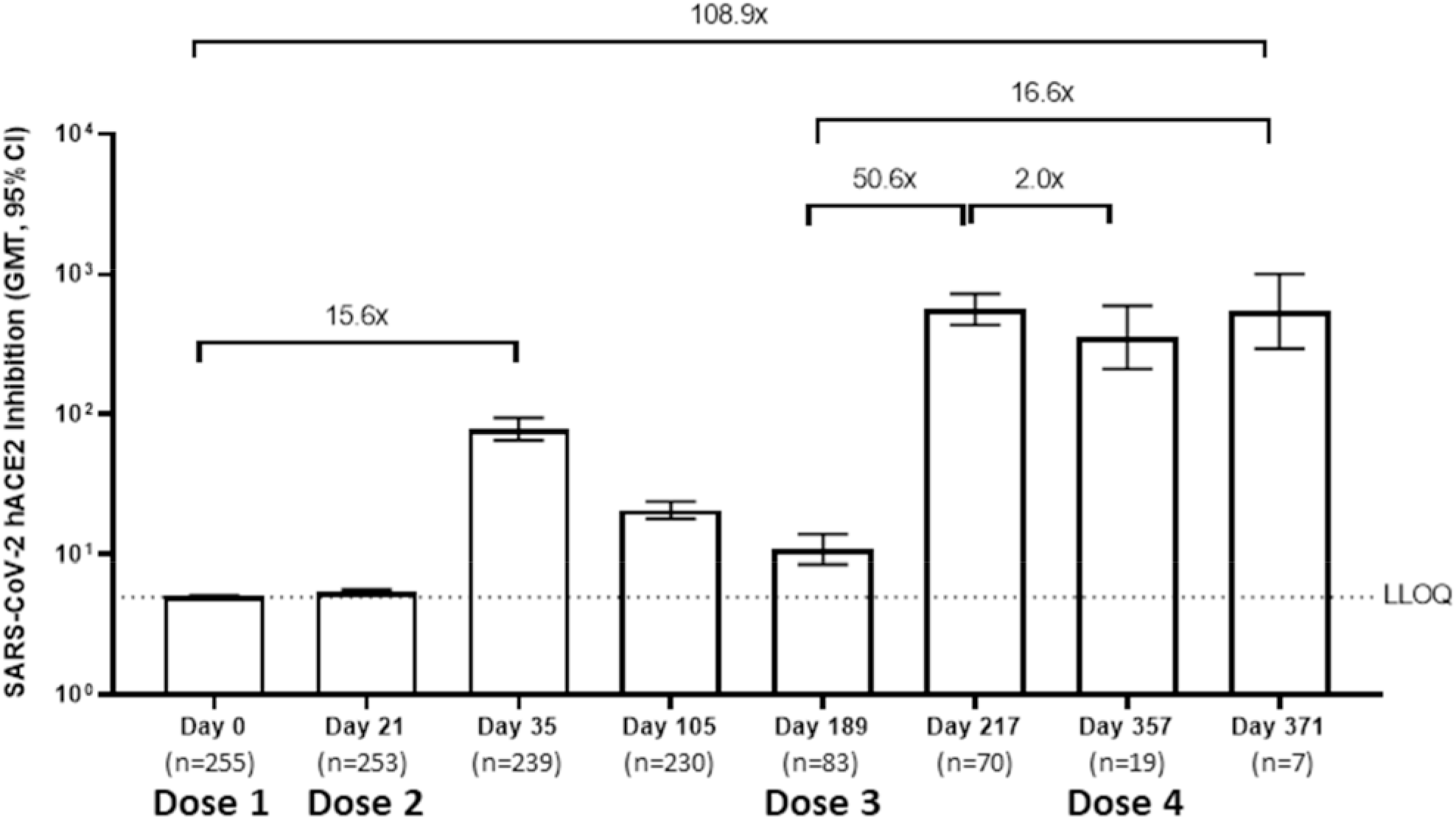
ACE2 inhibition after 4 doses of NVX-CoV2373 (5 µg SARS-CoV-2 rS with 50 µg Matrix-M™ adjuvant). GMFR for various comparisons are depicted above the bars. GMFR, geometric mean fold rise; LLOQ, lower limit of quantitation.

