## Supplementary material for "Immunogenicity and safety of a 4^th^ homologous booster dose of a SARS-CoV-2 recombinant spike protein vaccine (NVX-CoV2373): a phase 2, randomized, placebo-controlled trial": Study Protocol

### CLINICAL STUDY PROTOCOL

### A 2-PART, PHASE 1/2, RANDOMIZED, OBSERVER-BLINDED STUDY TO EVALUATE THE SAFETY AND IMMUNOGENICITY OF A SARS-CoV-2 RECOMBINANT SPIKE PROTEIN NANOPARTICLE VACCINE (SARS-CoV-2 rS) WITH OR WITHOUT MATRIX-M™ ADJUVANT IN HEALTHY SUBJECTS

#### PROTOCOL NO. 2019nCoV-101

| | |
|---|---|
| **Sponsor:** | Novavax, Inc.<br>21 Firstfield Road<br>Gaithersburg, MD 20878<br>United States |
| **Sponsor Contact:** | [redacted] |
| **Medical Monitor:** | [redacted]<br>PPD<br>[redacted] |
| **Bioanalytical Laboratories** | Novavax, Inc.<br>Clinical Immunology<br>21 Firstfield Road<br>Gaithersburg, MD 20878<br>[redacted]<br><br>360biolabs<br>[redacted]<br><br>Cellular Technology Limited<br>[redacted] |

P23 Labs, LLC
[redacted]

PPD Global Central Labs
[redacted]

**Version of Protocol:** Amendment 8/Version 9.0

**Date of Protocol:** 03 May 2021

#### CONFIDENTIAL

The concepts and information contained in this document or generated during the study are considered proprietary and may not be disclosed in whole or in part without the expressed, written consent of Novavax, Inc.

The study will be conducted according to the International Council for Harmonisation Guideline E6(R2): Good Clinical Practice.

### SIGNATURE PAGE

**PROTOCOL TITLE:** A 2-Part, Phase 1/2, Randomized, Observer-Blinded Study to Evaluate the Safety and Immunogenicity of a SARS-CoV-2 Recombinant Spike Protein Nanoparticle Vaccine (SARS-CoV-2 rS) With or Without Matrix-M™ Adjuvant in Healthy Subjects

**PROTOCOL NUMBER:** 2019-nCoV-101

[redacted]

03-May-21 | 11:31 EDT

Date

[redacted]

03-May-21 | 10:03 EDT

Date

[redacted] Clinical Operations
Novavax, Inc.

### INVESTIGATOR PROTOCOL AGREEMENT PAGE

I agree to conduct the study as outlined in the protocol titled "A 2-Part, Phase 1/2, Randomized, Observer-Blinded Study to Evaluate the Safety and Immunogenicity of a SARS-CoV-2 Recombinant Spike Protein Nanoparticle Vaccine (SARS-CoV-2 rS) With or Without Matrix-M™ Adjuvant in Healthy Subjects" in accordance with all guidelines, including International Council for Harmonisation guidelines, and all applicable government regulations. I have read and understand all sections of the protocol.

_______________________________________ _________________________

Signature of Investigator Date

_______________________________________

Printed Name of Investigator

14.3.4 Exploratory Analyses 135
14.4 HANDLING OF MISSING DATA 135
14.5 INTERIM ANALYSES 135
**15. REFERENCE LIST 137**
**16. APPENDICES 139**
16.1 APPENDIX 1: LIST OF ABBREVIATIONS 139
16.2 APPENDIX 2: STUDY GOVERNANCE 142
16.2.1 Data Quality Assurance 142
16.2.2 Investigator Obligations 142
16.2.2.1 Confidentiality 142
16.2.2.2 Institutional Review 143
16.2.2.3 Subject Consent 143
16.2.2.4 Study Reporting Requirements 144
16.2.2.5 Financial Disclosure and Obligations 144
16.2.2.6 Investigator Documentation 144
16.2.2.7 Study Conduct 145
16.2.2.8 Case Report Forms and Source Documents 145
16.2.2.9 Adherence to Protocol 146
16.2.2.10 Reporting Adverse Events 146
16.2.2.11 Investigator's Final Report 146
16.2.2.12 Records Retention 147
16.2.2.13 Publications 147
16.2.3 Study Management 147
16.2.3.1 Monitoring 147
16.2.3.2 Management of Protocol Amendments and Deviations 148
16.2.3.3 Study Termination 150
16.2.3.4 Final Report 150
16.3 APPENDIX 3: FDA TOXICITY GRADING SCALES 151
16.4 APPENDIX 4: LISTINGS OF ADVERSE EVENTS OF SPECIAL INTEREST 155

### LIST OF TABLES

Table 3-1 Treatment Groups (Part 1) ... 29

Table 3-2 Schedule of Events (Part 1) ... 32

Table 7-1 Width of the 95% Confidence Interval (based on the Clopper-Pearson method) ... 54

Table 10-1 Treatment Groups as Originally Planned (Part 2) ... 90

Table 10-2 Treatment Groups, Re-randomized At Day 189 (Part 2) ... 91

Table 10-3 Treatment Groups B and C, Additional Dose on Day 357 (Part 2) ... 91

Table 10-4 Schedule of Events (Part 2) for Subjects in All Treatment Groups ... 95

Table 10-5 Schedule of Events: Treatment Groups B and C, Day 357 to EOS (Part 2) ... 98

Table 13-1 Qualifying Symptoms of Suspected COVID-19 Disease ... 122

Table 13-2 Endpoint Definitions of COVID-19 Disease Severity ... 125

Table 14-1 Width of the 95% Confidence Interval (based on the Clopper-Pearson method) ... 128

Table 14-2 Power of Detecting Difference in Seroconversion Rates between Treatment Groups ... 128

Table 14-3 Power of Detecting Ratio of Geometric Means between Treatment Groups ... 130

Table 16-1 FDA Toxicity Grading Scale for Clinical Abnormalities (Local and General Systemic Reactogenicity) ... 151

Table 16-2 Modified FDA Toxicity Grading Scale for Clinical Abnormalities (Local and General Systemic Reactogenicity) ... 152

Table 16-3 FDA Toxicity Grading Scale for Laboratory Abnormalities ... 153

Table 16-4 FDA Toxicity Grading Scale for Clinical Abnormalities (Vital Signs) ... 154

Table 16-5 Potential Immune-Mediated Medical Conditions (PIMMC) ... 155

Table 16-6 Adverse Events of Special Interest Relevant to COVID-19[a] ... 156

### SECTION 1 - PART 1 OF STUDY

### PROTOCOL SYNOPSIS (PART 1)

**PROTOCOL NO.:** 2019nCoV-101

**PROTOCOL TITLE:** A 2-Part, Phase 1/2, Randomized, Observer-Blinded Study to Evaluate the Safety and Immunogenicity of a SARS-CoV-2 Recombinant Spike Protein Nanoparticle Vaccine (SARS-CoV-2 rS) With or Without Matrix-M™ Adjuvant in Healthy Subjects

**STUDY PHASE:** Phase 1 (Part 1), with expansion to Phase 2 (Part 2) following a review of safety and immunogenicity.

**STUDY SITES:** For Phase 1 (Part 1), up to 3 clinical sites across Australia (and potentially the United States). Refer to Section 2 for Phase 2 (Part 2) study site information.

**OBJECTIVES AND ENDPOINTS:**

**The primary objectives are:**

- To accumulate a safety experience for the candidate vaccine in healthy adult subjects based on solicited short-term reactogenicity and safety laboratory assessments following each vaccination (by toxicity grade).
- To assess the immune response (IgG antibody to SARS-CoV-2 rS protein) at selected early time points through Day 35 following first and second vaccination with or without Matrix-M™ adjuvant at 2 dose levels per SARS-CoV-2 rS construct.

**The secondary objectives are:**

- To assess overall safety through 49 days after first vaccination for all AEs; through 105 days after first vaccination for all MAAEs; and through 365 days after final vaccination for any MAAE attributed to study vaccine, AESI, or SAE.
- To describe the amplitude, kinetics, and durability of immune responses to the various regimens in terms of ELISA units of serum IgG antibodies to SARS-CoV-2 rS protein(s) at selected time points, including longer immune persistence; and amplitude of adjuvant effect. To include reverse cumulative distribution curves.
- To describe the immune responses to the various regimens in terms of titers or concentrations of serum antibodies competitive with a monoclonal antibody specific for the receptor binding domain of SARS coronavirus, or an alternative receptor domain-specific assay, at selected time points. Fc epitope mapping to include high affinity to the ACE-2 binding site.
- To describe the immune protection to the various regimens in terms of neutralizing antibody (eg, a chimeric test virus expressing the SARS-CoV-2 rS protein or wild-type virus under appropriate settings), when such assay systems are available.
- To assess cell-mediated response to differentiate Th1 or Th2 predominance by various vaccine regimens (eg, IL-2, IL-4, IL-5, IL-6, IL-13, TNFα, INFγ using flow cytometry, ELISpot, or other system) in whole blood or harvested PBMC cells (in response to in vitro stimulation with SARS-CoV-2 rS protein).

**Exploratory objectives are:**

- To describe the immune responses to the various regimens in terms of protection using a passive antibody live virus challenge model with wildtype SARS-CoV-2 or variant, when available and appropriate.
- To describe the amplitude, kinetics, and durability of immune responses to the various regimens in terms of ELISA units of other immunoglobulins and immunoglobulin subtypes (eg, IgG1, IgG2, IgE) at selected early time points.
- To develop additional assays to best characterize the immune response for future vaccine development needs.

**The primary endpoints are:**

- Numbers and percentages (with 95% CIs) of subjects with solicited AEs (local, systemic) for 7 days following each primary vaccination (Days 0, 21) by severity score, duration, and peak intensity. In the case of no reactogenicity, a toxicity score of zero (0) will be applied.
- Safety laboratory values (serum chemistry, hematology) by FDA toxicity scoring (absolute and change from baseline where identified) at 7 days after each vaccination. In the case of a toxicity score of zero (0) will be applied.
- Serum IgG antibody levels specific for the SARS-CoV-2 rS protein antigen(s) as detected by ELISA at Day 21 and Day 35. Derived/calculated endpoints based on these data will include geometric mean ELISA units, geometric mean fold rise, and seroconversion rate (proportion of subjects with ≥4-fold rises in ELISA units).

**The secondary endpoints are:**

- Unsolicited AEs (eg, treatment-emergent, serious, suspected unexpected serious, those of special interest, all MAAEs) through the first 49 days by MedDRA classification, severity score, and relatedness.
- Vital sign measurements with severity scoring immediately following vaccination. Descriptive statistics (mean, SD, change from baseline) by treatment group, by visit.
- All MAAEs through 105 days after first vaccination; and any MAAE related to vaccine (as determined by the investigator), AESI (predefined and including COVID-19 positivity), or SAE through 365 days after final vaccination.
- Serum IgG antibody levels specific for the SARS-CoV-2 rS protein antigen(s) as detected by ELISA (refer to primary endpoints), including seroconversion rates (proportion of subjects with ≥2-fold and ≥4-fold rises in antibody levels) and seroresponse rate (proportion of subjects with rises in ELISA units exceeding the 95$^{th}$ percentile of placebo recipients at the same time point) to be described across time points following first and second vaccination through Day 189.
- Epitope-specific immune responses to the SARS-CoV-2 rS protein receptor binding domain measured by serum titers/concentrations of antibody in a competition ELISA assay using a monoclonal antibody to the similar SARS receptor binding domain or other inhibition assay to that ACE-2 receptor. Derived/calculated endpoints based on

these data will include geometric mean titer or concentration, geometric mean fold rise, seroconversion rate, and seroresponse rate.

- Neutralizing antibody activity compared to baseline in all treatment groups at various time points. Expression units (eg, titers) to be determined during assay development.
- Cell-mediated (Th1/Th2) pathways as measured by whole blood (flow cytometry) and/or in vitro PBMC stimulation (eg, ELISpot, cytokine staining) with SARS-CoV-2 rS protein(s) as measured on Days 0, 7, and 28.

**Exploratory endpoints are:**

- Percentage of protection measured in a passive antibody live virus challenge model using SARS-CoV-2 virus or other modified construct at select time points if available and appropriate. Expression units to be determined during development.
- Additional serum Ig antibody ELISA measurements specific for the SARS-CoV-2 rS protein antigen(s) characterization. Expression units to be determined during development.
- Any additional assays to measure immune response, protection, or potential safety signals.

**STUDY DESIGN:**

Part 1 is a first-in-human, Phase 1, randomized, placebo-controlled, observer-blinded study to evaluate the safety and immunogenicity of a SARS-CoV-2 recombinant spike protein nanoparticle vaccine (SARS-CoV-2 rS) with or without Matrix-M adjuvant in male and female subjects. Subjects will be healthy adults based on medical history, physical examination, and baseline clinical laboratory testing and serology tests.

After signing the ICF, subjects may be screened within a window of up to 30 days. Subjects will be asked to provide consent for the use of samples for future testing or assay development specific to SARS-CoV-2 (or related variants). The study design for Part 1 allows up to 2 unique SARS-CoV-2 rS constructs to be investigated.

A minimum of 100 healthy adult male and female subjects between 18 and 59 years of age (inclusive) per SARS-CoV-2 rS construct, along with 25 control subjects receiving only placebo, are planned for block randomization. In addition, 6 sentinel subjects per unique construct will be enrolled in an unblinded fashion (ie, in addition to those block randomized). The total number of subjects in Part 1 will not exceed 131 subjects if a single cohort is included; 237 subjects if 2 cohorts are included and randomized in parallel; or 262 subjects if 2 constructs are included and randomized in sequence. Subjects who meet the criteria for study entry will be assigned to the current cohort and randomly assigned to a treatment group (Table S1-1).

Sentinel dosing will be utilized for the first vaccination, where the first 6 subjects enrolled in each cohort (ie, 6 sentinel subjects per unique SARS-CoV-2 rS construct investigated) are vaccinated and observed for reactogenicity through 48 hours before the remaining subjects are vaccinated. Sentinel subjects in Cohort 1 will be assigned in a 1:1 ratio to receive either 5 or 25 μg SARS-CoV-2 rS with Matrix-M adjuvant. Treatment assignment for the sentinel subjects will be unblinded.

Optional: Cohort 2 may be enrolled to investigate a unique second SARS-CoV-2 rS nanoparticle construct. If a unique second construct is not available, then the construct used in Cohort 1 may be investigated at lower doses in Cohort 2 (ie, lower doses of either SARS-CoV-2 rS or Matrix-M) based on early immune responses observed in Cohort 1. No more than the proposed total number of subjects will be allowed in Cohort 2; however, fewer treatment groups may be evaluated. For Cohort 2, sentinel subjects will be assigned in a 1:1 ratio to receive same treatment as Group G or Group H (Table S1-1).

**Table S1-1 Treatment Groups (Part 1)**

| | | n | | Day 0 | | | Day 21 (+ 5 days) | | |
|---|---|---|---|---|---|---|---|---|---|
| **Cohort** | **Treatment Group** | **Randomized** | **Sentinel** | **SARS-CoV-2 rS (µg)** | | **Matrix-M (µg)** | **SARS-CoV-2 rS (µg)** | | **Matrix-M (µg)** |
| 1 | A | 25 | – | Placebo | 0 | 0 | Placebo | 0 | 0 |
| | B | 25 | – | Construct A | 25 | 0 | Construct A | 25 | 0 |
| | C | 25 | 3 | Construct A | 5 | 50 | Construct A | 5 | 50 |
| | D | 25 | 3 | Construct A | 25 | 50 | Construct A | 25 | 50 |
| | E | 25 | – | Construct A | 25 | 50 | Placebo | 0 | 0 |
| Maximum number of subjects if 1 construct evaluated: | | 125 | 6 | Total = 131 | | | | | |
| 2[a,b] | A | 0[c] or 25 | – | Placebo | 0 | 0 | Placebo | 0 | 0 |
| | F | 25 | – | Construct B | X[d] | 0 | Construct B | X[d] | 0 |
| | G | 25 | 3 | Construct B | Y[d] | 50 | Construct B | Y[d] | 50 |
| | H | 25 | 3 | Construct B | X[d] | 50 | Construct B | X[d] | 50 |
| | I | 25 | – | Construct B | X[d] | 50 | Placebo | 0 | 0 |
| Maximum number of subjects if 2 constructs evaluated in sequence: | | 250 | 12 | Total = 262 | | | | | |

Note: At least 1 and up to two SARS-CoV-2 rS constructs (ie, Constructs A and B) will be evaluated during Part 1.

a If study vaccine for Cohort 2 (ie, Construct B) is not available at the same time as study vaccine for Cohort 1 (ie, Construct A), then Cohort 2 may be sequentially enrolled. Fewer treatment groups may be evaluated in such a case, based on early immune and safety results in Cohort 1.

b If a unique second construct is not available, then the construct used in Cohort 1 may be investigated at lower doses in Cohort 2 (ie, lower doses of either SARS-CoV-2 rS or Matrix-M) based on early immune responses observed in Cohort 1. No more than the proposed total number of subjects will be allowed in Cohort 2; however, fewer treatment groups may be evaluated.

c If Cohort 2 is enrolled in parallel with Cohort 1, no subjects will be randomized to Cohort 2, Group A.

d X and Y indicate 2 different doses, where X is the higher dose. Doses will be determined based on early immune responses observed in Cohort 1 or as new preclinical data becomes available.

Study vaccinations will comprise 2 IM injections at a 21-day interval (Day 0 and Day 21), ideally in alternate deltoids with the study treatment assigned (ie, placebo or a 5 or 25 µg dose of SARS-CoV-2 rS (Construct A or B), with or without a 50 µg dose of Matrix-M adjuvant), in an injection volume of approximately 0.5 mL (not to exceed 0.7 mL).

Vaccinations will be performed on an outpatient basis by site personnel using a methodology to maintain the blind (eg, unblinded personnel, blinded syringe).

Blood samples for immunogenicity assessments (B and T cell) will be collected before each vaccination and at selected time points following first and second vaccination.

Safety assessments will include monitoring and recording of solicited (local and systemic reactogenicity events) and unsolicited AEs; MAAEs; AESI; SAEs; clinical laboratory results including hematology and serum chemistry; vital sign measurements; and physical examination findings. Relatedness/causality and severity grading will be included.

Vaccination pause rules based on reactogenicity, safety laboratory results, and SAEs related to study participation will be in place to monitor subject safety during the study. A safety monitoring committee will be formed before the first subject is vaccinated and will follow a SMC charter. The SMC will review study progress and subject, clinical, safety, and reactogenicity data ad hoc when a vaccination pause rule is met, for immediate concerns regarding observations during this study, to allow advancement to Part 2 of study, or as needed.

Part 1 will consist of a screening period (Days −30 to 0); vaccination days (Days 0 and 21); outpatient study visits on Day 0, Day 2 (sentinels only), Days 7 (+ 3 days), 21 (+ 5 days), 28 (+3 days), 35 (+ 5 days), 49 (+ 5 days), 105 (± 7 days), and 189 (± 15 days); and an end-of-study telephone call on Day 386 (± 15 days). The duration of the Part 1, excluding screening, is approximately 387 days.

If the Novavax study vaccine or another vaccine from a different manufacturer is demonstrated to be safe and efficacious and approved and/or authorized for use by the regulatory authority in Australia, subjects for whom the new approved/authorized vaccine is recommended and available will be counseled with respect to their options. These subjects may be offered the opportunity to be unblinded so that those who received placebo may be offered the Novavax vaccine or another approved/authorized vaccine, as appropriate, outside the protocol procedures. Subjects who received the Novavax vaccine and who wish to receive an approved/authorized vaccine from another manufacturer will be advised to discuss this plan with their healthcare provider given the current lack of safety data regarding the sequential administration of vaccines made by different manufacturers. Subjects who are unblinded and receive an approved/authorized vaccine in this manner will be strongly encouraged to remain in study for safety follow-up as defined in the protocol. However, subjects also have the right to discontinue participation in the study at any time.

Due to the ongoing pandemic, recent national regulatory and local IRB and public health guidance will be applied at any site location requiring alternatives for a study subject’s ability to attend an investigational site for protocol-specified visits. While the site should do everything in its power to keep activities as per the defined schedule of events, the site should also have a process in place to conduct the needed assessments (eg, telephone contact, virtual visit, alternative location for assessment, including local laboratories or imaging centers) when necessary and feasible, as long as such visits are sufficient to assure the safety of study subjects.

**Vaccination Pause Rules:**
Adverse events meeting any one of the following criteria will result in a hold being placed on subsequent vaccinations pending further review by the safety monitoring committee:

- Any SAE attributed to vaccine.
- Any toxicity grade 3 (severe) solicited single AE term occurring in ≥7 subjects across any single SARS-CoV-2 rS construct following vaccination (first and second vaccinations to be assessed separately).
- Toxicity grade 3 (severe) solicited single prespecified laboratory value occurring in ≥7 subjects across any single SARS-CoV-2 rS construct following vaccination (first and second vaccinations to be assessed separately). Prespecified laboratory values to be evaluated include creatinine, ALT, AST, bilirubin, hemoglobin, complete white blood count, and platelets.
- Any grade 3 (severe) unsolicited single AE preferred term for which the investigator assesses as related which occurs in ≥7 subjects across any single SARS-CoV-2 rS construct, within 49 days following vaccination (first).

The sponsor, along with medical monitor, may request an SMC review for any safety concerns that may arise in the trial and not associated with any specific pause rule.

**STUDY POPULATION:**
**Inclusion Criteria:**
Each subject must meet all of the following criteria to be enrolled in this study:

1. Healthy adult males or females between 18 and 59 years of age, inclusive, at screening. Healthy status will be determined by the investigator based on medical history, clinical laboratory results, vital sign measurements, and physical examination at screening.
2. The subject has a body mass index 17 to 35 kg/m$^2$, inclusive, at screening.
3. Willing and able to give informed consent prior to study enrollment and comply with study procedures.
4. Female subjects of childbearing potential (defined as any female who has experienced menarche and who is NOT surgically sterile [ie, hysterectomy, bilateral tubal ligation, or bilateral oophorectomy] or postmenopausal [defined as amenorrhea at least 12 consecutive months or documented plasma follicle-stimulating hormone level ≥40 mIU/mL]) must agree to be heterosexually inactive from at least 21 days prior to enrollment and through 6 months after the last vaccination OR agree to consistently use any of the following methods of contraception from at least 21 days prior to enrollment and through 6 months after the last vaccination:
    a. Condoms (male or female) with spermicide (if acceptable in country)
    b. Diaphragm with spermicide
    c. Cervical cap with spermicide
    d. Intrauterine device
    e. Oral or patch contraceptives

f. Norplant®, Depo-Provera®, or other in country regulatory-approved contraceptive method that is designed to protect against pregnancy
g. Abstinence, as a form of contraception, is acceptable if in line with the subject's lifestyle

**NOTE**: Periodic abstinence (eg, calendar, ovulation, symptothermal, post-ovulation methods) and withdrawal are not acceptable methods of contraception. These procedures and laboratory test results must be confirmed by physical examination, by subject recall of specific date and hospital/facility of procedure, or by medical documentation of said procedure.

**Exclusion Criteria:**
Subjects meeting any of the following criteria will be excluded from the study:

1. Any ongoing, symptomatic acute or chronic illness requiring medical or surgical care, inclusive of changes in medication in the past 2 months indicating that chronic illness/disease is not stable (at the discretion of the investigator). This includes any current workup of undiagnosed illness that could lead to a new condition.
2. Chronic disease inclusive of: a) hypertension uncontrolled for age according to JNC 8 guidelines; b) congestive heart failure by NYHA functional classification of ≥II; c) chronic obstructive pulmonary disease by GOLD classification of ≥2; d) recent (within 6 months prior to first study vaccination) exacerbation of coronary artery disease as manifested by cardiac intervention, addition of new cardiac medications for control of symptoms, or unstable angina; e) asthma (diagnosed by spirometry showing reversibility of disease and must meet at least the Step 1 classification with current prescription/use of medications to control symptoms); f) diabetes requiring use of medicine (insulin or oral) or not controlled with diet.
3. Participation in research involving an investigational product (drug/biologic/device) within 45 days prior to first study vaccination.
4. History of a confirmed diagnosis of SARS or COVID-19 disease (confirmed by a specific test for each disease) or known exposure to a SARS-CoV-2 positive confirmed close contact (eg, family member, housemate, daycare provider, aged parent requiring care), at the discretion of the investigator.

**NOTE**: If exposure is suspected, subjects may be enrolled with subsequent documentation of a negative test for SARS-CoV-2, at the discretion of the investigator.

5. Currently working in an occupation with a high risk of exposure to SARS-CoV-2 (eg, healthcare worker, emergency response personnel).
6. Currently taking any product (investigational or off-label) for prevention of COVID-19 disease.
7. Positive tests for SARS-CoV-2 (either ELISA IgG or PCR) at screening or prior to first vaccination. Testing may be repeated during the screening period if exposure to SARS-CoV-2 is suspected, at the discretion of the investigator.
8. Received influenza vaccination within 14 days prior to first study vaccination, or any other vaccine within 4 weeks prior to first study vaccination.

9. Any autoimmune or immunodeficiency disease/condition (iatrogenic or congenital).

**NOTE:** Stable endocrine disorders that have a confirmed autoimmune etiology (eg, thyroid, pancreatic), including stable diet-controlled diabetes, are allowed at the discretion of the investigator.

10. Chronic administration (defined as more than 14 continuous days) of immunosuppressant, systemic glucocorticosteroids, or other immune-modifying drugs within 90 days prior to first study vaccination; or anticipation of the need for immunosuppressive treatment within 6 months after last vaccination.

**NOTE**: An immunosuppressant dose of glucocorticoid is defined as a systemic dose ≥10 mg of prednisone per day or equivalent. The use of topical and nasal glucocorticoids will be permitted. Use of inhaled glucocorticoids is prohibited.

11. Received immunoglobulin, blood-derived products, or other immunosuppressant drugs within 90 days prior to first study vaccination.
12. Any acute illness concurrent or within 14 days prior to first study vaccination (medical history and/or physical examination) or documented temperature of >38°C during this period. This includes respiratory or constitutional symptoms consistent with SARS-CoV-2 (COVID-19) exposure (ie, cough, sore throat, difficulty breathing)

**NOTE**: This is a temporary exclusion for which the subject may be re-evaluated if they remain free from acute illness after 14 days and are test-negative for COVID-19 if symptoms indicate exposure. Subjects may be re-evaluated anytime during the screening period.

13. Known disturbance of coagulation (iatrogenic or congenital).

**NOTE**: The use of ≤325 mg of aspirin per day as prophylaxis is permitted, but the use of other platelet aggregation inhibitors, thrombin inhibitors, Factor Xa inhibitors, or warfarin derivatives is exclusionary, regardless of bleeding history, because these imply treatment or prophylaxis of known cardiac or vascular disease.

14. Evidence of Hepatitis B or C or HIV by laboratory testing.
15. A positive test result for drugs of abuse (except a positive test result associated with prescription medication that has been reviewed and approved by the investigator) at screening.

**NOTE**: A positive tetrahydrocannabinol result will not be exclusionary but may result in a decision by the investigator to exclude subjects due to concerns of impairment in the quality of safety reporting.

16. Any neurological disease or history of significant neurological disorder (eg, meningitis, seizures, multiple sclerosis, vasculitis, migraines, Guillain-Barré syndrome [genetic/congenital or acquired]).

**NOTE**: Significant neurological migraine includes frequent migraine (2 times a month or greater), migraine with aura or migraine with complications (status migrainosus, persist aura without infarction, infarction or aura triggered seizure defined by International Classification of Headache Disorders-3.

17. Active cancer (malignancy) within 5 years prior to first study vaccination (with the exception of adequately treated non-melanomatous skin carcinoma, at the discretion of the investigator)
18. Vital sign (blood pressure, pulse, temperature) abnormalities of toxicity grade >1.

**NOTE**: Vital sign measurements may be repeated once during the screening period to allow inclusion with the most recent measurement taken as the baseline value.

19. Clinical laboratory abnormalities of toxicity grade >1 for selected serum chemistry and hematology parameters.

**NOTE**: Clinical laboratory testing may be repeated once during the screening period to allow inclusion with the most recent measurement taken as the baseline value.

20. Any known allergies to products contained in the investigational product or latex allergy.
21. Women who are pregnant, breastfeeding, or who plan to become pregnant during the study.
22. History of alcohol abuse or drug addiction within one year prior to the first study vaccination.
23. Any condition that, in the opinion of the investigator, would pose a health risk to the subject if enrolled or could interfere with evaluation of the study vaccine or interpretation of study results (including neurologic or psychiatric conditions deemed likely to impair the quality of safety reporting).
24. Study team member or first-degree relative of any study team member (inclusive of sponsor, PPD, and site personnel involved in the study).

**Other Considerations:**
Subjects meeting any of the following criteria may be delayed for a second vaccination:

- Respiratory symptoms in the past 3 days (ie, temperature of >38°C, cough, sore throat, difficulty breathing). Subject may be vaccinated once all symptoms have been resolved for >3 days. Out of window vaccination is allowed for this reason.
- Any acute illness (eg, gastroenteritis, migraine, urinary tract infection, injury) that is causing symptoms that could, in the opinion of the investigator, impact the assessment of reactogenicity or safety laboratory tests. Subject may be vaccinated once symptoms have resolved or are stabilized for >3 days. Out of window vaccination is allowed for this reason.
- Immunization with any vaccine within 14 days prior to second vaccination. Out of window vaccination is allowed for this reason.

**STUDY TREATMENTS:**
Study vaccinations will comprise 2 IM injections at a 21-day interval (Day 0 and Day 21), ideally in alternate deltoids with the study treatment assigned (ie, placebo or a 5 or 25 µg dose of SARS-CoV-2 rS [Construct A or B], with or without a 50 µg dose of Matrix-M adjuvant), in an injection volume of approximately 0.5 mL (not to exceed 0.7 mL).

**STUDY PROCEDURES:**

**Safety Assessments:**

Safety assessments will include monitoring and recording of solicited (local and systemic reactogenicity events) and unsolicited AEs; MAAEs; AESI; SAEs; clinical laboratory results including hematology and serum chemistry; vital sign measurements; and physical examination findings.

**Immunogenicity Assessments:**

Blood samples for immunogenicity assessments will be collected before vaccination and at selected early time points following vaccination. Immune measurements (ELISA) will be conducted on serum (IgG) for SARS-CoV-2 rS protein antigen(s). Additional immunogenicity assessments specific to SARS-CoV-2 (or related variants) include human ACE-2 receptor inhibition assay, specific monoclonal antibody competition assays to other SARS-CoV-2 epitopes, neutralizing antibody and other tests of immune protection using passive antibody live virus challenge models (if available and appropriate). Cell-mediated immunity by measurement of cytokines produced using whole blood or after in vitro stimulation of PBMCs will occur at selected time points for all subjects. Aliquots of all collected samples from this study may be retained for additional testing of antigens specific to SARS-CoV-2 (or related variants) for a maximum of 25 years (starting from the date at which the last subject had the last study visit), unless local rules, regulations, or guidelines require different timeframes or different procedures, in accord with subject consent.

**STATISTICAL ANALYSIS PLANS:**

**Sample Size:**

The sample size for this study is based on clinical and practical considerations and not on a formal statistical power calculation. The sample size is considered sufficient to evaluate the objectives of the study. With 25 subjects in each treatment group, there is a 92.8% probability to observe at least 1 subject with an AE if the true incidence of the AE is 10% and a 72.3% probability if the true incidence of the AE is 5%. With 100 subjects receiving at least 1 vaccination per SARS-CoV-2 rS construct, there is a greater than 99% probability to observe at least 1 subject with an AE if the true incidence of the AE is at least 5%.

**Analysis Sets:**

The safety analysis set will include all subjects who receive at least 1 dose of study vaccine (SARS-CoV-2 rS or placebo). Subjects will be analyzed according to the vaccine actually received.

The per-protocol analysis set will be determined for each study visit and will include all subjects who receive at least 1 dose of study vaccine (SARS-CoV-2 rS or placebo), have at least a baseline and 1 serum sample result available after vaccination, and have no major protocol violations that impact immunogenicity response at the corresponding study visit. All subjects in the per-protocol analysis set will be analyzed according to the study vaccine the subject was randomized to receive and not according to what was actually received, in the event there is a discrepancy.

The intent-to-treat analysis set will include all subjects who are randomized, regardless of protocol violations or missing data. The intent-to-treat analysis set will be used for supportive analyses.

**Safety Analyses:**
Numbers and percentages (with 95% CIs based on the Clopper-Pearson method) of subjects with solicited local and systemic AEs through 7 days after each vaccination will be summarized by treatment group and the maximum toxicity grade over 7 days after each vaccination. The duration of solicited local and systemic AEs after each vaccination will also be summarized by treatment group.

Unsolicited AEs will be coded by preferred term and system organ class using the latest version of MedDRA and summarized by treatment group as well as by severity and relationship to study vaccine. Adverse events through 49 days after first vaccination; all MAAEs through 105 days after first vaccination; and any MAAE related to vaccine, SAE, or AESI through 365 days after final vaccination will be listed separately and summarized by treatment group.

Actual values, changes from baseline (where indicated), and toxicity grading for clinical safety laboratory test results and vital sign measurements will be summarized by treatment group at each timepoint using descriptive statistics.

Concomitant medications will be summarized by treatment group and preferred drug name as coded using the World Health Organization drug dictionary.

**Immunogenicity Analyses:**
The primary immunogenicity analyses will be performed using the per-protocol analysis set.

For the serum IgG antibody levels specific for the SARS-CoV-2 rS protein antigen(s) as detected by ELISA, the geometric mean at each study visit, and the geometric mean fold rise comparing to the baseline (Day 0) at each post-vaccination study visit, along with 95% CI will be summarized by treatment group. The 95% CI will be calculated based on the t-distribution of the log-transformed values for geometric means or geometric mean fold rises, then back transformed to the original scale for presentation. The seroconversion rate (proportion of subjects with ≥2-fold and also ≥4-fold rises in ELISA units), and seroresponse rate (proportion of subjects with rises in ELISA units exceeding the 95$^{th}$ percentile of placebo recipients at the same time point) along with 95% CIs based on the Clopper-Pearson method will be summarized by treatment group at each post-vaccination study visit. An ANCOVA model will be constructed at each post-vaccination study visit on the log-transformed titer, including the treatment group as a fixed effect and the baseline log-transformed titer as a covariate. Comparisons of selected treatment groups will be performed within each visit. Additional covariates, such as site and age, may be explored as supportive analyses. Difference in the seroconversion rate and seroresponse rate between selected treatment groups along with 95% CIs within each visit will be calculated using the method of Miettinen and Nurminen.

Neutralization assay specific for the SARS-CoV-2 wildtype (or variant) will be developed and appropriate testing parameters similar to ELISA will be utilized to assess functional antibody response. Refer to the laboratory manual and SAP.

Cell-mediated response for both Th1 and Th2 pathways will be assessed by cytokine profiling in either whole blood with flow cytometry and/or harvested PBMCs and summarized by treatment group in the per-protocol analysis set. Refer to the laboratory manual and SAP.

Similar summaries will be generated for the other immunogenicity endpoints identified in the future.

**Interim Analyses:**
Due to the nature of the pandemic, rolling interim analyses are planned for rapid vaccine development. The sentinel safety group of 6 subjects (assigned unblinded active vaccine constructs with Matrix-M) will be tested (ELISA and neutralization assay) per construct (cohort) immediately after Day 21 and Day 35 for early awareness of vaccine responsiveness.

Interim limited analyses at the aggregate treatment level only will be conducted when all subjects in a cohort have completed their respective Days 21, 35, 49, 105, and/or 189 visits. Refer to the SAP for more details. These interim analyses will allow decisions to initiate investigation of further nanoparticle constructs/doses, including advancement to Part 2 of study (based on Day 35 data), and will include (at a minimum) the following:

- Day 21 – Demographics and baseline characteristics, reactogenicity (local and systemic) following first vaccination (by toxicity grade and day), ELISA IgG GMT (baseline and Day 21) and seroconversion rate (≥4-fold), neutralization response (baseline and Day 21) expressed as titer or LD50.
- Day 35 – Reactogenicity (local and systemic) following first and second vaccination (by toxicity grade and day), laboratory values (by toxicity grading), vital signs (by toxicity grading), AEs (by classifications), ELISA IgG GMT (baseline, Day 21, Day 35) and seroconversion rate (≥4-fold), neutralization response (baseline, Day 21, Day 35) expressed as titer or LD50.

Informational unblinded analysis will be provided to enable a safety review by the SMC through Day 35 on all subjects with notification of recommendation to advance into Part 2 of study. Refer to the SMC charter for more details. Additional limited interim analyses at the aggregate level may be performed at Days 49, 105, and/or Day 189 to inform on longer term safety and immune persistence if needed. The SAP will pre-specify such analyses but may be updated to reflect learnings based on the prior analyses in such instances. The final database lock will occur after Day 386 and include all primary and secondary endpoints.

**DATE OF PROTOCOL:** 03 May 2021

### 1. INTRODUCTION

#### 1.1 BACKGROUND

Novavax, Inc. is developing a recombinant vaccine adjuvanted with the saponin-based Matrix-M™ (previously referred to as Matrix-M) for the prevention of disease caused by the SARS-CoV-2 virus. The newly described coronavirus' genetic relationship with the 2002-2003 severe acute respiratory syndrome coronavirus (SARS-CoV) has resulted in adoption of the name SARS-CoV-2, with the associated disease being referred to as coronavirus disease 2019 (COVID-19). SARS-CoV-2 recombinant spike (S) protein nanoparticle vaccine (SARS-CoV-2 rS) is constructed from the full length wild-type SARS-CoV-2 S glycoprotein (GP) based upon the GenBank gene sequence MN908947, nucleotides 21563-25384. The S protein is a type 1 trimeric glycoprotein of 1,273 amino acids that is produced as an inactive S0 precursor. The *S*-gene was codon optimized for expression in *Spodoptera frugiperda* (Sf9) insect cells. The SARS-CoV-2 rS nanoparticle vaccine is intended for administration with or without Matrix-M adjuvant, which is a saponin-based adjuvant that has been shown to enhance the immunogenicity of nanoparticle vaccines in nonclinical and clinical studies. Reference the current version of the Matrix-M adjuvant safety supplement to the IB for additional details (Novavax 2020).

Further details on the study vaccine can be found in the IB (Novavax 2020).

#### 1.2 RATIONALE FOR STUDY

The purpose of this 2-Part, Phase 1/2, first-in-human, randomized, placebo-controlled, observer-blinded study, is to evaluate the safety and immunogenicity of a SARS-CoV-2 recombinant spike protein nanoparticle vaccine (SARS-CoV-2 rS) with or without Matrix-M adjuvant in healthy subjects. The primary objectives of Part 1 of this study are to accumulate a safety experience for the candidate vaccine in healthy adult subjects based on solicited short-term reactogenicity and safety laboratory assessments following each vaccination (by toxicity grade); and to assess the immune response (IgG antibody to SARS-CoV-2 rS protein) at selected early time points following first and second vaccination with or without Matrix-M adjuvant at 2 dose levels per SARS-CoV-2 rS construct. The information provided by the Part 1/Phase 1 will inform progression of the vaccine to the Part 2/Phase 2 portion of this study.

#### 1.3 RATIONALE FOR DOSE SELECTION

The proposed dose levels of SARS-CoV-2 rS to be evaluated in this study are based on several data sources, including nonclinical studies, and nonclinical and clinical studies with other Novavax, Inc. baculovirus-Sf9-produced nanoparticle vaccines including safety data in over 14,700 subjects from clinical studies with EBOV, RSF F, and influenza vaccines. For details on nonclinical studies of SARS-CoV-2 rS and for details on the ongoing clinical studies with Novavax, Inc. baculovirus-Sf9-produced nanoparticle vaccines, please refer to the IB (Novavax 2020).

The proposed dose level of Matrix-M adjuvant to be evaluated in this study is based on the fully-analyzed human experience to date with Matrix-M adjuvants, which is confined to adults who have received 1- to 3-dose series of IM doses of 25 to 75 µg. Further details regarding the Matrix-M adjuvant, including safety data from over 4200 subjects in clinical studies with EBOV, RSV F, malaria, rabies, herpes simplex virus, and influenza vaccines with Matrix-M, are provided in the current version of the Matrix-M adjuvant safety supplement to the IB (Novavax 2020).

### 2. STUDY OBJECTIVES AND ENDPOINTS

#### 2.1 STUDY OBJECTIVES

The primary objectives are:

- To accumulate a safety experience for the candidate vaccine in healthy adult subjects based on solicited short-term reactogenicity and safety laboratory assessments following each vaccination (by toxicity grade).
- To assess the immune response (IgG antibody to SARS-CoV-2 rS protein) at selected early time points through Day 35 following first and second vaccination with or without Matrix-M adjuvant at 2 dose levels per SARS-CoV-2 rS construct.

The secondary objectives are:

- To assess overall safety through 49 days after first vaccination for all AEs; through 105 days after first vaccination for all MAAEs; and through 365 days after final vaccination for any MAAE attributed to study vaccine, AESI, or SAE.

- To describe the amplitude, kinetics, and durability of immune responses to the various regimens in terms of ELISA units of serum IgG antibodies to SARS-CoV-2 rS protein(s) at selected time points, including longer immune persistence; and amplitude of adjuvant effect. To include reverse cumulative distribution curves.

- To describe the immune responses to the various regimens in terms of titers or concentrations of serum antibodies competitive with a monoclonal antibody specific for the receptor binding domain of SARS coronavirus, or an alternative receptor domain-specific assay, at selected time points. Fc epitope mapping to include high affinity to the ACE-2 binding site.

- To describe the immune protection to the various regimens in terms of neutralizing antibody (eg, a chimeric test virus expressing the SARS-CoV-2 rS protein or wild-type virus under appropriate settings), when such assay systems are available.

- To assess cell-mediated response to differentiate Th1 or Th2 predominance by various vaccine regimens (eg, IL-2, IL-4, IL-5, IL-6, IL-13, TNFα, INFγ using flow cytometry, ELISpot, or other system) in whole blood or harvested PBMC cells (in response to in vitro stimulation with SARS-CoV-2 rS protein).

The exploratory objectives are:

- To describe the immune responses to the various regimens in terms of protection using a passive antibody live virus challenge model with wildtype SARS-CoV-2 or variant, when available and appropriate.

- To describe the amplitude, kinetics, and durability of immune responses to the various regimens in terms of ELISA units of other immunoglobulins and immunoglobulin subtypes (eg, IgG1, IgG2, IgE) at selected early time points.

- To develop additional assays to best characterize the immune response for future vaccine development needs.

#### 2.2 STUDY ENDPOINTS

The primary endpoints are:

- Numbers and percentages (with 95% CIs) of subjects with solicited AEs (local, systemic) for 7 days following each primary vaccination (Days 0, 21) by severity score, duration, and peak intensity. In the case of no reactogenicity, a toxicity score of zero (0) will be applied.
- Safety laboratory values (serum chemistry, hematology) by FDA toxicity scoring (absolute and change from baseline where identified) at 7 days after each vaccination. In the case of a toxicity score of zero (0) will be applied.
- Serum IgG antibody levels specific for the SARS-CoV-2 rS protein antigen(s) as detected by ELISA at Day 21 and Day 35. Derived/calculated endpoints based on these data will include geometric mean ELISA units, geometric mean fold rise, and seroconversion rate (proportion of subjects with ≥4-fold rises in ELISA units).

The secondary endpoints are:

- Unsolicited AEs (eg, treatment-emergent, serious, suspected unexpected serious, those of special interest, all MAAEs) through the first 49 days by MedDRA classification, severity score, and relatedness.
- Vital sign measurements with severity scoring immediately following vaccination. Descriptive statistics (mean, SD, change from baseline) by treatment group, by visit.
- All MAAEs through 105 days after first vaccination; and any MAAE related to vaccine (as determined by the investigator), AESI (predefined and including COVID-19 positivity), or SAE through 365 days after final vaccination.
- Serum IgG antibody levels specific for the SARS-CoV-2 rS protein antigen(s) as detected by ELISA (refer to primary endpoints), including seroconversion rate (proportion of subjects with ≥2-fold and ≥4-fold rises in antibody levels) and seroresponse rate (proportion of subjects with rises in ELISA units exceeding the 95$^{th}$ percentile of placebo recipients at the same time point) to be described across time points following first and second vaccination through Day 189.

- Epitope-specific immune responses to the SARS-CoV-2 rS protein receptor binding domain measured by serum titers/concentrations of antibody in a competition ELISA assay using a monoclonal antibody to the similar SARS receptor binding domain or other inhibition assay to that ACE-2 receptor. Derived/calculated endpoints based on these data will include geometric mean titer or concentration, geometric mean fold rise, seroconversion rate, and seroresponse rate.

- Neutralizing antibody activity compared to baseline in all treatment groups at various time points. Expression units (eg, titers) to be determined during assay development.

- Cell-mediated (Th1/Th2) pathways as measured by whole blood (flow cytometry) and/or in vitro PBMC stimulation (eg, ELISpot, cytokine staining) with SARS-CoV-2 rS protein(s) as measured on Days 0, 7, and 28.

Exploratory endpoints are:

- Percentage of protection measured in a passive antibody live virus challenge model using SARS-CoV-2 virus or other modified construct at select time points if available and appropriate. Expression units to be determined during development.

- Additional serum Ig antibody ELISA measurements specific for the SARS-CoV-2 rS protein antigen(s) characterization. Expression units to be determined during development.

- Any additional assays to measure immune response, protection, or potential safety signals.

### 3. STUDY DESIGN

Part 1 is a first-in-human, Phase 1, randomized, placebo-controlled, observer-blinded study to evaluate the safety and immunogenicity of a SARS-CoV-2 recombinant spike protein nanoparticle vaccine (SARS-CoV-2 rS) with or without Matrix-M adjuvant in male and female subjects. Subjects will be healthy adults based on medical history, physical examination, and baseline clinical laboratory testing and serology tests.

After signing the ICF, subjects may be screened within a window of up to 30 days. Subjects will be asked to provide consent for the use of samples for future testing for other viruses and/or sequencing of the SARS-CoV-2 in positive specimens or assay development specific to SARS-CoV-2 (or related variants). The study design for Part 1 allows up to 2 unique SARS-CoV-2 rS constructs to be investigated.

A minimum of 100 healthy male and female subjects between 18 and 59 years of age (inclusive) per SARS-CoV-2 rS construct, along with 25 control subjects receiving only placebo, are planned for block randomization. In addition, 6 sentinel subjects per unique construct will be enrolled in an unblinded fashion (ie, in addition to those block randomized). The total number of subjects in Part 1 will not exceed 131 subjects if a single cohort is included; 237 subjects if 2 cohorts are included and randomized in parallel; or 262 subjects if 2 constructs are included and randomized in sequence. Subjects who meet the criteria for study entry will be assigned to the current cohort and randomly assigned to a treatment group (Table 3-1).

Sentinel dosing will be utilized for the first vaccination, where the first 6 subjects enrolled in each cohort (ie, 6 sentinel subjects per unique SARS-CoV-2 rS construct investigated) are vaccinated and observed for reactogenicity through 48 hours before the remaining subjects are vaccinated. Sentinel subjects in Cohort 1 will be assigned in a 1:1 ratio to receive either 5 or 25 μg SARS-CoV-2 rS with Matrix-M adjuvant. Treatment assignment for the sentinel subjects will be unblinded.

Optional: Cohort 2 may be enrolled to investigate a unique second SARS-CoV-2 rS nanoparticle construct. If a unique second construct is not available, then the construct used in Cohort 1 may be investigated at lower doses in Cohort 2 (ie, lower doses of either SARS-CoV-2 rS or Matrix-M) based on early immune responses observed in Cohort 1. No more than the proposed total number of subjects will be allowed in Cohort 2; however, fewer treatment groups may be evaluated. For Cohort 2, sentinel subjects will be assigned in a 1:1 ratio to receive same treatment as Group G or Group H (Table 3-1).

**Table 3-1 Treatment Groups (Part 1)**

| Cohort | Treatment Group | n | | Day 0 | | | Day 21 (+ 5 days) | | |
|---|---|---|---|---|---|---|---|---|---|
| | | Randomized | Sentinel | SARS-CoV-2 rS (µg) | | Matrix-M (µg) | SARS-CoV-2 rS (µg) | | Matrix-M (µg) |
| 1 | A | 25 | – | Placebo | 0 | 0 | Placebo | 0 | 0 |
| | B | 25 | – | Construct A | 25 | 0 | Construct A | 25 | 0 |
| | C | 25 | 3 | Construct A | 5 | 50 | Construct A | 5 | 50 |
| | D | 25 | 3 | Construct A | 25 | 50 | Construct A | 25 | 50 |
| | E | 25 | – | Construct A | 25 | 50 | Placebo | 0 | 0 |
| Maximum number of subjects if 1 construct evaluated: | | 125 | 6 | Total = 131 | | | | | |
| 2[a,b] | A | 0[c] or 25 | – | Placebo | 0 | 0 | Placebo | 0 | 0 |
| | F | 25 | – | Construct B | X[d] | 0 | Construct B | X[d] | 0 |
| | G | 25 | 3 | Construct B | Y[d] | 50 | Construct B | Y[d] | 50 |
| | H | 25 | 3 | Construct B | X[d] | 50 | Construct B | X[d] | 50 |
| | I | 25 | – | Construct B | X[d] | 50 | Placebo | 0 | 0 |
| Maximum number of subjects if 2 constructs evaluated in sequence: | | 250 | 12 | Total = 262 | | | | | |

Note: At least 1 and up to two SARS-CoV-2 rS constructs (ie, Constructs A and B) will be evaluated during Part 1.

e If study vaccine for Cohort 2 (ie, Construct B) is not available at the same time as study vaccine for Cohort 1 (ie, Construct A), then Cohort 2 may be sequentially enrolled. Fewer treatment groups may be evaluated in such a case, based on early immune and safety results in Cohort 1.

f If a unique second construct is not available, then the construct used in Cohort 1 may be investigated at lower doses in Cohort 2 (ie, lower doses of either SARS-CoV-2 rS or Matrix-M) based on early immune responses observed in Cohort 1. No more than the proposed total number of subjects will be allowed in Cohort 2; however, fewer treatment groups may be evaluated.

g If Cohort 2 is enrolled in parallel with Cohort 1, no subjects will be randomized to Cohort 2, Group A.

h X and Y indicate 2 different doses, where X is the higher dose. Doses will be determined based on early immune responses observed in Cohort 1 or as new preclinical data becomes available.

Study vaccinations will comprise 2 IM injections at a 21-day interval (Day 0 and Day 21), ideally in alternate deltoids with the study treatment assigned (ie, placebo or a 5 or 25 µg dose of SARS-CoV-2 rS (Construct A or B), with or without a 50 µg dose of Matrix-M adjuvant), in an injection volume of approximately 0.5 mL (not to exceed 0.7 mL). Vaccinations will be performed on an outpatient basis by site personnel using a methodology to maintain the blind (eg, unblinded personnel, blinded syringe).

Blood samples for immunogenicity assessments (B and T cell) will be collected before each vaccination and at selected time points following first and second vaccination.

Safety assessments will include monitoring and recording of solicited (local and systemic reactogenicity events) and unsolicited AEs; MAAEs; AESI; SAEs; clinical laboratory results including hematology and serum chemistry; vital sign measurements; and physical examination findings. Relatedness/causality and severity grading will be included.

Vaccination pause rules based on reactogenicity, safety laboratory results, and SAEs related to study participation will be in place to monitor subject safety during the study. A safety monitoring committee will be formed before the first subject is vaccinated and will follow a SMC charter. The SMC will review study progress and subject, clinical, safety, and reactogenicity data ad hoc when a vaccination pause rule is met, for immediate concerns regarding observations during this study, to allow advancement to Part 2 of study, or as needed.

Part 1 will consist of a screening period (Days −30 to 0); vaccination days (Days 0 and 21); outpatient study visits on Day 0, Day 2 (sentinels only), Days 7 (+ 3 days), 21 (+ 5 days), 28 (+3 days), 35 (+ 5 days), 49 (+ 5 days), 105 (± 7 days), and 189 (± 15 days); and an end-of-study telephone call on Day 386 (± 15 days). The duration of the Part 1, excluding screening, is approximately 387 days.

If the Novavax study vaccine or another vaccine from a different manufacturer is demonstrated to be safe and efficacious and approved and/or authorized for use by the regulatory authority in Australia, subjects for whom the new approved/authorized vaccine is recommended and available will be counseled with respect to their options. These subjects may be offered the opportunity to be unblinded so that those who received placebo may be offered the Novavax vaccine or another approved/authorized vaccine, as appropriate, outside the protocol procedures. Subjects who received the Novavax vaccine and who wish to receive an approved/authorized vaccine from another manufacturer will be advised to discuss this plan with their healthcare provider given the current lack of safety data regarding the sequential administration of vaccines made by different manufacturers. Subjects who are unblinded and receive an approved/authorized vaccine in this manner will be strongly encouraged to remain in study for safety follow-up as defined in the protocol. However, subjects also have the right to discontinue participation in the study at any time.

Due to the ongoing pandemic, recent national regulatory and local IRB and public health guidance will be applied at any site location requiring alternatives for a study subject’s ability to attend an investigational site for protocol-specified visits. While the site should do everything in its power to keep activities as per the defined schedule of events, the site should also have a process in place to conduct the needed assessments (eg, telephone contact, virtual visit, alternative location for assessment, including local laboratories or imaging centers) when necessary and feasible, as long as such visits are sufficient to assure the safety of study subjects.

#### 3.1 SCHEDULE OF EVENTS

**Table 3-2 Schedule of Events (Part 1)**

| Study Day: | –30 to 0 | 0 | 2[a] | 7 | 21 | 28 | 35 | 49 | 105 | Unscheduled | 189 | 386[b] |
|---|---|---|---|---|---|---|---|---|---|---|---|---|
| **Window (days)[c]:** | | **0** | **0** | **+3** | **+5** | **+3** | **+5** | **+5** | **±7** | **–** | **±15** | **±15** |
| **Study Visit:** | **Screening** | **1** | **S1[a]** | **2** | **3** | **4** | **5** | **6** | **7** | **Unscheduled** | **8** | **EOS[b]** |
| Informed consent | X | | | | | | | | | | | |
| Medical history[d] | X | | | | | | | | | | | |
| Inclusion/exclusion criteria | X | X[e,f] | | | X[e,f] | | | | | | | |
| Demographics[g] | X | | | | | | | | | | | |
| Prior/concomitant medications[h,i] | X | X[e,f] | X | X | X[e,f] | X | X | X | X | X | X | X |
| Vital sign measurements | X | XX[j] | X | X | XX[j] | X | X | X | X | X | X | |
| Urine pregnancy test[k] | X | X[f] | | | X[f] | | | | | | X | |
| Serum FSH[l] | X | | | | | | | | | | | |
| Serology[m] | X | | | | | | | | | | | |
| Testing for SARS-CoV-2 (ELISA)[n] | X | | | | | | | | | X | | |
| Urine drug screen[o] | X | | | | | | | | | | | |
| Physical examination[p] | X | X[f] | X | X | X[f] | X | X | X | X | X | | |
| Clinical laboratory testing[q] | X | | | X | X[f] | X | | | | X (if applicable) | | |
| Vaccination | | X | | | X | | | | | | | |
| Immediate reactogenicity[r] | | X | | | X | | | | | | | |
| Subject diary distribution[s] | | X | | | X | | | | | | | |
| Subject diary review and collection[s] | | | (review) | X | | X | | | | (review, if applicable) | | |
| Nasal swab(s) - nasopharyngeal and/or mid-turbinate; testing for SARS-CoV-2 (PCR)[t] | X | | | | | | X | | | X | | |
| Blood sampling for SARS-CoV-2 immunogenicity (ELISA) | | X[f] | | X | X[f] | X | X | X | X | | X | |
| Blood sampling for SARS-CoV-2 neutralization assay and (potential) challenge study | | X[f] | | | X[f] | | X | X | | | X | |
| Whole blood sampling and PBMC harvest for cell-mediated functional testing | | X[f] | | X | | X | | | | | | |
| Unsolicited AEs | | X | X | X | X | X | X | X | | | | |
| All MAAEs | | X | X | X | X | X | X | X | X | | | |
| Any MAAE attributed to vaccine | | X | X | X | X | X | X | X | X | X | X | X |
| SAEs | | X | X | X | X | X | X | X | X | X | X | X |
| AESI[u] | | X | X | X | X | X | X | X | X | X | X | X |

| Study Day: | –30 to 0 | 0 | 2[a] | 7 | 21 | 28 | 35 | 49 | 105 | Unscheduled | 189 | 386[b] |
|---|---|---|---|---|---|---|---|---|---|---|---|---|
| Window (days)[c]: | | 0 | 0 | +3 | +5 | +3 | +5 | +5 | ±7 | – | ±15 | ±15 |
| Study Visit: | Screening | 1 | S1[a] | 2 | 3 | 4 | 5 | 6 | 7 | Unscheduled | 8 | EOS[b] |
| EOS form[v] | | | | | | | | | | | | X |

[a] Sentinel subjects (only) will return to the clinic for Study Visit S1 at 48 (±8) hours after vaccination (ie, Day 2).

[b] End-of-study telephone call. Should subjects decide to terminate early, a telephone call will occur to collect the maximum safety data possible. At the EOS visit, it should be confirmed that no pregnancy has occurred in women of reproductive potential since the last contact.

[c] Days relative to vaccination are only estimates because the window allowance is not inclusive. Should a study pause occur, visits/windows will be adjusted to allow subjects to continue without protocol deviation. Visit schedule following the second vaccination is calculated relative to the day the second vaccination was received.

[d] Including prior and concomitant medical conditions, recent vaccinations (≤90 days), and significant surgical procedures.

[e] Specific exclusions to vaccination will be assessed (Section 4.3). Should subjects start specific medications or have specific diagnoses that are exclusionary at baseline, approval for vaccination must be given by medical monitor or sponsor.

[f] Performed prior to vaccination.

[g] Screening only. Including date of birth (day, month, and year), sex, race, ethnicity, weight, height, and BMI (derived).

[h] Recent and current medications at the time of screening to be reviewed to ensure eligibility criteria are fulfilled. Concomitant medications include all medications (including vaccines) taken by the subject from the time of first vaccination through Study Visit 6.

[i] For Study Visits 7 and 8 and EOS visit, only those medications associated with any MAAE attributed to vaccine, AESI, or SAE will be recorded.

[j] On vaccination days, vital sign measurements will be collected once before vaccination and again at 30 (+15) minutes after vaccination. FDA toxicity scoring will be applied for pulse and blood pressure (systolic and diastolic). FDA toxicity scoring for temperature will be applied during reactogenicity evaluation both post vaccination (clinic assessment) and daily (by subject) through 7 days following vaccination

[k] Women of childbearing potential only. A urine pregnancy test will be performed at screening and prior to each vaccination. A serum pregnancy test may be used at screening or at the discretion of the investigator. A positive urine pregnancy test at any time will result in the subject not receiving any further vaccination.

[l] Females only. A serum FSH test may be performed at screening to confirm postmenopausal status.

[m] Serology testing will include hepatitis B, hepatitis C, and HIV.

[n] All randomized subjects must have a documented negative test for SARS-CoV-2 by ELISA (IgG) based on screening labs. Testing may be repeated during the screening period if exposure to SARS-CoV-2 is suspected, at the discretion of the investigator.

[o] Urine drug screen will occur at screening. If a subject is on controlled substances at the discretion of their primary care physician for non-pain related diagnoses (eg, Adderall) and the medication dosage is stable, then the subject may be enrolled if all other inclusion/exclusion criteria are met. At the discretion of the investigator, an additional urine drug screen may be performed on any subject at any time. A positive tetrahydrocannabinol result will not be exclusionary but may result in a decision by the investigator to exclude subjects due to concerns of impairment in the quality of safety reporting.

[p] Examination at screening to include height and weight (calculated BMI), HEENT, lungs, heart, and abdomen as well as the lymphatic assessment of upper extremities to allow for vaccination. A targeted or symptom-directed physical examination will be performed at all other scheduled time points but always to include lymphatic assessment of injected upper extremity on vaccination days. Interim physical examinations will be performed at any unscheduled visit at the discretion of the investigator, if necessary.

| Study Day: | –30 to 0 | 0 | 2[a] | 7 | 21 | 28 | 35 | 49 | 105 | Unscheduled | 189 | 386[b] |
|---|---|---|---|---|---|---|---|---|---|---|---|---|
| Window (days)[c]: | | 0 | 0 | +3 | +5 | +3 | +5 | +5 | ±7 | – | ±15 | ±15 |
| Study Visit: | Screening | 1 | S1[a] | 2 | 3 | 4 | 5 | 6 | 7 | Unscheduled | 8 | EOS[b] |

[q] Blood samples will be collected under non-fasting conditions and will be prepared using standard procedures. A complete list of assessments is provided in Section 6.1.2. Laboratory testing may be repeated once during the 30-day screening period if specific values used for exclusion criteria exceed toxicity grade 1 (Section 6.1.2). Should any value reach FDA toxicity grade 3 or greater during the study, it must be recorded as an AE and followed with retesting and observing for resolution (Grade ≤1) or new clinical baseline. Laboratory toxicity grades ≥3 must be repeated within 7 days and may require an unscheduled visit for follow-up. Laboratory tests may be included in unscheduled visits if deemed necessary by the investigator.

[r] On vaccination days, reactogenicity will be collected at 30 (+15) minutes after vaccination.

[s] All subjects will record reactogenicity (local and systemic) in a daily subject diary starting on the same day of the vaccinations and for an additional 7 days (not counting vaccination day). Site personnel will review the information from the subject diary with the subjects on Visits 2 and 4 or if an unscheduled visit occurs due to toxicity grade ≥3 prior to those visits. For sentinel subjects, site personnel will review the information from the subject diary with the subjects at 48 (±8) hours after first vaccination (ie, Day 2, Visit S1). Should any reactogenicity event extend beyond 7 days after vaccination (toxicity grade ≥1), then it will be recorded as an AE and followed to resolution per FDA guidelines for dataset capture.

[t] All randomized subjects must have a documented negative test for SARS-CoV-2 by PCR at screening. Should testing advance in time for the study to allow nasal turbinate swabs to be highly accurate for SARS-CoV-2 PCR, then only nasal turbinate swabs will be required. Either swab (or both) may be performed at screening and Day 35 depending on availability and detection performance. Either swab (or both) and PCR testing may be performed at any visit (scheduled or unscheduled) if exposure to SARS-CoV-2 is suspected, at the discretion of the investigator, including rescreening at baseline.

[u] AEs of special interest: To include potentially immune mediated medical conditions (Appendix 4, Table 16-5), COVID-19 diagnosis with sequelae (Appendix 4, Table 16-6), or any newly identified potential AESI followed through 365 days after final vaccination.

[v] EOS form will be completed for all subjects, including those who are terminated early.

### 4. STUDY POPULATION

Healthy male or female subjects will be enrolled at up to 3 sites across Australia (and potentially the United States). A minimum of 100 healthy male and female subjects between 18 and 59 years of age (inclusive) per SARS-CoV-2 rS construct, along with 25 control subjects receiving only placebo, are planned for block randomization. In addition, 6 sentinel subjects per unique construct will be enrolled in an unblinded fashion (ie, in addition to those block randomized). The total number of subjects in Part 1 will not exceed 131 subjects if a single cohort is included; 237 subjects if 2 cohorts are included and randomized in parallel; or 262 subjects if 2 constructs are included and randomized in sequence.

#### 4.1 INCLUSION CRITERIA

Each subject must meet all of the following criteria to be enrolled in this study:

1. Healthy adult males or females between 18 and 59 years of age, inclusive, at screening. Healthy status will be determined by the investigator based on medical history, clinical laboratory results, vital sign measurements, and physical examination at screening.

2. The subject has a body mass index 17 to 35 kg/m$^2$, inclusive, at screening.

3. Willing and able to give informed consent prior to study enrollment and comply with study procedures.

4. Female subjects of childbearing potential (defined as any female who has experienced menarche and who is NOT surgically sterile [ie, hysterectomy, bilateral tubal ligation, or bilateral oophorectomy] or postmenopausal [defined as amenorrhea at least 12 consecutive months or documented plasma follicle-stimulating hormone level ≥40 mIU/mL]) must agree to be heterosexually inactive from at least 21 days prior to enrollment and through 6 months after the last vaccination OR agree to consistently use any of the following methods of contraception from at least 21 days prior to enrollment and through 6 months after the last vaccination:

    a. Condoms (male or female) with spermicide (if acceptable in country)

    b. Diaphragm with spermicide

    c. Cervical cap with spermicide

    d. Intrauterine device

e. Oral or patch contraceptives

f. Norplant®, Depo-Provera®, or other in country regulatory-approved contraceptive method that is designed to protect against pregnancy

g. Abstinence, as a form of contraception, is acceptable if in line with the subject's lifestyle

**NOTE**: Periodic abstinence (eg, calendar, ovulation, symptothermal, post-ovulation methods) and withdrawal are not acceptable methods of contraception. These procedures and laboratory test results must be confirmed by physical examination, by subject recall of specific date and hospital/facility of procedure, or by medical documentation of said procedure.

#### 4.2 EXCLUSION CRITERIA

Subjects meeting any of the following criteria will be excluded from the study:

1. Any ongoing, symptomatic acute or chronic illness requiring medical or surgical care, inclusive of changes in medication in the past 2 months indicating that chronic illness/disease is not stable (at the discretion of the investigator). This includes any current workup of undiagnosed illness that could lead to a new condition.

2. Chronic disease inclusive of: a) hypertension uncontrolled for age according to JNC 8 guidelines; b) congestive heart failure by NYHA functional classification of ≥II; c) chronic obstructive pulmonary disease by GOLD classification of ≥2; d) recent (within 6 months prior to first study vaccination) exacerbation of coronary artery disease as manifested by cardiac intervention, addition of new cardiac medications for control of symptoms, or unstable angina; e) asthma (diagnosed by spirometry showing reversibility of disease and must meet at least the Step 1 classification with current prescription/use of medications to control symptoms); f) diabetes requiring use of medicine (insulin or oral) or not controlled with diet.

3. Participation in research involving an investigational product (drug/biologic/device) within 45 days prior to first study vaccination.

4. History of a confirmed diagnosis of SARS or COVID-19 disease (confirmed by a specific test for each disease) or known exposure to a SARS-CoV-2 positive confirmed close contact (eg, family member, housemate, daycare provider, aged parent requiring care), at the discretion of the investigator.

**NOTE:** If exposure is suspected, subjects may be enrolled with subsequent documentation of a negative test for SARS-CoV-2, at the discretion of the investigator.

5. Currently working in an occupation with a high risk of exposure to SARS-CoV-2 (eg, healthcare worker, emergency response personnel).

6. Currently taking any product (investigational or off-label) for prevention of COVID-19 disease.

7. Positive tests for SARS-CoV-2 (either ELISA IgG or PCR) at screening or prior to first vaccination. Testing may be repeated during the screening period if exposure to SARS-CoV-2 is suspected, at the discretion of the investigator.

8. Received influenza vaccination within 14 days prior to first study vaccination, or any other vaccine within 4 weeks prior to first study vaccination.

9. Any autoimmune or immunodeficiency disease/condition (iatrogenic or congenital).

**NOTE:** Stable endocrine disorders that have a confirmed autoimmune etiology (eg, thyroid, pancreatic), including stable diet-controlled diabetes, are allowed at the discretion of the investigator.

10. Chronic administration (defined as more than 14 continuous days) of immunosuppressant, systemic glucocorticosteroids, or other immune-modifying drugs within 90 days prior to first study vaccination; or anticipation of the need for immunosuppressive treatment within 6 months after last vaccination.

**NOTE:** An immunosuppressant dose of glucocorticoid is defined as a systemic dose ≥10 mg of prednisone per day or equivalent. The use of topical and nasal glucocorticoids will be permitted. Use of inhaled glucocorticoids is prohibited.

11. Received immunoglobulin, blood-derived products, or other immunosuppressant drugs within 90 days prior to first study vaccination.

12. Any acute illness concurrent or within 14 days prior to first study vaccination (medical history and/or physical examination) or documented temperature of >38°C during this period. This includes respiratory or constitutional symptoms consistent with SARS-CoV-2 (COVID-19) exposure (ie, cough, sore throat, difficulty breathing)

**NOTE:** This is a temporary exclusion for which the subject may be re-evaluated if they remain free from acute illness after 14 days and are test-negative for COVID-19 if symptoms indicate exposure. Subjects may be re-evaluated anytime during the screening period.

13. Known disturbance of coagulation (iatrogenic or congenital).

**NOTE:** The use of ≤325 mg of aspirin per day as prophylaxis is permitted, but the use of other platelet aggregation inhibitors, thrombin inhibitors, Factor Xa inhibitors, or warfarin derivatives is exclusionary, regardless of bleeding history, because these imply treatment or prophylaxis of known cardiac or vascular disease.

14. Evidence of Hepatitis B or C or HIV by laboratory testing.

15. A positive test result for drugs of abuse (except a positive test result associated with prescription medication that has been reviewed and approved by the investigator) at screening.

**NOTE:** A positive tetrahydrocannabinol result will not be exclusionary but may result in a decision by the investigator to exclude subjects due to concerns of impairment in the quality of safety reporting.

16. Any neurological disease or history of significant neurological disorder (eg, meningitis, seizures, multiple sclerosis, vasculitis, migraines, Guillain-Barré syndrome [genetic/congenital or acquired]).

**NOTE:** Significant neurological migraine includes frequent migraine (2 times a month or greater), migraine with aura or migraine with complications (status migrainosus, persist aura without infarction, infarction or aura triggered seizure defined by International Classification of Headache Disorders-3 [ICHD-3 2018]).

17. Active cancer (malignancy) within 5 years prior to first study vaccination (with the exception of adequately treated non-melanomatous skin carcinoma, at the discretion of the investigator).

18. Vital sign (blood pressure, pulse, temperature) abnormalities of toxicity grade >1 (Appendix 3; Table 16-4).

**NOTE:** Vital sign measurements may be repeated once during the screening period to allow inclusion with the most recent measurement taken as the baseline value.

19. Clinical laboratory abnormalities of toxicity grade >1 for selected serum chemistry and hematology parameters (Appendix 3; Table 16-3).

**NOTE:** Clinical laboratory testing may be repeated once during the screening period to allow inclusion with the most recent measurement taken as the baseline value.

20. Any known allergies to products contained in the investigational product or latex allergy.

21. Women who are pregnant, breastfeeding, or who plan to become pregnant during the study.

22. History of alcohol abuse or drug addiction within one year prior to the first study vaccination.

23. Any condition that, in the opinion of the investigator, would pose a health risk to the subject if enrolled or could interfere with evaluation of the study vaccine or interpretation of study results (including neurologic or psychiatric conditions deemed likely to impair the quality of safety reporting).

24. Study team member or first-degree relative of any study team member (inclusive of sponsor, PPD, and site personnel involved in the study).

#### 4.3 OTHER CONSIDERATIONS

Subjects meeting any of the following criteria may be delayed for second vaccination:

- Respiratory symptoms in the past 3 days (ie, temperature of >38°C, cough, sore throat, difficulty breathing). Subject may be vaccinated once all symptoms have been resolved for >3 days. Out of window vaccination is allowed for this reason.

- Any acute illness (eg, gastroenteritis, migraine, urinary tract infection, injury) that is causing symptoms that could, in the opinion of the investigator, impact the assessment of reactogenicity or safety laboratory tests. Subject may be vaccinated once symptoms have resolved or are stabilized for >3 days. Out of window vaccination is allowed for this reason.

- Immunization with any vaccine within 14 days prior to second vaccination. Out of window vaccination is allowed for this reason.

#### 4.4 WITHDRAWAL OF SUBJECTS FROM THE STUDY

##### 4.4.1 Reasons for Withdrawal

Subjects can withdraw consent and discontinue from the study at any time, for any reason. Subjects may refuse further procedures (including vaccination) but are encouraged to remain in the study for safety follow-up. In such cases where only safety is being conducted, subject contact could be managed via telemedicine contact (eg, telephone, web chat, video, FaceTime).

The investigator may **withhold** further vaccination from a subject in the study if the subject:

1. Is non-compliant with the protocol;

2. Experiences an SAE or intolerable AE(s) for which vaccination is not advised by the investigator;

3. Has laboratory safety assessments that reveal clinically significant hematological or biochemical changes and is deemed to be best for the subject's health;

4. Pregnancy (discontinuation of further vaccination **required**).

The investigator can also withdraw a subject upon the request of the sponsor or if the sponsor terminates the study. Upon the occurrence of an SAE or intolerable AE, the investigator may confer with the sponsor before future vaccination.

##### 4.4.2 Handling of Withdrawals

Subjects are free to withdraw from the study at any time upon request. Subject participation in the study may be stopped at any time at the discretion of the investigator or at the request of the sponsor.

When a subject withdraws from the study, the reason(s) for withdrawal shall be recorded by the investigator on the relevant page of the eCRF. Whenever possible, any subject who withdraws from the study prematurely will undergo all EOS assessments. Any subject who fails to return for final assessments will be contacted by the site in an attempt to have them comply with the protocol. The status of subjects who fail to complete final assessments will be documented in the eCRF.

##### 4.4.3 Replacements

Subjects who withdraw, are withdrawn or terminated from this study, or are lost to follow-up after signing the ICF but prior to first study vaccination may be replaced. Subjects who receive study vaccine and subsequently withdraw, are discontinued from further vaccination, are terminated from the study, or are lost to follow-up, will not be replaced.

### 5. STUDY TREATMENTS

#### 5.1 TREATMENTS ADMINISTERED

Study vaccinations will comprise 2 IM injections at a 21-day interval (Day 0 and Day 21), ideally in alternate deltoids with the study treatment assigned (ie, placebo or a 5 or 25 µg dose of SARS-CoV-2 rS [Construct A or B], with or without a 50 µg dose of Matrix-M adjuvant), in an injection volume of approximately 0.5 mL (not to exceed 0.7 mL).

#### 5.2 INVESTIGATIONAL PRODUCTS

The following supplies will be used for vaccination in the study:

| Product | Supplied Formulation |
|---|---|
| SARS-CoV-2 recombinant spike protein nanoparticle vaccine, construct BV2373[a] | Solution for preparation for injection, 70 µg/mL |
| SARS-CoV-2 recombinant spike protein nanoparticle vaccine, construct TBD[b] | Solution for preparation for injection, 70 µg/mL |
| Matrix-M adjuvant | Solution for preparation for injection, 375 µg/mL |
| Sodium chloride injection (BP, sterile) | Solution for injection, 0.9% |

a Referred to as "Construct A" in protocol

b To be determined; referred to as "Construct B" in protocol. Cohort 2 may be enrolled to investigate a unique second SARS-CoV-2 rS nanoparticle construct (refer to Section 3).

SARS-CoV-2 recombinant spike protein nanoparticle vaccine [redacted]

Further details on the study vaccine can be found in the IB (Novavax 2020).

##### 5.2.1 Investigational Product Packaging and Storage

Novavax, Inc. will provide adequate quantities and appropriate labelling of SARS-CoV-2 rS and Matrix-M adjuvant and PPD will ensure distribution to the clinical sites from a designated depot. Sodium chloride injection (BP, sterile) is commercially available and will be supplied by PPD. The clinical unit pharmacy will prepare the study treatments for each subject. Detailed instructions for the preparation of study vaccine will be provided in a separate pharmacy manual.

All investigational product must be stored according to the labeled instructions in a secure cabinet or room with access restricted to necessary clinic personnel. The site will be required to keep a temperature log to establish a record of compliance with storage conditions.

##### 5.2.2 Investigational Product Accountability

The investigator (or delegate) will maintain accurate records of receipt of all investigational product, including dates of receipt. Accurate records will be kept regarding when and how much investigational product is dispensed and used for each subject in the study. Reasons for departure from the expected dispensing regimen must also be recorded. At the completion of the study, and to satisfy regulatory requirements regarding investigational product accountability, all investigational product will be reconciled and retained or destroyed according to applicable regulations. No investigational product will be destroyed until authorized in writing by the sponsor.

#### 5.3 METHOD OF ASSIGNING SUBJECTS TO TREATMENT GROUPS

Subjects will be randomly assigned in a blinded manner using the centralized IRT, with 25 subjects assigned to each treatment group, according to pre-generated randomization schedules. There will be no stratification. Details regarding the IRT process will be provided separately to the sites. The 6 sentinel subjects in each cohort will be in addition to the assigned blocked randomization of 25 subjects per treatment group and will be open-label with 3 subjects receiving 5 µg SARS-CoV-2 rS + 50 µg Matrix-M and 3 subjects receiving 25 µg SARS-CoV-2 rS + 50 µg Matrix-M (Table 3-1).

##### 5.3.1 Blinding Procedures

This is an observer-blinded study. To maintain the blind, placebo vaccination via IM route will be included, and unblinded site personnel will manage vaccine logistic, preparation, and administration (if necessary) so as to maintain the blind from the remainder of the site personnel and subjects. The unblinded site personnel will not be involved in study-related assessments or have subject contact for data collection following study vaccine administration.

##### 5.3.2 Breaking the Blind

A subject's treatment assignment will not be broken until the end of the study for the clinical site study team unless medical treatment of the subject depends on knowing the study treatment the subject received. A subject's treatment assignment may also be broken in the event that the Novavax study vaccine, or another vaccine from a different manufacturer, is demonstrated to be safe and efficacious and approved/authorized by the regulatory authority in Australia and the subject plans on receiving the approved/authorized vaccine. Subjects would be unblinded at EOS after all assessments are completed.

In the event that the blind needs to be broken because of a medical emergency, the investigator may unblind an individual subject's treatment allocation. As soon as possible, the investigator should first contact the medical monitor to discuss the medical emergency and the reason for revealing the actual treatment received by that subject. The treatment assignment will be unblinded through IRT. Reasons for treatment unblinding must be clearly explained and justified in the eCRF. The date on which the code was broken together with the identity of the person responsible must also be documented.

In addition to the aforementioned situations where the blind may be broken, the data will also be unblinded to a statistical team at specified time points for interim analyses, as outlined in Section 7.5.

#### 5.4 TREATMENT COMPLIANCE

All doses of the study vaccine will be administered in the clinical unit under direct observation of clinic personnel and recorded in the eCRF. Clinic personnel will confirm that the subject has received the entire dose.

The location (right or left arm), date, and timing of all doses of study vaccine will be recorded in the subjects' eCRF. If a subject does not receive a dose of the study vaccine or does not receive all of the planned dose of the study vaccine, the reason will be recorded.

##### 5.4.1 Prior Vaccinations and Concomitant Medications

Administration of medications, therapies, or vaccines will be recorded in the eCRF. Concomitant medications will include all medications (including vaccines) taken by the subject from the time of signing the ICF through EOS (or through the early termination visit if prior to that time). Prescription and over-the-counter drugs, as well as herbals, vitamins, and supplements, will be included.

### 6. STUDY PROCEDURES

Before performing any study procedures, all potential subjects will sign an ICF as outlined in Section 16.2.2.3. Subjects will undergo study procedures at the time points specified in the SOE (Table 3-2).

The total amount of blood collected from each subject over the duration of the study, including any extra assessments that may be required, will not exceed 500 mL.

#### 6.1 SAFETY ASSESSMENTS

The timing and frequency of all safety assessments are listed in the SOE (Table 3-2).

Safety assessments will include monitoring and recording of solicited (local and systemic reactogenicity events) and unsolicited AEs; MAAEs; AESI; SAEs; clinical laboratory results, hematology and serum chemistry; vital sign measurements; and physical examination findings.

Subjects who are unblinded for the purpose of receiving an approved/authorized vaccine, will be encouraged to remain in the study for safety follow-up as defined in the protocol, and safety assessments will be performed via the timelines and mechanisms as described above and throughout Section 6.1.

In addition, investigators will be required to report any AEs observed in subjects who received another manufacturer's approved/authorized vaccine to health care and/or regulatory authorities via the local Regulatory reporting guidance.

##### 6.1.1 Adverse Events

Adverse events will be assessed during the study as described in the SOE (Table 3-2) and should be followed until they are resolved, stable, or judged by the investigator to be not clinically significant. Adverse events will be captured after first dose of study vaccination administered with the exception of an AE related to study procedure or one that causes a delay in study vaccination administration (eg, acute illness).

The investigator is responsible for ensuring that all AEs and SAEs are recorded in the eCRF and reported to the sponsor, regardless of their relationship to study vaccine or clinical significance. If there is any doubt as to whether a clinical observation is an AE, the event should be reported.

###### 6.1.1.1 Adverse Event Definitions

The investigator is responsible for reporting all AEs that are observed or reported during the study, regardless of their relationship to study vaccination or their clinical significance.

An AE is defined as any untoward medical occurrence in a subject enrolled into this study regardless of its causal relationship to study vaccination. Subjects will be instructed to contact the investigator at any time after randomization if any symptoms develop.

###### 6.1.1.1.1 Serious Adverse Events

An SAE is defined as any event that

- results in death
- is immediately life threatening
- requires inpatient hospitalization or prolongation of existing hospitalization
- results in persistent or significant disability/incapacity
- is a congenital anomaly/birth defect

Important medical events that may not result in death, be life threatening, or require hospitalization may be considered SAEs when, based upon appropriate medical judgment, they may jeopardize the subject or may require medical or surgical intervention to prevent one of the outcomes listed in this definition. Examples of such medical events include allergic bronchospasm requiring intensive treatment in an emergency room or at home, blood dyscrasias or convulsions that do not result in inpatient hospitalization, or the development of drug dependency or drug abuse.

###### 6.1.1.1.2 Local and General Systemic Reactogenicity

Site-specific local (arm) and general systemic reactogenicity reactions including start and stop dates will be recorded and the investigator will apply a standard toxicology grading at the subsequent visit based on subject diary results (Table 16-1). If no reactogenicity is noted, then a zero (0) should be recorded for the day and parameter. If a subject does not complete a diary for any measurement, a missing value for that day and parameter will be noted. Subjects will be instructed that should they have reactogenicity with a potential toxicity grade 3 or greater at any time, they are to call the site for further evaluation, including a potential unscheduled outpatient visit. Should any reactogenicity event extend beyond 7 days after vaccination (ie, toxicity grade ≥1), then it will be recorded as an AE with the same start date as the reactogenicity event and followed to resolution.

###### 6.1.1.1.3 Adverse Events of Special Interest

Subjects will be assessed for diagnosis of an AESI at all study visits. Adverse events of special interest include potential immune-mediated medical conditions (PIMMC), AEs specific to COVID-19 disease, or other potential AEs that may be determined at any time by regulatory authorities as additional information concerning COVID-19 is obtained. Given the concern for cytokine storm, an AESI of cytokine release syndrome will be included as an AEs specific to COVID-19 disease. Listings of AESI are presented in Appendix 4 (Section 16.4).

###### 6.1.1.1.4 Medically Attended Adverse Events

A MAAE is defined as an AE that leads to an unscheduled visit to a healthcare practitioner.

###### 6.1.1.1.5 Pregnancy

Pregnancy is not considered an AE unless there is a suspicion that an investigational vaccine may have interfered with the effectiveness of a contraceptive medication. Any pregnancy that occurs during study participation must be reported using a clinical study pregnancy form. To ensure subject safety, each pregnancy must be reported to Novavax, Inc. within 2 weeks of learning of its occurrence. If pregnancy occurs, further vaccination will be discontinued. The pregnancy must be followed up to determine outcome (including spontaneous miscarriage, elective termination, normal birth, or congenital abnormality) and the status of both mother and child, even if the subject was discontinued from the study. Pregnancy complications and elective terminations for medical reasons must be reported as an AE or SAE. Spontaneous miscarriages must be reported as an SAE.

Any pregnancy brought to the investigator's attention after the subject has completed the study but occurring while the subject was in the study must be promptly reported to:

Sponsor Safety Monitor: [redacted]

###### 6.1.1.2 Eliciting and Documenting Adverse Events

At every study visit, subjects will be asked a standard question to elicit any medically-related changes in their well-being. They will also be asked if they have been hospitalized, had any accidents, used any new medications, or changed concomitant medication regimens (both prescription and over-the-counter medications).

In addition to subject observations, AEs will be documented from any data collected on the AE page of the eCRF or other documents that are relevant to subject safety.

###### 6.1.1.3 Reporting Adverse Events

All AEs reported or observed during the study will be recorded on the AE page of the eCRF. Information to be collected includes study treatment, type of event, time of onset, dosage, investigator-specified assessment of severity and relationship to study vaccine and/or study procedure, time of resolution of the event, seriousness, any required treatment or evaluations, and outcome. Any AEs resulting from concurrent illnesses, reactions to concurrent illnesses, reactions to concurrent medications, or progression of disease must also be reported. All AEs will be followed until they are resolved, stable, or judged by the investigator to be not clinically significant. MedDRA will be used to code all AEs.

Any medical condition that is present at the time that the subject is screened but does not deteriorate should not be reported as an AE. However, if it deteriorates at any time during the study, it should be recorded as an AE.

Due to the vaccination pause rules in this study, data entry of reactogenicity from either investigator-observed or subject-reported outcomes, any SAE, and all AEs attributed to vaccine through Day 49 should be entered into the database within 24 hours of documentation by site personnel.

Any AE that is considered serious by the investigator or that meets SAE criteria (Section 6.1.1.1) must be reported to the sponsor immediately (within 24 hours after the investigator has confirmed the occurrence of the SAE). The investigator will assess whether there is a reasonable possibility that the study vaccine caused the SAE. The sponsor will be responsible for notifying the relevant regulatory authorities of any SAE, in compliance with health authority requirements, as outlined in the relevant clinical trial guidelines. The investigator is responsible for notifying the independent HREC/IRB directly.

SAE reports received that may be attributed to a combination of the Novavax vaccine and an approved/authorized vaccine from a different manufacturer will be reported to the regulatory authority in Australia as applicable.

SAE reporting forms allow for the notation of other factors that may have impacted the investigator's assessment of causality. Investigators will be instructed to utilize this section of the reporting form to note the impact of an approved/authorized vaccine from a different manufacturer on the event, if applicable. Investigators will be required to report any SAEs in subjects who received a different manufacturer's approved/authorized vaccine to local health care and/or regulatory authorities in Australia as per the local regulatory guidelines.

For this study, the following contact information will be used for SAE reporting:

PPD Medical Monitor: [redacted]

###### 6.1.1.4 Assessment of Severity

The severity (or intensity) of an AE refers to the extent to which it affects the subject's daily activities and will be classified as mild, moderate, or severe using the following criteria:

- Mild (grade 1): These events require minimal or no treatment and do not interfere with the subject's daily activities.

- Moderate (grade 2): These events result in a low level of inconvenience or require minor therapeutic measures. Moderate events may cause some interference with normal functioning.

- Severe (grade 3): These events interrupt a subject's usual daily activity and may require systemic drug therapy or other treatment. Severe events are usually incapacitating.

If the severity of an AE changes, the most intense severity should be reported. An AE characterized as intermittent does not require documentation of the onset and duration of each episode.

###### 6.1.1.5 Assessment of Causality

The investigator's assessment of an AE's relationship to study vaccine is part of the documentation process, but it is not a factor in determining what is or is not reported in the study. If there is any doubt as to whether a clinical observation is an AE, the event should be reported.

The investigator will assess causality (ie, whether there is a reasonable possibility that the study vaccine caused the event) for all AEs and SAEs (solicited reactions are to be considered as being related to vaccination).

The relationship will be classified as follows:

- Not related: There is not a reasonable possibility of relationship to study vaccine. The AE does not follow a reasonable temporal sequence from administration of study vaccine or can be reasonably explained by the subject's clinical state or other factors (eg, disease under study, concurrent diseases, and concomitant medications).

- Related: There is a reasonable possibility of relationship to study vaccine. The AE follows a reasonable temporal sequence from administration of study vaccine and cannot be reasonably explained by the subject's clinical state or other factors (eg, disease under study, concurrent diseases, or concomitant medications), represents a known reaction to study vaccine or other vaccines in its class, is consistent with the known pharmacological properties of the study vaccine, and/or resolves with discontinuation of the study vaccine (and/or recurs with re-challenge, if applicable).

###### 6.1.1.6 Follow-up of Adverse Events

All AEs must be reported in detail on the appropriate page of the eCRF and followed until they are resolved, stable, or judged by the investigator to be not clinically significant.

##### 6.1.2 Clinical Laboratory Testing

Clinical laboratory tests will be performed at a designated laboratory. Blood and urine samples will be collected under non-fasting conditions and will be prepared using standard procedures.

Repeat clinical laboratory tests may be performed at the discretion of the investigator, if necessary, to evaluate inclusion and exclusion criteria or clinical laboratory abnormalities. The clinical laboratory that will perform the tests will provide the reference ranges for all clinical laboratory parameters.

The following clinical laboratory assessments will be recorded:

| | |
|---|---|
| Hematology | Hemoglobin*, hematocrit, platelet count*, and complete white blood cell count* |
| Serum Chemistry | Alanine aminotransferase*, aspartate aminotransferase*, total bilirubin*, urea (ie, BUN), creatinine*, random glucose, potassium, sodium, and total protein |
| Serology | Hepatitis B surface antigen, hepatitis C virus antibody, and human immunodeficiency virus antibody types 1 and 2 (Screening only) |
| Other analyses | All subjects: Urine drug screen (amphetamines, methamphetamines, methadone, barbiturates, benzodiazepines, cocaine, opiates, methylenedioxymethamphetamine, phencyclidine, tetrahydrocannabinol) |
| | Female subjects: Urine pregnancy test (human chorionic gonadotropin) |

Notes: Hematology or serum chemistry parameters will be graded according to the FDA toxicity grading scale (Appendix 3, Table 16-3) or DAIDS toxicity grading and will be adjusted according to local laboratory reference ranges. FDA toxicity grades >1 as listed in Table 16-3 (Appendix 3), AND after local laboratory grading adjustments are made, are exclusionary. Subjects have the ability to rescreen once (Section 4.2). A serum pregnancy test may be substituted for a urine pregnancy test at screening or at the discretion of the investigator.

* Selected clinical laboratory abnormalities to be included in vaccination pause determination (Section 6.1.5).

Any abnormal laboratory test results (hematology or serum chemistry) resulting in a grade 3 or greater FDA toxicity score which is associated with the scheduled follow-up testing must be recorded as an AE and followed with retesting and observing for resolution (grade ≤1) or until a new baseline is established. If an underlying disease or new condition is identified that accounts for the elevation, this disease or condition should be documented as the AE.

Any clinically significant safety assessments that are associated with the underlying or new onset condition for which laboratory testing occurs during the study will not be recorded as an AE per se, rather the new onset or worsening condition should be recorded as judged by the investigator.

##### 6.1.3 Vital Sign Measurements

Vital signs, including oral temperature, pulse rate and diastolic and systolic blood pressure (after subject is seated for at least 5 minutes) will be graded according to the FDA toxicity grading scale (Appendix 3, Table 16-4). Oral temperature will be recorded and graded during general systemic reactogenicity evaluation (Section 6.1.1.1.1).

##### 6.1.4 Physical Examinations

Examination at screening to include height and weight (calculated BMI), HEENT, lungs, heart, and abdomen as well as the lymphatic assessment of upper extremities to allow for vaccination.

A targeted or symptom-directed physical examination will be performed at the time points specified in the SOE (Table 3-2) but always to include lymphatic assessment of injected upper extremity on vaccination days. Interim physical examinations will be performed at any unscheduled visit at the discretion of the investigator, if necessary.

##### 6.1.5 Vaccination Pause Rules

Adverse events meeting any one of the following criteria will result in a hold being placed on subsequent vaccinations pending further review by the SMC:

- Any SAE attributed to vaccine.
- Any toxicity grade 3 (severe) solicited single AE term occurring in ≥ 7 subjects across any single SARS-CoV-2 rS construct following vaccination (first and second vaccinations to be assessed separately).
- Toxicity grade 3 (severe) solicited single prespecified laboratory value occurring in ≥7 subjects across any single SARS-CoV-2 rS construct following injection (first and

second vaccinations to be assessed separately). Prespecified laboratory values to be evaluated include creatinine, ALT, AST, bilirubin, hemoglobin, complete white blood count, and platelets.

- Any grade 3 (severe) unsolicited single AE preferred term for which the investigator assesses as related which occurs in ≥ 7 subjects across any single SARS-CoV-2 rS construct, within 49 days following vaccination (first).

The sponsor, along with medical monitor, may request an SMC review for any safety concerns that may arise in the trial and not associated with any specific pause rule.

##### 6.1.6 Safety Monitoring

Safety oversight will be conducted by a SMC that is an independent group of experts that monitors subject safety and advises Novavax, Inc. The SMC members will be separate and independent of site personnel participating in this study and should not have a scientific, financial, or other conflict of interest related to this study or the sponsor. The SMC will consist of members with appropriate expertise to contribute to the interpretation of the data from this study.

The SMC will operate under the rules of a sponsor-approved charter that will be approved at the organizational meeting of the SMC. At this time, data elements that the SMC needs to assess will be clearly defined based on the vaccination pause rules (Section 6.1.5). Procedures for SMC reviews/meetings will be defined in the charter. The SMC will review applicable data for safety assessments and any clinical data that may be of significance to this review (eg, demographics, vaccination timing, medications) based on the reason for invoking the pause rule. If needed, a narrative will be supplied by the medical monitor (eg, SAE). Additional data may be requested by the SMC, and interim reports may be generated as deemed necessary and appropriate by Novavax, Inc. The SMC is expected to review data in an unblinded fashion if required for safety assessment.

Novavax, Inc. or the SMC chair may convene the SMC on an ad hoc basis when a vaccination pause rule is met, for immediate concerns regarding observations during this study, or as needed. The SMC will also convene to review unblinded data through Day 35 to recommend advancement to Part 2 of study.

#### 6.2 IMMUNOGENICITY ASSESSMENTS

Blood samples will be taken at the time points specified in the SOE (Table 3-2) to assess immune response. Immune measurements (ELISA) will be conducted on serum (IgG) for SARS-CoV-2 rS protein antigen(s). Additional immunogenicity assessments specific to SARS-CoV-2 (or related variants) include human ACE-2 receptor inhibition assay, specific monoclonal Abs competition assays to other SARS-CoV-2 epitopes, neutralizing antibody and other tests of immune protection using passive antibody live virus challenge models (if available and appropriate). Additional assays may be developed or used for vaccine characterization.

Whole blood and blood samples for PBMC harvesting will be taken at the time points specified in the SOE (Table 3-2) for assessing cell-mediated immunity by measurement of cytokines produced using whole blood or after in vitro stimulation of PBMCs.

Details for the collection, processing, storage, and shipping of immunogenicity samples will be provided to the clinical unit separately.

Immunogenicity samples will be shipped to the clinical immunology department at Novavax, Inc. (Gaithersburg, MD) or 360biolabs (Melbourne, Victoria, Australia) for initial analyses.

Subjects will be asked to provide consent for the use of samples for future testing for other viruses and/or sequencing of the SARS-CoV-2 in positive specimens or assay development specific to SARS-CoV-2 (or related variants). Aliquots of all collected samples from this study may be retained for the stated purposes for a maximum of 25 years (starting from the date at which the last subject had the last study visit), unless local rules, regulations, or guidelines require different timeframes or different procedures, in accordance with subject consent.

### 7. STATISTICAL ANALYSIS PLANS

#### 7.1 SAMPLE SIZE CALCULATIONS

The sample size for this study is based on clinical and practical considerations and not on a formal statistical power calculation. The sample size is considered sufficient to evaluate the objectives of the study. With 25 subjects in each treatment group, there is a 92.8% probability to observe at least 1 subject with an AE if the true incidence of the AE is 10% and a 72.3% probability if the true incidence of the AE is 5%. With 100 subjects receiving at least

1 vaccination per SARS-CoV-2 rS construct, there is a greater than 99% probability to observe at least 1 subject with an AE if the true incidence of the AE is at least 5%.

Table 7-1 presents the width of the 95% CI (based on the Clopper-Pearson method) for 23 subjects in each treatment group (assuming approximately 5% of dosed subjects will be excluded from the per-protocol analysis set) under seroresponse rate assumptions between 50% and 90%.

**Table 7-1 Width of the 95% Confidence Interval (based on the Clopper-Pearson method)**

| Seroresponse Rate (%) | 95% CI (%) | Width (%) of 95% CI |
|---|---|---|
| 50 | 28.7, 71.3 | 42.6 |
| 60 | 37.7, 79.6 | 41.9 |
| 70 | 47.5, 87.1 | 39.6 |
| 80 | 58.2, 93.6 | 35.3 |
| 90 | 70.3, 98.5 | 28.2 |

Notes: Width of 2-sided 95% confidence interval (based on the Clopper-Pearson method) for the specified seroresponse rate produced by a sample size of 23 subjects is presented. PASS 15.0.7 Confidence Intervals for One Proportion is used in the calculation.

#### 7.2 ANALYSIS SETS

The safety analysis set will include all subjects who receive at least 1 dose of study vaccine (SARS-CoV-2 rS or placebo). Subjects will be analyzed according to the vaccine actually received.

The per-protocol analysis set will be determined for each study visit and will include all subjects who receive at least 1 dose of study vaccine (SARS-CoV-2 rS or placebo), have at least a baseline and 1 serum sample result available after vaccination, and have no major protocol violations that impact immunogenicity response at the corresponding study visit. All subjects in the per-protocol analysis set will be analyzed according to the study vaccine the subject was randomized to receive and not according to what was actually received, in the event there is a discrepancy.

The intent-to-treat analysis set will include all subjects who are randomized, regardless of protocol violations or missing data. The intent-to-treat analysis set will be used for supportive analyses.

#### 7.3 STATISTICAL ANALYSIS

Details of all statistical analyses will be described in a SAP.

All data collected will be presented in data listings. Data from subjects excluded from an analysis set will be presented in the data listings but not included in the calculation of summary statistics for that analysis set.

Data from subjects receiving placebo will be pooled across cohorts for all presentations.

For categorical variables, frequencies and percentages will be presented. Continuous variables will be summarized using descriptive statistics (number of subjects, mean, median, minimum, and maximum).

Baseline demographic and background variables will be summarized by treatment group. The number of subjects who enroll in the study and the number and percentage of subjects who complete the study will be presented. Frequency and percentage of subjects who withdraw or discontinue from the study, and the reason for withdrawal or discontinuation, will also be summarized.

##### 7.3.1 Safety Analyses

Numbers and percentages (with 95% CIs based on the Clopper-Pearson method) of subjects with solicited local and systemic AEs though 7 days after each vaccination will be summarized by treatment group and the maximum toxicity grade over 7 days after each vaccination. The duration of solicited local and systemic AEs after each vaccination will also be summarized by treatment group.

Unsolicited AEs will be coded by preferred term and system organ class using the latest version of MedDRA and summarized by treatment group as well as by severity and relationship to study vaccine. Adverse events through 49 days after first vaccination; all MAAEs through 105 days after first vaccination; and any MAAE related to vaccine, SAE, or AESI through 365 days after final vaccination will be listed separately and summarized by treatment group.

Actual values, changes from baseline (where indicated), and toxicity grading for clinical safety laboratory test results and vital sign measurements will be summarized by treatment group at each timepoint using descriptive statistics. Shift from baseline in toxicity grades will also be summarized.

Concomitant medications will be summarized by treatment group and preferred drug name as coded using the World Health Organization drug dictionary.

##### 7.3.2 Immunogenicity Analyses

The primary immunogenicity analyses will be performed using the per-protocol analysis set.

For the serum IgG antibody levels specific for the SARS-CoV-2 rS protein antigen(s) as detected by ELISA, the geometric mean at each study visit, and the geometric mean fold rise comparing to the baseline (Day 0) at each post-vaccination study visit, along with 95% CI will be summarized by treatment group. The 95% CI will be calculated based on the t-distribution of the log-transformed values for geometric means or geometric mean fold rises, then back transformed to the original scale for presentation. The seroconversion rate (proportion of subjects with ≥2-fold and also ≥4-fold rises in ELISA units), and seroresponse rate (proportion of subjects with rises in ELISA units exceeding the $95^{th}$ percentile of placebo recipients at the same time point) along with 95% CIs based on the Clopper-Pearson method will be summarized by treatment group at each post-vaccination study visit. An ANCOVA model will be constructed at each post-vaccination study visit on the log-transformed titer, including the treatment group as a fixed effect and the baseline log-transformed titer as a covariate. Comparisons of selected treatment groups will be performed within each visit. Additional covariates, such as site and age, may be explored as supportive analyses. Difference in the seroconversion rate and seroresponse rate between selected treatment groups along with 95% CIs within each visit will be calculated using the method of Miettinen and Nurminen (Miettinen and Nurminen 1985).

Neutralization assay specific for the SARS-CoV-2 wildtype (or variant) will be developed and appropriate testing parameters similar to ELISA will be utilized to assess functional antibody response. Refer to the laboratory manual and SAP.

Cell-mediated response for both Th1 and Th2 pathways will be assessed by cytokine profiling in either whole blood with flow cytometry and/or harvested PBMCs and summarized by treatment group in the per-protocol analysis set. Refer to the laboratory manual and SAP.

Similar summaries will be generated for the other immunogenicity endpoints identified in the future.

#### 7.4 HANDLING OF MISSING DATA

For calculating geometric means and geometric mean fold rises, immunogenicity values reported as below the LLOQ will be replaced by 0.5 × LLOQ. Values that are greater than the ULOQ will be replaced by the ULOQ. Missing results will not be imputed.

#### 7.5 INTERIM ANALYSES

Due to the nature of the pandemic, rolling interim analyses are planned for rapid vaccine development. The sentinel safety group of 6 subjects (assigned unblinded active vaccine constructs with Matrix-M) will be tested (ELISA and neutralization assay) per construct (cohort) immediately after Day 21 and Day 35 for early awareness of vaccine responsiveness.

Interim limited analyses at the aggregate treatment level only will be conducted when all subjects in a cohort have completed their respective Days 21, 35, 49, 105, and/or 189 visits. Refer to the SAP for more details. These interim analyses will allow decisions to initiate investigation of further nanoparticle constructs/doses, including advancement to Part 2 of study (based on Day 35 data), and will include (at a minimum) the following:

- Day 21 – Demographics and baseline characteristics, reactogenicity (local and systemic) following first vaccination (by toxicity grade and day), ELISA IgG GMT (baseline and Day 21) and seroconversion rate (≥4-fold), neutralization response (baseline and Day 21) expressed as titer or LD50.

- Day 35 – Reactogenicity (local and systemic) following first and second vaccination (by toxicity grade and day), laboratory values (by toxicity grading), vital signs (by toxicity grading), AEs (by classifications), ELISA IgG GMT (baseline, Day 21, Day 35) and seroconversion rate (≥4-fold), neutralization response (baseline, Day 21, Day 35) expressed as titer or LD50.

- Informational unblinded analysis will be provided to enable a safety review by the SMC through Day 35 on all subjects with notification of recommendation to advance into Part 2 of study. Refer to the SMC charter for more details. Additional limited interim analyses at the aggregate level may be performed at Days 49, 105, and/or Day 189 to inform on longer term safety and immune persistence if needed. The SAP will pre-specify such analyses but may be updated to reflect learnings based on the prior analyses in such instances. The final database lock will occur after Day 386 and include all primary and secondary endpoints.

### SECTION 2 - PART 2 OF STUDY

### PROTOCOL SYNOPSIS (PART 2)

**PROTOCOL NO.:** 2019nCoV-101

**TITLE:** A 2-Part, Phase 1/2, Randomized, Observer-Blinded Study to Evaluate the Safety and Immunogenicity of a SARS-CoV-2 Recombinant Spike Protein Nanoparticle Vaccine (SARS-CoV-2 rS) With or Without Matrix-M™ Adjuvant in Healthy Subjects

**STUDY PHASE:** Phase 1 (Part 1), with expansion to Phase 2 (Part 2) following a review of safety and immunogenicity.

**STUDY SITES:** For Phase 2 (Part 2), up to 40 clinical sites across Australia and/or the United States. Refer to Section 1 for Phase 1 study site information.

**OBJECTIVES AND ENDPOINTS:**

**The primary objectives are:**

- To identify the optimal dose across age strata based on immune response (IgG antibody to SARS-CoV-2 rS) at Day 35 and whether baseline immune status has an impact.
- To accumulate a safety experience for the candidate vaccine in healthy adult subjects based on solicited short-term reactogenicity across a broad age spectrum (by toxicity grade) and by AE profile for primary vaccination (through Day 35).
- To identify dose(s) to potentially take forward in an emergency use authorization setting and/or for Phase 3 efficacy or effectiveness trial(s).

**The secondary objectives are:**

- To assess if the single-dose regimens can provide similar (or adequate) priming immune response at Day 35 (compared to the two-dose regimens and to placebo) and whether baseline immune status alters such response (IgG antibody to SARS-CoV-2 rS).
- To define the optimal dosing regimen in subjects who are naïve and those with pre-existing antibodies to SARS-CoV-2 (if enough subjects are identified with pre-existing antibodies) as assessed by the immune response (IgG antibody to SARS-CoV-2 rS and angiotensin-converting enzyme 2 [ACE2] receptor binding inhibition) to the various regimens at Day 21 (post first dose), Day 35 (post second dose), and Day 217. Optimal dosing regimen to be assessed across full age spectrum and by age strata (18-59, 60-84 years).
- To describe the amplitude, kinetics, and durability of immune responses to the various regimens in terms of ELISA units of serum IgG antibodies to SARS-CoV-2 rS and titers of ACE2 receptor binding inhibition at selected time points and relative to whether subjects had pre-existing antibodies to SARS-CoV-2. To include reverse cumulative distribution curves.
- To describe the immune responses to the various regimens in terms of titers of neutralizing antibody at selected time points and relative to whether subjects had pre-existing antibodies to SARS-CoV-2 (subset of subjects). Optimal dosing regimen to be assessed across full age spectrum and by age strata (18-59, 60-84 years).

- To assess immune responses to the various regimens at 6 months and whether a boost at 6 months for a subset of the subjects enrolled in the 5-µg dose regimens (Treatment Groups B and C) induces immune memory and is beneficial to maintain immune response in terms of IgG and neutralizing antibodies to SARS-CoV-2 rS and ACE2 receptor binding inhibition.
- To assess immune responses to the various regimens at 12 months (all treatment groups) and whether a boost at 12 months for subjects enrolled in the 5-µg dose regimens (Treatment Groups B and C) induces immune memory and is beneficial to maintain immune responses in terms of IgG and neutralizing antibodies to SARS-CoV-2 rS and ACE2 receptor binding inhibition.
- To assess overall safety through 35 days after prime vaccination is initiated (1 or 2 doses) for all AEs; from 6-month boost (Day 189) through 28 days after 6-month boost (Day 217); and, for subjects in Treatment Groups B and C, from 12-month boost (Day 357) through 28 days after 12-month boost (Day 385) (Table S2-3) for all AEs; and through the EOS for any MAAE attributed to vaccine, AESI, or SAE.
- To assess for occurrence of COVID-19 disease as measured by PCR confirmation following subject-reported symptoms and to assess disease severity (virologically confirmed, mild, moderate, severe) and duration by patient-reported outcomes (eg, InFLUenza Patient-Reported Outcome [FLU-PRO©]) in those immunized with SARS-CoV-2 rS compared to placebo.
- To assess cell-mediated response: Type 1 T helper (Th1) or Type 2 T helper (Th2) predominance by various vaccine regimens (eg, IL-2, IL-4, IL-5, IL-6, IL-13, TNFα, INFγ using flow cytometry, ELISpot, or other system) in whole blood and/or harvested PBMC cells (in response to in vitro stimulation with SARS-CoV-2 rS protein) (subset of subjects).

**Exploratory objectives are:**

- To explore application of the FLU-PRO instrument to categorize COVID-19 severity, for maximal illness severity assessment, and the time course and severity of clinical symptomatology for COVID-19 cases for treatment groups.
- To utilize additional assays (current or to be developed) to best characterize the immune response for future vaccine development needs.
- To assess overall safety through 35 days after prime vaccination is initiated (1 or 2 doses) for solicited AEs; by SARS-CoV-2 positivity at Day 0, if greater than 10% of total subjects are SARS-CoV-2 positive at Day 0.
- To assess immune responses to the various regimens at 12 months and whether a boost at 12 months for subjects enrolled in the 5-µg dose regimens (Treatment Groups B and C) induces immune memory and is beneficial to maintain immune responses in terms of IgG and neutralizing antibodies to SARS-CoV-2 rS and ACE2 receptor binding inhibition for new variants, including, but not limited to, the South Africa variant B.1.351.

**The primary endpoints are comparisons of treatment regimens**:

- Serum IgG antibody levels specific for the SARS-CoV-2 rS protein antigen(s) as detected by ELISA using GMT or SCR for the two-dose regimens by dose at Day 35 regardless of baseline immune status and stratified by baseline immune status. Derived/calculated endpoints based on these data will include geometric mean ELISA units (GMEUs), geometric mean fold rise (GMFR), and SCR.

- Two-dose regimens by dose compared to placebo

**SCR** is defined as the percentage of subjects with a post-vaccination titer ≥ 4-fold.

**Positive baseline status** (+/-) using GMT and/or positive PCR at baseline.

- Numbers and percentages (with 95% CIs) of subjects with solicited AEs (local, systemic) for 7 days following each primary vaccination (Days 0 and 21) by severity score, duration, and peak intensity. Unsolicited AEs (eg, treatment-emergent, serious, suspected unexpected serious, those of special interest, MAAEs) through 35 days by MedDRA classification, severity score, and relatedness.

**The secondary endpoints are comparisons of treatment regimens**:

- Serum IgG antibody levels specific for the SARS-CoV-2 rS protein antigen(s) as detected by ELISA using GMT or SCR (≥ 4-fold change) for the single-dose regimens compared to the two-dose regimens and to placebo at Day 21 and Day 35 regardless of baseline immune status and stratified by baseline immune status. Derived/calculated endpoints based on these data will include GMEUs, GMFR, and SCR (≥ 4-fold change).
- Serum IgG antibody levels specific for the SARS-CoV-2 rS protein antigen as detected by ELISA, described across study time points with derived/calculated endpoints to include GMEUs, GMFR, and SCR (≥ 4-fold change) for the single-dose regimens compared to the two-dose regimens and to placebo, stratified by baseline immune response.
- Epitope-specific immune responses to the SARS-CoV-2 rS protein receptor binding domain measured by serum titers in an ACE2 receptor binding inhibition assay to include GMT or concentration, GMFR, and SCR (≥ 4-fold change) for the single-dose regimens compared to the two-dose regimens and to placebo.
- Neutralizing antibody activity at Days 35, 217, and at 357 for all Treatment Groups, and additionally at Days 371 and 546 for Treatment Groups B and C relative to baseline (Table 10-5) in a subset of subjects by absolute titers and change from baseline, including the SCR (≥ 4-fold change). Analysis to include subjects by treatment group, by age (18-59, 60-64 years) and relative to whether subjects had pre-existing antibodies to SARS-CoV-2. A sampling scheme to identify a subset of such subjects will be deployed.
- Serum IgG antibody levels specific for the SARS-CoV-2 rS protein antigen(s) as detected by ELISA using GMT or GMFR at Days 189, 217, and 357 for all Treatment Groups, and additionally at Days 371 and 546 for Treatment Groups B and C (Table 10-5) for boosting assessment with either placebo or active boost.
- All MAAEs through Day 217, and then related MAAEs until EOS, and all AESIs, or SAEs through the EOS by MedDRA classification, severity score, and relatedness.
- Vital sign measurement before vaccination and as clinically needed during the 30-minute post-vaccination observation period. Vital sign measurements at all other time points to be classified by descriptive statistics (eg, mean, median, SD) by visit.

- Percentage of subjects with SARS-CoV-2 positivity as diagnosed by qualitative PCR following COVID-19 symptoms assessment from Day 28 through 6 months with severity classification, overall and by age strata (18-59, 60-84 years). If frequent clinical PCR-confirmed SARS-CoV-2 infections occur during the study follow-up period, vaccine efficacy assessments for primary vaccination for treatment groups compared to placebo may be generated. The criteria to determine whether this will occur will be documented in the SAP.
- Assessment of SARS-CoV-2 by qualitative PCR based on routine screening by nasal mid-turbinate sample self-collection from Day 28 through 6 months without symptomatology to further describe epidemiologic evolution of the pandemic and potential effect of vaccination.
- Assessment of cell-mediated (Th1/Th2) pathways as measured by whole blood (flow cytometry) and/or in vitro PBMC stimulation (eg, ELISpot, cytokine staining) with SARS-CoV-2 rS protein(s) as measured on Days 0, 7, and 28.

**Exploratory endpoints are:**

- Assessment of immune responses to the various regimens at 6 and 12 months for all treatment groups and at 18 months for Treatment Groups B and C and whether a boost at 6 months (all Treatment Groups) and again at 12 months (Treatment Groups B and C) induces immune memory and is beneficial to maintain immune responses to SARS-CoV-2 rS in terms of IgG and neutralizing antibodies and ACE2 receptor binding inhibition for new variants, including, but not limited to, the South Africa variant B.1.351.
- Assessment of COVID-19 severity and the time course of symptom scores using the FLU-PRO instrument for up to 10 days following a qualifying illness episode, to assess the potential use of patient-reported symptom-based severity criteria for application in future clinical endpoint studies, with assessment of maximal severity of illness episodes and the relationship between FLU-PRO measures and COVID-19 clinically defined severity categories.
- Any additional assays to measure immune response, protection, or potential safety signals.
- Numbers and percentages (with 95% CIs) of subjects with solicited AEs (local, systemic) for 7 days following each primary vaccination (Days 0 and 21) by severity score, duration, and peak intensity by SARS-CoV-2 positivity at Day 0, if greater than 10% of total subjects are SARS-CoV-2 positive at Day 0.

**STUDY DESIGN:**

If adequate safety and desired immune responses are observed at interim analyses during Part 1, the study may be immediately extended, at the sponsor's discretion, to include Part 2. Only 1 construct for SARS-CoV-2 rS will be evaluated in Part 2.

Part 2 is a Phase 2, randomized, placebo-controlled, observer-blinded study to evaluate the safety and immunogenicity of a SARS-CoV-2 rS with Matrix-M™ adjuvant in male and female subjects. Subjects will be healthy adults based on medical history and physical examination. It is anticipated that Matrix-M adjuvant will be required for an adequate response in this population and for dose-sparing needs.

After signing the informed consent form (ICF), subjects may be screened within a window of up to 45 days; however, SARS-CoV-2 serostatus across the enrolling sites should not be expected to be more than 15% of the overall population will be SARS-CoV-2 positive at baseline. Subjects will be asked to provide consent for the use of samples for future testing for other viruses and/or sequencing of the SARS-CoV-2 in positive specimens or assay development specific to SARS-CoV-2 (or related variants).

Approximately 750 healthy male and female subjects between 18 and 84 years of age inclusive (≥18 years to < 85 years) will be randomized in each country, with approximately 50% of subjects overall in the study ≥60 years of age. Two-factor, 2-level stratification will be employed (ages 18-59 and 60-84; study site). Subjects who meet the criteria for study entry will initially be randomized in a 1:1:1:1:1 ratio to 1 of 5 vaccine groups (Table S2-1).

Up to approximately 1500 subjects could be enrolled for the 5 vaccine groups across the 2 countries to potentially mitigate the risk of enrollment in either country being delayed, or the expected availability of study data from either country being delayed due to the potential impact of the pandemic on feasibility of study vaccine delivery, possibilities for adequate specimen and data collection, and feasible specimen transportation. This flexibility would allow review of study data on approximately 500 subjects to facilitate initiation of Phase 3 clinical development as soon as possible.

For the Part 2 component of the study, following study initiation, enrollment of older adult subjects (≥ 60 to 84 years of age) will be paused when approximately 50 subjects in the older age group are enrolled in each of the 5 study vaccine groups (ie, approximately 250 older adult subjects in total enrolled subjects across all sites). The Safety Monitoring Committee (SMC) will then review solicited reactogenicity data for both study age groups for 5 days solicited reactogenicity following the first vaccine dose (Day 0 through Day 4, inclusive), when all subjects in the older adult group have accrued these 5 days of data. Enrollment of adult subjects (≥ 18 to 59 years of age) will not be paused during this SMC review unless general vaccination pause rules detailed in Section 13.1.5 are also met. Details of this review, and processes to restart enrollment of older adult subjects will be documented in the SMC Charter.

**Table S2-1 Treatment Groups as Originally Planned (Part 2)**

| Treatment Group | Number of Subjects | Day 0 | Day 21 (−1 to +3 days) | Day 189 (±15 days) |
|---|---|---|---|---|
| | | SARS-CoV-2 rS + Matrix-M Adjuvant | SARS-CoV-2 rS + Matrix-M Adjuvant | SARS-CoV-2 rS + Matrix-M Adjuvant |
| A | 150-300 | Placebo | Placebo | Placebo |
| B | 150-300 | 5 µg + 50 µg | 5 µg + 50 µg | Placebo |
| C | 150-300 | 5 µg + 50 µg | Placebo | 5 µg + 50 µg |
| D | 150-300 | 25 µg + 50 µg | 25 µg + 50 µg | Placebo |
| E | 150-300 | 25 µg + 50 µg | Placebo | 5 µg + 50 µg |

Note: The first dose represents the amount of antigen (SARS-CoV-2 rS) and the second dose represents the amount of adjuvant (Matrix-M). For example, 5 µg + 50 µg represents 5 µg SARS-CoV-2 rS + 50 µg Matrix-M adjuvant.

Subjects initially randomized to treatment groups B and C through Day 21 will be re-randomized at a 1:1 ratio to receive either a booster dose of vaccine (B2 and C2) or placebo (B1 or C1) at Day 189 (Table S2-2). This change to the boosting regime is being implemented in order to gain additional immunogenicity and safety data for subjects who receive 1 or 2 doses of 5 µg SARS-CoV-2 rS + 50 µg Matrix-M1 adjuvant on Days 0 and 21 (ie, the dose level intended for licensure). In addition, subjects in treatment group E who were initially scheduled to receive a booster dose of vaccine at Day 189 will now receive placebo. Treatment group E is no longer being boosted as the 25-µg dose of SARS-CoV-2 rS is no longer being taken forward into later phase studies of the vaccine and the value of gathering additional boosting immunogenicity data for this group is considered minimal.

**Table S2-2 Treatment Groups, Re-randomized at Day 189 (Part 2)**

| Treatment Group | Number of Subjects | Day 0<br>SARS-CoV-2 rS + Matrix-M Adjuvant | Day 21 (−1 to +3 days)<br>SARS-CoV-2 rS + Matrix-M Adjuvant | Day 189 (±15 days)<br>SARS-CoV-2 rS + Matrix-M Adjuvant |
|---|---|---|---|---|
| A | 300 | Placebo | Placebo | Placebo |
| B1 | 150 | 5 µg + 50 µg | 5 µg + 50 µg | Placebo |
| B2 | 150 | 5 µg + 50 µg | 5 µg + 50 µg | 5 µg + 50 µg |
| C1 | 150 | 5 µg + 50 µg | Placebo | Placebo |
| C2 | 150 | 5 µg + 50 µg | Placebo | 5 µg + 50 µg |
| D | 300 | 25 µg + 50 µg | 25 µg + 50 µg | Placebo |
| E | 300 | 25 µg + 50 µg | Placebo | Placebo |

Note: The first dose represents the amount of antigen (SARS-CoV-2 rS) and the second dose represents the amount of adjuvant (Matrix-M). For example, 5 µg + 50 µg represents 5 µg SARS-CoV-2 rS + 50 µg Matrix-M adjuvant.

Subjects in Treatment Groups B and C who agree to continue study participation for an additional 6 months will receive a booster dose of vaccine (B1, B2, and C1) or placebo (C2) at Day 357 (Table S2-3). This change to the boosting regime is being implemented in order to gain additional immunogenicity and safety data for subjects who receive 1 or 2 doses of 5 µg SARS-CoV-2 rS + 50 µg Matrix-M1 adjuvant on Days 0 and 21 (ie, the dose level intended for licensure) and a booster dose of 5 µg SARS-CoV-2 rS + 50 µg Matrix-M1 adjuvant on Day 189.

**Table S2-3 Treatment Groups B and C, Additional Dose on Day 357 (Part 2)**

| Treatment Group | Number of Subjects | Day 0<br>SARS-CoV-2 rS + Matrix-M Adjuvant | Day 21 (−1 to +3 days)<br>SARS-CoV-2 rS + Matrix-M Adjuvant | Day 189 (±15 days)<br>SARS-CoV-2 rS + Matrix-M Adjuvant | Day 357 (±15 days)<br>SARS-CoV-2 rS + Matrix-M Adjuvant |
|---|---|---|---|---|---|
| B1 | 150 | 5 µg + 50 µg | 5 µg + 50 µg | Placebo | 5 µg + 50 µg |
| B2 | 150 | 5 µg + 50 µg | 5 µg + 50 µg | 5 µg + 50 µg | 5 µg + 50 µg |
| C1 | 150 | 5 µg + 50 µg | Placebo | Placebo | 5 µg + 50 µg |
| C2 | 150 | 5 µg + 50 µg | Placebo | 5 µg + 50 µg | Placebo |

Note: The first dose represents the amount of antigen (SARS-CoV-2 rS) and the second dose represents the amount of adjuvant (Matrix-M). For example, 5 µg + 50 µg represents 5 µg SARS-CoV-2 rS + 50 µg Matrix-M adjuvant.

Study vaccinations will comprise up to 3 intramuscular (IM) injections (Day 0 and Day 21 [priming doses], and Day 189 [6-month booster], in Treatment Groups A, D, and E, and up to 4 IM injections (Day 0 and Day 21 [priming doses], Day 189 [6-month booster], and Day 357 [12-month booster]) in Treatment Groups B and C, ideally in alternating deltoids for priming doses, with the study vaccine assigned in a full dose injection volume of approximately 0.5 mL. All vaccinations will be administered on an outpatient basis by designated site personnel in a way to maintain the blind, and if subjects are not able to attend sites due to local recommendations/restrictions related to the COVID-19 pandemic, home visits for vaccine administration may be utilized, or alternative models of vaccine administration such as drive-through clinics. Following study vaccination, subjects should either remain in the clinic or under study staff observation if vaccinated during home visits or at other locations, for at least 30 minutes post-vaccination to be monitored for any immediate anaphylaxis reactions.

Any pharmacy preparation with unblinded product will require unblinded site personnel who will not otherwise be involved in the study procedures or observation of subjects. The goal of Part 2 will be to utilize co-formulated product; however, the Part 2 site(s) may initiate dosing using the approach of bedside mixing as noted in the Part 1 portion of the protocol (with the appropriate concentrations being tested in Part 2).

Blood samples for immunogenicity assessments will be collected before vaccination and at selected time points following each vaccination. Immune measurements (ELISA) will be conducted on serum (IgG) for SARS-CoV-2 rS protein antigen(s) and ACE2 receptor binding inhibition. Additional immunogenicity assessments specific to SARS-CoV-2 (or related variants), including anti-nucleocapsid protein serology, will include a neutralizing antibody assay. Blood samples for serology will be collected at baseline but will not be used for inclusion/exclusion for randomization as a medical history will suffice; however, individuals with positive serologies (hepatitis B, hepatitis C, human immunodeficiency virus [HIV]) will not be included in the primary immunogenicity analysis.

Subgroup analyses of the primary immunogenicity analysis (ie, serum IgG antibody levels specific for the SARS-CoV-2 rS protein antigen[s] as detected by ELISA using GMT or SCR for the two-dose regimens by dose at Day 35 regardless of baseline immune status and stratified by baseline immune status), as measured by in-house SARS-CoV-2 IgG assay at the Novavax central immunology laboratory, will be undertaken. All formal analyses will utilize the cut-off of SARS-CoV-2 IgG from the Novavax central immunology laboratory.

Operational processes will aim to manage the number of SARS-CoV-2 antibody-positive subjects enrolled in the study, such that not more than 15% of subjects test SARS-CoV-2 antibody-positive at baseline. This will be done using community seroprevalence estimates considered during site selection processes, and baseline SARS-CoV-2 antibody results are not required to be known prior to subject enrollment.

Safety assessments will include monitoring and recording of solicited (local and systemic reactogenicity events) and unsolicited AEs; MAAEs; AESI; SAEs; vital sign measurements; and targeted physical examination findings. Symptoms related to either a suspected, probable, or confirmed COVID-19 case for illness events starting prior to Day 28 should be recorded as unsolicited AEs, or multiple symptom-based AEs can be aggregated into a single AE of a suspected, probable, or confirmed COVID-19 case.

If a US subject is unblinded in the study and receives an approved/authorized vaccine from a different manufacturer, all vaccine administration errors, SAEs, cases of multisystem inflammatory syndrome, and hospitalized or fatal cases of COVID-19 following vaccination with the approved/authorized vaccine must be reported to the Vaccine Adverse Event Reporting System (VAERS). Investigators in Australia should follow local regulatory reporting guidance for safety events that occur in subjects who receive approved/authorized vaccines within the study.

COVID-19 disease monitoring for qualifying symptoms of suspected COVID-19 will commence every 14 days beginning at Day 28. Table S2-4 details qualifying symptoms of suspected COVID-19. If a subject has qualifying symptoms, this will trigger a request to subjects to self-sample according to protocol instructions (see below).

For efficacy assessments, COVID-19 severity will be categorized as virologically confirmed, mild, moderate, or severe according to protocol-specified criteria. Solicited and unsolicited AEs and any COVID-19 disease assessments may be conducted by electronic data capture/reporting. Telemedicine is acceptable in lieu of site visits for COVID-19 containment issues at any time (either community-, site-, or subject-defined).

An independent SMC will remain in effect from Part 1 of the study and will review safety data in aggregate when data snapshots are available (see Section 14.5) from the Day 21 and/or Day 35 time point (to inform potential stage-gate decisions for follow-on clinical studies) and every 3 to 4 months during Part 2 of the study or sooner if there are new safety signals noted by sponsor internal safety reviews or if an excess number of COVID-19 cases occur. The study will continue as planned during the SMC reviews, with the exception of when an enrollment pause will occur following initial enrollment of approximately 250 older adult subjects. An SMC Charter will document SMC membership and processes, data review time points, and safety tables that will be reviewed.

For Treatment Groups A, D, and E, Part 2 will consist of a screening period (Days −45 to 0); vaccination days (Days 0, 21, and 189); outpatient study visits on Day 0 and on Days 21 (−1 to +3 days), 35 (−1 to +3 days), 105 (± 7 days), 189 (± 15 days), 217 (+7 days), 357 ± 15 days), and a phone visit at Days 273 (± 15 days). Subjects in an immunogenicity subset for PBMC collection will additionally have outpatient study visits on Days 7 (−1 to +3 days) and 28 (−1 to +3 days) (See Table 10-4). For Treatment Groups B and C, Part 2 will consist of a screening period (Days −45 to 0); vaccination days (Days 0, 21, 189, and 357); outpatient study visits on Day 0 and on Days 21 (−1 to +3 days), 35 (−1 to +3 days), 105 (± 7 days), 189 (± 15 days), 217 (+7 days), 357 (± 15 days), 371 (−1 to +3 days), and Day 546/EOS (± 15 days), and phone visits at Day 273 (± 15 days), Day 385 (−1 to +3 days), and Day 475 (± 15 days). Subjects in an immunogenicity subset for PBMC collection will additionally have outpatient study visits on Days 7 (−1 to +3 days) and 28 (−1 to +3 days) (See Table 10-4 and Table 10-5).

The duration of the Part 2, excluding screening, is approximately 12 months for subjects enrolled in Treatment Groups A, D, and E and approximately 18 months for those subjects in Treatment Groups B and C who agree to stay in the study through the Day 357 boost and 6-month follow up. Visits requiring vaccination or blood sampling may occur via home visits or using local services, if conducted by qualified personnel (eg, phlebotomy, home health, external laboratory, home visit by site staff).

If the Novavax study vaccine or another vaccine from a different manufacturer is demonstrated to be safe and efficacious and approved and/or authorized for use by regulatory authorities in the US or Australia, subjects for whom the new approved/authorized vaccine is recommended and available will be counseled with respect to their options. These subjects may be offered the opportunity to be unblinded so that those who received placebo may be offered the Novavax vaccine or another approved/authorized vaccine, as appropriate, outside the protocol procedures. Subjects who received the Novavax vaccine and who wish to receive an approved/authorized vaccine from another manufacturer will be advised to discuss this plan with their healthcare provider given the current lack of safety data regarding the sequential administration of vaccines made by different manufacturers. Subjects who are unblinded and receive an approved/authorized vaccine in this manner will be strongly encouraged to remain in study for safety follow-up as defined in the protocol. However, subjects also have the right to discontinue participation in the study at any time.

Due to the ongoing pandemic, recent national regulatory and local HREC/IRB and public health guidance will be applied at the site locations regarding alternations in the ability of study subjects to attend an investigational site for protocol specified visits, with the site's investigator being allowed to conduct safety assessments (eg, telephone contact, virtual visit via telemedicine, alternative location for assessment, including local laboratories or imaging centers) when necessary and feasible, as long as such visits are sufficient to assure the safety of study subjects. Serum samples may be drawn using local phlebotomy services, home health, or other modalities if site visits cannot occur. Vaccination visits must have adequate oversight for issues associated with immediate severe reactions but may need to occur outside of the clinical site depending on the pandemic situation (eg, home vaccinations).

**COVID-19 Disease Monitoring**

**Monitoring of Qualifying Symptoms of Suspected COVID-19 Disease**

Note: Based on the extremely low incidence of SARS-CoV-2 transmission in Australia, monitoring for symptoms associated with SARS-CoV-2 infection via the ePRO system, self-sampling and completion of the electronic FLU-PRO instrument will no longer be required for subjects enrolled at Australian sites following approval of Amendment 7 (Version 8.0) of the protocol.

Subjects at US sites will continue to be monitored via ePRO system (data capture) for symptoms associated with SARS-CoV-2 infection (eg, cough, fever, sore throat, difficulty breathing, and other symptoms ([Table S2-4]) every 14 days beginning at Day 28 until approximately Day 217. When COVID-19 disease symptom scoring indicates the need for sample collection for potential PCR confirmation, subjects at sites in the US will self-collect a nasal mid-turbinate sample for 2 consecutive days taken as close to the onset of symptoms as possible (ideal timing within 3 days). Symptom-based collection can be either due to passive or active surveillance using an ePRO system. Samples will not be collected after 14 days as a new symptom-based query will be initiated (eg, every 2 weeks).

Since self-sampling specimens for PCR confirmation under consideration for use in the study are not currently approved for diagnostic use, and depending on the site locations in relation to central study laboratories and associated delays in reporting related to specimen transportation, study PCR sampling is not an alternative for subjects to obtain diagnosis of potential COVID-19 in the context of individual clinical and public health case management. Subjects will be notified by sites of any positive or indeterminant PCR results when they are available and liaise with subjects' regular treating physicians in this instance. Negative study PCR results will not be routinely communicated to subjects, since a negative result for a non-diagnostic level test should not be interpreted as confirmation of absent SARS-CoV-2 infection. If subjects request results for PCR and results were negative, sites may provide the negative result to subjects, along with the context that negative results for the test should not be interpreted as excluding SARS-CoV-2 infection. Sites will follow required local processes around the notification of positive SARS-CoV-2 infections, which may involve public health reporting. Sites will assess local processes and availability of diagnostic quality SARS-CoV-2 infection testing and will communicate which options are locally available to subjects if they meet local clinical or public health criteria for testing.

Clinical symptoms of suspected COVID-19 disease from Day 0 to Day 28 will be reported to sites by subjects directly to sites by site-established communication channels outside of the ePRO system, and site staff will clinically assess the symptomatology and actions required that are consistent with locally applicable guidance for diagnosis of SARS-CoV-2 infection, which may vary by jurisdiction. If clinically indicated, actions could include unscheduled visits with site staff sampling of subjects, or referral of subjects into locally established PCR testing, with co-ordination with regular subject clinicians or clinicians locally established to assess and monitor potential COVID-19 cases.

**Table S2-4 Qualifying Symptoms of Suspected COVID-19 Disease**

| |
|---|
| Chills or fever or feverishness (reported by the subject, or fever ≥ 37.8°C, regardless of use of anti-pyretic medications) OR |
| New onset of any respiratory symptoms (cough, rapid breathing, shortness of breath, difficulty breathing, runny nose, nasal congestion, or sore throat) OR |
| New onset of the following other symptoms:<br>• Anosmia (smell disturbances)<br>• Ageusia (taste disturbances)<br>• Fatigue or tiredness or weakness<br>• Myalgia (muscle aches)<br>• Headache<br>• Nausea or vomiting<br>• Diarrhea |

Abbreviations: COVID-19 = coronavirus disease 2019.

**COVID-19 Disease Case Ascertainment**

**Sample collection for COVID-19 laboratory confirmation**

When a subject at a US site reports any of the qualifying symptoms of suspected COVID-19 disease listed in Table S2-4 after Day 28, he/she will begin daily self-collection of a nasal mid-turbinate sample for 2 consecutive days (self-collection on any of these days may be replaced by sample collection by an HCP, if the subject is admitted to the hospital or COVID-19 disease treatment facility and self-collection is not available). Timing of self-collection should be as close to the onset of symptoms as possible (ideally within 3 days). All nasal mid-turbinate samples collected over the 2-day period will be sent to a prespecified central laboratory where a validated PCR test will be performed for confirmation of SARS-CoV-2 infection. If both self-collected samples that are initially collected are PCR negative or indeterminate, and the subject has a clinical illness consistent with COVID-19 disease, site staff may request subjects to resample through nasal mid-turbinate sample collection for a further 2 consecutive days any time from 5 days after symptom onset.

Subjects enrolled at both US and Australian sites may also be seen by site personnel within the surveillance period to evaluate for potential COVID-19 disease, if acceptable based on the ongoing pandemic and subject containment requirements; and if seen, then a nasal mid-turbinate sample and an additional blood sample for immunogenicity assessments will be obtained. Any subject admitted to the hospital or COVID-19 treatment facility may utilize hospital testing for SARS-CoV-2 as the method of testing positivity. Endpoint collection will be obtained using hospital derived information which may include electronic medical records. For subjects enrolled at both US and Australian sites, any outpatient or public health PCR sample testing that has confirmed SARS-CoV-2 infection may be used to establish subject positivity for COVID-19 for study endpoints, provided subjects can provide some form of appropriate confirmation documentation.

Should a medical visit be warranted based on symptomatology (and allowed via local isolation guidance), such a visit may occur using telemedicine, home visitation, or clinic visit. Subjects experiencing shortness of breath or difficulty breathing must be seen by an HCP to assess tachypnea and blood oxygen saturation levels.

**PCR Resampling for Subsequent Illness Episodes**

If a subject becomes symptom-free (of the qualifying symptoms of suspected COVID-19) for 3 consecutive days, then the subject will be eligible for PCR resampling at the next onset of qualifying symptoms of suspected COVID-19 diseases.

**FLU-PRO Disease/Symptom Severity Patient-Reported Outcome Instrument**

The severity and symptom duration after qualifying symptoms of suspected COVID-19 disease have been met will also be monitored by subjects enrolled at US sites using the electronic FLU-PRO instrument, which may also include additional questions for COVID-19 disease depending on the status of validation of modified questions at the time of the study. The FLU-PRO will be completed daily up to 10 consecutive days if possible within the electronic collection tool by the patient, when a subject actively initiates reporting of qualifying symptoms for a potential COVID disease within the electronic patient diary, starting after qualifying symptoms of suspected COVID-19 disease have been met

(Table S2-4). However, if a subject reports in the additional questions after the FLU-PRO instrument symptoms questions within the electronic diary for 3 consecutive days during the 10-day follow-up period that they have returned to their usual health, then completion of the FLU-PRO tool on further days need not continue. If subjects then have the return of potential qualifying symptoms as part of a new illness episode, then the subject can have the new illness episode assessed again for potential qualifying symptoms through the electronic diary. FLU-PRO assessments therefore should only be associated with one potential qualifying illness episode at a time.

In case of severe clinical deterioration, the instrument may not be able to be completed for the full 10-day period. Subjects can only be reporting FLU-PRO scores in relation to one illness event at a time. Sites may use the FLU-PRO scores over time to monitor subjects' self-reported symptomatology during illness episodes.

##### COVID-19 Clinical Endpoint Disease Severity Definitions

Virologically confirmed COVID-19 disease (assessed by severity) will be graded as follows (virologically confirmed, mild, moderate, or severe) based on progression to the greatest severity during the course of illness (Table S2-5).

**Table S2-5 Endpoint Definitions of COVID-19 Disease Severity**

| COVID-19 Severity | Endpoint Definitions |
|---|---|
| **Virologically Confirmed** | ≥ 1 COVID-19 disease symptom in Table S2-4<br>**AND**<br>Does not meet criteria for mild, moderate or severe disease |
| **Mild** | ≥ 1 of:<br>• Fever (defined by subjective or objective measure, regardless of use of anti-pyretic medications)<br>• New onset cough<br>• ≥ 2 COVID-19 respiratory/non-respiratory symptoms in Table S2-4<br>**AND**<br>• Does not meet criteria for moderate or severe |
| **Moderate** | ≥ 1 of:<br>• Fever (defined by subjective or objective measure, regardless of use of anti-pyretic medications) + any 2 COVID-19 respiratory/non-respiratory symptoms in Table S2-4 for ≥ 3 days (need not be contiguous days)<br>• High fever (≥ 38.4°C) for ≥ 3 days (need not be contiguous days)<br>• Any evidence of significant LRTI:<br>– Shortness of breath (or breathlessness or difficulty breathing) with or without exertion (greater than baseline)<br>– Tachypnea: 20 to 29 breaths per minute at rest<br>– $SpO_2$: 94% to 95% on room air<br>– Abnormal chest x-ray or CT consistent with pneumonia or LRTI<br>– Adventitious sounds on lung auscultation (eg, crackles/rales, wheeze, rhonchi, pleural rub, stridor)<br>**AND**<br>• Does not meet criteria for severe disease |
| **Severe** | ≥ 1 of:<br>• Tachypnea: ≥ 30 breaths per minute at rest<br>• Resting heart rate ≥ 125 beats per minute |

**Table S2-5 Endpoint Definitions of COVID-19 Disease Severity**

| COVID-19 Severity | Endpoint Definitions |
|---|---|
| | • $SpO_2 \leq 93\%$ on room air or $PaO_2/FiO_2 < 300$ mmHg<br>• High-flow oxygen therapy or NIV/NIPPV (eg, BiPAP or CPAP)<br>• Mechanical ventilation or ECMO<br>• One or more major organ system dysfunction or failure (eg, cardiac/circulatory, pulmonary, renal, hepatic, and/or neurological, to be defined by diagnostic testing/clinical syndrome/interventions), including any of the following<br> o Acute respiratory distress syndrome (ARDS)<br> o Acute renal failure<br> o Acute hepatic failure<br> o Acute right or left heart failure<br> o Septic or cardiogenic shock (with shock defined as SBP < 90 mm Hg OR DBP < 60 mm Hg)<br> o Acute stroke (ischemic or hemorrhagic)<br> o Acute thrombotic event: AMI, DVT, PE<br> o Requirement for: vasopressors, systemic corticosteroids, or hemodialysis<br> o Admission to an ICU<br> o Death |

Abbreviations: AMI = acute myocardial infarction; BiPAP = bi-level positive airway pressure; CPAP = continuous positive air pressure; CT = computed tomography; DBP = diastolic blood pressure; DVT = deep vein thrombosis; ECMO = extracorporeal membrane oxygenation; $FiO_2$ = fraction of inspired oxygen; ICU = intensive care unit; LRTI = lower respiratory tract infection; NIPPA = non-invasive positive pressure ventilation; NIV = non-invasive ventilation; $PAO_2$ = partial pressure of oxygen in the alveolus; PE = pulmonary embolism; SBP = systolic blood pressure; $SpO_2$ = oxygen saturation.

#### COVID-19 Clinical Endpoint Case Definition

A subject whose nasal mid-turbinate sample tests positive for SARS-CoV-2 infection by qualitative PCR [ie, (+)-PCR-confirmed SARS-CoV-2 illness) and whose initial qualifying symptom(s) of suspected COVID-19 disease (Table S2-4) meets the criteria for symptomatic virologically confirmed, mild, moderate, or severe COVID-19 disease (Table S2-5) within 14 days of the initial qualifying symptom(s) will be defined as a single case in the secondary efficacy analysis.

#### STUDY POPULATION:

Healthy male or female subjects will be enrolled at up to 40 clinical sites across Australia and/or the United States. Approximately 750 healthy male and female subjects between 18 and 84 years of age (inclusive) will be randomized in each country, with approximately 50% of subjects overall in the study ≥60 years of age. Two-factor, 2-level stratification will be employed (ages 18-59 and 60-84; study site). Up to approximately 1500 subjects could be enrolled for the 5 vaccine groups across the 2 countries to potentially mitigate the risk of enrollment in either country being delayed, or the expected availability of study data from either country being delayed due to the potential impact of the pandemic on feasibility of study vaccine delivery, possibilities for adequate specimen and data collection, and feasible specimen transportation. This flexibility would allow review of study data on approximately 500 to facilitate initiation of Phase 3 clinical development as soon as possible.

For the Part 2 component of the study, following study initiation, enrollment of older adult subjects (≥ 60 to 84 years of age) will be paused when approximately 50 subjects in the older age group are enrolled in each of the 5 study vaccine groups (ie, approximately 250 older adult subjects in total enrolled subjects across all sites). The Safety Monitoring Committee (SMC) will then review reactogenicity data for both study age groups for 5 days following the first vaccine dose (Day 0 through Day 4, inclusive) when all subjects in the older adult group have accrued these 5 days of data. Enrollment of adult subjects (≥ 18 to 59 years of age) will not be paused during this SMC review unless general vaccination pause rules detailed in Section 13.1.5 are also met. Details of this review, and processes to restart enrollment of older adult subjects will be documented in the SMC Charter.

Depending on the current pandemic situation and ability to rapidly test, enrollment of subjects with prior SARS-COV-2 exposure/positive testing will be limited on a site-by-site basis with an overall goal of not more than 15% of the population being SARS-CoV-2 positive at baseline; however, the result for baseline seropositivity will not need to be known for subjects prior to enrollment, rather it will be retrospectively assessed from baseline serology, with potential shifts to enrollment in sites with lower levels of baseline seropositivity if needed during the enrollment period.

**Inclusion Criteria:**
Subject eligibility for the study (ie, subject reported to successfully meet all Inclusion Criteria and not meet any Exclusion Criteria) will be recorded in the subject source notes. Clinical validation of self-reported subject information related to eligibility is not routinely required for enrolment, unless specified for particular eligibility criteria.

Each subject must meet all of the following criteria to be enrolled in this study:

1. Healthy adult males or females between 18 and 84 years of age, inclusive, at screening who are of legal adult age in their local jurisdiction. Healthy status will be determined by the investigator based on medical history, vital sign measurements, and physical examination at screening.
2. The subject has a body mass index 17 to 35 kg/m$^2$, inclusive, at screening.
3. Willing and able to give informed consent prior to study enrollment and comply with study procedures.
4. Female subjects of childbearing potential (defined as any female who has experienced menarche and who is NOT surgically sterile [ie, hysterectomy, bilateral tubal ligation, or bilateral oophorectomy] or postmenopausal [defined as amenorrhea at least 12 consecutive months or documented plasma FSH level ≥40 mIU/mL]) must agree to be heterosexually inactive from at least 21 days prior to enrollment and through 6 months after the last vaccination OR agree to consistently use any of the following methods of contraception from at least 21 days prior to enrollment and through 6 months after the last vaccination:
    a. Condoms (male or female) with spermicide (if acceptable in country)
    b. Diaphragm with spermicide
    c. Cervical cap with spermicide
    d. Intrauterine device
    e. Oral or patch contraceptives

f. Norplant®, Depo-Provera®, or other in country regulatory-approved contraceptive method that is designed to protect against pregnancy

g. Abstinence, as a form of contraception, is acceptable if in line with the subject's lifestyle

**NOTE**: Periodic abstinence (eg, calendar, ovulation, symptothermal, post-ovulation methods), male partner vasectomy, and withdrawal are not acceptable methods of contraception. These procedures and laboratory test results must be confirmed by physical examination, by subject recall of specific date and hospital/facility of procedure, or by medical documentation of said procedure.

**Exclusion Criteria:**
Subjects meeting any of the following criteria will be excluded from the study (subject-reported unless otherwise indicated):

1. Subjects who are having any current workup of undiagnosed illness within the last 8 weeks, which is either subject-reported or has been clinician-assessed, that could lead to a new condition diagnosis.
2. Participation in research involving receipt of an investigational product (drug/biologic/device) within 45 days prior to first study vaccination.
3. History of a confirmed diagnosis of SARS or history of a confirmed diagnosis of COVID-19 disease resulting in medical intervention.

   **NOTE**: Subjects with a history of confirmed COVID-19 disease resulting in mild symptoms are allowed. Mild symptoms are defined as those in which treatment was symptom relief and not systemic or respiratory supportive care (eg, IV hydration, oxygen, nebulizer), with OTC medications being allowed.
4. Received influenza vaccination within 14 days prior to first study vaccination, or any other vaccine within 4 weeks prior to first study vaccination.
5. Have clinically significant chronic cardiovascular, endocrine, gastrointestinal/ hepatic, renal, neurological, respiratory, or other medical disorders not excluded by other exclusion criteria, that are assessed by the Investigator as being clinically unstable within the prior 4 weeks evidenced by: a) hospitalization for the condition, including day surgical interventions, b) new significant organ function deterioration, c) needing addition of new treatments or major dose adjustments of current treatments.
6. Diabetes mellitus requiring insulin therapy (either type 1 or type 2 diabetes mellitus).
7. Chronic obstructive pulmonary disease with a history of an acute exacerbation of any severity in the prior year.
8. Any history of congestive heart failure.
9. Any history of chronic kidney disease (the presence of impaired or reduced kidney function lasting at least 3 months). Clinical validation of potential cases of chronic kidney disease should be conducted.
10. Evidence of unstable coronary artery disease as manifested by cardiac intervention, addition of new cardiac medications for control of symptoms, or unstable angina in the past 3 months.

11. History of chronic neurological disorders that have required prior specialist physician review for diagnosis and management (such as multiple sclerosis, dementia, transient ischemic attacks, Parkinson's disease, degenerative neurological conditions, neuropathy, and epilepsy) or a history of stroke or previous neurological disorder within 12 months with residual symptoms. Subjects with a history of migraine or chronic headaches or nerve root compression that have been stable on treatment for the last 4 weeks are not excluded.
12. Any autoimmune or immunodeficiency disease/condition (iatrogenic or congenital).

    **NOTE**: Stable endocrine disorders that have a confirmed autoimmune etiology (eg, thyroid, pancreatic) are allowed.
13. Chronic administration (defined as more than 14 continuous days) of immunosuppressants, systemic glucocorticosteroids reaching an immunosuppressive dose, or other immune-modifying drugs within 90 days prior to first study vaccination.

    **NOTE**: An immunosuppressant dose of glucocorticoid is defined as a systemic dose ≥10 mg of prednisone per day or equivalent. The use of topical, inhaled, and nasal glucocorticoids will be permitted if other chronic disease conditions are not exclusionary.
14. Received immunoglobulin, blood-derived products, or other immunosuppressant drugs within 90 days prior to first study vaccination.
15. Known disturbance of coagulation (iatrogenic or congenital).

    **NOTE**: The use of ≤325 mg of aspirin per day as prophylaxis is permitted, but the use of other platelet aggregation inhibitors, thrombin inhibitors, Factor Xa inhibitors, or warfarin derivatives is exclusionary, due to potential bleeding following IM injection.
16. Active cancer (malignancy) within 5 years prior to first study vaccination (with the exception of adequately treated non-melanomatous skin carcinoma, at the discretion of the investigator).
17. Any known allergies to products contained in the investigational product or latex allergy.
18. Women who are breastfeeding or who plan to become pregnant during the study.
19. History of alcohol abuse or drug addiction within one year prior to the first study vaccination.
20. Any condition that, in the opinion of the investigator, would pose a health risk to the subject if enrolled or could interfere with evaluation of the study vaccine or interpretation of study results (including neurologic or psychiatric conditions deemed likely to impair the quality of safety reporting).
21. Study team member or first-degree relative of any study team member (inclusive of sponsor, PPD, and site personnel involved in the study).

**Other Considerations:**

Subjects meeting any of the following criteria may have planned study vaccination deferred for a later date, but these criteria are not exclusionary for study enrollment. The sponsor may advise sites of dates after which potential subjects who have been deferred need to have been enrolled, due to the need to rapidly enroll subjects in this study to allow rapid reporting of results to allow initiation of future studies.

- Respiratory symptoms in the past 3 days (ie, cough, sore throat, difficulty breathing). Subject may be vaccinated once all symptoms have been resolved for >3 days. Out of window vaccination is allowed for this reason.
- Temperature of >38°C within 24 hours of planned vaccination (site measured or subject measured). Subject may be vaccinated once the fever has resolved and there has not been any temperature measured as being >38°C for >3 days. Out of window vaccination is allowed for this reason.

  **NOTE:** Screening for COVID-19 disease symptoms may be indicated for either of the above-mentioned reasons or if COVID-19 disease is suspected based on potential exposure to SARS-CoV-2 infection through either close contacts or based on local epidemiology. In such a case, subjects should also have study samples collection for qualitative PCR testing on the day of any subsequent study vaccination, but the results of the qualitative PCR test are not needed before study vaccination can be given for the first or subsequent doses. Any subjects with new positive PCR-confirmed SARS-CoV-2 infections occurring from screening and prior to the end of immunogenicity assessments will be removed from applicable immunogenicity analyses as defined in the statistical analysis plan.
- Any acute illness (cardiovascular, endocrine, gastrointestinal/ hepatic, renal, neurological, respiratory, or other medical disorders) that is actively causing symptoms that could, in the opinion of the investigator, impact the assessment of reactogenicity or other study assessments. Subject may be vaccinated once symptoms have resolved or are stabilized for >3 days. Out of window vaccination is allowed for this reason.
- Immunization with any vaccine within 14 days prior to vaccination. Out of window vaccination is allowed for this reason.
- Blood pressure that exceeds the United States Eighth Joint National Committee control levels (by age) prior to vaccination (BP is 150/90 mm Hg or higher in adults 60 years and older, or 140/90 mm Hg or higher in adults younger than 60 years). Repeated BP testing following a high reading can occur up to a total of a further 2 times during the same visit after a pause of not less than 5 minutes between measures, and if the reading is then within the permitted range the subject may be randomized and vaccinated. Out of window vaccination is allowed for this reason if subjects are deferred on any planned vaccination day due to transient abnormal BP readings.

**STUDY TREATMENTS:**
Study vaccinations will comprise up to 3 IM injections (Day 0 and Day 21 [priming doses] and Day 189 [booster – active or placebo] in Treatment Groups A, D, and E, and up to 4 IM injections (Day 0 and Day 21 [priming doses] and Day 189 [6-month booster – active or placebo] and Day 357 [booster – active or placebo]) in Treatment Groups B and C, ideally in alternating deltoids for priming doses, with the study vaccine assigned in an full dose injection volume of approximately 0.5 mL. Dose levels of SARS-CoV-2 rS in Part 2 will be 5 µg and up to 25 µg, with 50 µg Matrix-M adjuvant.

**STUDY PROCEDURES:**

**Safety Assessments:**

Safety assessments will include monitoring and recording of solicited (local and systemic reactogenicity events) and unsolicited AEs; MAAEs; AESI; SAEs; vital sign measurements; and physical examination findings Recording of solicited and unsolicited AEs may be conducted by electronic data capture/reporting. Monitoring for PIMMC and AESI specific to potential disease enhancement for COVID-19 will be continued as noted for Part 1 of study.

**Immunogenicity Assessments:**

Blood samples for immunogenicity assessments will be collected before vaccination and at selected early time points following vaccination. Immune measurements (ELISA) will be conducted on serum (IgG and ACE2 receptor inhibition) for SARS-CoV-2 rS protein antigen(s). Neutralization testing and cell-mediated immunity following in vitro stimulation of PBMCs will occur at selected time points for a subset of subjects. In the case of the cell-mediated immunity testing subset, a sampling scheme based on visit, age strata, or other factors (such as location in relation to PBMC processing capacity) may be utilized. Additional testing will occur with further assay development. Aliquots of all collected samples from this study may be retained for the stated purposes for a maximum of 25 years (starting from the date at which the last subject had the last study visit), unless local rules, regulations, or guidelines require different timeframes or different procedures, in accordance with subject consent. Based on the emergence of new variants, including B.1.1.7, B.1.351, P.1 and others, immunogenicity testing may be performed for these new variants as well as against the original prototype Wuhan strain.

**COVID-19 Disease Assessments (Efficacy):**

Subjects will be instructed on self-collection using a nasal mid-turbinate sample approach (depending on assay availability and appropriate validation) and should demonstrate competency during an early clinic visit. An ePRO system will be employed at US sites for monitoring/documenting potential COVID-19 disease symptoms based on the occurrence of symptoms from a standard list (see Table S2-4), with subjects being queried every 14 days. Subjects may also trigger collection between the 14-day query periods if symptoms are reported spontaneously and conform to the algorithm to trigger self-collection. Self-collection should occur within 3 days of symptom onset (if possible). Subjects will also be notified through the electronic patient diary to self-collect specimens every 28 days if symptom free to assess for asymptomatic carrier status.

Should a subject be admitted to the hospital or COVID-19 intensive care ward and sample self-collection is unavailable, then a local public health or hospital test will be taken as a valid result. All PCR samples taken through the study will be sent to a prespecified central laboratory where a validated PCR test will be performed. When a subject actively initiates reporting of qualifying symptoms for a potential COVID disease within the electronic patient diary, follow-up patient-reported outcomes will be collected using the FLU-PRO instrument for up to 10 days, and the duration of resolved illnesses (number of days from the start of illness symptoms until the day at which health returned to normal). Exploratory analyses may also be conducted looking at the time course and maximal values of FLU-PRO total and domain level scores during an assessed illness episode, and how the proportion of subjects with maximal scores relate to COVID-19 study-defined illness severity categories. Should a medical visit be warranted based on symptomatology (and allowed via local isolation guidance), such a visit may occur using telemedicine, home visitation, or clinic visit.

Subjects will be notified of new positive PCR-confirmed SARS-CoV-2 infection status due to requirements of self-isolation and potential transmission. Since self-collection PCR-confirmation assay methodologies that are likely to be used in this study are not expected to be licensed for diagnostic purposes, and likely relative delays in the availability of results due to transportation and testing timelines, collection of study samples for PCR-confirmation do not replace the need for subjects to also be tested and followed up through public health testing and management processes that are in place locally if they meet criteria for local testing.

To further describe the epidemiologic evolution of the pandemic and potential effect of vaccination, assessment of SARS-CoV-2 by PCR testing based on routine screening without symptomatology will be done by nasal mid-turbinate sample self-collection from Day 28 through approximately 6 months.

Asymptomatic infection, period of transmission, and other defined infection parameters will be assessed along with severity and progression of disease.

- **Asymptomatic infection** will be defined as a positive PCR-confirmed SARS-CoV-2 infection with no symptoms (regardless of past positivity) in the 7 days prior to self-collection.
- **Primary infection** will be defined as the first positive PCR-confirmed SARS-CoV-2 infection regardless of symptoms.
- **Primary symptomatic infection** will be defined as the first positive PCR-confirmed SARS-CoV-2 infection that is triggered by the symptomatic algorithm. A symptomatic infection will initiate further evaluation of physical status with severity scoring applied and monitoring of duration.
- **Period of transmission** for any subject (eg, transmission potential) is the period from a positive PCR test until the time of the first negative PCR test that follows a positive PCR test (if follow-up PCR test results are available for that subject).
- **Duration of resolved illness episodes** (ie, number of days from the start of illness symptoms for illnesses followed up by FLU-PRO assessments until the day at which health returned to normal as reported in the electronic diary) classified by COVID-19 disease severity (Table S2-4). Duration will not be calculated for asymptomatic infections or illness episodes in which the subject's health has not returned to normal at the time of the data cut.

**STATISTICAL ANALYSIS PLANS:**
**Sample Size:**
The decision on the choice of formulation and dosing regimen will be made based on the totality of the immunogenicity and safety data rather than any individual measurement. No multiplicity adjustment will be made for this early phase study where multiple treatment groups and endpoints are being evaluated.

With 150 subjects in each treatment group, there is a greater than 99.9% probability to observe at least 1 subject with an AE if the true incidence of the AE is 5% and a 77.9% probability if the true incidence of the AE is 1%.

**Analysis Sets:**

The All Screened Subjects Analysis Set will include all subjects who sign the ICF.

The Intent-to-Treat (ITT) Analysis Set will include all subjects who are randomized, regardless of protocol violations or missing data.

The Full Analysis Set will include all ITT subjects who receive at least 1 dose of study vaccine (SARS-CoV-2 or placebo). The Full Analysis Set will be used for supportive analyses. Subjects will be analyzed according to the randomized treatment assignment.

The Safety Analysis Set will include all subjects who receive at least 1 dose of study vaccine (SARS-CoV-2 rS or placebo). Subjects will be analyzed according to the vaccine actually received. Actual treatment received will be assumed to correspond with randomized treatment assignment except in cases where a site reports having used incorrect investigational product during vaccine administration.

The Per-Protocol (PP) Analysis Set will be determined for each study visit and will include all subjects who receive the initial dose of study vaccine (SARS-CoV-2 rS or placebo) for all analyses through Day 21, all subjects who receive vaccine doses at both Day 0 and Day 21 for all analyses starting at Day 28, all subjects who receive vaccine doses at Days 0, 21, and 189 for all analyses starting at Day 217, and all subjects who receive vaccine doses at Days, 0, 21, 189, and 357 for all analyses starting at Day 371, have at least a baseline and 1 post-baseline serum sample IgG result available at the corresponding study visit, have no major protocol violations (eg, having randomized treatment assignment unblinded due to medical emergency, having taken prohibited medication on-study such as chronic systemic glucocorticoids, having received a vaccination containing investigational product that differs from randomized treatment assignment) that impact immunogenicity response at the corresponding study visit. The review and determination for exclusion from the PP Analysis Set will be carried out in a blinded fashion by a study clinician prior to unblinding for each analysis based on all available information from either the locked database or from a database freeze, depending on the SAP defined analysis. All subjects in the PP Analysis Set will be analyzed according to the randomized treatment assignment. Any subject who is SARS-CoV-2 positive by qualitative PCR testing from screening and prior to the immunogenicity assessment for a particular time point will be excluded from the PP Analysis Set from that time point onward. The protocol violations that impact the immunogenicity response and impact upon the PP Analysis Set are detailed in the study deviation rules document.

The PP-Immunogenicity PBMC Subset will include all subjects in the PP Analysis Set but further restricted to subjects who had blood samples taken and PBMCs harvested for analysis of cell-mediated immunity.

The PP-Immunogenicity Analysis Set will include all subjects who received their full vaccination schedule (ie, Day 0 and Day 21, or Days 0, 21, and Day 189, or Days 0, 21, 189, and Day 357), have at least a baseline and a Day 35 serum sample IgG result available for analysis, have no major protocol violations up to and including Day 35 that impact immunogenicity response, and no evidence of positive serology at baseline (ie, hepatitis B, hepatitis C, or HIV). The PP-Immunogenicity Analysis Set will be used for the primary immunogenicity analysis (ie, serum IgG antibody levels specific for the SARS-CoV-2 rS protein antigen[s] as detected by ELISA using GMT or SCR for the 2-dose regimens at Day 35 regardless of baseline immune status). If the number of subjects in the PP-Immunogenicity and PP Analysis Set differ (defined as the difference divided by the total number of subjects in the given PP Analysis Set) by more than 10%, supportive analyses of immunogenicity may be conducted using the PP Analysis Set.

The PP-Efficacy Analysis Set will be the PP Analysis Set further restricted to subjects who received their full vaccination schedule (ie, Day 0 and Day 21). All subjects in the PP-Efficacy Analysis Set will be analyzed according to the randomized treatment assignment.

A listing of analysis sets (with reasons for exclusion, if applicable) will be provided.

**Safety Analyses:**
Numbers and percentages (with 95% CIs based on the Clopper-Pearson method) of subjects with solicited local and systemic AEs through 7 days after each vaccination will be summarized by treatment group and the maximum toxicity grade over 7 days after each vaccination. The duration of solicited local and systemic AEs after each vaccination will also be summarized by treatment group. If more than 10% of total subjects are seropositive to SARS-CoV-2 infection at baseline, exploratory analyses by SARS-CoV-2 positivity at Day 0 may be performed.

Unsolicited AEs will be coded by preferred term and system organ class using the latest version of MedDRA and summarized by treatment group as well as by severity and relationship to study vaccine. Unsolicited AEs from the first vaccination until before the second vaccination, from the second vaccination through 35 days after first vaccination, and from boost until 28 days after boost vaccination; all MAAEs through 217 days after first vaccination; and any MAAE related to vaccine, SAE, or AESI through end of study will be listed separately and summarized by treatment group and age strata (18-59, 60-84 years). Adverse events of special interest associated with PIMMC and COVID-19 disease potential exacerbation will be listed separately.

Vital sign measurements will be summarized by treatment group at each time point using descriptive statistics. Vital sign toxicity grading will be derived on day of vaccination.

Concomitant medications will be summarized by treatment group and preferred drug name as coded using the World Health Organization drug dictionary.

**Immunogenicity Analyses:**
The primary immunogenicity analyses will be performed using the PP Immunogenicity Analysis Set (which is restricted relative to subjects in the PP Analysis Set who received their full vaccination schedule and have a Day 35 sample available for analysis regardless of baseline SARS-CoV-2 IgG serostatus), excluding those subjects with evidence of positive serology at baseline (ie, hepatitis B, hepatitis C, or HIV). Subgroup analyses of the primary immunogenicity analyses by SARS-CoV-2 IgG serostatus at baseline (as measured by

in-house SARS-CoV-2 IgG assay at the Novavax central immunology laboratory) will be undertaken.

If the number of subjects in the PP-Immunogenicity and PP Analysis Set differ (defined as the difference divided by the total number of subjects in the PP Analysis Set) by more than 10%, supportive analyses of immunogenicity may be conducted using the PP Analysis Set.

For the serum IgG antibody levels and ACE2 receptor binding inhibition specific for the SARS-CoV-2 rS protein antigen(s) as detected by ELISA, the geometric mean, the GMFR compared to the baseline (Day 0) and the SCR (proportion of subjects with ≥ 4-fold rises in ELISA units) at each post-vaccination study visit (except the Day 7 and Day 28 visits for the PBMC subset), and the GMFR comparing post- (Day 217) to pre- (Day 189), and post- (Day 371) to pre- (Day 357) along with 95% CI will be summarized by treatment group by visit. Both age strata (18-59, 60-84 years) and naïve versus non-naïve subjects at baseline will be analyzed. Definitions for naïve versus non-naïve subjects at baseline will be defined in the SAP.

A subset of subjects will have immunogenicity analyses performed using a wild-type SARS-CoV-2 neutralization assay at a minimum of the time points baseline (Day 0), Day 35, and Day 217). More details for these neutralization assays will be documented in the SAP.

A subset of subjects will have neutralization assays, IgG, and hACE-2 testing for both the Wuhan and the B.1.351 strain at Days 189, 217, 357, 371, and 546. Immunogenicity assessments for other variants may also be performed.

The 95% CI will be calculated based on the t-distribution of the log-transformed values for geometric means or GMFRs, then back transformed to the original scale for presentation. The SCR (proportion of subjects with ≥4-fold rises in ELISA units) along with 95% CIs based on the Clopper-Pearson method will be summarized by treatment group at each post-vaccination study visit. An ANCOVA model will be constructed at each post-vaccination study visit on the log-transformed titer, including the treatment group as a fixed effect and the baseline log-transformed titer as a covariate. Comparisons of selected treatment groups will be performed within each visit. Additional covariates such as site and age strata (18-59, 60-84 years) will be included as covariates in the ANCOVA model used to analyze serum IgG antibody levels, serum ACE-2 receptor binding inhibition, and neutralization assay results. The difference in the SCR between selected treatment groups along with 95% CIs within each visit will be calculated using the method of Miettinen and Nurminen.

Similar summaries will be generated for the other immunogenicity endpoints.

Cell-mediated immunity will be measured using select cytokines (eg, IL-2, IL-4, IL-5, IL-6, IL-13, TNFα, INFγ) in harvested PBMC cells using flow cytometry and will be summarized by treatment group, overall and by age strata (18-59, 60-84 years) (subset of subjects).

**Efficacy Analyses:**
Numbers and percentages (with 95% CIs based on the Clopper-Pearson method) of subjects with occurrence of SARS-CoV-2 positivity and classified by COVID-19 disease severity will be summarized by treatment group, overall and by age strata (18-59, 60-84 years) by those receiving SARS-CoV-2 rS compared to placebo. If frequent clinical PCR-confirmed SARS-CoV-2 infections occur during the study follow-up period, vaccine efficacy

assessments for primary vaccination for treatment groups compared to placebo may be generated. The criteria to determine whether this will occur will be documented in the SAP.

Vaccine Efficacy (VE) is defined as VE (%) = (1 – RR) x 100, where RR = relative risk of incidence rates between the 2 treatment groups (SARS-CoV-2 rS/Placebo). The interim and final analyses will be carried out at the one-sided Type I error rate of 0.025 overall. A two-sided 95% CI for the VE for each primary endpoint will accompany the point estimate. The estimated RR and its CI will be derived using Poisson regression with robust error variance [Zou, 2004]. The explanatory variables in the model will include treatment group and age group. The dependent variable will be the incidence rate of the endpoint of interest. The robust error variances will be estimated using the repeated statement and the subject identifier. The Poisson distribution will be used with a logarithmic link function.

The duration of resolved illness episodes (number of days from the start of illness symptoms for illnesses followed up by FLU-PRO assessments, until the day at which health returned to normal as reported in the electronic diary) and classified by COVID-19 disease severity will be summarized by treatment group, overall and by age strata (18-59, 60-84 years).

**Exploratory Analyses:**

Daily FLU-PRO scores (for total score and/or domain subgroup scores) following subject reported SARS-CoV-2 positive illnesses may be summarized by treatment group and age subgroup using descriptive statistics. Maximal FLU-PRO scores during the daily follow-up for an illness episode (for total score and/or domain subgroup scores) by treatment group and age subgroup may also be assessed by study defined COVID-19 illness severity categories. Further details of exploratory analyses related to FLU-PRO scores will be defined in the SAP.

**Interim Analyses:**

A primary database lock will occur following Day 35 data for all primary endpoints and selected secondary endpoints defined in the SAP, to allow assessment of treatment group responses in the Part 2 study, to inform decisions around recommended dose regimens by age strata, and to facilitate the initiation of the Phase 3 efficacy trial.

However, earlier database freezes may occur to potentially allow earlier initiation of Phase 3 efficacy trials in consultation with the SMC and regulatory agencies. These database freezes may be based on data accrued in either Australia or the United States or accrued across the 2 countries combined (depending on the timing of availability of results to allow assessments to occur).

Database freezes may occur when approximately 50 subjects for either age group (18-59, 60-84 years) of each of the 2 first dose formulations (5 µg + 50 µg and 25 µg + 50 µg) have: a) reached Day 21 for safety follow-up; and b) baseline (Day 0) and Day 21 serum IgG antibody levels are available for immunogenicity follow-up. Data for database freezes will be based on partially cleaned and verified data. Immunogenicity data at these reviews may contain neutralization data or other immunogenicity assessments that are available at the time of the database freeze.

Database freezes may also occur to assess the Day 35 data when approximately 50 subjects for either age group (18-59, 60-84 years) in vaccine groups B and D have: a) reached Day 35 for safety follow-up; and b) baseline (Day 0) and Day 35 serum IgG antibody levels are available for immunogenicity follow-up. Data for database freezes will be based on partially cleaned and verified data. Immunogenicity data at these reviews may contain neutralization data or other immunogenicity assessments that are available at the time of the database freeze.

Subsequent database freezes will occur at Days 105 and 217 (all treatment groups), and at Day 371 (Treatment Groups B and C) for analysis of selected primary and/or secondary endpoints with a final database lock at Day 546 (EOS). In addition, planned analyses for COVID-19 endpoints will be included in all scheduled SMC reviews to allow review of potential vaccine efficacy (if sufficient COVID-19 events occur in the jurisdiction(s) where the study is conducted) or safety concerns. The planned primary analyses will be performed by an unblinded biostatistics and programming team. The sponsor biostatistics and programming team may be provided with the immunogenicity data set with a limited number of variables and dummy subject identifiers if sponsor analyses are planned, such that sponsor biostatistics and programming team remain blinded at the subject level. The variables to be included in this blinded immunogenicity data transfer are the dummy subject identifiers, treatment group assignments, visit numbers, and assay identifiers, and results only with the dataset subset by the unblinded team to only those subjects meeting per-protocol population criteria for the planned analyses. Only group level unblinded summaries will be generated at planned analyses and subject level treatment assignment will not be released until the sponsor is unblinded at the subject level, or regulatory authorities request rolling submission of data from earlier study time points (which would require sponsor subject level unblinding). To the extent possible based on the availability of authorized COVID-19 vaccines, investigator/site and CRO subject level blinding will be maintained until the study is completed. Planned analyses will be documented in the SAP prior to initiating that specific analysis.

**DATE OF PROTOCOL:** 03 May 2021

### 8. INTRODUCTION

#### 8.1 BACKGROUND

Novavax, Inc. is developing a recombinant vaccine adjuvanted with the saponin-based Matrix-M™ (previously referred to as Matrix-M) for the prevention of disease caused by the SARS-CoV-2 virus. The newly described coronavirus' genetic relationship with the 2002-2003 SARS-CoV has resulted in adoption of the name SARS-CoV-2, with the associated disease being referred to as coronavirus disease 2019 (COVID-19). SARS-CoV-2 recombinant spike (S) protein nanoparticle vaccine (SARS-CoV-2 rS) is constructed from the full length wild-type SARS-CoV-2 S glycoprotein based upon the GenBank gene sequence MN908947, nucleotides 21563-25384. The S protein is a type 1 trimeric glycoprotein of 1,273 amino acids that is produced as an inactive S0 precursor. The S-gene was codon optimized for expression in Sf9 insect cells. SARS-CoV-2 rS is intended for administration with or without Matrix-M adjuvant, which is a saponin-based adjuvant that has been shown to enhance the immunogenicity of nanoparticle vaccines in nonclinical and clinical studies. Reference the current version of the Matrix-M adjuvant safety supplement to the IB for additional details.

Further details on the study vaccine can be found in the IB (Novavax 2020).

#### 8.2 RATIONALE FOR STUDY

The purpose of this 2-Part, Phase 1/2, first-in-human, randomized, placebo-controlled, observer-blinded study is to evaluate the safety and immunogenicity of SARS-CoV-2 rS with or without Matrix-M adjuvant in healthy subjects. The primary objectives of Part 2 of this study are to identify the optimal dose based on immune response (IgG antibody to SARS-CoV-2 rS) and whether baseline immune status has an impact; and to accumulate a safety experience for the candidate vaccine in healthy adult subjects based on solicited short-term reactogenicity across a broad age spectrum (by toxicity grade) and by AE profile for primary vaccination. The information provided by the Phase 2 portion of this study will inform progression of the vaccine and identify dose(s) to take forward in an emergency use authorization setting and/or for Phase 3 efficacy or effectiveness trial(s).

#### 8.3 RATIONALE FOR DOSE SELECTION

The proposed dose levels of SARS-CoV-2 rS to be evaluated in this study are based on several data sources, including nonclinical studies; nonclinical and clinical studies with other Novavax, Inc. baculovirus-Sf9-produced nanoparticle vaccines including safety data in over

14,700 subjects from clinical studies with EBOV, RSF F, and influenza vaccines; and the results from Part 1 of this study. For details on nonclinical studies of SARS-CoV-2 rS and for details on the ongoing clinical studies with Novavax, Inc. baculovirus-Sf9-produced nanoparticle vaccines, please refer to the IB (Novavax 2020).

The proposed dose level of Matrix-M adjuvant to be evaluated in this study is based on the fully-analyzed human experience to date with Matrix-M adjuvants, which is confined to adults who have received 1- to 3-dose series of IM doses of 25 to 75 µg. Further details regarding the Matrix-M adjuvant, including safety data from over 4200 subjects in clinical studies with EBOV, RSV F, malaria, rabies, herpes simplex virus, and influenza vaccines with Matrix-M, are provided in the current version of the Matrix-M adjuvant safety supplement to the IB (Novavax 2020). It is anticipated that Matrix-M adjuvant will be required to provide a robust immunological response with appropriate cell mediated immune pathway activation.

### 9. STUDY OBJECTIVES AND ENDPOINTS

#### 9.1 STUDY OBJECTIVES

The primary objectives are:

- To identify the optimal dose across age strata based on immune response (IgG antibody to SARS-CoV-2 rS) at Day 35 and whether baseline immune status has an impact.
- To accumulate a safety experience for the candidate vaccine in healthy adult subjects based on solicited short-term reactogenicity across a broad age spectrum (by toxicity grade) and by AE profile for primary vaccination (through Day 35).
- To identify dose(s) to potentially take forward in an emergency use authorization setting and/or for Phase 3 efficacy or effectiveness trial(s).

The secondary objectives are:

- To assess if the single-dose regimens can provide similar (or adequate) priming immune response at Day 35 (compared to the two-dose regimens and to placebo) and whether baseline immune status alters such response (IgG antibody to SARS-CoV-2 rS).
- To define the optimal dosing regimen in subjects who are naïve and those with pre-existing antibodies to SARS-CoV-2 (if enough subjects are identified with pre-existing antibodies) as assessed by the immune response (IgG antibody to

SARS-CoV-2 rS and angiotensin-converting enzyme 2 [ACE2] receptor binding inhibition) to the various regimens at Day 21 (post first dose), Day 35 (post second dose), and Day 217. Optimal dosing regimen to be assessed across full age spectrum and by age strata (18-59, 60-84 years).

- To describe the amplitude, kinetics, and durability of immune responses to the various regimens in terms of ELISA units of serum IgG antibodies to SARS-CoV-2 rS and titers of ACE2 receptor binding inhibition at selected time points and relative to whether subjects had pre-existing antibodies to SARS-CoV-2. To include reverse cumulative distribution curves.

- To describe the immune responses to the various regimens in terms of titers of neutralizing antibody at selected time points and relative to whether subjects had pre-existing antibodies to SARS-CoV-2 (subset of subjects). Optimal dosing regimen to be assessed across full age spectrum and by age strata (18-59, 60-84 years).

- To assess immune responses to the various regimens at 6 months and whether a boost at 6 months for a subset of the subjects enrolled in the 5-µg dose regimens (Treatment Groups B and C) induces immune memory and is beneficial to maintain immune response in terms of IgG and neutralizing antibodies to SARS-CoV-2 rS and ACE2 receptor binding inhibition.

- To assess immune responses to the various regimens at 12 months (all treatment groups) and whether a boost at 12 months for subjects enrolled in the 5-µg dose regimens (Treatment Groups B and C) induces immune memory and is beneficial to maintain immune responses in terms of IgG and neutralizing antibodies to SARS-CoV-2 rS and ACE2 receptor binding inhibition.

- To assess overall safety through 35 days after prime vaccination is initiated (1 or 2 doses) for all AEs; from 6-month boost (Day 189) through 28 days after 6-month boost (Day 217); and, for participants in Treatment Groups B and C, from 12-month boost (Day 357) through 28 days after 12-month boost (Day 385) (Table 10-4) for all AEs; and through the EOS for any MAAE attributed to vaccine, AESI, or SAE.

- To assess for occurrence and duration of COVID-19 disease as measured by PCR following subject-reported symptoms and to assess disease severity (virologically confirmed, mild, moderate, severe) and duration by patient-reported outcomes (eg, InFLUenza Patient-Reported Outcome [FLU-PRO©]) in those immunized with SARS-CoV-2 rS compared to placebo.

- To assess cell-mediated response: Type 1 T helper (Th1) or Type 2 T helper (Th2) predominance by various vaccine regimens (eg, IL-2, IL-4, IL-5, IL-6, IL-13, TNFα, INFγ using flow cytometry, ELISpot, or other system) in whole blood and/or harvested PBMC cells (in response to in vitro stimulation with SARS-CoV-2 rS protein) (subset of subjects).

Exploratory objectives are:

- To explore application of the FLU-PRO instrument to categorize COVID-19 severity, for maximal illness severity assessment, and the time course and severity of clinical symptomatology for COVID-19 cases for treatment groups.

- To utilize additional assays (current or to be developed) to best characterize the immune response for future vaccine development needs.

- To assess overall safety through 35 days after prime vaccination is initiated (1 or 2 doses) for solicited AEs; by SARS-CoV-2 positivity at Day 0, if greater than 10% of total subjects are SARS-CoV-2 positive at Day 0.

- To assess immune responses to the various regimens at 12 months and whether a boost at 12 months for subjects enrolled in the 5-µg dose regimens (Treatment Groups B and C) induces immune memory and is beneficial to maintain immune responses in terms of IgG and neutralizing antibodies to SARS-CoV-2 rS and ACE2 receptor binding inhibition for new variants, including, but not limited to, the South Africa variant B.1.351.

#### 9.2 STUDY ENDPOINTS

The primary endpoints are comparisons of treatment regimens:

- Serum IgG antibody levels specific for the SARS-CoV-2 rS protein antigen(s) as detected by ELISA using GMT or SCR for the two-dose regimens by dose at Day 35 regardless of baseline immune status and stratified by baseline immune status. Derived/calculated endpoints based on these data will include geometric mean ELISA units (GMEUs), geometric mean fold rise (GMFR), and SCR.

    - Two-dose regimens by dose compared to placebo

    **SCR** is defined as the percentage of subjects with a post-vaccination titer ≥ 4-fold.

    **Positive baseline status** (+/-) using GMT and/or positive PCR at baseline.

- Numbers and percentages (with 95% CIs) of subjects with solicited AEs (local, systemic) for 7 days following each primary vaccination (Days 0 and 21) by severity score, duration, and peak intensity. Unsolicited AEs (eg, treatment-emergent, serious, suspected unexpected serious, those of special interest, MAAEs) through 35 days by MedDRA classification, severity score, and relatedness.

The secondary endpoints are comparisons of treatment regimens:

- Serum IgG antibody levels specific for the SARS-CoV-2 rS protein antigen(s) as detected by ELISA using GMT or SCR (≥ 4-fold change) for the single-dose regimens compared to the two-dose regimens and to placebo at Day 21 and Day 35 regardless of baseline immune status and stratified by baseline immune status. Derived/calculated endpoints based on these data will include GMEUs, GMFR, and SCR.

- Serum IgG antibody levels specific for the SARS-CoV-2 rS protein antigen as detected by ELISA, described across study time points with derived/calculated endpoints to include GMEUs, GMFR, and SCR (≥ 4-fold change) for the single-dose regimens compared to the two-dose regimens and to placebo, stratified by baseline immune response.

- Epitope-specific immune responses to the SARS-CoV-2 rS protein receptor binding domain measured by serum titers in an ACE2 receptor binding inhibition assay to include GMT or concentration, GMFR, and SCR (≥ 4-fold change) for the single-dose regimens compared to the two-dose regimens and to placebo.

- Neutralizing antibody activity at Days 35, 217, and at 357 for all treatment groups, and additionally at Days 371 and 546 for Treatment groups B and C relative to baseline (Table 10-5) in a subset of subjects by absolute titers and change from baseline, including the SCR (≥ 4-fold change). Analysis to include subjects by treatment group, by age (18-59, 60-84 years) and relative to whether subjects had pre-existing antibodies to SARS-CoV-2. A sampling scheme to identify a subset of such subjects will be deployed.

- Serum IgG antibody levels specific for the SARS-CoV-2 rS protein antigen(s) as detected by ELISA using GMT or GMFR at Days 189, 217, and 357 for all treatment groups and additionally at Days 371 and 546 for Treatment Groups B and C (Table 10-5) for boosting assessment with either placebo or active boost.

- All MAAEs through Day 217, and then related MAAEs until EOS, and all AESIs, or SAEs through the EOS by MedDRA classification, severity score, and relatedness.

- Vital sign measurement before vaccination and as clinically needed during the 30-minute post-vaccination observation period. Vital sign measurements at all other time points to be classified by descriptive statistics (eg, mean, median, SD) by visit.

- Percentage of subjects with SARS-CoV-2 positivity as diagnosed by qualitative PCR following COVID-19 symptoms assessment from Day 28 through 6 months with severity classification, overall and by age strata (18-59, 60-84 years). If frequent clinical PCR-confirmed SARS-CoV-2 infections occur during the study follow-up period, vaccine efficacy assessments for primary vaccination for treatment groups compared to placebo may be generated. The criteria to determine whether this will occur will be documented in the SAP.

- Assessment of SARS-CoV-2 by qualitative PCR based on routine screening by nasal mid-turbinate sample self-collection from Day 28 through 6 months without symptomatology to further describe epidemiologic evolution of the pandemic and potential effect of vaccination.

- Assessment of cell-mediated (Th1/Th2) pathways as measured by whole blood (flow cytometry) and/or in vitro PBMC stimulation (eg, ELISpot, cytokine staining) with SARS-CoV-2 rS protein(s) as measured on Days 0, 7, and 28.

Exploratory endpoints are:

- Assessment of immune responses to the various regimens at 6 and 12 months for all treatment groups and at 18 months for Treatment Groups B and C and whether a boost at 6 months (all treatment groups) and again at 12 months (Treatment Groups B and C) induces immune memory and is beneficial to maintain immune responses to SARS-CoV-2 rS in terms of IgG and neutralizing antibodies and ACE2 receptor binding inhibition for new variants, including, but not limited to, the South Africa variant B.1.351.

- Assessment of COVID-19 severity and the time course of symptom scores using the FLU-PRO instrument for up to 10 days following a qualifying illness episode, to assess the potential use of patient-reported symptom-based severity criteria for application in future clinical endpoint studies, with assessment of maximal severity of illness episodes and the relationship between FLU-PRO measures and COVID-19 clinically defined severity categories.

- Any additional assays to measure immune response, protection, or potential safety signals.

- Numbers and percentages (with 95% CIs) of subjects with solicited AEs (local, systemic) for 7 days following each primary vaccination (Days 0 and 21) by severity score, duration, and peak intensity by SARS-CoV-2 positivity at Day 0, if greater than 10% of total subjects are SARS-CoV-2 positive at Day 0.

### 10. STUDY DESIGN

If adequate safety and desired immune responses are observed at interim analyses during Part 1, the study may be immediately extended, at the sponsor's discretion, to include Part 2. Only 1 construct for SARS-CoV-2 rS will be evaluated in Part 2.

Part 2 is a Phase 2, randomized, placebo-controlled, observer-blinded study to evaluate the safety and immunogenicity of a SARS-CoV-2 rS with Matrix-M™ adjuvant in male and female subjects. Subjects will be healthy adults based on medical history and physical examination. It is anticipated that Matrix-M adjuvant will be required for an adequate response in this population and for dose-sparing needs.

After signing the informed consent form (ICF), subjects may be screened within a window of up to 45 days; however, SARS-CoV-2 serostatus across the enrolling sites should not be expected to be more than 15% of the overall population will be SARS-CoV-2 positive at baseline. Subjects will be asked to provide consent for the use of samples for future testing for other viruses and/or sequencing of the SARS-CoV-2 in positive specimens or assay development specific to SARS-CoV-2 (or related variants).

Approximately 750 healthy male and female subjects between 18 and 84 years of age inclusive (≥ 18 years to < 85 years) will be randomized in each country, with approximately 50% of subjects overall in the study ≥ 60 years of age. Two-factor, 2-level stratification will be employed (ages 18-59 and 60-84; study site). Subjects who meet the criteria for study entry will initially be randomized in a 1:1:1:1:1 ratio to 1 of 5 vaccine groups (Table 10-1).

Up to approximately 1500 subjects could be enrolled for the 5 vaccine groups across the 2 countries to potentially mitigate the risk of enrollment in either country being delayed, or the expected availability of study data from either country being delayed due to the potential impact of the pandemic on feasibility of study vaccine delivery, possibilities for adequate specimen and data collection, and feasible specimen transportation. This flexibility would allow review of study data on approximately 500 subjects to facilitate initiation of Phase 3 clinical development as soon as possible.

For the Part 2 component of the study, following study initiation, enrollment of older adult subjects (≥ 60 to 84 years of age) will be paused when approximately 50 subjects in the older age group are enrolled in each of the 5 study vaccine groups (ie, approximately 250 older adult subjects in total enrolled subjects across all sites). The Safety Monitoring Committee (SMC) will then review reactogenicity data for both study age groups for 5 days following the first vaccine dose (Day 0 through Day 4, inclusive) when all subjects in the older adult group have accrued these 5 days of data. Enrollment of adult subjects (≥ 18 to 59 years of age) will not be paused during this SMC review unless general vaccination pause rules detailed in Section 13.1.5 are also met. Details of this review, and processes to restart enrollment of older adult subjects will be documented in the SMC Charter.

**Table 10-1 Treatment Groups as Originally Planned (Part 2)**

| Treatment Group | Number of Subjects | Day 0 | Day 21 (−1 to +3 days) | Day 189 (±15 days) |
|---|---|---|---|---|
| | | SARS-CoV-2 rS + Matrix-M Adjuvant | SARS-CoV-2 rS + Matrix-M Adjuvant | SARS-CoV-2 rS + Matrix-M Adjuvant |
| A | 150-300 | Placebo | Placebo | Placebo |
| B | 150-300 | 5 µg + 50 µg | 5 µg + 50 µg | Placebo |
| C | 150-300 | 5 µg + 50 µg | Placebo | 5 µg + 50 µg |
| D | 150-300 | 25 µg + 50 µg | 25 µg + 50 µg | Placebo |
| E | 150-300 | 25 µg + 50 µg | Placebo | 5 µg + 50 µg |

Note: The first dose represents the amount of antigen (SARS-CoV-2 rS) and the second dose represents the amount of adjuvant (Matrix-M). For example, 5 µg + 50 µg represents 5 µg SARS-CoV-2 rS + 50 µg Matrix-M adjuvant.

Subjects initially randomized to treatment groups B and C through Day 21 will be re-randomized at a 1:1 ratio to receive either a booster dose of vaccine (B2 and C2) or placebo (B1 or C1) at Day 189 (Table 10-2). This change to the boosting regime is being implemented in order to gain additional immunogenicity and safety data for subjects who receive 1 or 2 doses of 5 µg SARS-CoV-2 rS + 50 µg Matrix-M1 adjuvant on Days 0 and 21 (ie, the dose level intended for licensure). In addition, subjects in treatment group E who were initially scheduled to receive a booster dose of vaccine at Day 189 will now receive placebo. Treatment group E is no longer being boosted, as the 25-µg dose of SARS-CoV-2-rS is no longer being taken forward into later phase studies of the vaccine and the value of gathering additional boosting immunogenicity data for this group is considered minimal.

**Table 10-2 Treatment Groups, Re-randomized At Day 189 (Part 2)**

| Treatment Group | Number of Subjects | Day 0<br>SARS-CoV-2 rS + Matrix-M Adjuvant | Day 21 (−1 to +3 days)<br>SARS-CoV-2 rS + Matrix-M Adjuvant | Day 189 (±15 days)<br>SARS-CoV-2 rS + Matrix-M Adjuvant |
|---|---|---|---|---|
| A | 300 | Placebo | Placebo | Placebo |
| B1 | 150 | 5 µg + 50 µg | 5 µg + 50 µg | Placebo |
| B2 | 150 | 5 µg + 50 µg | 5 µg + 50 µg | 5 µg + 50 µg |
| C1 | 150 | 5 µg + 50 µg | Placebo | Placebo |
| C2 | 150 | 5 µg + 50 µg | Placebo | 5 µg + 50 µg |
| D | 300 | 25 µg + 50 µg | 25 µg + 50 µg | Placebo |
| E | 300 | 25 µg + 50 µg | Placebo | Placebo |

Note: The first dose represents the amount of antigen (SARS-CoV-2 rS) and the second dose represents the amount of adjuvant (Matrix-M). For example, 5 µg + 50 µg represents 5 µg SARS-CoV-2 rS + 50 µg Matrix-M adjuvant.

Subjects in Treatment Groups B and C who agree to continue study participation for an additional 6 months will receive a booster dose of vaccine (B1, B2, and C1) or placebo (C2) at Day 357 (Table 10-3). This change to the boosting regime is being implemented in order to gain additional immunogenicity and safety data for subjects who receive 1 or 2 doses of 5 µg SARS-CoV-2 rS + 50 µg Matrix-M1 adjuvant on Days 0 and 21 (ie, the dose level intended for licensure) and a booster dose of 5 µg SARS-CoV-2 rS + 50 µg Matrix-M1 adjuvant on Day 189.

**Table 10-3 Treatment Groups B and C, Additional Dose on Day 357 (Part 2)**

| Treatment Group | Number of Subjects | Day 0<br>SARS-CoV-2 rS + Matrix-M Adjuvant | Day 21 (−1 to +3 days)<br>SARS-CoV-2 rS + Matrix-M Adjuvant | Day 189 (±15 days)<br>SARS-CoV-2 rS + Matrix-M Adjuvant | Day 357 (±15 days)<br>SARS-CoV-2 rS + Matrix-M Adjuvant |
|---|---|---|---|---|---|
| B1 | 150 | 5 µg + 50 µg | 5 µg + 50 µg | Placebo | 5 µg + 50 µg |
| B2 | 150 | 5 µg + 50 µg | 5 µg + 50 µg | 5 µg + 50 µg | 5 µg + 50 µg |
| C1 | 150 | 5 µg + 50 µg | Placebo | Placebo | 5 µg + 50 µg |
| C2 | 150 | 5 µg + 50 µg | Placebo | 5 µg + 50 µg | Placebo |

Note: The first dose represents the amount of antigen (SARS-CoV-2 rS) and the second dose represents the amount of adjuvant (Matrix-M). For example, 5 µg + 50 µg represents 5 µg SARS-CoV-2 rS + 50 µg Matrix-M adjuvant.

Study vaccinations will comprise up to 3 IM injections (Day 0 and Day 21 [priming doses] and Day 189 [booster], in Treatment Groups A, D, and E, and up to 4 IM injections (Day 0 and Day 21 [priming doses], Day 189 [6-month booster], and Day 357 [12-month booster]), in Treatment Groups B and C, ideally in alternating deltoids for priming doses, with the study vaccine assigned in a full dose injection volume of approximately 0.5 mL. All vaccinations will be administered on an outpatient basis by designated site personnel in a way to maintain the blind, and if subjects are not able to attend sites due to local recommendations/restrictions related to the COVID-19 pandemic, home visits for vaccine administration may be utilized, or alternative models of vaccine administration such as drive-through clinics. Following study vaccination, subjects should either remain in the clinic or under study staff observation if vaccinated during home visits or at other locations, for at least 30 minutes post-vaccination to be monitored for any immediate anaphylaxis reactions.

Any pharmacy preparation with unblinded product will require unblinded site personnel who will not otherwise be involved in the study procedures or observation of subjects. The goal of Part 2 will be to utilize co-formulated product; however, the Part 2 site(s) may initiate dosing using the approach of bedside mixing as noted in the Part 1 portion of the protocol (with the appropriate concentrations being tested in Part 2).

Blood samples for immunogenicity assessments will be collected before vaccination and at selected time points following each vaccination. Immune measurements (ELISA) will be conducted on serum (IgG) for SARS-CoV-2 rS protein antigen(s) and ACE2 receptor binding inhibition. Additional immunogenicity assessments specific to SARS-CoV-2 (or related variants), including anti-nucleocapsid protein serology, will include a neutralizing antibody assay. Blood samples for serology will be collected at baseline but will not be used for inclusion/exclusion for randomization as a medical history will suffice; however, individuals with positive serologies (hepatitis B, hepatitis C, human immunodeficiency virus [HIV]) will not be included in the primary immunogenicity analysis.

Subgroup analyses of the primary immunogenicity analysis (ie, serum IgG antibody levels specific for the SARS-CoV-2 rS protein antigen[s] as detected by ELISA using GMT or SCR for the two-dose regimens by dose at Day 35 regardless of baseline immune status and stratified by baseline immune status), as measured by in-house SARS-CoV-2 IgG assay at the Novavax central immunology laboratory, will be undertaken. All formal analyses will utilize the cut-off of SARS-CoV-2 IgG from the Novavax central immunology laboratory.

Operational process will aim to manage the number of SARS-CoV-2 antibody-positive subjects enrolled in the study, such that not more than 15% of subjects test SARS-CoV-2 antibody-positive at baseline. This will be done using community seroprevalence estimates considered during site selection processes, and baseline SARS-CoV-2 antibody results are not required to be known prior to subject enrollment.

Safety assessments will include monitoring and recording of solicited (local and systemic reactogenicity events) and unsolicited AEs; MAAEs; AESI; SAEs; vital sign measurements; and targeted physical examination findings. Symptoms related to either a suspected, probable, or confirmed COVID-19 case for illness events starting prior to Day 28 should be recorded as unsolicited AEs, or multiple symptom-based AEs can be aggregated into a single AE of a suspected, probable, or confirmed COVID-19 case.

If a US subject is unblinded in the study and receives an approved/authorized vaccine from different manufacturer, all vaccine administration errors, all SAEs, cases of multisystem inflammatory syndrome, and hospitalized or fatal cases of COVID-19 following vaccination with the approved/authorized vaccine must be reported to the Vaccine Adverse Event Reporting System (VAERS). Investigators in Australia should follow local Regulatory reporting guidance for safety events that occur in subjects who receive approved/authorized vaccines within the study.

COVID-19 disease monitoring for qualifying symptoms of suspected COVID-19 will commence every 14 days beginning at Day 28 until. Details of COVID-19 disease monitoring can be found in Section 13.3.1 (see also Section 13.3.1.1 for a note about the removal of the requirement for the use of ePRO for self-monitoring of qualifying symptoms of suspected COVID-19, applicable only to subjects enrolled at Australian sites). Table 13-1 details qualifying symptoms of suspected COVID-19. If a subject has qualifying symptoms, this will trigger a request to subjects to self-sample according to protocol instructions (see below).

An independent SMC will remain in effect from Part 1 of the study and will review safety data in aggregate when data snapshots are available (see Section 14.5) from the Day 21 and/or Day 35 time point (to inform potential stage-gate decisions for follow-on clinical studies) and every 3 to 4 months during Part 2 of the study or sooner if there are new safety signals noted by sponsor internal safety reviews or if an excess number of COVID-19 cases occur. The study will continue as planned during the SMC reviews, with the exception of when an enrollment pause will occur following initial enrollment of approximately 250 older adult subjects. An SMC Charter will document SMC membership and processes, data review time points, and safety tables that will be reviewed.

For Treatment Groups A, D, and E, Part 2 will consist of a screening period (Days −45 to 0); vaccination days (Days 0, 21, and 189); outpatient study visits on Day 0 and on Days 21 (−1 to +3 days), 35 (−1 to +3 days), 105 (± 7 days), 189 (± 15 days), 217 (+7 days), 357 (± 15 days), and a phone visit at Day 273 (± 15 days). Subjects in an immunogenicity subset for PBMC collection will additionally have outpatient study visits on Days 7 (−1 to +3 days) and 28 (−1 to +3 days) (See Table 10-4). For Treatment Groups B and C, Part 2 will consist of a screening period (Days −45 to 0); vaccination days (Days 0, 21, 189, and 357); outpatient study visits on Day 0 and on Days 21 (−1 to +3 days), 35 (−1 to +3 days), 105 (± 7 days),

189 (± 15 days), 217 (+7 from booster dose), 357 (± 15 days), 371 (−1 to +3 days), and Day 546/EOS, and phone visits at Days 273 (± 15 days), Day 385 (−1 to +3 days) and Day 475 (± 15 days). Subjects in an immunogenicity subset for PBMC collection will additionally have outpatient study visits on Days 7 (−1 to +3 days) and 28 (−1 to +3 days) (See Table 10-4 and Table 10-5).

The duration of the Part 2, excluding screening, is approximately 12 months for subjects enrolled in Treatment Groups A, D, and E and approximately 18 months for those subjects in Treatment Groups B and C who agree to stay in the study through the Day 357 boost and 6-month follow up. Visits requiring vaccination or blood sampling may occur via home visits or using local services, if conducted by qualified personnel (eg, phlebotomy, home health, external laboratory, home visit by site staff).If the Novavax study vaccine or another vaccine from a different manufacturer is demonstrated to be safe and efficacious and approved and/or authorized for use by regulatory authorities in the US or Australia, subjects for whom the new approved/authorized vaccine is recommended and available will be counseled with respect to their options. These subjects may be offered the opportunity to be unblinded so that those who received placebo may be offered the Novavax vaccine or another approved/authorized vaccine, as appropriate, outside the protocol procedures. Subjects who received the Novavax vaccine and who wish to receive an approved/authorized vaccine from another manufacturer will be advised to discuss this plan with their healthcare provider given the current lack of safety data regarding the sequential administration of vaccines made by different manufacturers. Subjects who are unblinded and receive an approved/authorized vaccine in this manner will be strongly encouraged to remain in study for safety follow-up as defined in the protocol. However, subjects also have the right to discontinue participation in the study at any time.

Due to the ongoing pandemic, recent national regulatory and local HREC/IRB and public health guidance will be applied at the site locations regarding alternations in the ability of study subjects to attend an investigational site for protocol specified visits, with the site's investigator being allowed to conduct safety assessments (eg, telephone contact, virtual visit via telemedicine, alternative location for assessment, including local laboratories or imaging centers) when necessary and feasible, as long as such visits are sufficient to assure the safety of study subjects. Serum samples may be drawn using local phlebotomy services, home health, or other modalities if site visits cannot occur. Vaccination visits must have adequate oversight for issues associated with immediate severe reactions but may need to occur outside of the clinical site depending on the pandemic situation (eg, home vaccinations).

#### 10.1 SCHEDULE OF EVENTS

**Table 10-4 Schedule of Events (Part 2) for Subjects in All Treatment Groups**

| **Study Day:** | **–45 to 0** | **0[a]** | **7** | **21** | **28** | **35** | **105** | **189** | **217** | **Unscheduled[p]** | **273[b]** | **357[x]** |
|---|---|---|---|---|---|---|---|---|---|---|---|---|
| **Window (days)[c]:** | – | 0 | –1 to +3 | –1 to +3 | –1 to +3 | –1 to +3 | ±7 | ±15 | +7 | – | ±15 | ±15 |
| **Minimum days following most recent vaccination:[c]** | – | 0 | 6 | 20 | 6 | 13 | 77 | 153 | 28 | – | 69 | 153 |
| **Study Visit:** | **Screening** | **1** | **2** | **3** | **4** | **5** | **6** | **7** | **8** | **Unscheduled** | **9[b]** | **10/EOS** |
| Informed consent | X | | | | | | | | | | | |
| Medical history[d] | X | | | | | | | | | | | |
| Inclusion/exclusion criteria | X | X[e,f] | | X[e,f] | | | | X[e,f] | | | | |
| Demographics[g] | X | | | | | | | | | | | |
| Prior/concomitant medications[h] | X | X[e,f] | | X[e,f] | | X | X | X[e,f] | X | X | X[i] | X[i] |
| Vital sign measurements | X | X[j] | | X[j] | | | | X[j] | | X[j] | | |
| Urine pregnancy test[k] | X | X[f] | | X[f] | | | | X[f] | | | | |
| Serum FSH[l] | X | | | | | | | | | | | |
| Serology (not exclusionary for subject entry)[m] | X | | | | | | | | | | | |
| Baseline testing for SARS-CoV-2 serostatus (ELISA)[n] | | X | | | | | | | | | | |
| Targeted physical examination[o] | X | X[f] | | X[f] | | X | X | X[f] | X | X | | |
| Clinical laboratory testing[q] | X | | | | | | | | | X | | |
| Vaccination | | X | | X | | | | X | | | | |
| Reactogenicity[r,s] | | X | | X | | | | X | | | | |
| Blood sampling for SARS-CoV-2 immunogenicity[t] | | X[f] | | X[f] | | X | X | X[f] | X | X | | X |
| PBMC collection (subset) | | X[f] | X | | X | | | | | | | |
| Monitoring for COVID-19 disease endpoints | | | | | Day 28 through Day 217 | | | | | | | |
| Sample collection for SARS-CoV-2 qualitative PCR test (self-collected from Day 28, or site collected at Visit 1 onwards, or at unscheduled visits if clinically indicated) | | X | | | X[u] | | | | | X[v] | | |
| All unsolicited AEs since prior visit | | X | | X | | X | | | X | | | |
| All MAAEs | | X | | X | | X | X | X | X | | | |
| Any MAAE attributed to vaccine | | X | | X | | X | X | X | X | X | X | X |
| SAEs | | X | | X | | X | X | X | X | X | X | X |
| AESI[w] | | X | | X | | X | X | X | X | X | X | X |
| EOS form[x] | | | | | | | | | | | | X |

| **Study Day:** | **–45 to 0** | **0[a]** | **7** | **21** | **28** | **35** | **105** | **189** | **217** | **Unscheduled[p]** | **273[b]** | **357[x]** |
|---|---|---|---|---|---|---|---|---|---|---|---|---|
| **Window (days)[c]:** | – | **0** | **–1 to +3** | **–1 to +3** | **–1 to +3** | **–1 to +3** | **±7** | **±15** | **+7** | – | **±15** | **±15** |
| **Minimum days following most recent vaccination:[c]** | – | **0** | **6** | **20** | **6** | **13** | **77** | **153** | **28** | – | **69** | **153** |
| **Study Visit:** | **Screening** | **1** | **2** | **3** | **4** | **5** | **6** | **7** | **8** | **Unscheduled** | **9[b]** | **10/EOS** |
| Analyses to be performed after all subjects have completed designated visit (interim analyses through Day 357) | | | | X | | X | X | | X | | | X |

Abbreviations: ACE2 = angiotensin-converting enzyme; AE = adverse event; AESI = adverse event of special interest; BMI = body mass index; COVID-19 = coronavirus disease 2019; ELISA = enzyme-linked immunosorbent assay; EOS = end of study; ePRO = electronic patient-reported outcome; FSH = follicle-stimulating hormone; HIV = human immunodeficiency virus; IgG = immunoglobulin G; MAAE = medically attended adverse event; PBMC = peripheral blood mononuclear cells; PCR = polymerase chain reaction; SAE = serious adverse event; SARS-CoV-2 = severe acute respiratory syndrome coronavirus 2.

[a] If screening and randomization occur on the same day (ie, Day 0), study visit procedures should not be duplicated.

[b] Telephone call. Should subjects decide to terminate early, an EOS visit will occur to collect the maximum safety data possible.

[c] Days relative to vaccination are estimates due to window allowance. Should a study pause occur, visits/windows will be adjusted to allow subjects to continue without protocol deviation. Visit schedules following the second and third vaccinations are calculated relative to the day the vaccinations were received.

[d] Including prior and concomitant medical conditions, recent vaccinations (≤90 days), and significant surgical procedures.

[e] Specific exclusions to vaccination or potential reasons for vaccination deferral will be assessed (Section 11.3). Should subjects start specific medications or develop specific diagnoses that are exclusionary at baseline (but not present at baseline) prior to later vaccination time points, then approval for vaccination at those latter time points must be given by medical monitor or sponsor.

[f] Performed prior to vaccination.

[g] Screening only. Including date of birth (day, month, and year), sex, race, ethnicity, weight, height, and BMI (derived).

[h] Recent and current medications at the time of screening to be reviewed to ensure eligibility criteria are fulfilled. Concomitant medications include all medications (including vaccines in the past 90 days) taken by the subject.

[i] Concomitant medications associated with any MAAE attributed to vaccine, potential AESI, or SAE will be recorded. Receipt of any authorized or approved COVID-19 vaccine will also be recorded as a concomitant medication.

[j] On vaccination days, vital sign measurements (including blood pressure and temperature) will be collected at least once before vaccination and as clinically needed during the 30-minute post-vaccination observation period. Subjects must have controlled blood pressure and no evidence of fever prior to vaccination (Section 11.3). Vital sign measurements may be collected during an unscheduled visit at the discretion of the investigator.

[k] Women of childbearing potential only. A urine pregnancy test will be performed at screening and prior to each vaccination. A serum pregnancy test may be used at screening or at the discretion of the investigator. A positive urine pregnancy test at any time will result in the subject not receiving any further vaccination.

[l] Females only. A serum FSH test may be performed at screening to confirm postmenopausal status.

| **Study Day:** | **–45 to 0** | **0[a]** | **7** | **21** | **28** | **35** | **105** | **189** | **217** | **Unscheduled[p]** | **273[b]** | **357[x]** |
|---|---|---|---|---|---|---|---|---|---|---|---|---|
| **Window (days)[c]:** | – | 0 | –1 to +3 | –1 to +3 | –1 to +3 | –1 to +3 | ±7 | ±15 | +7 | – | ±15 | ±15 |
| **Minimum days following most recent vaccination:[c]** | – | 0 | 6 | 20 | 6 | 13 | 77 | 153 | 28 | – | 69 | 153 |
| **Study Visit:** | Screening | 1 | 2 | 3 | 4 | 5 | 6 | 7 | 8 | Unscheduled | 9[b] | 10/EOS |

[m] Serology testing will include hepatitis B, hepatitis C, and HIV. Note if serology tests are positive, then subjects will be excluded from the per-protocol analysis set but not from the safety analysis set.

[n] Serostatus at time of study enrollment will be based on Day 0 seropositivity to serum IgG antibody to SARS-CoV-2 proteins from blood samples for study endpoint assessments and will not be a separate blood draw/test.

[o] Examination at screening to include height and weight (derived BMI based on gender), lungs, heart, and abdomen as well as the lymphatic assessment of upper extremities to allow for vaccination; symptom-directed (targeted) physical examination at all other scheduled time points but always to include lymphatic assessment of injected upper extremity on vaccination days. Interim physical examinations will be performed at any unscheduled visit at the discretion of the investigator, if necessary.

[p] If applicable, based on reason for unscheduled visit. Subjects may also be evaluated by other methods (eg, telemedicine, hospital/COVID-19 intensive care ward records, home visits).

[q] Baseline clinical laboratory testing will not include toxicity grading but will be recorded for future AE assessment. A complete list of assessments is provided in Section 13.1.2. None of these laboratory tests are exclusionary for enrollment or vaccination. Laboratory tests may be included in unscheduled visits if deemed necessary by the investigator.

[r] On vaccination days, subjects should remain in the clinic or under study staff observation for at least 30 minutes post-vaccination to be monitored for any immediate anaphylaxis reactions.

[s] Subjects will utilize an electronic subject-reported outcome application to record reactogenicity following vaccination. All subjects will record reactogenicity on the day of vaccination and for an additional 6 days after vaccination. Should any reactogenicity event extend beyond 6 days after vaccination (toxicity grade ≥1), then it will be recorded as an AE with start dates of Day 7, Day 28, or Day 196 (the first day after completion of the subject solicited symptom electronic diary) and followed to resolution per FDA guidelines for dataset capture. The standard toxicology grading scale implemented in the electronic diary for Part 2 of the study is included in Section 16.3 (Appendix 3, Table 16-2).

[t] IgG antibody to SARS-CoV-2 protein and ACE2 receptor binding inhibition assays will be performed at each of the time points indicated for all subjects. Neutralization testing will be conducted in a subset of subjects at a minimum of the Day 0, Day 35, Day 217, and Day 357 time points as specified in the Statistical Analysis plan. PBMC collection for cell-mediated immunity testing will be assessed in a subset of subjects and time points, with a target PBMC subset of approximately 150-200 evaluable subjects across study treatment groups at the Day 0, Day 7, and Day 28 time points.

[u] Subjects will be instructed on self-collection using a nasal mid-turbinate sample approach from Day 28 and will demonstrate competency during an early clinic visit (eg, Day 21). An ePRO algorithm will be developed to instruct when self-collection is required (Section 13.3.1 and Section 13.3.1.1). Self-collection of samples for ad-hoc potential COVID-19 illnesses should occur within 3 days of symptom onset if possible. Subjects will also be notified to self-collect samples every 28 days starting from Day 28 if symptom free (ie, once monthly) to assess for asymptomatic carrier status. Should a subject be admitted to the hospital or COVID-19 intensive care ward and self-collection is unavailable, then a local public health test will be taken as a valid result.

[v] For unscheduled visits, site staff may collect samples for PCR confirmation.

| Study Day: | –45 to 0 | 0[a] | 7 | 21 | 28 | 35 | 105 | 189 | 217 | Unscheduled[p] | 273[b] | 357[x] |
|---|---|---|---|---|---|---|---|---|---|---|---|---|
| Window (days)[c]: | – | 0 | –1 to +3 | –1 to +3 | –1 to +3 | –1 to +3 | ±7 | ±15 | +7 | – | ±15 | ±15 |
| Minimum days following most recent vaccination:[c] | – | 0 | 6 | 20 | 6 | 13 | 77 | 153 | 28 | – | 69 | 153 |
| Study Visit: | Screening | 1 | 2 | 3 | 4 | 5 | 6 | 7 | 8 | Unscheduled | 9[b] | 10/EOS |

[w] To include PIMMC (Appendix 4, Table 16-5), AESIs relevant to COVID-19 disease (Appendix 4, Table 16-6), or any newly identified potential AESI followed through 365 days after final vaccination.

[x] EOS form and assessments will be completed at the final study visit for all subjects in Treatment Groups A, D, and E, and those in Treatment Groups B and C who are terminated early.

**Table 10-5 Schedule of Events: Treatment Groups B and C, Day 357 to EOS (Part 2)**

| Study Day: | 357 | 371 | Unscheduled[j] | 385[m] | 475[m] | 546 |
|---|---|---|---|---|---|---|
| Window (days)[a]: | ±15 | –1 to +3 | – | –1 to +3 | ±15 | ±15 |
| Minimum days following most recent vaccination:[a] | 153 | 13 | – | 27 | 103 | 174 |
| Study Visit: | 10 | 11 | Unscheduled | 12 | 13 | 14/EOS |
| Inclusion/exclusion criteria | X[b,c] | | | | | |
| Review COVID-19 vaccination status to exclude participants who received an authorized vaccine | X | | | | | |
| Prior/concomitant medications[n] | X[b,d] | X | X | X | X | X |
| Vital sign measurements | X[g] | | X | | | |
| Urine pregnancy test[e] | X[c] | | | | | |
| Targeted physical examination[f] | X[c] | X | X | | | |
| Vaccination | X | | | | | |
| Reactogenicity[g,h] | X | | | | | |
| Blood sampling for SARS-CoV-2 immunogenicity[i] | X[c] | X | | | | X |
| Blood sampling for SARS-CoV-2 (ELISA for anti N-protein serology) | | X | | | | X |
| All unsolicited AEs since prior visit | | X | | X | | |
| All MAAEs | | | | | | |
| Any MAAE attributed to vaccine | X | X | X | X | X | X |
| SAEs | X | X | X | X | X | X |
| AESI[k] | X | X | X | X | X | X |
| EOS form[l] | | | | | | X |

Abbreviations: ACE2 = angiotensin-converting enzyme; AE = adverse event; AESI = adverse event of special interest; COVID 19 = coronavirus disease 2019; ELISA = enzyme-linked immunosorbent assay; EOS = end of study; ePRO = electronic patient-reported outcome; IgG = immunoglobulin G;

MAAE = medically attended adverse event; anti-N = anti-nucleocapsid; SAE = serious adverse event; SARS-CoV-2 = severe acute respiratory syndrome coronavirus 2.

[a] Days relative to vaccination are estimates due to window allowance. Should a study pause occur, visits/windows will be adjusted to allow subjects to continue without protocol deviation. Visit schedules following the Day 357 vaccination are calculated relative to the day the vaccination was received.

[b] Specific exclusions to vaccination or potential reasons for vaccination deferral will be assessed (Section 11.3). Should subjects start specific medications or develop specific diagnoses that are exclusionary at baseline (but not present at baseline) prior to later vaccination time points, then approval for vaccination at those latter time points must be given by medical monitor or sponsor.

[c] Performed prior to vaccination.

[d] Current medications to be reviewed to ensure eligibility for dosing.

[e] Women of childbearing potential only. A urine pregnancy test will be performed prior to vaccination. A positive pregnancy test at any time will result in the subject not receiving any further vaccination.

[f] Symptom-directed (targeted) physical examination. Interim physical examinations will be performed at any unscheduled visit at the discretion of the investigator, if necessary.

[g] On vaccination days, subjects should remain in the clinic or under study staff observation for at least 30 minutes post-vaccination to be monitored for any immediate anaphylaxis reactions.

[h] Subjects will utilize an electronic subject-reported outcome application to record reactogenicity following vaccination. All subjects will record reactogenicity on the day of vaccination and for an additional 6 days after vaccination. Should any reactogenicity event extend beyond 6 days after vaccination (toxicity grade ≥ 1), then it will be recorded as an AE with a start date of Day 364 (the first day after completion of the subject solicited symptom electronic diary) and followed to resolution per FDA guidelines for dataset capture. The toxicology grading scale implemented in the electronic diary for Part 2 of the study is included in Section 16.3 (Appendix 3, Table 16-2).

[i] IgG antibody to SARS-CoV-2 protein and ACE2 receptor binding inhibition assays (Wuhan and B.1.351 strain or other variants) will be performed at each of the time points indicated.

[j] If applicable, based on reason for unscheduled visit. Subjects may also be evaluated by other methods (eg, telemedicine, hospital/COVID-19 intensive care ward records, home visits).

[k] To include PIMMC (Appendix 4, Table 16-5), AESIs relevant to COVID-19 disease (Appendix 4, Table 16-6), or any newly identified potential AESI followed through EOS.

[l] EOS form will be completed for all participants, including those who terminate early.

[m] Telephone call. Should subjects decide to terminate early, an EOS visit will occur to collect the maximum safety data possible.

[n] Concomitant medications associated with any MAAE attributed to vaccine, potential AESI, or SAE will be recorded. Receipt of any authorized or approved COVID-19 vaccine will also be recorded as a concomitant medication.

### 11. STUDY POPULATION

Healthy male or female subjects will be enrolled at up to 40 clinical sites across Australia and/or the United States. Approximately 750 healthy male and female subjects between 18 and 84 years of age (inclusive) will be randomized in each country, with approximately 50% of subjects overall in the study ≥60 years of age. Two-factor, 2-level stratification will be employed (ages 18-59 and 60-84 years; study site). Up to approximately 1500 subjects could be enrolled for the 5 vaccine groups across the 2 countries to potentially mitigate the risk of enrollment in either country being delayed, or the expected availability of study data from either country being delayed due to the potential impact of the pandemic on feasibility of study vaccine delivery, possibilities for adequate specimen and data collection, and feasible specimen transportation. This flexibility would allow review of study data on approximately 500 subjects to facilitate initiation of Phase 3 clinical development as soon as possible.

For the Part 2 component of the study, following study initiation, enrollment of older adult subjects (≥ 60 to 84 years of age) will be paused when approximately 50 subjects in the older age group are enrolled in each of the 5 study vaccine groups (ie, approximately 250 older adult subjects in total enrolled subjects across all sites). The SMC will then review reactogenicity data for both study age groups for 5 days following the first vaccine dose (Day 0 through Day 4, inclusive) when all subjects in the older adult group have accrued these 5 days of data. Enrollment of adult subjects (≥ 18 to 59 years of age) will not be paused during this SMC review unless general vaccination pause rules detailed in Section 13.1.5 are also met. Details of this review, and processes to restart enrollment of older adult subjects will be documented in the SMC Charter.

Depending on the current pandemic situation and ability to rapidly test, enrollment of subjects with prior SARS-COV-2 exposure/positive testing will be limited on a site-by-site basis with an overall goal of not more than 15% of the population being SARS-CoV-2 positive at baseline; however, the result for baseline seropositivity will not need to be known for subjects prior to enrollment, rather it will be retrospectively assessed from baseline serology, with potential shifts to enrollment in sites with lower levels of baseline seropositivity if needed during the enrollment period.

#### 11.1 INCLUSION CRITERIA

Subject eligibility for the study (ie, subject reported to successfully meet all Inclusion Criteria and not meet any Exclusion Criteria) will be recorded in the subject source notes. Clinical validation of self-reported subject information related to eligibility is not routinely required for enrollment, unless specified for particular eligibility criteria.

Each subject must meet all of the following criteria to be enrolled in this study:

1. Healthy adult males or females between 18 and 84 years of age, inclusive, at screening who are of legal adult age in their local jurisdiction. Healthy status will be determined by the investigator based on medical history, vital sign measurements, and physical examination at screening.

2. The subject has a body mass index 17 to 35 kg/m$^2$, inclusive, at screening.

3. Willing and able to give informed consent prior to study enrollment and comply with study procedures.

4. Female subjects of childbearing potential (defined as any female who has experienced menarche and who is NOT surgically sterile [ie, hysterectomy, bilateral tubal ligation, or bilateral oophorectomy] or postmenopausal [defined as amenorrhea at least 12 consecutive months or documented plasma FSH ≥40 mIU/mL]) must agree to be heterosexually inactive from at least 21 days prior to enrollment and through 6 months after the last vaccination OR agree to consistently use any of the following methods of contraception from at least 21 days prior to enrollment and through 6 months after the last vaccination:

    a. Condoms (male or female) with spermicide (if acceptable in country)

    b. Diaphragm with spermicide

    c. Cervical cap with spermicide

    d. Intrauterine device

    e. Oral or patch contraceptives

    f. Norplant®, Depo-Provera®, or other in country regulatory-approved contraceptive method that is designed to protect against pregnancy

g. Abstinence, as a form of contraception, is acceptable if in line with the subject's lifestyle

**NOTE**: Periodic abstinence (eg, calendar, ovulation, symptothermal, post-ovulation methods), male partner vasectomy, and withdrawal are not acceptable methods of contraception. These procedures and laboratory test results must be confirmed by physical examination, by subject recall of specific date and hospital/facility of procedure, or by medical documentation of said procedure.

#### 11.2 EXCLUSION CRITERIA

Subjects meeting any of the following criteria will be excluded from the study (subject-reported unless otherwise indicated):

1. Subjects who are having any current workup of undiagnosed illness within the last 8 weeks, which is either subject-reported or has been clinician-assessed, that could lead to a new condition diagnosis.

2. Participation in research involving receipt of an investigational product (drug/biologic/device) within 45 days prior to first study vaccination.

3. History of a confirmed diagnosis of SARS or history of a confirmed diagnosis of COVID-19 disease resulting in medical intervention.

   **NOTE**: Subjects with a history of confirmed COVID-19 disease resulting in mild symptoms are allowed. Mild symptoms are defined as those in which treatment was symptom relief and not systemic or respiratory supportive care (eg, IV hydration, oxygen, nebulizer), with OTC medications being allowed.

4. Received influenza vaccination within 14 days prior to first study vaccination, or any other vaccine within 4 weeks prior to first study vaccination.

5. Have clinically significant chronic cardiovascular, endocrine, gastrointestinal/ hepatic, renal, neurological, respiratory, or other medical disorders not excluded by other exclusion criteria, that are assessed by the investigator as being clinically unstable within the prior 4 weeks evidenced by: a) hospitalization for the condition, including day surgical interventions, b) new significant organ function deterioration, c) needing addition of new treatments or major dose adjustments of current treatments.

6. Diabetes mellitus requiring insulin therapy (either type 1 or type 2 diabetes mellitus).

7. Chronic obstructive pulmonary disease with a history of an acute exacerbation of any severity in the prior year.

8. Any history of congestive heart failure.

9. Any history of chronic kidney disease (the presence of impaired or reduced kidney function lasting at least 3 months). Clinical validation of potential cases of chronic kidney disease should be conducted.

10. Evidence of unstable coronary artery disease as manifested by cardiac intervention, addition of new cardiac medications for control of symptoms, or unstable angina in the past 3 months.

11. History of chronic neurological disorders that have required prior specialist physician review for diagnosis and management (such as multiple sclerosis, dementia, transient ischemic attacks, Parkinson's disease, degenerative neurological conditions, neuropathy and epilepsy) or a history of stroke or previous neurological disorder within 12 months with residual symptoms. Subjects with a history of migraine or chronic headaches or nerve root compression that have been stable on treatment for the last 4 weeks are not excluded.

12. Any autoimmune or immunodeficiency disease/condition (iatrogenic or congenital).

    **NOTE:** Stable endocrine disorders that have a confirmed autoimmune etiology (eg, thyroid, pancreatic) are allowed.

13. Chronic administration (defined as more than 14 continuous days) of immunosuppressants, systemic glucocorticosteroids reaching an immunosuppressive dose, or other immune-modifying drugs within 90 days prior to first study vaccination.

    **NOTE:** An immunosuppressant dose of glucocorticoid is defined as a systemic dose ≥10 mg of prednisone per day or equivalent. The use of topical, inhaled, and nasal glucocorticoids will be permitted if other chronic disease conditions are not exclusionary.

14. Received immunoglobulin, blood-derived products, or other immunosuppressant drugs within 90 days prior to first study vaccination.

15. Known disturbance of coagulation (iatrogenic or congenital).

**NOTE:** The use of ≤325 mg of aspirin per day as prophylaxis is permitted, but the use of other platelet aggregation inhibitors, thrombin inhibitors, Factor Xa inhibitors, or warfarin derivatives is exclusionary due to potential bleeding following IM injection.

16. Active cancer (malignancy) within 5 years prior to first study vaccination (with the exception of adequately treated non-melanomatous skin carcinoma, at the discretion of the investigator).
17. Any known allergies to products contained in the investigational product or latex allergy.
18. Women who are breastfeeding or who plan to become pregnant during the study.
19. History of alcohol abuse or drug addiction within one year prior to the first study vaccination.
20. Any condition that, in the opinion of the investigator, would pose a health risk to the subject if enrolled or could interfere with evaluation of the study vaccine or interpretation of study results (including neurologic or psychiatric conditions deemed likely to impair the quality of safety reporting).
21. Study team member or first-degree relative of any study team member (inclusive of sponsor, PPD, and site personnel involved in the study).

#### 11.3 OTHER CONSIDERATIONS

Subjects meeting any of the following criteria may have planned study vaccination deferred for a later date, but these criteria are not exclusionary for study enrollment. The sponsor may advise sites of dates after which potential subjects who have been deferred need to have been enrolled, due to the need to rapidly enroll subjects in this study to allow rapid reporting of results to allow initiation of future studies.

- Respiratory symptoms in the past 3 days (ie, cough, sore throat, difficulty breathing). Subject may be vaccinated once all symptoms have been resolved for >3 days. Out of window vaccination is allowed for this reason.
- Temperature of >38°C within 24 hours of planned vaccination (site measured or subject measured). Subject may be vaccinated once the fever has resolved and there has not been any temperature measured as being >38°C for >3 days. Out of window vaccination is allowed for this reason.

NOTE: Screening for COVID-19 disease symptoms may be indicated for either of the above-mentioned reasons or if COVID-19 disease is suspected based on potential exposure to SARS-CoV-2 infection through either close contacts or based on local epidemiology. In such a case, subjects should also have study samples collection for qualitative PCR testing on the day of any subsequent study vaccination, but the results of the qualitative PCR test are not needed before study vaccination can be given for the first or subsequent doses. Any subjects with new positive PCR-confirmed SARS-CoV-2 infections occurring from screening and prior to the end of immunogenicity assessments will be removed from applicable immunogenicity analyses as defined in the statistical analysis plan.

- Any acute illness (cardiovascular, endocrine, gastrointestinal/hepatic, renal, neurological, respiratory, or other medical disorders) that is actively causing symptoms that could, in the opinion of the investigator, impact the assessment of reactogenicity or other study assessments. Subject may be vaccinated once symptoms have resolved or are stabilized for >3 days. Out of window vaccination is allowed for this reason.

- Immunization with any vaccine within 14 days prior to vaccination. Out of window vaccination is allowed for this reason.

- Blood pressure that exceeds the United States Eighth Joint National Committee control levels (by age) prior to vaccination (BP is 150/90 mm Hg or higher in adults 60 years and older, or 140/90 mm Hg or higher in adults younger than 60 years). Repeated BP testing following a high reading can occur up to a total of a further 2 times during the same visit after a pause of not less than 5 minutes between measures, and if the reading is then within the permitted range the subject may be randomized and vaccinated. Out of window vaccination is allowed for this reason if subjects are deferred on any planned vaccination day due to transient abnormal BP readings.

#### 11.4 WITHDRAWAL OF SUBJECTS FROM THE STUDY

##### 11.4.1 Reasons for Withdrawal

Subjects can withdraw consent and discontinue from the study at any time, for any reason. Subjects may refuse further procedures (including vaccination) but are encouraged to remain in the study for safety follow-up. In such cases where only safety is being conducted, subject contact could be managed via telemedicine contact (eg, telephone, web chat, video, FaceTime).

The investigator may **withhold** further vaccination from a subject in the study if after study enrollment the subject:

1. Is non-compliant with the protocol;
2. Experiences an SAE or intolerable AE(s) for which vaccination is not advised by the investigator;
3. Has laboratory safety assessments that reveal clinically significant hematological or biochemical changes and is deemed to be best for the subject’s health (post-enrollment evaluation);
4. Becomes pregnant (discontinuation of further vaccination **required**).
5. Receives approved/authorized vaccine to prevent COVID-19.

The investigator can also withdraw a subject upon the request of the sponsor or if the sponsor terminates the study. Upon the occurrence of an SAE or intolerable AE, the investigator may confer with the sponsor before future vaccination.

##### 11.4.2 Handling of Withdrawals

Subjects are free to withdraw from the study at any time upon request. Subject participation in the study may be stopped at any time at the discretion of the investigator or at the request of the sponsor.

When a subject withdraws from the study, the reason(s) for withdrawal shall be recorded by the investigator on the relevant page of the eCRF. Whenever possible, any subject who withdraws from the study prematurely will undergo all EOS assessments. Any subject who fails to return for final assessments will be contacted by the site in an attempt to have them comply with the protocol. The status of subjects who fail to complete final assessments will be documented in the eCRF.

##### 11.4.3 Replacements

Subjects who withdraw, are withdrawn or terminated from this study, or are lost to follow-up after signing the ICF but prior to first study vaccination may be replaced. Subjects who receive study vaccine and subsequently withdraw, are discontinued from further vaccination, are terminated from the study, or are lost to follow-up will not be replaced.

### 12. STUDY TREATMENTS

#### 12.1 TREATMENTS ADMINISTERED

Study vaccinations will comprise up to 3 IM injections (Day 0 and Day 21 [priming doses] and Day 189 [6-month booster – active or placebo] in Treatment Groups A, D, and E, and up to 4 IM injections (Day 0 and Day 21 [priming doses] and Day 189 [6-month booster – active or placebo] and Day 357 [booster – active or placebo]) in Treatment Groups B and C, ideally in alternating deltoids for priming doses, with the study vaccine assigned in an full dose injection volume of approximately 0.5 mL. Dose levels of SARS-CoV-2 rS in Part 2 will be 5 µg and up to 25 µg, with 50 µg Matrix-M adjuvant).

#### 12.2 INVESTIGATIONAL PRODUCTS

The following supplies will be used for vaccination in the study:

| Product | Supplied Formulation |
|---|---|
| SARS-CoV-2 recombinant spike protein nanoparticle vaccine, co-formulated with Matrix-M adjuvant[a] | Co-formulated for injection, with the following formulations (single-dose or multi-dose vials):<br>1. 10 µg/mL SARS-CoV-2 rS with 100 µg/mL Matrix-M adjuvant, 0.5 mL or 6.0 mL vial<br>2. 50 µg/mL SARS-CoV-2 rS with 100 µg/mL Matrix-M adjuvant, 0.5 mL vial |
| Sodium chloride injection (sterile)[a] | Solution for injection, 0.9%, 10 mL vial |

[a] Novavax or its vendor will supply all investigational product materials with labelling.

It is anticipated that the product will be available in a co-formulated single vial for Part 2 of this study. If the co-formulated product is not available for the start of Part 2, then the product will be provided as a bed-side mix as in Part 1 of the study may be used. The co-formulated drug substance [redacted]

[redacted]

Further details on the study vaccine can be found in pharmacy manual.

##### 12.2.1 Investigational Product Packaging and Storage

Novavax, Inc. will provide adequate quantities and appropriate labelling of SARS-CoV-2 rS with Matrix-M adjuvant and placebo (sterile sodium chloride injection) and PPD will ensure distribution to the clinical sites from a designated depot. The clinical unit pharmacy will prepare the study treatments for each subject. Detailed instructions for the preparation of study vaccine will be provided in a separate pharmacy manual.

All investigational products must be stored according to the labeled instructions in a secure cabinet or room with access restricted to necessary clinic personnel. The site will be required to keep a temperature log to establish a record of compliance with storage conditions.

##### 12.2.2 Investigational Product Accountability

The investigator (or delegate) will maintain accurate records of receipt of all investigational product, including dates of receipt. Accurate records will be kept regarding when and how much investigational product is dispensed and used by each subject in the study. Reasons for departure from the expected dispensing regimen must also be recorded. At the completion of the study, and to satisfy regulatory requirements regarding investigational product accountability, all investigational product will be reconciled and retained or destroyed according to applicable regulations. No investigational product will be destroyed until authorized in writing by the sponsor.

#### 12.3 METHOD OF ASSIGNING SUBJECTS TO TREATMENT GROUPS

Original Plan – Subjects will be randomly assigned in a blinded manner using the centralized IRT, with subjects assigned in a 1:1:1:1:1 ratio to each treatment group, according to pre-generated randomization schedules. Approximately 750 healthy male and female subjects between 18 and 84 years of age (inclusive) will be randomized in each country, with approximately 50% of subjects overall in the study ≥60 years of age. Two-factor, 2-level stratification will be employed (ages 18-59 and 60-84; study site). Up to approximately 1500 subjects could be enrolled for the 5 vaccine groups across the 2 countries to potentially mitigate the risk of enrollment in either country being delayed, or the expected availability of study data from either country being delayed due to the potential impact of the pandemic on feasibility of study vaccine delivery, possibilities for adequate specimen and data collection, and feasible specimen transportation. This flexibility would allow review of study data on approximately 500 subjects to facilitate initiation of Phase 3 clinical development as soon as possible. Details regarding the IRT process will be provided separately to the sites.

For the Part 2 component of the study, following study initiation, enrollment of older adult subjects (≥ 60 to 84 years of age) will be paused when approximately 50 subjects in the older age group are enrolled in each of the 5 study vaccine groups (ie, approximately 250 older adult subjects in total enrolled subjects across all sites). The SMC will then review reactogenicity data for both study age groups for 5 days following the first vaccine dose (Day 0 through Day 4, inclusive) when all subjects in the older adult group have accrued these 5 days of data. Enrollment of adult subjects (≥ 18 to 59 years of age) will not be paused during this SMC review unless general vaccination pause rules detailed in Section 13.1.5 are also met. Details of this review, and processes to restart enrollment of older adult subjects will be documented in the SMC Charter.

D189 Re-randomization - Participants initially randomized to Groups B and C will be re-randomized for the D189 booster dose at a 1:1 ratio to receive either 5 µg SARS-CoV-2 + 50 µg Matrix M adjuvant or placebo. In addition, subjects in Treatment Group E who were initially scheduled to receive a booster dose of vaccine at Day 189 will now receive placebo.

D357 Additional Dose - Subjects in Treatment Groups B and C who agree to continue study participation for an additional 6 months will receive a booster dose of vaccine (B1, B2, and C1) or placebo (C2) at Day 357 via a non-random assignment.

##### 12.3.1 Blinding Procedures

This is an observer-blinded study. To maintain the blind, placebo vaccination via IM route will be included, and unblinded site personnel will manage vaccine logistic, preparation, and administration (if necessary) so as to maintain the blind from the remainder of the site personnel and subjects.

For emergency unblinding associated with safety issues, site personnel who become unblinded will not be involved in study-related assessments or have subject contact for data collection following study vaccine administration. In this case, personnel at the clinical study site including, investigators and study staff, research site, and study subjects will remain blinded to subject treatment assignments until the end of study.

For unblinding associated with receipt of an approved/authorized vaccine, site personnel who become unblinded may continue to be involved in follow-up safety monitoring. It is expected that a significant number of study subjects will have the opportunity to receive an approved/authorized vaccine to prevent COVID-19 following the interim analysis at Day 35. It will be difficult for sites to maintain the blind for these subjects since the approved/authorized vaccine will be documented as a concomitant medication in the subject's medical records and in the study database which are readily available to blinded staff.

##### 12.3.2 Breaking the Blind

A subject's treatment assignment will not be broken until the end of the study for the clinical site study team unless medical treatment of the subject depends on knowing the study treatment the subject received or in the event that the Novavax study vaccine or another vaccine from a different manufacturer, is demonstrated to be safe and efficacious and approved/authorized by regulatory authorities in the US or Australia and the subject plans on receiving the approved/authorized vaccine. Subjects would be unblinded at EOS after all assessments are completed.

In the event that the blind needs to be broken because of a medical emergency or planned receipt of an approved/ authorized vaccine, the investigator may unblind an individual subject's treatment allocation. For a medical emergency, as soon as possible, the investigator should first contact the medical monitor to discuss the medical emergency and the reason for revealing the actual treatment received by that subject. In either case, the treatment assignment will be unblinded through IRT. Reasons for treatment unblinding must be clearly explained and justified in the eCRF. The date on which the code was broken together with the identity of the person responsible must also be documented.

In addition to the aforementioned situations where the blind may be broken, the data will also be unblinded to a statistical team at specified time points for interim analyses, as outlined in Section 14.5.

#### 12.4 TREATMENT COMPLIANCE

All doses of the study vaccine will be administered in the clinical unit under direct observation of clinic personnel and recorded in the eCRF. Clinic personnel will confirm that the subject has received the entire dose.

The location (right or left arm), date, and timing of all doses of study vaccine will be recorded in the subjects' eCRF. If a subject does not receive a dose of the study vaccine or does not receive the complete planned dose of the study vaccine, the reason will be recorded.

##### 12.4.1 Prior Vaccinations and Concomitant Medications

Administration of medications, therapies, or vaccines will be recorded in the eCRF. Concomitant medications will include all medications (including vaccines taken by the subject from the time of signing the ICF until Day 273 (or through the early termination visit if prior to that time) and thereafter will include only those medications associated with any MAAE attributed to vaccine, potential AESI, or SAE. Prescription and over-the-counter drugs, as well as herbals, vitamins, and supplements, will be included. Receipt of an authorized or approved COVID-19 vaccine from another manufacturer should be recorded as a concomitant medication through EOS.

### 13. STUDY PROCEDURES

Before performing any study procedures, all potential subjects will sign an ICF as outlined in Section 16.2.2.3. Subjects will undergo study procedures at the time points specified in the SOE (Table 10-4 for all treatment groups and additionally in Table 10-5 for Treatment Groups B and C). The total amount of blood collected from each subject over the duration of the study, including any extra assessments that may be required, will not exceed 500 mL. Subjects in the PBMC immunogenicity subset will have scheduled blood draws that should not exceed a total of 300 mL in volume over the duration of the study, which includes three 51 mL PBMC whole blood draws (at Day 0, Day 7, and Day 28). Subjects not in the PBMC immunogenicity subset will have scheduled blood draws that should not exceed a total of 150 mL in volume over the duration of the study. Unscheduled visits may be associated with additional blood draws, the volume of which would depend on clinical or study requirements following investigator assessment.

#### 13.1 SAFETY ASSESSMENTS

The timing and frequency of all safety assessments are listed in the SOE (Table 10-4 for all treatment groups and additionally in Table 10-5 for Treatment Groups B and C).

Safety assessments will include monitoring and recording of solicited (local and systemic reactogenicity events) and unsolicited AEs; MAAEs; AESI; SAEs; vital sign measurements; and physical examination findings. Recording of solicited and unsolicited AEs may be conducted by electronic data capture/reporting. Monitoring for PIMMC and AESI specific to potential disease enhancement for COVID-19 will be continued as noted for Part 1 of study.

**Note:** If a US subject is unblinded in the study and receives an approved/authorized vaccine from different manufacturer, all vaccine administration errors, all SAEs, cases of multisystem inflammatory syndrome, and hospitalized or fatal cases of COVID-19 following vaccination with the approved/authorized vaccine must be reported to the Vaccine Adverse Event Reporting System (VAERS). Investigators in Australia should follow local regulatory reporting guidance for safety events that occur in subjects who receive approved/authorized vaccines within the study.

##### 13.1.1 Adverse Events

Adverse events will be assessed during the study as described in the SOE (Table 10-4 for all treatment groups and additionally in Table 10-5 for Treatment Groups B and C) and should be followed until they are resolved, stable, or judged by the investigator to be not clinically significant. Adverse events will be captured after first dose of study vaccination administered with the exception of an AE related to study procedure or one that causes a delay in study vaccination administration (eg, acute illness).

The investigator is responsible for ensuring that all AEs and SAEs are recorded in the eCRF and reported to the sponsor, regardless of their relationship to study vaccine or clinical significance. If there is any doubt as to whether a clinical observation is an AE, the event should be reported.

###### 13.1.1.1 Adverse Event Definitions

The investigator is responsible for reporting all AEs that are observed or reported during the study, regardless of their relationship to study vaccination or their clinical significance.

An AE is defined as any untoward medical occurrence in a subject enrolled into this study regardless of its causal relationship to study vaccination. Subjects will be instructed to contact the investigator at any time after randomization if any symptoms develop.

###### 13.1.1.1.1 Serious Adverse Events

An SAE is defined as any event that

- results in death
- is immediately life threatening
- requires inpatient hospitalization or prolongation of existing hospitalization
- results in persistent or significant disability/incapacity
- is a congenital anomaly/birth defect

Important medical events that may not result in death, be life threatening, or require hospitalization may be considered SAEs when, based upon appropriate medical judgment, they may jeopardize the subject or may require medical or surgical intervention to prevent one of the outcomes listed in this definition. Examples of such medical events include allergic bronchospasm requiring intensive treatment in an emergency room or at home, blood dyscrasias or convulsions that do not result in inpatient hospitalization, or the development of drug dependency or drug abuse.

If investigators become aware of any SAEs after the study follow-up period has finished that they believe are related to study vaccination, such as potential cases of vaccine-induced immune enhancement of COVID-19, investigators should report these events to the sponsor.

[redacted]

###### 13.1.1.1.2 Abnormal Laboratory Tests Reported as Adverse Events

Any abnormal laboratory test results (hematology or serum chemistry) collected after first study vaccination considered clinically significant by the investigator should be recorded as an AE. If the laboratory finding did not worsen from baseline, the abnormality is considered part of medical history and not an AE.

However, if an underlying disease or new condition is identified that accounts for the clinically significant laboratory finding, the disease or condition should be documented as the AE instead of the laboratory abnormality.

###### 13.1.1.1.3 Local and General Systemic Reactogenicity Symptoms

Subjects will utilize an electronic subject-reported outcome application to record injection site-specific local (arm) and general systemic reactogenicity following vaccination. All subjects will record reactogenicity on the day of vaccination and for an additional 6 days after vaccination. Subject reported solicited symptoms are assumed to be causally related. Should any reactogenicity event extend beyond 6 days after vaccination and be clinically significant by toxicity grade 1 or greater, then it will be recorded as an AE with start dates of Day 7 and Day 28 (the first day after completion of the subject solicited symptom diary) and followed to resolution per FDA guidelines for dataset capture. For the 6-month and 12-month boosts, reactogenicity will be recorded as an AE with start dates of Day 196 and Day 364, respectively. The standard toxicology grading scale implemented in the electronic diary for Part 2 of the study is included in Section 16.3 (Appendix 3, Table 16-2).

###### 13.1.1.1.4 Adverse Events of Special Interest

Subjects will be assessed for diagnosis of an AESI at all study visits. Adverse events of special interest include PIMMC, AESIs relevant to COVID-19 disease, or other potential AEs that may be determined at any time by regulatory authorities as additional information concerning COVID-19 is obtained. Given the concern for cytokine storm, an AESI of cytokine release syndrome will be included as an AESI relevant to COVID-19 disease. Listings of AESI are presented in Appendix 4 (Section 16.4).

###### 13.1.1.1.5 Medically Attended Adverse Events

A MAAE is defined as an AE that leads to an unscheduled visit to a healthcare practitioner.

###### 13.1.1.1.6 Pregnancy

Pregnancy is not considered an AE unless there is a suspicion that an investigational vaccine may have interfered with the effectiveness of a contraceptive medication. Any pregnancy that occurs during study participation must be reported using a clinical study pregnancy form. To ensure subject safety, each pregnancy must be reported to Novavax, Inc. within 2 weeks of learning of its occurrence. If pregnancy occurs, further vaccination will be discontinued. The pregnancy must be followed up to determine outcome (including spontaneous miscarriage, elective termination, normal birth, or congenital abnormality) and the status of both mother and child, even if the subject was discontinued from the study. Pregnancy complications and elective terminations for medical reasons must be reported as an AE or SAE. Spontaneous miscarriages must be reported as an SAE.

Any pregnancy brought to the investigator's attention before the study is completed should be reported to PPD Pharmacovigilance using the pregnancy reporting forms provided to sites.

[redacted]

Any pregnancy brought to the investigator's attention after the subject has completed the study but occurring while the subject was in the study must be promptly reported to:

Sponsor Safety Monitor: [redacted]

###### 13.1.1.2 Eliciting and Documenting Adverse Events

At every study visit, subjects will be asked a standard question to elicit any medically related changes in their well-being. They will also be asked if they have been hospitalized, had any accidents, used any new medications, or changed concomitant medication regimens (both prescription and over-the-counter medications).

In addition to subject observations, AEs will be documented from any data collected on the AE page of the eCRF or other documents that are relevant to subject safety.

###### 13.1.1.3 Reporting Adverse Events

All AEs reported or observed during the study will be recorded on the AE page of the eCRF. Information to be collected includes study treatment, type of event, time of onset, dosage, investigator-specified assessment of severity and relationship to study vaccine and/or study procedure, time of resolution of the event, seriousness, any required treatment or evaluations, and outcome. Any AEs resulting from concurrent illnesses, reactions to concurrent illnesses, reactions to concurrent medications, or progression of disease must also be reported. All AEs will be followed until they are resolved, stable, or judged by the investigator to be not clinically significant. MedDRA will be used to code all AEs.

Any medical condition or laboratory finding that is present at the time that the subject is screened but does not deteriorate should not be reported as an AE. However, if it deteriorates at any time during the study, it should be recorded as an AE.

Any AE that is considered serious by the investigator or that meets SAE criteria (Section 6.1.1.1) must be reported to the sponsor immediately (within 24 hours after the investigator has confirmed the occurrence of the SAE). The investigator will assess whether there is a reasonable possibility that the study vaccine caused the SAE. The sponsor will be responsible for notifying the relevant regulatory authorities of any SAE, in compliance with health authority requirements, as outlined in the relevant clinical trial guidelines. The investigator is responsible for notifying the independent HREC/IRB directly.

For this study, the following contact information will be used for SAE reporting:

PPD Medical Monitor: [redacted]

###### 13.1.1.4 Assessment of Severity

The severity (or intensity) of an AE refers to the extent to which it affects the subject's daily activities and will be classified as mild, moderate, or severe using the following criteria:

- Mild (grade 1): These events require minimal or no treatment and do not interfere with the subject's daily activities.
- Moderate (grade 2): These events result in a low level of inconvenience or require minor therapeutic measures. Moderate events may cause some interference with normal functioning.
- Severe (grade 3): These events interrupt a subject's usual daily activity and may require systemic drug therapy or other treatment. Severe events are usually incapacitating.

If the severity of an AE changes, the most intense severity should be reported. An AE characterized as intermittent does not require documentation of the onset and duration of each episode.

###### 13.1.1.5 Assessment of Causality

The investigator's assessment of an AE's relationship to study vaccine is part of the documentation process, but it is not a factor in determining what is or is not reported in the study. If there is any doubt as to whether a clinical observation is an AE, the event should be reported.

The investigator will assess causality (ie, whether there is a reasonable possibility that the study vaccine caused the event) for all AEs and SAEs (solicited reactions are to be considered as being related to vaccination).

The relationship will be classified as follows:

- Not related: There is not a reasonable possibility of relationship to study vaccine. The AE does not follow a reasonable temporal sequence from administration of study vaccine or can be reasonably explained by the subject's clinical state or other factors (eg, disease under study, concurrent diseases, and concomitant medications).
- Related: There is a reasonable possibility of relationship to study vaccine. The AE follows a reasonable temporal sequence from administration of study vaccine and cannot be reasonably explained by the subject's clinical state or other factors (eg, disease under study, concurrent diseases, or concomitant medications), represents a known reaction to study vaccine or other vaccines in its class, is consistent with the known pharmacological properties of the study vaccine, and/or resolves with discontinuation of the study vaccine (and/or recurs with re-challenge, if applicable).

###### 13.1.1.6 Follow-up of Adverse Events

All AEs must be reported in detail on the appropriate page of the eCRF and followed until they are resolved, stable, or judged by the investigator to be not clinically significant.

##### 13.1.2 Clinical Laboratory Testing

Clinical laboratory tests will be performed at a designated laboratory (to be determined prior to initiation of Part 2). The clinical laboratory that will perform the tests will provide the reference ranges for all clinical laboratory parameters.

The following clinical laboratory assessments will be performed at screening for testing to provide a reference point for evaluation of later AEs should they occur, as needed. None of these laboratory tests are exclusionary for enrollment or vaccination. Laboratory tests may be included in unscheduled visits if deemed necessary by the investigator.

| | |
|---|---|
| Hematology | Hemoglobin, hematocrit, platelet count, and complete white blood cell count |
| Serum Chemistry | Alanine aminotransferase, aspartate aminotransferase, total bilirubin, creatinine |
| Serology | Hepatitis B surface antigen, hepatitis C virus antibody, and human immunodeficiency virus antibody types 1 and 2 |
| Other analyses | Female subjects: Urine pregnancy test at screening and prior to each vaccination (human chorionic gonadotropin); serum follicle-stimulating hormone may be requested to confirm postmenopausal status |

Note: A serum pregnancy test may be substituted for a urine pregnancy test at screening or at the discretion of the investigator.

Any abnormal laboratory test results (hematology or serum chemistry) or serology results identified from bloods collected prior to randomization may be followed up by investigators if considered clinically significant. Since screening and randomization visits may occur on the same day, baseline laboratory test results are not exclusionary for enrollment and will not always be available prior to randomization and first vaccine administration. Additional unscheduled laboratory tests may be collected at unscheduled or later scheduled visits at the investigator's discretion.

##### 13.1.3 Vital Sign Measurements

Vital sign measurements by the site will include body temperature (measured by preferred site method), pulse rate and diastolic and systolic BP (after subject is seated for at least 5 minutes). Subjects will record oral temperature using the provided thermometers during general systemic reactogenicity evaluation. The other vital sign measurements will be recorded as continuous variables prior to each vaccination.

On vaccination days, vital sign measurements will be collected at least once before vaccination to ensure subject has controlled BP and heart rate and no evidence of fever prior to vaccination and as clinically needed during the 30-minute post-vaccination observation period. If individual vital sign measurements are considered clinically significant by the investigator, vaccination may be withheld that day, and subjects may return on a subsequent day for re-evaluation and vaccination, ideally, within the time window specified in the SOE (Table 10-4 for all treatment groups and additionally in Table 10-5 for Treatment Groups B and C).

##### 13.1.4 Physical Examinations

A physical examination will be performed at screening (at minimum, assessment of skin, neck, thyroid, lungs, heart, cardiovascular, abdomen, lymph nodes of the upper extremities and neck, and musculoskeletal system/extremities). Height and weight will be measured and BMI will be calculated at screening only.

A targeted or symptom-directed physical examination will be performed at the time points specified in the SOE (Table 10-4 for all treatment groups and additionally in Table 10-5 for Treatment Groups B and C).

##### 13.1.5 Vaccination Pause Rules

AEs meeting any one of the following criteria will result in an immediate enrollment or further dosing hold for Part 2 of the study, pending further review by the SMC at the direction of the SMC Chair:

- Any toxicity grade 3 (severe) solicited single AE term across all study vaccine groups (blinded) occurring in ≥5% of subjects for either study age group (after a minimum of 100 subjects are enrolled in either age group) following vaccination (first, second vaccinations and booster doses to be assessed separately).

- Any grade 3 (severe) unsolicited single AE preferred term for which the investigator assesses as related which occurs in ≥7 subjects overall (both age groups) within 35 days following first dose vaccination.

In addition, any SAE assessed as related to vaccine (by study investigator and/or the sponsor) will be reported by the sponsor to the SMC Chair as soon as possible, and within 24 hours of the sponsor's awareness of the event. Based on this initial report of the event to the SMC Chair, the Chair may advise the sponsor to immediately pause enrolment and further dosing in either some or all subjects in the study and to convene an ad hoc meeting, or make alternative recommendations. The SMC Charter defines processes for how this review will occur and how the Chair's recommendations will be documented.

The sponsor, along with medical monitor, may request an SMC review for any safety concerns that may arise in the trial and not associated with any specific pause rule.

##### 13.1.6 Safety Monitoring

Safety oversight will be conducted by an SMC that is an independent group of experts that monitors subject safety and advises Novavax, Inc. The SMC members will be separate and independent of site personnel participating in this study and should not have a scientific, financial, or other conflict of interest related to this study or the sponsor. The SMC will consist of members with appropriate expertise to contribute to the interpretation of the data from this study.

The SMC will operate under the rules of a sponsor-approved charter that will be approved at the organizational meeting of the SMC. At this time, each data element that the SMC needs to assess will be clearly defined. Procedures for SMC reviews/meetings will be defined in the charter. Additional data may be requested by the SMC, and interim statistical reports may be generated as deemed necessary and appropriate for reviews. The SMC may receive data in aggregate and presented by treatment group. The SMC may also be provided with expected and observed rates of the expected AEs in an unblinded fashion and may request the treatment assignment be unblinded for an individual subject if required for safety assessment.

#### 13.2 IMMUNOGENICITY ASSESSMENTS

Blood samples will be taken at the time points specified in the SOE (Table 10-4 for all treatment groups and additionally in Table 10-5) for Treatment Groups B and C) to assess immune response. An additional blood sample may be collected within 14 days of the onset of any symptoms associated with SARS-CoV-2 infection, if possible, up through Day 217. Immune measurements (ELISA) will be conducted on serum (IgG and ACE2 receptor inhibition) for SARS-CoV-2 rS protein antigen(s). Neutralization testing and cell-mediated immunity following in vitro stimulation of PBMCs will occur at selected time points for a subset of subjects. In the case of the cell-mediated immunity testing subset, a sampling scheme based on visit, age strata, or other factors (such as location in relation to PBMC processing capacity) may be utilized. Additional testing will occur with further assay development. Based on the emergence of new variants, including B.1.1.7, B.1.351, P.1 and others, immunogenicity testing may be performed for these new variants as well as against the original prototype Wuhan strain.

The details on the handling, processing, and shipping of immunogenicity samples will be provided separately in a laboratory manual.

Subjects will be asked to provide consent for the use of samples for future testing for other viruses and/or sequencing of the SARS-CoV-2 in positive specimens or assay development specific to SARS-CoV-2 (or related variants). Aliquots of all collected samples from this study may be retained for the stated purposes for a maximum of 25 years (starting from the date at which the last subject had the last study visit), unless local rules, regulations, or guidelines require different timeframes or different procedures, in accordance with subject consent.

#### 13.3 COVID-19 DISEASE ASSESSMENTS (EFFICACY)

##### 13.3.1 COVID-19 Disease Monitoring

###### 13.3.1.1 Monitoring of Qualifying Symptoms of Suspected COVID-19 Disease

Note: Based on the extremely low incidence of SARS-CoV-2 transmission in Australia, monitoring for symptoms associated with SARS-CoV-2 infection via the ePRO system, self-sampling and completion of the electronic FLU-PRO instrument will no longer be conducted for participants enrolled at Australian sites following approval of Amendment 7 (Version 8.0) of the protocol.

Subjects at US sites will continue to be monitored via ePRO system (data capture) for symptoms associated with SARS-CoV-2 infection (eg, cough, fever, sore throat, difficulty breathing, and other symptoms [Table 13-1]) every 14 days beginning at Day 28 until approximately Day 217. When COVID-19 disease symptom scoring indicates the need for sample collection for potential PCR confirmation, subjects at sites in the US will self-collect a nasal mid-turbinate sample taken as close to the onset of symptoms as possible (ideal timing within 3 days). Symptom-based collection can be either due to passive or active surveillance using an ePRO system. Samples will not be collected after 14 days as a new symptom-based query will be initiated (eg, every 2 weeks).

Since self-sampling specimens for PCR confirmation under consideration for use in the study are not currently approved for diagnostic use, and depending on the site locations in relation to central study laboratories and associated delays in reporting related to specimen transportation, study PCR sampling is not an alternative for subjects to obtain diagnosis of potential COVID-19 in the context of individual clinical and public health case management. Subjects will be notified by sites of any positive or indeterminant PCR results when they are available and liaise with subjects' regular treating physicians in this instance. Negative study PCR results will not be routinely communicated to subjects, since a negative result for a non-diagnostic level test should not be interpreted as confirmation of absent SARS-CoV-2 infection. If subjects request results for PCR and results were negative, sites may provide the negative result to subjects, along with the context that negative results for the test should not be interpreted as excluding SARS-CoV-2 infection. Sites will follow required local processes around the notification of positive SARS-CoV-2 infections, which may involve public health reporting. Sites will assess local processes and availability of diagnostic quality SARS-CoV-2 infection testing and will communicate which options are locally available to subjects if they meet local clinical or public health criteria for testing.

Clinical symptoms of suspected COVID-19 disease from Day 0 to Day 28 will be reported to sites by subjects directly to sites by site-established communication channels outside of the ePRO system, and site staff will clinically assess the symptomatology and actions required that are consistent with locally applicable guidance for diagnosis of SARS-CoV-2 infection, which may vary by jurisdiction. If clinically indicated, actions could include unscheduled visits with site staff sampling of subjects, or referral of subjects into locally established PCR testing, with co-ordination with regular subject clinicians or clinicians locally established to assess and monitor potential COVID-19 cases.

**Table 13-1 Qualifying Symptoms of Suspected COVID-19 Disease**

| |
|---|
| Chills or fever or feverishness (reported by the subject, or fever ≥ 37.8°C, regardless of use of anti-pyretic medications) OR |
| New onset of any respiratory symptoms (cough, rapid breathing, shortness of breath, difficulty breathing, runny nose, nasal congestion, or sore throat) OR |
| New onset of the following other symptoms:<br>• Anosmia (smell disturbances)<br>• Ageusia (taste disturbances)<br>• Fatigue or tiredness or weakness<br>• Myalgia (muscle aches)<br>• Headache<br>• Nausea or vomiting<br>• Diarrhea |

Abbreviations: COVID-19 = coronavirus disease 2019.

###### 13.3.1.2 COVID-19 Disease Case Ascertainment – Day 28 Through Day 217

###### 13.3.1.2.1 Sample Collection for COVID-19 Laboratory Confirmation

When a subject at a US site reports any of the qualifying symptoms of suspected COVID-19 disease listed in Table 13-1 after Day 28, he/she will begin daily self-collection of a nasal mid-turbinate sample for 2 consecutive days (self-collection on any of these days may be replaced by sample collection by an HCP, if the subject is admitted to the hospital or COVID-19 disease treatment facility and self-collection is not available). Timing of self-collection should be as close to the onset of symptoms as possible (ideally within 3 days). All nasal samples collected over the 2-day period will be sent to a prespecified central laboratory where a validated PCR test will be performed for confirmation of SARS-CoV-2 infection. If both self-collected samples that are initially collected are PCR negative or indeterminate, and the subject has a clinical illness consistent with COVID-19 disease, site staff may request subjects to resample by nasal mid-turbinate sample collection for a further 2 consecutive days any time from 5 days after symptom onset.

Subjects enrolled at both US and Australian sites may also be seen by site personnel within the surveillance period to evaluate for potential COVID-19 disease, if acceptable based on the ongoing pandemic and subject containment requirements; and if seen, then a nasal mid-turbinate sample and an additional blood sample for immunogenicity assessments will be obtained. Any subject admitted to the hospital or COVID-19 treatment facility may utilize hospital testing for SARS-CoV-2 as the method of testing positivity. Endpoint collection will be obtained using hospital derived information which may include electronic medical records. For subjects enrolled at both US and Australian sites, any outpatient or public health PCR sample testing that has confirmed SARS-CoV-2 infection may be used to establish subject positivity for COVID-19 for study endpoints, provided subjects can provide some form of appropriate confirmation documentation.

Should a medical visit be warranted based on symptomatology (and allowed via local isolation guidance), such a visit may occur using telemedicine, home visitation, or clinic visit. Subjects experiencing shortness of breath or difficulty breathing must be seen by an HCP to assess tachypnea and blood oxygen saturation levels.

###### 13.3.1.2.2 PCR Resampling for Subsequent Illness Episodes

If a subject becomes symptom-free (of the qualifying symptoms of suspected COVID-19) for 3 consecutive days, then the subject will be eligible for PCR resampling at the next onset of qualifying symptoms of suspected COVID-19 diseases.

###### 13.3.1.2.3 FLU-PRO Disease/Symptom Severity Patient-Reported Outcome Instrument

The severity and symptom duration after qualifying symptoms of suspected COVID-19 disease have been met will also be monitored by subjects enrolled at US sites using the electronic FLU-PRO instrument, which may also include additional questions for COVID-19 disease depending on the status of validation of modified questions at the time of the study. The FLU-PRO will be completed daily up to 10 consecutive days if possible within the electronic collection tool by the patient, when a subject actively initiates reporting of qualifying symptoms for a potential COVID disease within the electronic patient diary, starting after qualifying symptoms of suspected COVID-19 disease have been met (Table 13-1). However, if a subject reports in the additional questions after the FLU-PRO instrument symptoms questions within the electronic diary for 3 consecutive days during the 10-day follow-up period that they have returned to their usual health, then completion of the FLU-PRO tool on further days need not continue. If subjects then have the return of potential qualifying symptoms as part of a new illness episode, then the subject can have the new illness episode assessed again for potential qualifying symptoms through the electronic diary. FLU-PRO assessments therefore should only be associated with one potential qualifying illness episode at a time.

In case of severe clinical deterioration, the instrument may not be able to be completed for the full 10-day period. Subjects can only be reporting FLU-PRO scores in relation to one illness event at a time. Sites may use the FLU-PRO scores over time to monitor subjects' self-reported symptomatology during illness episodes.

###### 13.3.1.2.4 COVID-19 Clinical Endpoint Disease Severity Definitions

Virologically confirmed COVID-19 disease (assessed by severity) will be graded as follows (virologically confirmed, mild, moderate, or severe) based on progression to the greatest severity during the course of illness (Table 13-2).

**Table 13-2 Endpoint Definitions of COVID-19 Disease Severity**

| COVID-19 Severity | Endpoint Definitions |
|---|---|
| **Virologically Confirmed** | ≥ 1 COVID-19 disease symptom in Table 13-1<br>**AND**<br>Does not meet criteria for mild, moderate or severe disease |
| **Mild** | ≥ 1 of:<br>• Fever (defined by subjective or objective measure, regardless of use of anti-pyretic medications)<br>• New onset cough<br>• ≥ 2 COVID-19 respiratory/non-respiratory symptoms in Table 13-1<br>**AND**<br>• Does not meet criteria for moderate or severe |
| **Moderate** | ≥ 1 of:<br>• Fever(defined by subjective or objective measure, regardless of use of anti-pyretic medications) + any 2 COVID-19 respiratory/non-respiratory symptoms in Table 13-1 for ≥ 3 days (need not be contiguous days)<br>• High fever (≥ 38.4°C) for ≥ 3 days (need not be contiguous days)<br>• Any evidence of significant LRTI:<br>– Shortness of breath (or breathlessness or difficulty breathing) with or without exertion (greater than baseline)<br>– Tachypnea: 20 to 29 breaths per minute at rest<br>– $SpO_2$: 94% to 95% on room air<br>– Abnormal chest x-ray or CT consistent with pneumonia or LRTI<br>– Adventitious sounds on lung auscultation (eg, crackles/rales, wheeze, rhonchi, pleural rub, stridor)<br>**AND**<br>• Does not meet criteria for severe disease |
| **Severe** | ≥ 1 of:<br>• Tachypnea: ≥ 30 breaths per minute at rest<br>• Resting heart rate ≥ 125 beats per minute<br>• $SpO_2 \leq 93\%$ on room air or $PaO_2/FiO_2 < 300$ mmHg<br>• High-flow oxygen therapy or NIV/NIPPV (eg, BiPAP or CPAP),<br>• Mechanical ventilation or ECMO<br>• One or more major organ system dysfunction or failure (eg, cardiac/circulatory, pulmonary, renal, hepatic, and/or neurological, to be defined by diagnostic testing/clinical syndrome/interventions), including any of the following<br>○ Acute respiratory distress syndrome (ARDS)<br>○ Acute renal failure<br>○ Acute hepatic failure<br>○ Acute right or left heart failure<br>○ Septic or cardiogenic shock (with shock defined as SBP < 90 mm Hg OR DBP < 60 mm Hg)<br>○ Acute stroke (ischemic or hemorrhagic)<br>○ Acute thrombotic event: AMI, DVT, PE<br>○ Requirement for: vasopressors, systemic corticosteroids, or hemodialysis<br>○ Admission to an ICU<br>○ Death |

Abbreviations: AMI = acute myocardial infarction; BiPAP = bi-level positive airway pressure; CPAP = continuous positive air pressure; CT = computed tomography; DBP = diastolic blood pressure; DVT = deep vein thrombosis; ECMO = extracorporeal membrane oxygenation; $FiO_2$ = fraction of inspired oxygen; ICU = intensive care unit; LRTI = lower respiratory tract infection; NIPPA = non-invasive positive pressure ventilation; NIV = non-invasive ventilation; $PAO_2$ =

partial pressure of oxygen in the alveolus; PE = pulmonary embolism; SBP = systolic blood pressure; $SpO_2$ = oxygen saturation.

###### 13.3.1.2.5 COVID-19 Clinical Endpoint Case Definition

A subject whose nasal mid-turbinate sample tests positive for SARS-CoV-2 infection by qualitative PCR [ie, (+)-PCR-confirmed SARS-CoV-2 illness) and whose initial qualifying symptom(s) of suspected COVID-19 disease (Table 13-1) meets the criteria for symptomatic virologically confirmed, mild, moderate, or severe COVID-19 disease (Table 13-2) within 14 days of the initial qualifying symptom(s) will be defined as a single case in the secondary efficacy analysis.

Subjects will be instructed on self-collection using a nasal mid-turbinate sample approach and should demonstrate competency during an early clinic visit. An ePRO system will be employed for monitoring/documenting potential COVID-19 disease symptoms based on the occurrence of symptoms from a standard list (see Table 13-1), with subjects being queried every 14 days. Subjects may also trigger collection between the 14-day query periods if symptoms are reported spontaneously and conform to the algorithm to trigger self-collection. Self-collection should occur within 3 days of symptom onset (if possible). Subjects will also be notified through the electronic patient diary to self-collect specimens every 28 days if symptom free to assess for asymptomatic carrier status.

Should a subject be admitted to the hospital or COVID-19 intensive care ward and sample self-collection is unavailable, then a local public health or hospital test will be taken as a valid result. All PCR samples taken through the study will be sent to a prespecified central laboratory where a validated PCR test will be performed. When a subject actively initiates reporting of qualifying symptoms for a potential COVID disease within the electronic patient diary, follow-up patient-reported outcomes will be collected using the FLU-PRO instrument for up to 10 days, and the duration of resolved illnesses (number of days from the start of illness symptoms until the day at which health returned to normal). Exploratory analyses may also be conducted looking at the time course and maximal values of FLU-PRO total and domain level scores during an assessed illness episode, and how the proportion of subjects with maximal scores relate to COVID-19 study-defined illness severity categories. Should a medical visit be warranted based on symptomatology (and allowed via local isolation guidance), such a visit may occur using telemedicine, home visitation, or clinic visit.

Subjects will be notified of new positive PCR-confirmed SARS-CoV-2 infection status due to requirements of self-isolation and potential transmission. Since self-collection PCR-confirmation assay methodologies that are likely to be used in this study are not expected to be licensed for diagnostic purposes, and likely relative delays in the availability of results due to transportation and testing timelines, collection of study samples for PCR-confirmation do not replace the need for subjects to also be tested and followed up through public health testing and management processes that are in place locally if they meet criteria for local testing.

To further describe the epidemiologic evolution of the pandemic and potential effect of vaccination, assessment of SARS-CoV-2 by qualitative PCR testing based on routine screening without symptomatology will be done by nasal mid-turbinate sample self-collection from Day 28 through approximately 6 months.

Asymptomatic infection, period of transmission, and other defined infection parameters will be assessed along with severity and progression of disease.

- **Asymptomatic infection** will be defined as a positive PCR-confirmed SARS-CoV-2 infection with no symptoms (regardless of past positivity) in the 7 days prior to self-collection.

- **Primary infection** will be defined as the first positive PCR-confirmed SARS-CoV-2 infection regardless of symptoms.

- **Primary symptomatic infection** will be defined as the first positive PCR-confirmed SARS-CoV-2 infection that is triggered by the symptomatic algorithm. A symptomatic infection will initiate further evaluation of physical status with severity scoring applied and monitoring of duration.

- **Period of transmission** for any subject (eg, transmission potential) is the period from a positive PCR test until the time of the first negative PCR test that follows a positive PCR test (if follow-up PCR test results are available for that subject).

- **Duration of resolved illness episodes** (ie, number of days from the start of illness symptoms for illnesses followed up by FLU-PRO assessments until the day at which health returned to normal as reported in the electronic diary) classified by COVID-19 disease severity (Table 13-2). Duration will not be calculated for asymptomatic infections or illness episodes in which the subject's health has not returned to normal at the time of the data cut.

### 14. STATISTICAL ANALYSIS PLANS

#### 14.1 SAMPLE SIZE CALCULATIONS

The decision on the choice of formulation and dosing regimen will be made based on the totality of the immunogenicity and safety data rather than any individual measurement. No multiplicity adjustment will be made for this early phase study where multiple treatment groups and endpoints are being evaluated.

With 150 subjects in each treatment group, there is a greater than 99.9% probability to observe at least 1 subject with an AE if the true incidence of the AE is 5% and a 77.9% probability if the true incidence of the AE is 1%.

Table 14-1 presents the width of the 95% CI (based on the Clopper-Pearson method) for 150 subjects in each treatment group (assuming 5% of dosed subjects will be excluded from the per-protocol analysis set) under SCR assumptions between 50% and 90%.

**Table 14-1 Width of the 95% Confidence Interval (based on the Clopper-Pearson method)**

| Seroconversion Rate (%) | 95% CI (%) | Width (%) of 95% CI |
|---|---|---|
| 50 | 41.7, 58.3 | 16.5 |
| 60 | 51.7, 67.9 | 16.2 |
| 70 | 62.0, 77.2 | 15.2 |
| 80 | 72.7, 86.1 | 13.4 |
| 90 | 84.0, 94.3 | 10.3 |

Notes: Width of 2-sided 95% confidence interval (based on the Clopper-Pearson method) for the specified seroconversion rate produced by a sample size of 150 subjects is presented. PASS 15.0.7 Confidence Intervals for One Proportion is used in the calculation.

Table 14-2 presents the power of detecting difference in SCRs between treatment groups for 150 subjects in each treatment group (assuming 5% of dosed subjects will be excluded from the per-protocol analysis set).

**Table 14-2 Power of Detecting Difference in Seroconversion Rates between Treatment Groups**

| Seroconversion Rate (%) in the Control Group | Seroconversion Rate (%) in the Comparison Group | Power of Detecting the Difference in Seroconversion Rate |
|---|---|---|
| 50 | 55 | 14.6% |
| | 60 | 42.9% |

| Seroconversion Rate (%) in the Control Group | Seroconversion Rate (%) in the Comparison Group | Power of Detecting the Difference in Seroconversion Rate |
|---|---|---|
| | 65 | 76.1% |
| | 70 | 94.7% |
| 60 | 65 | 14.3% |
| | 70 | 44.0% |
| | 75 | 79.6% |
| | 80 | 97.0% |
| 70 | 75 | 16.1% |
| | 80 | 51.9% |
| | 85 | 88.3% |
| | 90 | 99.4% |
| 80 | 85 | 20.6% |
| | 90 | 68.6% |
| | 95 | 98.5% |
| 90 | 95 | 36.9% |

Notes: Power of detecting the specified difference in SCRs between treatment groups, assuming a significance level of 0.05, produced by a sample size of 150 subjects in each treatment group is presented. PASS 15.0.7 Superiority by a Margin Tests for the Difference Between Two Proportions (Miettinen & Nurminen Likelihood Score Test) is used in the calculation.

Table 14-3 presents the power of detecting ratio of geometric means between treatment groups for 150 subjects in each treatment group (assuming 5% of dosed subjects will be excluded from the per-protocol analysis set).

**Table 14-3 Power of Detecting Ratio of Geometric Means between Treatment Groups**

| Ratio of GM (Difference of $Log_{10}$ Transformed Geometric Mean) | SD of Difference of $Log_{10}$ Transformed Geometric Mean) | Power |
|---|---|---|
| 1.5 (0.176) | 0.4 | 96.7% |
| | 0.5 | 86.0% |
| | 0.6 | 71.6% |
| 2 (0.301) | 0.4 | >99.9% |
| | 0.5 | >99.9% |
| | 0.6 | 99.1% |
| 4 (0.602) | 0.4 | >99.9% |
| | 0.5 | >99.9% |
| | 0.6 | >99.9% |

Notes: Power of detecting the specified difference of $log_{10}$ transformed geometric mean, assuming same SD for both treatment groups and a significance level of 0.05, produced by a sample size of 150 subjects in each treatment group is presented. PASS 15.0.7 Two-Sample T-Tests Assuming Equal Variance is used in the calculation.

#### 14.2 ANALYSIS SETS

The All Screened Subjects Analysis Set will include all subjects who sign the ICF.

The Intent-to-Treat (ITT) Analysis Set will include all subjects who are randomized, regardless of protocol violations or missing data.
The Full Analysis Set will include all ITT subjects who receive at least 1 dose of study vaccine (SARS-CoV-2 or placebo). The Full Analysis Set will be used for supportive analyses. Subjects will be analyzed according to the randomized treatment assignment.

The Safety Analysis Set will include all subjects who receive at least 1 dose of study vaccine (SARS-CoV-2 rS or placebo). Subjects will be analyzed according to the vaccine actually received. Actual treatment received will be assumed to correspond with randomized treatment assignment except in cases where a site reports having used incorrect investigational product during vaccine administration.

The Per-Protocol (PP) Analysis Set will be determined for each study visit and will include all subjects who receive the initial dose of study vaccine (SARS-CoV-2 rS or placebo) for all analyses through Day 21, all subjects who receive vaccine doses at both Day 0 and Day 21 for all analyses starting at Day 28, all subjects who receive vaccine doses at Days 0, 21, and 189 for all analyses starting at Day 217, and all subjects who receive vaccine doses at Days, 0, 21, 189, and 357 for all analyses starting at Day 371, have at least a baseline and 1 post-baseline serum sample IgG result available at the corresponding study visit have no major protocol violations (eg, having randomized treatment assignment unblinded due to medical emergency, having taken prohibited medication on-study such as chronic systemic glucocorticoids, having received a vaccination containing investigational product that differs from randomized treatment assignment) that impact immunogenicity response at the corresponding study visit. The review and determination for exclusion from the PP Analysis Set will be carried out in a blinded fashion by a study clinician prior to unblinding for each analysis based on all available information from either the locked database or from a database freeze, depending on the SAP defined analyses. All subjects in the PP Analysis Set will be analyzed according to the randomized treatment assignment. Any subject who is SARS-CoV-2 positive by qualitative PCR testing from screening and prior to the immunogenicity assessment for a particular time point will be excluded from the PP Analysis Set for that time point onward. The protocol violations that impact the immunogenicity response and impact upon the PP Analysis Set are detailed in the study deviation rules document.

The PP-Immunogenicity PBMC Subset will include all subjects in the PP Analysis Set but further restricted to subjects who had blood samples taken and PBMCs harvested for analysis of cell-mediated immunity.

The PP-Immunogenicity Analysis Set will include all subjects who received their full vaccination schedule (ie, Day 0 and Day 21, or Days 0, 21, and Day 189, or Days 0, 21, 189, and Day 357), have at least a baseline and a Day 35 serum sample IgG result available for analysis, have no major protocol violations up to and including Day 35 that impact immunogenicity response, and no evidence of positive serology at baseline (ie, hepatitis B, hepatitis C, or HIV). The PP-Immunogenicity Analysis Set will be used for the primary immunogenicity analysis (ie, serum IgG antibody levels specific for the SARS-CoV-2 rS protein antigen[s] as detected by ELISA using GMT or SCR for the 2-dose regimens at Day 35 regardless of baseline immune status). If the number of subjects in the PP-Immunogenicity and PP Analysis Set differ (defined as the difference divided by the total number of subjects in the given PP Analysis Set) by more than 10%, supportive analyses of immunogenicity may be conducted using the PP Analysis Set.

The PP-Efficacy Analysis Set will be the PP Analysis Set further restricted to subjects who received their full vaccination schedule (ie, Day 0 and Day 21). All subjects in the PP-Efficacy Analysis Set will be analyzed according to the randomized treatment assignment.

A listing of analysis sets (with reasons for exclusion, if applicable) will be provided.

#### 14.3 STATISTICAL ANALYSIS

Details of all statistical analyses will be described in a SAP.

All data collected will be presented in data listings. Data from subjects excluded from an analysis set will be presented in the data listings but not included in the calculation of summary statistics for that analysis set.

For categorical variables, frequencies and percentages will be presented. Continuous variables will be summarized using descriptive statistics (number of subjects, mean, median, minimum, and maximum).

Baseline demographic and background variables will be summarized by treatment group and age group. The number of subjects who enroll in the study and the number and percentage of subjects who complete the study will be presented. Frequency and percentage of subjects who withdraw or discontinue from the study, and the reason for withdrawal or discontinuation, will also be summarized.

##### 14.3.1 Safety Analyses

Numbers and percentages (with 95% CIs based on the Clopper-Pearson method) of subjects with solicited local and systemic AEs through 7 days after each vaccination will be summarized by treatment group and the maximum toxicity grade over 7 days after each vaccination. The duration of solicited local and systemic AEs after each vaccination will also be summarized by treatment group. If more than 10% of total subjects are seropositive to SARS-CoV-2 infection at baseline, exploratory analyses by SARS-CoV-2 positivity at Day 0 may be performed.

Unsolicited AEs will be coded by preferred term and system organ class using the latest version of MedDRA and summarized by treatment group as well as by severity and relationship to study vaccine. Unsolicited AEs from the first vaccination until before the second vaccination, from second vaccination through 35 days after first vaccination, and from boost until 28 days after boost vaccination; all MAAEs through 217 days after first vaccination; and any MAAE related to vaccine, SAE, or AESI through the end of study will be listed separately and summarized by treatment group and age strata (18-59, 60-84 years). Adverse events of special interest associated with PIMMC and COVID-19 disease potential exacerbation will be listed separately.

Vital sign measurements will be summarized by treatment group at each time point using descriptive statistics. Vital sign toxicity grading will be derived on day of vaccination.

Concomitant medications will be summarized by treatment group and preferred drug name as coded using the World Health Organization drug dictionary.

##### 14.3.2 Immunogenicity Analyses

The primary immunogenicity analyses will be performed using the PP-Immunogenicity Analysis Set (which is restricted relative to subjects in the PP Analysis Set who received their full vaccination schedule and have a Day 35 sample available for analysis regardless of baseline SARS-CoV-2 IgG serostatus), excluding those subjects with evidence of positive serology at baseline (ie, hepatitis B, hepatitis C, or HIV). Subgroup analyses of the primary immunogenicity analyses by SARS-CoV-2 IgG serostatus at baseline (as measured by in-house SARS-CoV-2 IgG assay at the Novavax central immunology laboratory) will be undertaken.

If the number of subjects in the Immunogenicity and PP Analysis Set differ (defined as the difference divided by the total number of subjects in the per-protocol analysis set) by more than 10%, supportive analyses of immunogenicity may be conducted using the PP Analysis Set.

For the serum IgG antibody levels and ACE2 receptor binding inhibition specific for the SARS-CoV-2 rS protein antigen(s) as detected by ELISA the geometric mean, the GMFR compared to the baseline (Day 0) and the SCR (proportion of subjects with ≥4-fold rises in ELISA units) at each post-vaccination study visit (except the Day 7 and Day 28 visits for the PBMC subset), and the GMFR comparing post- (Day 217) to pre- (Day 189), and post- (Day 371) to pre- (Day 357) along with 95% CI will be summarized by treatment group by visit. Both age strata (18-59, 60-84 years) and naïve versus non-naïve subjects at baseline will be analyzed. Definitions for naïve versus non-naïve subjects at baseline will be defined in the SAP.

A subset of subjects will have immunogenicity analyses performed using a wild-type SARS-CoV-2 neutralization assay at a minimum of the time points baseline (Day 0), Day 35, and Day 217). More details for these neutralization assays are documented in the SAP.

A subset of subjects will have neutralization assays, IgG, and hACE-2 testing for both the Wuhan and the B.1.351 strain at Days 189, 217, 357, 371, and 546. Immunogenicity assessments for other variants may also be performed.

The 95% CI will be calculated based on the t-distribution of the log-transformed values for geometric means or GMFRs, then back transformed to the original scale for presentation. The

SCR (proportion of subjects with ≥4-fold rises in ELISA units) along with 95% CIs based on the Clopper-Pearson method will be summarized by treatment group at each post-vaccination study visit. An ANCOVA model will be constructed at each post-vaccination study visit on the log-transformed titer, including the treatment group as a fixed effect and the baseline log-transformed titer as a covariate. Comparisons of selected treatment groups will be performed within each visit. Additional covariates such as site and age strata (18-59, 60-84 years) will be included as covariates in the ANCOVA model used to analyze serum IgG antibody levels, serum ACE-2 receptor binding inhibition, and neutralization assay results. The difference in the SCR between selected treatment groups along with 95% CIs will be calculated using the method of Miettinen and Nurminen (Miettinen and Nurminen 1985).

Similar summaries will be generated for the other immunogenicity endpoints.

Cell-mediated immunity will be measured using select cytokines (eg, IL-2, IL-4, IL-5, IL-6, IL-13, TNFα, INFγ) in harvested PBMC cells and will be summarized by treatment group, overall and by age strata (18-59, 60-84 years) (subset of subjects).

##### 14.3.3 Efficacy Analyses

Numbers and percentages (with 95% CIs based on the Clopper-Pearson method) of subjects with occurrence of SARS-CoV-2 positivity and classified by COVID-19 disease severity will be summarized by treatment group, overall and by age strata (18-59, 60-84 years) by those receiving SARS-CoV-2 rS compared to placebo. If frequent clinical PCR-confirmed SARS-CoV-2 infections occur during the study follow-up period, vaccine efficacy assessments for primary vaccination for treatment groups compared to placebo may be generated. The criteria to determine whether this will occur will be documented in the SAP.

Vaccine Efficacy (VE) is defined as VE (%) = (1 – RR) x 100, where RR = relative risk of incidence rates between the 2 treatment groups (SARS-CoV-2 rS / Placebo). The interim and final analyses will be carried out at the one-sided Type I error rate of 0.025 overall. A two-sided 95% CI for the VE for each primary endpoint will accompany the point estimate. The estimated RR and its CI will be derived using Poisson regression with robust error variance [Zou, 2004]. The explanatory variables in the model will include treatment group and age group. The dependent variable will be the incidence rate of the endpoint of interest. The robust error variances will be estimated using the repeated statement and the subject identifier. The Poisson distribution will be used with a logarithmic link function.

The duration of resolved illness episodes (number of days from the start of illness symptoms for illnesses followed up by FLU-PRO assessments, until the day at which health returned to normal as reported in the electronic diary) and classified by COVID-19 disease severity will be summarized by treatment group, overall and by age strata (18-59, 60-84 years).

###### 14.3.4 Exploratory Analyses

Daily FLU-PRO scores (for total score and/or domain subgroup scores) following subject reported SARS-CoV-2 positive illnesses may be summarized by treatment group and age subgroup using descriptive statistics. Maximal FLU-PRO scores during the daily follow-up for an illness episode (for total score and/or domain subgroup scores) by treatment group and age subgroup may also be assessed by study defined COVID-19 illness severity categories. Further details of exploratory analyses related to FLU-PRO scores will be defined in the SAP.

#### 14.4 HANDLING OF MISSING DATA

For calculating geometric means and GMFRs, immunogenicity values reported as below the LLOQ will be replaced by 0.5 × LLOQ. Values that are greater than the ULOQ will be replaced by the ULOQ. Missing results will not be imputed.

#### 14.5 INTERIM ANALYSES

A primary database lock will occur following Day 35 data for all primary endpoints and selected secondary endpoints defined in the SAP to allow assessment of treatment group responses in the Part 2 study, to inform decisions around recommended dose regimens by age strata, and to facilitate the initiation of the Phase 3 efficacy trial.

However, earlier database freezes may occur to potentially allow earlier initiation of Phase 3 efficacy trials in consultation with the SMC and regulatory agencies. These database freezes may be based on data accrued in either Australia or the United States or accrued across the 2 countries combined (depending on the timing of availability of results to allow assessments to occur).

Database freezes may occur when approximately 50 subjects for either age group (18-59, 60-84 years) of each of the 2 first dose formulations (5 µg + 50 µg and 25 µg + 50 µg) have: a) reached Day 21 for safety follow-up; and b) baseline (Day 0) and Day 21 serum IgG antibody levels are available for immunogenicity follow-up. Data for database freezes will be based on partially cleaned and verified data. Immunogenicity data at these reviews may contain neutralization data or other immunogenicity assessments that are available at the time of the database freeze.

Database freezes may also occur to assess the Day 35 data when approximately 50 subjects for either age group (18-59, 60-84 years) in vaccine groups B and D have: a) reached Day 35 for safety follow-up; and b) baseline (Day 0) and Day 35 serum IgG antibody levels are available for immunogenicity follow-up. Data for database freezes will be based on partially cleaned and verified data. Immunogenicity data at these reviews may contain neutralization data or other immunogenicity assessments that are available at the time of the database freeze.

Subsequent database freezes will occur at Days 105 and 217 (all treatment groups) and at Day 371 (Treatment Groups B and C) for analysis of selected primary and/or secondary endpoints with a final database lock at Day 546 (EOS). In addition, planned analyses for COVID-19 endpoints will be included in all scheduled SMC reviews to allow review of potential vaccine efficacy (if sufficient COVID-19 events occur in the jurisdiction(s) where the study is conducted) or safety concerns. The planned primary analyses will be performed by an unblinded biostatistics and programming team. The sponsor biostatistics and programming team may be provided with the immunogenicity data set with a limited number of variables and dummy subject identifiers if sponsor analyses are planned, such that sponsor biostatistics and programming team remain blinded at the subject level. The variables to be included in this blinded immunogenicity data transfer are the dummy subject identifiers, treatment group assignments, visit numbers, and assay identifiers, and results only with the dataset subset by the unblinded team to only those subjects meeting per-protocol population criteria for the planned analysis. Only group level unblinded summaries will be generated at planned analyses and subject level treatment assignment will not be released until the sponsor is unblinded at the subject level, or regulatory authorities request rolling submission of data from earlier study time points (which would require sponsor subject level unblinding). To the extent possible based on the availability of authorized COVID-19 vaccines, investigator/site and CRO subject level blinding will be maintained until the study is completed. Planned analyses will be documented in the SAP prior to initiating that specific analysis.

Novavax, Inc. SARS-CoV-2 Recombinant Spike Protein Nanoparticle Vaccine (SARS-CoV-2 rS). Investigator's brochure, 1st ed. Gaithersburg (MD); 2020. 43 p.

SPEAC: Safety Platform for Emergency Vaccines. D2.3 Priority List of Adverse Events of Special Interest: COVID-19 [internet]. V2.0; 2020 May 25 [cited 2020 Jun 19]. Available from: https://brightoncollaboration.us/wp-content/uploads/2020/06/SPEAC_D2.3_V2.0_COVID-19_20200525_public.pdf.

The Criteria Committee of the New York Heart Association (NYHA). Nomenclature and Criteria for Diagnosis of Diseases of the Heart and Great Vessels. 9th ed. Boston, Mass: Little, Brown & Co; 1994:253-256.

Zou G. A modified poisson regression approach to prospective studies with binary data. Am J Epidemiol. 2004; 159(7):702-6.

### 16. APPENDICES

#### 16.1 APPENDIX 1: LIST OF ABBREVIATIONS

| Abbreviation | Term |
|---|---|
| ACE2 | angiotensin converting enzyme 2 |
| AE | adverse event |
| AESI | adverse event(s) of special interest |
| ALT | alanine aminotransferase |
| ANCOVA | analysis of covariance |
| AST | aspartate aminotransferase |
| BMI | body mass index |
| BP | blood pressure |
| BUN | blood urea nitrogen (also referred to as urea) |
| CFR | Code of Federal Regulations |
| CI | confidence interval |
| COVID-19 | coronavirus disease 2019 |
| CV | coefficient of variation |
| EBOV | ebolavirus |
| eCRF | electronic case report form |
| ELISA | enzyme-linked immunosorbent assay |
| ELISpot | enzyme-linked immune absorbent spot |
| EOS | end of study |
| ePRO | electronic patient-reported outcomes |
| ER | emergency room |
| EU | ELISA units |
| FDA | US Food and Drug Administration |
| FLU-PRO | InFLUenza Patient-Reported Outcome |
| FSH | follicle-stimulating hormone |
| GLP | Good Laboratory Practice |
| GMC | geometric mean concentration |
| GMEU | geometric mean ELISA units |
| GMFR | geometric mean fold rise |
| GMT | geometric mean titer |
| GMEU | geometric mean ELISA units |
| GOLD | Global Initiative for Chronic Obstructive Lung Disease |

| Abbreviation | Term |
|---|---|
| HCG | human chorionic gonadotropin |
| HCP | healthcare provider |
| HIV | human immunodeficiency virus |
| HREC | human research ethics committee |
| IB | investigator's brochure |
| ICF | informed consent form |
| ICH | International Council for Harmonisation |
| IgG | immunoglobulin G |
| IL-13 | interleukin 13 |
| IL-2 | interleukin 2 |
| IL-4 | interleukin 4 |
| IL-5 | interleukin 5 |
| IL-6 | interleukin 6 |
| IM | intramuscular |
| INFγ | interferon gamma |
| IRB | institutional review board |
| IRT | Interactive Response Technology |
| ITT | Intent-to-Treat |
| JNC 8 | Eighth Joint National Committee |
| LLOQ | lower limit of quantification |
| MAAE | medically attended adverse event |
| MedDRA | Medical Dictionary for Regulatory Activities |
| NYHA | New York Heart Association |
| OTC | over-the-counter |
| PBMC | peripheral blood mononuclear cell |
| PCR | polymerase chain reaction |
| PIMMC | potential immune-mediated medical conditions |
| POC | point-of-care |
| PP | per-protocol |
| PT | preferred term |
| RBD | receptor binding domain |
| RSV | respiratory syncytial virus |
| SAE | serious adverse event |
| SAP | statistical analysis plan |
| SARS | severe acute respiratory syndrome |

| Abbreviation | Term |
|---|---|
| SARS-CoV | severe acute respiratory syndrome coronavirus |
| SARS-CoV-2 rS | severe acute respiratory syndrome coronavirus 2 recombinant spike (S) protein nanoparticle vaccine |
| SCR | seroconversion rate |
| Sf9 | *Spodoptera frugiperda* |
| SMC | safety monitoring committee |
| SOC | system organ class |
| SOE | schedule of events |
| SPR | seroprotection rate |
| Th1 | Type 1 T helper |
| Th2 | Type 2 T helper |
| TNFα | tumor necrosis factor alpha |
| ULN | upper limit of normal |
| ULOQ | upper limit of quantification |
| VE | vaccine efficacy |
| WBC | white blood cell |

#### 16.2 APPENDIX 2: STUDY GOVERNANCE

##### 16.2.1 Data Quality Assurance

This study will be conducted using the quality processes described in applicable procedural documents. The quality management approach to be implemented will be documented and will comply with current ICH guidance on quality and risk management. All aspects of the study will be monitored for compliance with applicable government regulatory requirements, current Good Clinical Practice, the protocol, and standard operating procedures. The monitor will maintain current personal knowledge of the study through observation, review of study records and source documentation, and discussion of the conduct of the study with the investigator and personnel. Electronic CRFs and electronic data capture will be utilized. The electronic data capture system is validated and compliant with US Title 21 CFR Part 11 and local regulations. Each person involved with the study will have an individual identification code and password that allows for record traceability.

Major protocol deviations, should they occur during the study, will be presented in Section 10.2 of the clinical study report, and all major and minor protocol deviations will be included in submission datasets.

##### 16.2.2 Investigator Obligations

The following administrative items are meant to guide the investigator in the conduct of the study and may be subject to change based on industry and government standard operating procedures, working practice documents, or guidelines. Changes will be reported to the HREC/IRB but will not result in protocol amendments.

###### 16.2.2.1 Confidentiality

All laboratory specimens, evaluation forms, reports, and other records will be identified in a manner designed to maintain subject confidentiality. All records will be kept in a secure storage area with limited access. Clinical information will not be released without the written permission of the subject, except as necessary for monitoring and auditing by the sponsor, its designee, relevant regulatory authority, or the HREC/IRB.

The investigator and all employees and coworkers involved with this study may not disclose or use for any purpose other than performance of the study, any data, record, or other unpublished, confidential information disclosed to those individuals for the purpose of the study. Prior written agreement from the sponsor or its designee must be obtained for the disclosure of any said confidential information to other parties.

###### 16.2.2.2 Institutional Review

Prior to initiation of a study site, regulatory authority regulations and the ICH E6(R2) guidelines require that approval be obtained from an HREC/IRB before participation of human subjects in research studies. Before study onset, the protocol, informed consent, advertisements to be used for the recruitment of study subjects, and any other written information regarding this study to be provided to the subject must be approved by the HREC/IRB. Documentation of all HREC/IRB approvals and of the HREC/IRB compliance with the ICH E6(R2) guidelines will be maintained by the study site and will be available for review by the sponsor or its designee.

All HREC/IRB approvals should be signed by the HREC/IRB chairman or designee and must identify the HREC/IRB name and address, the clinical protocol by title or protocol number or both and the date approval or a favorable opinion was granted.

###### 16.2.2.3 Subject Consent

Written informed consent in compliance with US Title 21 CFR Part 50 and local regulatory authority requirements shall be obtained from each subject before he or she enters the study or before any unusual or nonroutine procedure that involves risk to the subject is performed. If any institution-specific modifications to study-related procedures are proposed or made by the study site, the consent should be reviewed by the sponsor or its designee or both before HREC/IRB submission. Once reviewed, the investigator will submit the ICF to the HREC/IRB for review and approval before the start of the study. If the ICF is revised during the course of the study, all active participating subjects must sign the revised form.

Before recruitment and enrollment, each prospective subject will be given a full explanation of the study and be allowed to read the approved ICF. Once the investigator is assured that the subject understands the implications of participating in the study, the subject will be asked to give his or her consent to participate in the study by signing the ICF.

The investigator or designee will provide a copy of the ICF to the subject. The original form shall be maintained in the subject’s medical records at the study site.

Where possible, completion of informed consent should occur at the start of a screening visit, and ICF documentation may be provided to potential subjects in advance of screening visits through email or regular mail. If certain categories of potential subjects (such as older adults with co-morbid conditions) have limited ability to attend sites for screening visits or vaccination visits due to local restrictions related to the coronavirus pandemic, then remote processes for completion of parts of the informed consent process may be implemented, or home visit approaches may occur to allow completion of informed consent, screening and vaccination.

###### 16.2.2.4 Study Reporting Requirements

By participating in this study, the investigator agrees to submit reports of SAEs according to the timeline and method outlined in this protocol. In addition, the investigator agrees to submit annual reports to his or her HREC/IRB as appropriate.

###### 16.2.2.5 Financial Disclosure and Obligations

The investigator is required to provide financial disclosure information to allow the sponsor to submit the complete and accurate certification or disclosure statements required under US Title 21 CFR Part 54 and local regulations. In addition, the investigator must provide to the sponsor a commitment to promptly update this information if any relevant changes occur during the course of the investigation and for 1 year following the completion of the study.

Neither the sponsor nor PPD nor the study site is financially responsible for further testing or treatment of any medical condition that may be detected during the screening process. In addition, in the absence of specific arrangements, neither the sponsor nor PPD nor the study site is financially responsible for further treatment of the disease under study.

###### 16.2.2.6 Investigator Documentation

Prior to beginning the study, the investigator will be asked to comply with ICH E6(R2) Section 8.2, US Title 21 of the CFR, and local regulations by providing essential documents, including but not limited to, the following:

- HREC/IRB approval;
- An original investigator-signed investigator agreement page of the protocol;
- Curriculum vitae for the principal investigator and each subinvestigator. Current licensure must be noted on the curriculum vitae. They will be signed and dated by the

principal investigators and subinvestigators at study start-up, indicating that they are accurate and current;

- Financial disclosure information to allow the sponsor to submit complete and accurate certification or disclosure statements required under US Title 21 CFR Part 54 and local regulations. In addition, the investigators must provide to the sponsor a commitment to promptly update this information if any relevant changes occur during the course of the investigation and for 1 year after the completion of the study;

- An HREC/IRB-approved ICF, samples of study site advertisements for recruitment for this study, and any other written information about this study that is to be provided to the subject; and

- Laboratory certifications and reference ranges for any local laboratories used by the study site, in accordance with US Title 42 CFR Part 493 and local regulations.

###### 16.2.2.7 Study Conduct

The investigator agrees that the study will be conducted according to the principles of ICH E6(R2). The investigator will conduct all aspects of this study in accordance with all national, state, and local laws or regulations. The study will be conducted in compliance with the protocol, current Good Clinical Practice guidelines – adopting the principles of the Declaration of Helsinki – and all applicable regulatory requirements.

Prior to study initiation, the protocol and the informed consent documents will be reviewed and approved by the sponsor and an appropriate ethics committee. Any amendment to the protocol or consent materials must also be approved by the study sponsor and HREC/IRB and must be submitted/notified to the regulatory authority, as required, before they are implemented.

###### 16.2.2.8 Case Report Forms and Source Documents

Site personnel will maintain source documentation, enter subject data into the eCRF as accurately as possible, and will rapidly respond to any reported discrepancies.

Electronic CRFs and electronic data capture will be utilized. The electronic data capture system is validated and compliant with US Title 21 CFR Part 11 and local regulations. Each person involved with the study will have an individual identification code and password that allows for record traceability. Thus, the system, and any subsequent investigative reviews, can identify coordinators, investigators, and individuals who have entered or modified records, as well as the time and date of any modifications. There may be an internal quality review audit of the data and additional reviews by the clinical monitor.

Each eCRF is presented as an electronic copy, allowing data entry by site personnel, who can add and edit data, add new subjects, identify and resolve discrepancies, and view records. This system provides immediate direct data transfer to the database, as well as immediate detection of discrepancies, enabling site coordinators to resolve and manage discrepancies in a timely manner.

Paper copies of the eCRFs and other database reports may be printed and signed by the investigator. This system provides site personnel, monitors, and reviewers with access to hardcopy audits, discrepancy reviews, and investigator comment information.

###### 16.2.2.9 Adherence to Protocol

The investigator agrees to conduct the study as outlined in this protocol, in accordance with ICH E6(R2) and all applicable guidelines and regulations.

###### 16.2.2.10 Reporting Adverse Events

By participating in this study, the investigator agrees to submit reports of SAEs according to the timeline and method outlined in this protocol. In addition, the investigator agrees to submit annual reports to his or her HREC/IRB as appropriate. The investigator also agrees to provide the sponsor with an adequate report, if applicable, shortly after completion of the investigator's participation in the study.

###### 16.2.2.11 Investigator's Final Report

Upon completion of the study, the investigator, where applicable, should inform the institution; the investigator/institution should provide the HREC/IRB with a summary of the study's outcome and the sponsor and regulatory authority(ies) with any reports required. Interim reports are expected to be provided to regulatory authorities to allow vaccine development advancement given the pandemic situation. These reports are planned to be aggregate and at the treatment level unless the SMC deems additional data at the individual level (eg, select listings of select subjects) will be beneficial. In such a case, a firewall will be in place to maintain the blind for those individuals involved in the study conduct to ensure unbiased assessment continue.

###### 16.2.2.12 Records Retention

Essential documents should be retained until at least 2 years after the last approval of a marketing application in an ICH region and until there are no pending or contemplated marketing applications in an ICH region or until at least 2 years have elapsed since the formal discontinuation of clinical development of the study vaccine or per local regulation, whichever is longer. These documents should be retained for a longer period, however, if required by applicable regulatory requirements or by an agreement with the sponsor. It is the sponsor's responsibility to inform the investigator/institution as to when these documents no longer need to be retained.

###### 16.2.2.13 Publications

After completion of the study, the data may be considered for reporting at a scientific meeting or for publication in a scientific journal. In these cases, the sponsor will be responsible for these activities and will work with the investigators to determine how the manuscript is written and edited, the number and order of authors, the publication to which it will be submitted, and any other related issues. The sponsor has final approval authority over all such issues.

Data are the property of the sponsor and cannot be published without their prior authorization, but data and any publication thereof will not be unduly withheld.

##### 16.2.3 Study Management

###### 16.2.3.1 Monitoring

###### 16.2.3.1.1 Monitoring of the Study

The clinical monitor, as a representative of the sponsor, is obligated to follow the study closely. In doing so, the monitor will visit the investigator and study site at periodic intervals in addition to maintaining necessary telephone and email contact. The monitor will maintain current personal knowledge of the study through observation, review of study records and source documentation, and discussion of the conduct of the study with the investigator and personnel. The monitor will be blinded to treatment assignment. A separate unblinded study monitor will be responsible for drug accountability.

All aspects of the study will be carefully monitored by the sponsor or its designee for compliance with applicable government regulation with respect to current ICH E6(R2) guidelines and standard operating procedures.

###### 16.2.3.1.2 Inspection of Records

The investigator and institution involved in the study will permit study-related monitoring, audits, HREC/IRB review, and regulatory inspections by providing direct access to all study records. In the event of an audit, the investigator agrees to allow the sponsor, their representatives, or the regulatory authority access to all study records.

The investigator should promptly notify the sponsor of any audits scheduled by any regulatory authorities and promptly forward copies of any audit reports received to the sponsor.

##### 16.2.3.2 Management of Protocol Amendments and Deviations

###### 16.2.3.2.1 Modification of the Protocol

This is a Phase 1/2 study to evaluate the safety and immunogenicity of a SARS-CoV-2 rS protein nanoparticle vaccine. This protocol is written with some flexibility to accommodate the inherent dynamic nature and dose finding of Phase 1 clinical studies as well as the evolving pandemic and urgency for efficacious vaccine availability. Modifications to the dose, dosing regimen, and/or clinical or laboratory procedures currently outlined below may be required to achieve the scientific goals of the study objectives and/or to ensure appropriate safety monitoring of the study subjects:

- Dose and/or dosing regimen in Part 2 will be selected and/or updated prior to the initiation of Part 2; however, the maximum dose may not exceed that evaluated in Part 1, and no subject shall receive more than 3 vaccinations in Part 2.

- The timing of procedures for assessment of safety procedures in Part 2 may be modified based on newly available safety and tolerability data or evolving COVID-19 data.

- Due to the evolving COVID-19 epidemic, should a large portion of the population appear to have previous exposure to SARS-CoV-2 rS, treatment groups with a single priming dose may be increased to represent proportionally more of the randomized population.

- Treatment groups may be omitted.

- Up to an additional 50 mL of blood may be drawn for safety or immunogenicity analyses. The total blood volume withdrawn from any single subject will not exceed the maximum allowable volume during his or her participation in the entire study.

- Additional database freezes may occur as the study evolves and should the ongoing epidemic progression warrant rapid decision-making on product manufacturing. The study will continue in a blinded fashion (at the subject level) until the EOS.

- Rapid diagnostic testing for SARS-CoV-2 by POC tests may be available and substituted for centralized testing if accepted by regulatory authorities as a secondary endpoint in this study and hold validity for vaccine advancement.

It is understood that the current study may employ some or none of the alterations described above. Any changes in this research activity, except those necessary to remove an apparent immediate hazard to the subject, must be reviewed and approved by the sponsor or designee. Amendments to the protocol must be approved by the IRB before subjects can be enrolled into an amended protocol.

###### 16.2.3.2.2 Protocol Deviations

The investigator or designee must document and explain in the subject's source documentation any deviation from the approved protocol. The investigator may implement a deviation from, or a change to, the protocol to eliminate an immediate hazard to study subjects without prior HREC/IRB approval. As soon as possible after such an occurrence, the implemented deviation or change, the reasons for it, and any proposed protocol amendments should be submitted to the HREC/IRB for review and approval, to the sponsor for agreement, and to the regulatory authorities, if required.

A protocol deviation is any change, divergence, or departure from the study design or procedures defined in the protocol. A major deviation is a subset of protocol deviations that leads to a subject being discontinued from the study or significantly affects the subject's rights, safety, or well-being and/or the completeness, accuracy, and reliability of the study data. A major deviation can include nonadherence to inclusion or exclusion criteria or nonadherence to regulatory authority including ICH E6(R2) guidelines. A minor protocol deviation are deviations that do not meet criteria to be classed as a major protocol deviation.

Protocol deviations will be documented by the clinical monitor throughout the course of monitoring visits. The investigator will be notified in writing by the monitor of deviations.

The HREC/IRB should be notified of all protocol deviations, if appropriate, in a timely manner.

Review and categorization of protocol deviations as either major or minor protocol deviations will occur prospectively during the study prior to database locks.

###### 16.2.3.3 Study Termination

Although the sponsor has every intention of completing the study, they reserve the right to discontinue it at any time for clinical or administrative reasons.

The end of the study is defined as the date on which the last subject completes the last study visit (including the EOS visit and any additional long-term follow-up). Any additional long-term follow-up that is required for monitoring of the resolution of an AE or finding may be appended to the clinical study report.

###### 16.2.3.4 Final Report

Regardless of whether the study is completed or prematurely terminated, the sponsor will ensure that clinical study reports are prepared and provided to regulatory agency(ies) as required by the applicable regulatory requirement(s). The sponsor will also ensure that clinical study reports in marketing applications meet the standards of the ICH Harmonised Tripartite Guideline E3: Structure and Content of Clinical Study Reports.

Where required by applicable regulatory requirements, an investigator signatory will be identified for the approval of the clinical study report. The investigator will be provided reasonable access to statistical tables, figures, and relevant reports and will have the opportunity to review complete study results.

Upon completion of the clinical study report, the sponsor will provide the investigator(s) with the final approved clinical study report.

#### 16.3 APPENDIX 3: FDA TOXICITY GRADING SCALES

**Table 16-1 FDA Toxicity Grading Scale for Clinical Abnormalities (Local and General Systemic Reactogenicity)**

| **Local Reaction to Injectable Product** | | | | |
|---|---|---|---|---|
| | **Mild (Grade 1)** | **Moderate (Grade 2)** | **Severe (Grade 3)** | **Potentially Life Threatening (Grade 4)** |
| Pain | Does not interfere with activity | Repeated use of nonnarcotic pain reliever >24 hours or interferes with activity | Any use of narcotic pain reliever or prevents daily activity | ER visit or hospitalization |
| Tenderness | Mild discomfort to touch | Discomfort with movement | Significant discomfort at rest | ER visit or hospitalization |
| Erythema/redness[a] | 2.5 – 5 cm | 5.1 – 10 cm | >10 cm | Necrosis or exfoliative dermatitis |
| Induration/swelling[b] | 2.5 – 5 cm and does not interfere with activity | 5.1 – 10 cm or interferes with activity | >10 cm or prevents daily activity | Necrosis |
| **Systemic (General)** | | | | |
| | **Mild (Grade 1)** | **Moderate (Grade 2)** | **Severe (Grade 3)** | **Potentially Life Threatening (Grade 4)** |
| Fever (°C)<br>(°F) | 38.0 – 38.4<br>100.4 – 101.1 | 38.5 – 38.9<br>101.2 – 102.0 | 39.0 – 40<br>102.1 – 104 | >40<br>>104 |
| Nausea/vomiting | No interference with activity or 1 – 2 episodes/24 hours | Some interference with activity or >2 episodes/24 hours | Prevents daily activity, or requires outpatient IV hydration | ER visit or hospitalization for hypotensive shock |
| Headache | No interference with activity | Repeated use of nonnarcotic pain reliever >24 hours or some interference with activity | Significant; any use of narcotic pain reliever or prevents daily activity | ER visit or hospitalization |
| Fatigue/Malaise | No interference with activity | Some interference with activity | Significant; prevents daily activity | ER visit or hospitalization |
| Myalgia | No interference with activity | Some interference with activity | Significant; prevents daily activity | ER visit or hospitalization |
| Arthralgia | No interference with activity | Some interference with activity | Significant; prevents daily activity | ER visit or hospitalization |

[a] In addition to grading the measured local reaction at the greatest single diameter, the measurement should be recorded as a continuous variable.

[b] Induration/swelling should be evaluated and graded using the functional scale as well as the actual measurement.

[c] Oral temperature; no recent hot or cold beverages.

Source: DHHS 2007.

**Table 16-2 Modified FDA Toxicity Grading Scale for Clinical Abnormalities (Local and General Systemic Reactogenicity)**

| **Local Reaction to Injectable Product** | | | | |
|---|---|---|---|---|
| | **Mild (Grade 1)** | **Moderate (Grade 2)** | **Severe (Grade 3)** | **Potentially Life Threatening (Grade 4)** |
| Pain | Does not interfere with activity | Repeated use of non-prescription pain reliever >24 hours or interferes with activity | Significant; any use of prescription pain reliever or prevents daily activity | Requires ER visit or hospitalization |
| Tenderness | Mild discomfort to touch | Discomfort with movement | Significant discomfort at rest | Requires ER visit or hospitalization |
| Erythema/redness[a] | 2.5 − 5 cm | 5.1 − 10 cm | >10 cm | Necrosis or exfoliative dermatitis[b] |
| Induration/swelling[a] | 2.5 − 5 cm | 5.1 − 10 cm | >10 cm | Necrosis[b] |
| **Systemic (General)** | | | | |
| | **Mild (Grade 1)** | **Moderate (Grade 2)** | **Severe (Grade 3)** | **Potentially Life Threatening (Grade 4)** |
| Fever[c] (°C)<br>(°F) | 38.0 − 38.4<br>100.4 − 101.1 | 38.5 − 38.9<br>101.2 − 102.0 | 39.0 − 40<br>102.1 − 104 | >40<br>>104 |
| Nausea/vomiting | Does not interfere with activity or 1 − 2 episodes/24 hours | Some interference with activity or >2 episodes/24 hours | Prevents daily activity, or requires IV hydration outside of hospital | Requires ER visit or hospitalization |
| Headache | Does not interfere with activity | Repeated use of non-prescription pain reliever >24 hours or interferes with activity | Significant; any use of prescription pain reliever or prevents daily activity | Requires ER visit or hospitalization |
| Fatigue/Malaise | Does not interfere with activity | Some interference with activity | Significant, prevents daily activity | Requires ER visit or hospitalization |
| Myalgia | Does not interfere with activity | Some interference with activity | Significant, prevents daily activity | Requires ER visit or hospitalization |
| Arthralgia | Does not interfere with activity | Some interference with activity | Significant, prevents daily activity | Requires ER visit or hospitalization |

a The measurements should be recorded as a continuous variable.

b These events are not subject reported through the electronic diary and will be monitored through the AE pages of the study database.

c Oral temperature if subject collected, sites may collect temperature using local clinic practices/devices. Toxicity grade will be derived.

Source: DHHS 2007.

**Table 16-3 FDA Toxicity Grading Scale for Laboratory Abnormalities**

| Serum Chemistry and Hematology | Mild (Grade 1) | Moderate (Grade 2) | Severe (Grade 3) | Potentially Life Threatening (Grade 4)[a] |
|---|---|---|---|---|
| Sodium – hyponatremia (mEq/L) | 132 – 134 | 130 – 131 | 125 – 129 | <125 |
| Sodium – hypernatremia (mEq/L) | 144 – 145 | 146 – 147 | 148 – 150 | >150 |
| Potassium – hyperkalemia (mEq/L) | 5.1 – 5.2 | 5.3 – 5.4 | 5.5 – 5.6 | >5.6 |
| Potassium – hypokalemia (mEq/L) | 3.5 – 3.6 | 3.3 – 3.4 | 3.1 – 3.2 | <3.1 |
| Glucose – hyperglycemia<br>Random (mg/dL)<br>*(mmol/L)*[c] | <br>116 to 160<br>*6.44 to <8.89* | <br>>160 to 250<br>*8.89 to <13.89* | <br>>250 to 500<br>*13.89 to <27.75* | <br>≥500<br>*≥27.75* |
| Urea (ie, BUN) (mg/dL) | 23 – 26 | 27 – 31 | >31 | Requires dialysis |
| Creatinine (mg/dL)[b] | 1.5 – 1.7 | 1.8 – 2.0 | 2.1 – 2.5 | >2.5 or requires dialysis |
| Total protein – hypoproteinemia (g/dL) | 5.5 – 6.0 | 5.0 – 5.4 | <5.0 | – |
| Liver function tests – ALT, AST; increase by factor[b] | 1.1 – 2.5 × ULN | 2.6-5.0 × ULN | 5.1 – 10 × ULN | >10 × ULN |
| Total bilirubin – when accompanied by any increase in liver function test; increase by factor[b] | 1.1 – 1.25 × ULN | 1.26 – 1.5 × ULN | 1.51 – 1.75 × ULN | >1.75 × ULN |
| Total bilirubin – when liver function test is normal; increase by factor[b] | 1.1 – 1.5 × ULN | 1.6 – 2.0 × ULN | 2.0 – 3.0 × ULN | >3.0 × ULN |
| Hemoglobin (Female) (g/dL)[b] | 11.0 – 12.0 | 9.5 – 10.9 | 8.0 – 9.4 | <8.0 |
| Hemoglobin (Female) change from baseline value (g/dL) | Any decrease – 1.5 | 1.6 – 2.0 | 2.1 – 5.0 | >5.0 |
| Hemoglobin (Male) (g/dL)[b] | 12.5 – 13.5 | 10.5 – 12.4 | 8.5 – 10.4 | <8.5 |
| Hemoglobin (Male) change from baseline value (g/dL) | Any decrease – 1.5 | 1.6 – 2.0 | 2.1 – 5.0 | >5.0 |
| WBC increase (cell/mm$^3$)[b] | 10,800 – 15,000 | 15,001 – 20,000 | 20,001 – 25,000 | >25,000 |
| WBC decrease (cell/mm$^3$)[b] | 2,500 – 3,500 | 1,500 – 2,499 | 1,000 – 1,499 | <1,000 |
| Platelets decreased (cell/mm$^3$)[b] | 125,000 – 140,000 | 100,000 – 124,000 | 25,000 – 99,000 | <25,000 |

Notes: The laboratory values provided in the table serve as guidelines and are dependent upon institutional normal parameters. Institutional normal reference ranges should be provided to demonstrate that they are appropriate. Adjustments will be made for formal toxicity grading based on the local parameter and overlap with the above FDA ranges. **Clinical laboratory abnormalities with FDA toxicity grade >1 at screening are exclusion criteria for Part 1 only (Section 6.1.2). See laboratory manual for final scoring ranges. For Part 2, clinical laboratory assessments will be recorded at screening to provide a reference point for evaluation of later AEs, as needed (Section 13.1.2).**

[a] The clinical signs or symptoms associated with laboratory abnormalities might result in characterization of the laboratory abnormalities as Potentially Life Threatening (grade 4). For example, a low sodium value that falls within a grade 3 parameter (125-129 mE/L) should be recorded as a grade 4 hyponatremia event if the subject had a new seizure associated with the low sodium value.

[b] Values to be included in vaccination pause rules (highlighted in light grey) for Part 1 (Section 6.1.5).

[c] DAIDS toxicity scoring scale.

Sources: DAIDS 2017; DHHS 2007.

**Table 16-4 FDA Toxicity Grading Scale for Clinical Abnormalities (Vital Signs)**

| | Mild (Grade 1) | Moderate (Grade 2) | Severe (Grade 3) | Potentially Life Threatening (Grade 4) |
|---|---|---|---|---|
| Tachycardia (bpm) | 101 – 115 | 116 – 130 | >130 | ER visit or hospitalization for arrhythmia |
| Bradycardia (bpm)[a] | 50 – 54 | 45 – 49 | <45 | ER visit or hospitalization for arrhythmia |
| Hypertension (systolic) (mm Hg) | 141 – 150 | 151 – 155 | >155 | ER visit or hospitalization for malignant hypertension |
| Hypertension (diastolic) (mm Hg) | 91 – 95 | 96 – 100 | >100 | ER visit or hospitalization for malignant hypertension |
| Hypotension (systolic) (mm Hg) | 85 – 89 | 80 – 84 | <80 | ER visit or hospitalization for hypotensive shock |
| Respiratory Rate (breaths per minute) | 17 – 20 | 21 – 25 | >25 | Intubation |

Note: Subject should be at rest for all vital sign measurements. For Part 2, vital signs will only be toxicity scored on day of vaccination.

[d] When resting heart rate is between 60 – 100 bpm. Use clinical judgement when characterizing bradycardia among some healthy subject populations (eg, conditioned athletes).

Source: DHHS 2007.

#### 16.4 APPENDIX 4: LISTINGS OF ADVERSE EVENTS OF SPECIAL INTEREST

Because it has been hypothesized that immunizations with or without adjuvant may be associated with autoimmunity, regulatory authorities have requested that Novavax instruct investigators to be especially vigilant regarding the PIMMC listed in Table 16-5. Note that this regulatory request is not specific to Novavax's SARS-CoV-rS or Matrix-M adjuvant; and there is no current evidence to suggest that the investigational products in this protocol are, or are not, associated with these illnesses. The list is not intended to be exhaustive, nor does it exclude the possibility that other diagnoses may be AESI.

**Table 16-5 Potential Immune-Mediated Medical Conditions (PIMMC)**

| Categories | Diagnoses (as MedDRA Preferred Terms) |
|---|---|
| Neuroinflammatory Disorders: | Acute disseminated encephalomyelitis (including site specific variants: eg, non-infectious encephalitis, encephalomyelitis, myelitis, myeloradiculomyelitis), cranial nerve disorders including paralyses/paresis (eg, Bell's palsy), generalized convulsion, Guillain-Barre syndrome (including Miller Fisher syndrome and other variants), immune-mediated peripheral neuropathies and plexopathies (including chronic inflammatory demyelinating polyneuropathy, multifocal motor neuropathy and polyneuropathies associated with monoclonal gammopathy), myasthenia gravis, multiple sclerosis, narcolepsy, optic neuritis, transverse myelitis, uveitis |
| Musculoskeletal and Connective Tissue Disorders: | Antisynthetase syndrome, dermatomyositis, juvenile chronic arthritis (including Still's disease), mixed connective tissue disorder, polymyalgia rheumatic, polymyositis, psoriatic arthropathy, relapsing polychondritis, rheumatoid arthritis, scleroderma (including diffuse systemic form and CREST syndrome), spondyloarthritis (including ankylosing spondylitis, reactive arthritis [Reiter's Syndrome] and undifferentiated spondyloarthritis), systemic lupus erythematosus, systemic sclerosis, Sjogren's syndrome |
| Vasculidities: | Large vessels vasculitis (including giant cell arteritis such as Takayasu's arteritis and temporal arteritis), medium sized and/or small vessels vasculitis (including polyarteritis nodosa, Kawasaki's disease, microscopic polyangiitis, Wegener's granulomatosis, Churg–Strauss syndrome [allergic granulomatous angiitis], Buerger's disease [thromboangiitis obliterans], necrotizing vasculitis and anti-neutrophil cytoplasmic antibody [ANCA] positive vasculitis [type unspecified], Henoch-Schonlein purpura, Behcet's syndrome, leukocytoclastic vasculitis) |
| Gastrointestinal Disorders: | Crohn's disease, celiac disease, ulcerative colitis, ulcerative proctitis |
| Hepatic Disorders: | Autoimmune hepatitis, autoimmune cholangitis, primary sclerosing cholangitis, primary biliary cirrhosis |

| Categories | Diagnoses (as MedDRA Preferred Terms) |
|---|---|
| Renal Disorders: | Autoimmune glomerulonephritis (including IgA nephropathy, glomerulonephritis rapidly progressive, membranous glomerulonephritis, membranoproliferative glomerulonephritis, and mesangioproliferative glomerulonephritis |
| Cardiac Disorders: | Autoimmune myocarditis/cardiomyopathy |
| Skin Disorders: | Alopecia areata, psoriasis, vitiligo, Raynaud's phenomenon, erythema nodosum, autoimmune bullous skin diseases (including pemphigus, pemphigoid and dermatitis herpetiformis), cutaneous lupus erythematosus, morphoea, lichen planus, Stevens-Johnson syndrome, Sweet's syndrome |
| Hematologic Disorders: | Autoimmune hemolytic anemia, autoimmune thrombocytopenia, antiphospholipid syndrome, thrombocytopenia |
| Metabolic Disorders: | Autoimmune thyroiditis, Grave's or Basedow's disease, Hashimoto thyroiditis[a], diabetes mellitus type 1, Addison's disease |
| Other Disorders: | Goodpasture syndrome, idiopathic pulmonary fibrosis, pernicious anemia, sarcoidosis |

[a] For Hashimoto thyroiditis: new onset only.

Adverse events of special interest relevant to COVID-19 are listed in Table 16-6. The list is not intended to be exhaustive, nor does it exclude the possibility that other diagnoses may be AESI. It is anticipated that additional AESI may be associated with COVID-19 disease. Investigators should stay updated regarding such public health notifications.

**Table 16-6 Adverse Events of Special Interest Relevant to COVID-19[a]**

| Body System | Diagnoses |
|---|---|
| Immunologic | Enhanced disease following immunization, cytokine release syndrome related to COVID-19 disease[b], multisystem inflammatory syndrome in children (MIS-C) |
| Respiratory | Acute respiratory distress syndrome (ARDS) |
| Cardiac | Acute cardiac injury including:<br>• Microangiopathy<br>• Heart failure and cardiogenic shock<br>• Stress cardiomyopathy<br>• Coronary artery disease<br>• Arrhythmia<br>• Myocarditis, pericarditis |
| Hematologic | Coagulation disorder<br>• Deep vein thrombosis<br>• Pulmonary embolus<br>• Cerebrovascular stroke<br>• Limb ischemia<br>• Hemorrhagic disease<br>• Thrombotic complications |

| Body System | Diagnoses |
|---|---|
| Renal | Acute kidney injury |
| Gastrointestinal | Liver injury |
| Neurologic | Guillain Barré Syndrome, anosmia, ageusia, meningoencephalitis |
| Dermatologic | Chilblain-like lesions, single organ cutaneous vasulitis, erythema multiforme |

[a] COVID-19 disease manifestations associated with more severe presentation and decompensation with consideration of enhanced disease potential. The current listing is based on Safety Platform for Emergency Vaccines (SPEAC) D2.3 Priority List of Adverse Events of Special Interest: COVID-19 (SPEAC 2020).

[b] Cytokine Release Syndrome related to COVID-19 disease is a disorder characterized by nausea, headache, tachycardia, hypotension, rash, and/or shortness of breath (DAIDS 2017).
