## Supplementary material for "Immunogenicity and safety of a 4^th^ homologous booster dose of a SARS-CoV-2 recombinant spike protein vaccine (NVX-CoV2373): a phase 2, randomized, placebo-controlled trial": Addendum

**Novavax, Inc.**

### 2019nCoV-101

### A 2-PART, PHASE 1/2, RANDOMIZED, OBSERVER-BLINDED STUDY TO EVALUATE THE SAFETY AND IMMUNOGENICITY OF A SARS-CoV-2 RECOMBINANT SPIKE PROTEIN NANOPARTICLE VACCINE (SARS-CoV-2 rS) WITH OR WITHOUT MATRIX-M™ ADJUVANT IN HEALTHY SUBJECTS

**06 OCT 2022**

Part 2 Statistical Analysis Plan

**Version 2.1, Addendum 4.0**

Prepared by:
PPD
3900 Paramount Parkway
Morrisville, NC 27560 USA

#### Approval Signatures

DocuSigned by:
Wei-Ti Huang
Signer Name: Wei-Ti Huang
Signing Reason: I approve this document
Signing Time: [illegible]2022 | 1:44:52 PM GMT
[illegible]F47C7A207CAEA10F2DD91

| Date | Prepared by: Wei-Ti Huang<br>Lead Biostatistician, Biostatistics PPD |
|---|---|

DocuSigned by:
Jay Pottala
Signer Name: Jay Pottala
Signing Reason: I approve this document
Signing Time: 11-Oct-2022 | 1:52:53 PM GMT
CC5EF4DE359C46C0A9F2B4123D2D40FE

| Date | Reviewed by: Jay Pottala<br>Senior Reviewer, Biostatistics PPD |
|---|---|

DocuSigned by:
Shirley Galbiati
Signer Name: Shirley Galbiati
Signing Reason: I approve this document
Signing Time: 11-Oct-2022 | 9:45:41 AM EDT
B290CF83776147A6B0D6A2867BC2FBAA

| Date | Approved by: Shirley Galbiati<br>Senior Manager, Biostatistics Novavax |
|---|---|

8.2. IMMUNOGENICITY ANALYSIS ... 32
*8.2.1. Serum IgG Antibody Levels and Serum ACE-2 Receptor Binding Inhibition Specific for SARS-CoV-2 rS Protein Antigen ... 33*
*8.2.2. Neutralization Assay Specific for SARS-CoV-2 Wildtype (or Variant) ... 36*
*8.2.3. Cell-Mediated Response for Type 1 T Helper (Th1) and Type 2 T Helper (Th2) 36*
*8.2.4. Anti-Nucleocapsid Protein ... 37*
8.3. EXPLORATORY ANALYSIS ... 37

**9. SAFETY ANALYSIS ... 39**

9.1. UNSOLICITED ADVERSE EVENTS ... 43
*9.1.1. Incidence of Adverse Events ... 44*
*9.1.2. Relationship of Adverse Events to Study Vaccine ... 46*
*9.1.3. Severity of Adverse Event ... 47*
*9.1.4. Serious Adverse Events ... 47*
*9.1.5. Medically-Attended Adverse Events ... 48*
*9.1.6. Adverse Events of Special Interest ... 48*
*9.1.7. Adverse Events Leading to Study Vaccine Discontinuation ... 48*
*9.1.8. Adverse Events Leading to Discontinuation from Study ... 48*
9.2. SOLICITED ADVERSE EVENTS ... 48
9.3. CLINICAL LABORATORY EVALUATIONS ... 50
9.4. VITAL SIGN MEASUREMENTS ... 50
9.5. PHYSICAL EXAMINATION ... 50

**10. INTERIM ANALYSES ... 51**

**11. CHANGES FROM ANALYSES PLANNED IN THE PROTOCOL ... 52**

**12. REFERENCES ... 53**

**13. APPENDICES ... 54**

APPENDIX 1 SCHEDULE OF EVENTS (PART 2) FOR SUBJECT IN ALL TREATMENT GROUPS 54

#### Summary of Changes

##### Statistical Analysis Plan Revision History

| Version | Date | Summary of Changes |
|---|---|---|
| Version 1.0 | 23SEP2020 | Original version |
| Version 2.0 | 02DEC2020 | 1) Section 4.3 Analysis Sets, added intention-to-treat analysis set<br>2) Sections 3.6.2 Secondary Endpoints & 8.2 Immunogenicity Analysis, additional pairwise treatment comparisons for secondary endpoints<br>3) Section 8.2.3 Cell Mediated Responses, added details for intracellular cytokine staining<br>4) Section 9.1.1 Incidence of AE, removed D21 summary, clarified AESI as relevant to COVID-19 (not COVID-19 positivity)<br>5) Section 10 Interim Analysis, sponsor to be unblinded at the subject-level at Day 35 instead of EOS. |
| Version 2.1 | 23FEB2021 | 1) Sections 2.2 Secondary Objectives, 3.6.2 Secondary Endpoints, 8.2 Immunogenicity Analysis and Appendix 1 Schedule of Events, added immunogenicity analysis at Day 357<br>2) Sections 3.1 Overall Study Design and Plan, 3.2 COVID-19 Disease Monitoring, 3.3 COVID-19 Disease Case Ascertainment Sections, 4.2 Randomization, Stratification and Blinding, and 4.3 Analysis Sets added:<br>a) content of Day 189 re-randomization of group split change (i.e., group B becomes B1 and B2, group C becomes C1 and C2)<br>b) discontinued FLU-PRO data collection in Australia due to extremely low incidence of SARS-CoV-2 transmission<br>c) allowed subjects to be unblinded to receive authorized COVID-19 vaccine<br>3) Section 7.1 Recent Vaccination and Concomitant Medication, changed to safety analysis set to summarize recent vaccination and concomitant medication usage<br>4) Section 8.1 Efficacy Analysis, omitted duration of resolved illness episodes from Day 105 analysis<br>5) Section 8.3 Exploratory Analysis, clarified analysis of Flu-Pro instrument<br>6) Section 9 Safety Analysis, added safety sensitivity analysis for subjects that were unblinded and/or received an approved COVID-19 vaccine<br>7) Section 11 Changes from Analyses Planned in the Protocol, updated as required due to above |
| Version 2.1, Addendum 1.0 | 10MAY2021 | 1) Sections 2.2 Secondary Objectives, 2.3 Exploratory Objectives, 3.6.2 Secondary Endpoints, 3.6.3 Exploratory Endpoints, 4.2 Randomization, Stratification and Blinding, 7.1 Recent Vaccinations and Concomitant Medications, 7.3 Study Vaccine Administration, 8.1 Efficacy Analysis, 8.2 Immunogenicity Analysis, 9. Safety Analysis, 9.3 Clinical Laboratory |

| | | |
|---|---|---|
| | | Evaluations, 9.4.Vital Sign Measurements, 9.5 Physical Examination, added:<br>a) clarified the downstream impact of Day 189 re-randomization on treatment group presentation.<br>b) described the treatment group comparison for GMFR and SCR will only be implemented before Day 189; but cross study visits comparison will still be presented.<br>2) Section 8.2. Immunogenicity Analysis- re-randomization treatment groups after D189 with ANCOVA Geometric LS Mean only; no group comparison will be performed.<br>3) Section 8.2.3 no longer present ICCS assay by % of CD4 cells (only counts per 1 million cells)<br>4) Section 9. Safety Analysis, added Vaccine-Exposure Adjusted Event Rate (VEAER) content and clarified the concept of time at risk.<br>5) Section 8.3. Exploratory Analysis, enhanced the content of COVID-19 disease episode and Flu-Pro instrument<br>6) Sections 10. Interim Analyses, 11. Changes from Analyses Planned in the Protocol, PPD Biostatistics group unblinded after approval of SAP v2.1, Addendum 1.0 (instead of at EOS). |
| Version 2.1, Addendum 1.1 | 16JUL2021 | 1) Section 2.3. Exploratory Objectives, 3.6.3. Exploratory Endpoints, and 8.2. Immunogenicity Analysis; added downstream impact for South Africa variant B.1.351.<br>2) Section 7.3. Study Vaccine Administration, corrected to planned treatment from actual treatment assignment.<br>3) Section 9. Safety Analysis, clarified in details for VEAER algorithm. |
| Version 2.1, Addendum 1.2 | 30SEP2021 | 1) Section 4.2. Randomization, Stratification and Blinding; explained downstream impact for approved/authorized vaccine and introduced coalescent header.<br>2) Section 9. Safety Analysis; introduced dynamic header to adjust for subject who exposed to study vaccine in safety analysis.<br>3) 10. Interim Analyses, 11. Changes from Analyses Planned in the Protocol; added Day 217 and 385-IA database lock for early submission purpose. Also, mentioned there will be no Day 371 database freeze.<br>4) Section 4.3. Analysis Sets, 11. Changes from Analyses Planned in the Protocol; clarified the definition for PP-Immunogenicity Analysis Set . |
| Version 2.1, Addendum 1.3 | 12OCT2021 | 1) Section 5.1 Disposition, 7.1. Recent Vaccinations and Concomitant Medications, added missing dates imputation rules. |
| Version 2.1, Addendum 1.4 | 02NOV2021 | 1) Section 4.3. Analysis Sets, 11. Changes from Analyses Planned in the Protocol; clarified the definition for PP-Efficacy Analysis Set regarding SARS-CoV-2 infection status. |
| Version 2.1, Addendum 2.0 | 23DEC2021 | 1) Sections 2.2 Secondary Objectives, 2.3 Exploratory Objectives, 3.1. Overall Study Design and Plan, 3.6.2 Secondary Endpoints, 3.6.3 Exploratory Endpoints, 4.2 Randomization, Stratification and Blinding, 5.1 |

| | | |
|---|---|---|
| | | Disposition, 7.3 Study Vaccine Administration, 8.2 Immunogenicity Analysis, 9. Safety Analysis<br>a) clarified the downstream impact of study 12-month booster for treatment group B and C to support PA v9.0.<br>b) described the treatment group and visit comparison for GMFR and SCR for 12-month booster.<br>c) added more content for VEAER to subject who consent to study 12-month booster.<br>2) Section 4. General Statistical Considerations: spelled out Visit 1, Visit 7 and Visit 10 as three different baselines.<br>3) Sections 4.3 Analysis Sets, 8.2.4 Anti-Nucleocapsid Protein, and 11 Changes from Analyses Planned in the Protocol: Modified PP-set by adding consideration of vaccination at Days, 0, 21, 189, and 357; also, adding Anti-N protein content. |
| Version 2.1 Addendum 3.0 | 10 JUN2022 | Section 4.3 Analysis Sets, 11 Changes from Analysis Planned in the Protocol: The Sensitivity analysis may be performed for immunogenicity analysis at Day 357 to Day 546 (EOS) by using the same definition of the Per-Protocol (PP) Analysis Set, except the unblinding restriction. |
| Version 2.1 Addendum 4.0 | 06 OCT2022 | Section 8 Efficacy Analysis and Immunogenicity Analysis: made language flexible for immunogenicity analysis at various timepoints for exploratory variants (e.g. Beta, Delta, Omicron). |

#### List of Abbreviations

| | |
|---|---|
| ACE-2 | angiotensin converting enzyme 2 |
| AE | adverse event |
| AESI | adverse event of special interest |
| CI | confidence interval |
| CMI | cell mediated immunity |
| eCRF | electronic case report form |
| CSR | clinical study report |
| ELISA | enzyme-linked immunosorbent assay |
| ELISpot | enzyme-linked immune absorbent spot |
| EOS | end of study |
| EUA | Emergency Use Authorization |
| ePRO | electronic patient reported outcomes |
| FAS | Full Analysis Set |
| FDA | Food and Drug Administration |
| FLU-PRO | InFLUenza Patient-Reported Outcome |
| GMEU | geometric mean ELISA units |
| GMFR | geometric mean fold rise |
| GMT | geometric mean titer |
| HCP | healthcare provider |
| HREC | human research ethics committee |
| ICCS | Intracellular Cytokine Staining |
| ICF | informed consent form |
| IgG | immunoglobulin G |
| IL | interleukin |
| IM | intramuscular |
| IP | Investigational product |
| INFg | interferon gamma |
| IRB | institutional review board |
| IRT | interactive response technology |
| ITT | Intent-to-Treat |
| LLOQ | lower limit of quantification |
| MedDRA | Medical Dictionary for Regulatory Activities |
| MAAE | medically-attended adverse event |
| PBMC | peripheral blood mononuclear cell |
| PCR | polymerase chain reaction |
| PIMMC | potential immune-mediated medical conditions |
| PP | Per-Protocol |
| PT | preferred term |
| SAE | serious adverse event |
| SAP | Statistical analysis plan |

| | |
|---|---|
| SARS-CoV-2 rS | SARS-CoV-2 Recombinant Spike Protein Nanoparticle Vaccine |
| SCR | seroconversion rate |
| SD | standard deviation |
| SMC | safety monitoring committee |
| SOC | system organ class |
| TEAE | treatment-emergent adverse event |
| Th1 | type 1 helper |
| Th2 | type 2 helper |
| TNFa | tumor necrosis factor alpha |
| ULOQ | upper limit of quantification |
| VEAER | Vaccine-Exposure Adjusted Event Rate |

### 1. Introduction

This document outlines the statistical methods to be implemented in the analysis of data collected within the scope of Novavax, Inc., protocol 2019nCoV-101 (A 2-Part, Phase 1/2, Randomized, Observer-Blinded Study to Evaluate the Safety and Immunogenicity of a SARS-CoV-2 Recombinant Spike Protein Nanoparticle Vaccine [SARS-CoV-2 rS] With or Without Matrix-M™ Adjuvant in Healthy Subjects), Version 9.0, protocol amendment 8, dated May 3rd, 2021.

Novavax, Inc. is developing a recombinant vaccine adjuvanted with the saponin-based Matrix-M™ (previously referred to as Matrix-M) for the prevention of disease caused by the SARS-CoV-2 virus.

The purpose of this 2-Part, Phase 1/2, first-in-human, randomized, placebo-controlled, observer-blinded study is to evaluate the safety and immunogenicity of a SARS-CoV-2 recombinant spike protein nanoparticle vaccine (SARS-CoV-2 rS) with or without Matrix-M adjuvant in healthy subjects. The primary objectives of Part 2 of this study are to identify the optimal dose across age strata based on immune response (IgG antibody to SARS-CoV-2 rS vaccine) and whether baseline immune status has an impact; and to accumulate a safety experience for the candidate vaccine in healthy adult subjects based on solicited short-term reactogenicity across a broad age spectrum (by toxicity grade) and by adverse event (AE) profile for primary vaccination (through Day 35). The information provided by the Phase 2 portion of this study will inform progression of the vaccine and identify dose(s) to take forward in an emergency use authorization setting and/or for Phase 3 efficacy or effectiveness trial(s).

The purpose of this statistical analysis plan (SAP) is to define the planned statistical analysis of the study data consistent with the study objectives in Part 2/Phase 2.

References throughout this SAP to specific visits by study day (e.g. Day 0) refer to study day as defined in schedule of events in Appendix 1.

### 2. Objectives

#### 2.1. Primary Objectives

The primary objectives of the Part 2/Phase 2 portion of this study are:

- To identify the optimal dose across age strata based on immune response (immunoglobulin G [IgG] antibody to SARS-CoV-2 rS vaccine) at Day 35 and whether baseline immune status has an impact.

- To accumulate a safety experience for the candidate vaccine in healthy adult subjects based on solicited short-term reactogenicity across a broad age spectrum (by toxicity grade) and by AE profile for primary vaccination (through Day 35).
- To identify dose(s) to potentially take forward in an emergency use authorization setting and/or for Phase 3 efficacy or effectiveness trial(s).

#### 2.2. Secondary Objectives

The secondary objectives of the Part 2/Phase 2 portion of this study are as follows:

- To assess if the single-dose regimens can provide similar (or adequate) priming immune response at Day 35 (compared to the two-dose regimens and to placebo) and whether baseline immune status alters such response (IgG antibody to SARS-CoV-2 rS protein).
- To define the optimal dosing regimen in subjects who are naïve and those with pre-existing antibodies to SARS-CoV-2 (if enough subjects are identified with preexisting antibodies) as assessed by the immune response (IgG antibody to SARS-CoV-2 rS protein and angiotensin-converting enzyme 2 [ACE2] receptor binding inhibition) to the various regimens at Day 21 (post first dose), Day 35 (post second dose), and Day 217. Optimal dosing regimen to be assessed across full age spectrum and by age strata (18-59, 60-84 years). Due to Day 189 re-randomization, immune response data collected on or after Day 189 will be presented as re-randomization group split (Table 1B).
- To describe the amplitude, kinetics, and durability of immune responses to the various regimens in terms of enzyme-linked immunosorbent assay (ELISA) units of serum IgG antibodies and titers of ACE2 receptor binding inhibition to SARS-CoV-2 rS protein(s) at selected time points and relative to whether subjects had pre-existing antibodies to SARS-CoV-2. To include reverse cumulative distribution curves.
- To describe the immune responses to the various regimens in terms of titers of neutralizing antibody at selected time points and relative to whether subjects had pre-existing antibodies to SARS-CoV-2 (subset of subjects). Optimal dosing regimen to be assessed across full age spectrum and by age strata (18-59, 60-84 years).
- To assess immune responses to the various regimens at 6 months and whether a boost at 6 months for the 5- µg dose regimen (Treatment Groups B and C) induces immune memory and is beneficial to maintain immune response in terms of IgG and neutralizing antibodies and ACE2 receptor binding inhibition to SARS-CoV-2 rS protein.
- To assess immune responses to the various regimens at 12 months (all treatment groups) and whether a boost at 12 months for subjects enrolled in the 5-µg dose regimens (Treatment Groups B and C) induces immune memory and is beneficial

to maintain immune responses in terms of IgG and neutralizing antibodies to SARS-CoV-2 rS and ACE2 receptor binding inhibition.

- To assess overall safety through 35 days after prime vaccination (1 or 2 doses) for all AEs; from boost (Day 189) through 28 days after 6-month boost (Day 217);; and, for subjects in Treatment Groups B and C, from 12-month boost (Day 357) through 28 days after 12-month boost (Day 385) (Table 1C) for all AEs; and through the end of study (EOS) for any medically-attended adverse event (MAAE) attributed to vaccine, adverse event of special interest (AESI), or serious adverse event (SAE). Due to Day 189 re-randomization, safety data collected on or after Day 189 will be presented as re-randomization group split (Table 1B).
- To assess for occurrence and duration of COVID-19 disease as measured by polymerase chain reaction (PCR) following subject-reported symptoms and to assess disease severity (virologically confirmed, mild, moderate, severe) and duration by patient-reported outcomes (eg, InFLUenza Patient-Reported Outcome [FLU PRO©]) in those immunized with SARS-CoV-2 rS compared to placebo.
- To assess cell-mediated response: Type 1 helper (Th1) or Type 2 helper (Th2) predominance by various vaccine regimens (e.g. interleukin [IL]-2, IL-4, IL-5, IL-6, IL-13, tumor necrosis factor alpha [TNFα], interferon gamma [INFγ] using flow cytometry, enzyme-linked immune absorbent spot [ELISpot], or other system) in whole blood and/or harvested peripheral blood mononuclear cell (PBMC) cells (in response to in vitro stimulation with SARS-CoV-2 rS protein) (subset of subjects).

#### 2.3. Exploratory Objectives

The exploratory objectives of the Part 2/Phase 2 portion of this study are as follows:

- To explore application of the FLU-PRO instrument to categorize COVID-19 severity, for maximal illness severity assessment, and the time course and severity of clinical symptomatology for COVID-19 cases for treatment groups.
- To utilize additional assays (current or to be developed) to best characterize the immune response for future vaccine development needs.
- To assess overall safety through 35 days after prime vaccination is initiated (1 or 2 doses) for solicited AEs; by SARS-CoV-2 positivity at Day 0, if greater than 10% of total subjects are SARS-CoV-2 positive at Day 0.
- To assess immune responses to the various regimens at, but not limited to, 12 months and whether a boost at 12 months for subjects enrolled in the 5-ug dose regimens (Treatment Groups B and C) induces immune memory and is beneficial to maintain immune response in terms of IgG and neutralizing antibodies to SARS-CoV-2 rS and ACE2 receptor binding inhibition for new variants, including, but not limited to, the South Africa variant B.1.351. Due to Day 189 re-randomization, exploratory data collected on or after Day 189 will be presented as re-randomization group split (Table 1B).

### 3. Investigational Plan

#### 3.1. Overall Study Design and Plan

If adequate safety and desired immune responses are observed at interim analyses during Part 1, the study may be immediately extended, at the sponsor's discretion, to include Part 2. Only 1 construct for SARS-CoV-2 rS will be evaluated in Part 2.

Part 2 is a Phase 2, randomized, placebo-controlled, observer-blinded study to evaluate the safety and immunogenicity of a SARS-CoV-2 rS with Matrix-M™ adjuvant in male and female subjects. Subjects will be healthy adults based on medical history and physical examination. It is anticipated that Matrix-M adjuvant will be required for an adequate response in this population and for dose sparing needs.

After signing the informed consent form (ICF), subjects may be screened within a window of up to 45 days; however, SARS-CoV-2 serostatus across the enrolling sites should not be expected to be more than 15% of the overall population with SARS-CoV-2 positivity at baseline. Subjects will be asked to provide consent for the use of samples for future testing for other viruses and/or sequencing of the SARS-CoV-2 in positive specimens or assay development specific to SARS-CoV-2 (or related variants).

Approximately 750 healthy male and female subjects between 18 and 84 years of age (inclusive) will be randomized in both Australia and the United States, with approximately 50% of subjects overall in the study ≥60 years of age. Two-factor, 2-level stratification will be employed (ages 18-59 and 60-84; study site). Subjects who meet the criteria for study entry will initially be randomized in a 1:1:1:1:1 ratio to 1 of 5 treatment groups (Table 1A).

**Table 1A Treatment Groups as Originally Planned (Part 2)**

| Treatment Group | Number of Subjects | Day 0 | Day 21 (−1 to +3 days) | Day 189 (±15 days) |
|---|---|---|---|---|
| | | **SARS-CoV-2 rS + Matrix-M Adjuvant** | **SARS-CoV-2 rS + Matrix-M Adjuvant** | **SARS-CoV-2 rS + Matrix-M Adjuvant** |
| A | 300 | Placebo | Placebo | Placebo |
| B | 300 | 5 µg + 50 µg | 5 µg + 50 µg | Placebo |
| C | 300 | 5 µg + 50 µg | Placebo | 5 µg + 50 µg |
| D | 300 | 25 µg + 50 µg | 25 µg + 50 µg | Placebo |
| E | 300 | 25 µg + 50 µg | Placebo | 5 µg + 50 µg |

COVID-19 disease monitoring for qualifying symptoms of suspected COVID-19 will commence every 14 days beginning at Day 28. Table 2 details qualifying symptoms of suspected COVID-19. If a subject has qualifying symptoms, this will trigger a request to subjects to self-sample according to protocol instructions (see below). COVID-19 symptoms collected as part of COVID-19 disease monitoring processes for clinical endpoint assessments will not be reported as Adverse Events unless they relate to illness events starting prior to Day 35 (see also Section 3.2.1 for a note about the removal of the requirement for the use of ePRO for self-monitoring of qualifying symptoms of suspected COVID-19, applicable only to subjects enrolled at Australian sites).

Subjects at US sites will continue to be monitored via electronic patient reported outcomes (ePRO) system (data capture) for symptoms associated with SARS-CoV-2 infection (e.g. cough, fever, sore throat, difficulty breathing, and other symptoms [Table 2]) every 14 days beginning at Day 28 through Day 217. When COVID-19 disease symptom scoring indicates the need for sample collection for potential PCR confirmation at sites in the US, subjects will self-collect a nasal mid turbinate sample taken as close to the onset of symptoms as possible (ideal timing within 3 days). Symptom-based collection can be either due to passive or active surveillance using an ePRO system. Samples will not be collected after 14 days as a new symptom-based query will be initiated (e.g. every 2 weeks).

**Table 3 Endpoint Definitions of COVID-19 Disease Severity**

| COVID-19 Severity | Endpoint Definitions |
|---|---|
| **Virologically Confirmed** | ≥ 1 COVID-19 disease symptom in Table 2<br>**AND**<br>Does not meet criteria for mild, moderate or severe disease |
| **Mild** | ≥ 1 of:<br>• Fever (defined by subjective or objective measure, regardless of use of anti-pyretic medications)<br>• New onset cough<br>• ≥ 2 COVID-19 respiratory/non-respiratory symptoms in Table 2<br>**AND**<br>• Does not meet criteria for moderate or severe |
| **Moderate** | ≥ 1 of:<br>• Fever (defined by subjective or objective measure, regardless of use of anti-pyretic medications) + any 2 COVID-19 disease symptoms in Table 2 for ≥ 3 days (need not be contiguous days)<br>• High fever (≥ 38.4°C) for ≥ 3 days (need not be contiguous days)<br>• Any evidence of significant LRTI:<br>– Shortness of breath (or breathlessness or difficulty breathing) with or without exertion (greater than baseline)<br>– Tachypnea: 20 to 29 breaths per minute at rest<br>– $SpO_2$: 94% to 95% on room air<br>– Abnormal chest x-ray or chest CT consistent with pneumonia or LRTI<br>– Adventitious sounds on lung auscultation (eg, crackles/rales, wheeze, rhonchi, pleural rub, stridor)<br>**AND**<br>– Does not meet criteria for severe disease |
| **Severe** | ≥ 1 of:<br>• Tachypnea: ≥ 30 breaths per minute at rest<br>• Resting heart rate ≥ 125 beats per minute<br>• SpO2: ≤ 93% on room air or PaO2/FiO2 < 300 mmHg<br>• High flow oxygen therapy or NIV/NIPPV (eg, CPAP or BiPAP)<br>• Mechanical ventilation or ECMO |

| | |
|---|---|
| | • One or more major organ system dysfunction or failure (eg, cardiac/circulatory, pulmonary, renal, hepatic, and/or neurological, to be defined by diagnostic testing/clinical syndrome/interventions), including any of the following<br>○ Acute respiratory distress syndrome (ARDS)<br>○ Acute renal failure<br>○ Acute hepatic failure<br>○ Acute right or left heart failure<br>○ Septic or cardiogenic shock (with shock defined as SBP < 90 mm Hg or DBP < 60 mm Hg<br>○ Acute stroke (ischemic or hemorrhagic)<br>○ Acute thrombotic event: AMI, DVT, PE<br>○ Requirement for: vasopressors, systemic corticosteroids, or hemodialysis.<br>○ Admission to an ICU<br>○ Death |

Abbreviations: BiPAP = bi-level positive airway pressure; CPAP = continuous positive air pressure; CT = computed tomography; DBP = diastolic blood pressure; ECMO = extracorporeal membrane oxygenation; $FiO_2$ = fraction of inspired oxygen; HCP = healthcare provider; ICU = intensive care unit; LRTI = lower respiratory tract infection; NIV = non-invasive ventilation; $PAO_2$ = partial pressure of oxygen in the alveolus; SBP = systolic blood pressure; $SpO_2$ = oxygen saturation.

#### 3.5. COVID-19 Clinical Endpoint Case Definitions

A subject whose nasal mid-turbinate sample tests positive for SARS-CoV-2 infection by qualitative PCR [ie, (+)-PCR-confirmed SARS-CoV-2 illness) and whose initial qualifying symptom(s) of suspected COVID-19 disease (Table 2) meets the criteria for symptomatic virologically confirmed, mild, moderate, or severe COVID-19 disease (Table 3) within 14 days of the initial qualifying symptom(s) will be defined as a single case in the secondary efficacy analysis.

Due to the ongoing pandemic, recent national regulatory and local human research ethics committee (HREC) / institutional review board (IRB) and public health guidance will be applied at the site locations regarding alternations in the ability of study subjects to attend an investigational site for protocol specified visits, with the site's investigator being allowed to conduct safety assessments (eg, telephone contact, virtual visit via telemedicine, alternative location for assessment, including local laboratories or imaging centers) when necessary and feasible, as long as such visits are sufficient to assure the safety of study subjects. Serum samples may be drawn using local phlebotomy services, home health, or other modalities if site visits cannot occur. Vaccination visits must have adequate oversight for issues associated with immediate severe reactions but may need to occur outside of the clinical site depending on the pandemic situation (eg, home vaccinations).

#### 3.6. Study Endpoints

##### 3.6.1. Primary Endpoints

The primary endpoints of the Part 2/Phase 2 portion of this study are:

- Serum IgG antibody levels specific for the SARS-CoV-2 rS protein antigen(s) as detected by ELISA using geometric mean titer (GMT) or SCR for the two-dose regimens by dose at Day 35 regardless of baseline immune status. Derived/calculated endpoints based on these data will include geometric mean ELISA units (GMEUs), geometric mean fold rise (GMFR), and SCR.
  - Two-dose regimens by dose compared to placebo:
    B vs A; D vs A; (B+D) vs A (Table 1A)

  **SCR** is defined as the percentage of subjects with a post-vaccination titer ≥4 fold.
  **Positive baseline status** (+/-) using GMT and/or positive PCR at baseline.

- Numbers and percentages (with 95% CIs) of subjects with solicited AEs (local, systemic) for 7 days following each primary vaccination (Days 0 and 21) by toxicity grade (daily and maximum across 7 days) and duration. Unsolicited AEs (eg, treatment-emergent, serious, suspected unexpected serious, those of special interest, MAAEs) through 35 days by Medical Dictionary for Regulatory Activities (MedDRA) classification, severity score, and relatedness.

##### 3.6.2. Secondary Endpoints

The secondary endpoints of the Part 2/Phase 2 portion of this study are:

- Serum IgG antibody levels specific for the SARS-CoV-2 rS protein antigen(s) as detected by ELISA using GMT or SCR for the single-dose regimens compared to the two-dose regimens and to placebo at Day 21 and Day 35 regardless of baseline immune status. Derived/calculated endpoints based on these data will include GMEUs, GMFR, and SCR.
  - Treatment comparisons will consist of the following including single-dose regimens compared to the two-dose regimens and to placebo:
    B vs A;
    C vs B; C vs D; C vs (B+D) (at Day 35); C vs A;
    D vs A;
    E vs B; E vs D; E vs (B+D) (at Day 35); E vs A;
    (B+D) vs A (at Day 35); (C+E) vs A (at Day 35);
    (B+C) vs A (at Day 21 only); (B+C) vs (D+E) (at Day 21 only);
    (B+D) vs (C+E) (at Day 35);
    (D+E) vs A (at Day 21 only) (Table 1A).

- Serum IgG antibody levels specific for the SARS-CoV-2 rS protein antigen as detected by ELISA, described across study time points with derived/calculated endpoints to include GMEUs, GMFR, and SCR (≥ 4-fold change) for the single-dose regimens compared to the two-dose regimens and to placebo, stratified by baseline immune response.
    - Treatment comparisons will consist of the following including single-dose regimens compared to the two-dose regimens and to placebo:
      B vs A;
      C vs B; C vs D; C vs (B+D) (at Day 35 and following); C vs A;
      D vs A;
      E vs B; E vs D; E vs (B+D) (at Day 35 and following); E vs A;
      (B+D) vs A (at Day 35 and following); (C+E) vs A (at Day 35 and following);
      (B+C) vs A (at Day 21 only); (B+C) vs (D+E) (at Day 21 only);
      (B+D) vs (C+E) (at Day 35 and following);
      (D+E) vs A (at Day 21 only) (Table 1A). However, treatment comparisons for GMFR and SCR will not be performed on or after Day 189.
    - Serum IgG antibody levels specific for the SARS CoV 2 rS protein antigen(s) as detected by ELISA using GMT will be summarized based on the Day 189 re-randomization treatment group (i.e., A, B1, B2, C1, C2, D, and E) (Table 1B) from Day 189 (included as the second reference timepoint) to Day 546 (Day 357 if subjects disagree to 12-month booster dose) regardless of baseline immune status. GMFR and SCR (≥ 4-fold change) will only be presented across study visit comparison.
    - Serum IgG antibody levels specific for the SARS CoV 2 rS protein antigen(s) as detected by ELISA using GMT will be summarized for subjects in Treatment Groups B and C who agree to 12-month booster dose (Table 1C) from Day 357 (included as the third reference timepoint) to Day 546 regardless of baseline immune status. GMFR and SCR (≥ 4-fold change) will only be presented across study visit comparison.
- Epitope-specific immune responses to the SARS-CoV-2 rS protein receptor binding domain measured by serum titers in an ACE2 receptor binding inhibition assay to include GMT or concentration, GMFR, and SCR (≥ 4-fold change) for the single-dose regimens compared to the two-dose regimens and to placebo.
    - Treatment comparisons will consist of the following including single-dose regimens by dose compared to two-dose regimens and to placebo:
      B vs A;
      C vs B; C vs D; C vs (B+D) (at Day 35 and following); C vs A;
      D vs A;
      E vs B; E vs D; E vs (B+D) (at Day 35 and following); E vs A;
      (B+D) vs A (at Day 35 and following); (C+E) vs A (at Day 35 and following);
      (B+C) vs A (at Day 21 only); (B+C) vs (D+E) (at Day 21 only);

(B+C) vs (D+E) (at Day 21 only);

(B+D) vs (C+E) (at Day 35 and following);

(D+E) vs A (at Day 21 only) (Table 1A). However, treatment comparisons for GMFR and SCR (≥ 4-fold change) will not be performed on or after Day 189.

  - Epitope-specific immune responses to the SARS-CoV-2 rS protein receptor binding domain measured by serum titers in an ACE2 receptor binding inhibition assay to include GMT will be summarized based on the Day 189 re-randomization treatment group (i.e., A, B1, B2, C1, C2, D, and E) (Table 1B) from Day 189 (as the second reference time point) to Day 546 regardless of baseline immune status. GMFR and SCR (≥ 4-fold change) will only be presented across study visit comparison.

    For treatment Groups B and C with 12-month booster dose (Table 1C), epitope-specific immune responses to the SARS-CoV-2 rS protein receptor binding domain measured by serum titers in an ACE2 receptor binding inhibition assay to include GMT will be summarized based on the Day 189 re-randomization treatment group (i.e., B1, B2, C1, C2). GMFR and SCR (≥ 4-fold change) will only be presented across study visit comparison based on Day 357 (as the third reference time point) to Day 371 and Day 546 and regardless of baseline immune status.

- Neutralizing antibody activity at Days 35, 189, 217, and 357 for all treatment groups, and additionally at Days 371 and 546 for Treatment groups B and C relative to multiple baselines (i.e. Days 0, 189 and 357) in a subset of subjects by GMT and change from baseline, including GMFR and the SCR (≥ 4-fold change). Analysis to include subjects by treatment group, by age (18-59, 60-84 years) and relative to whether subjects had pre-existing antibodies to SARS-CoV-2. A sampling scheme to identify a subset of such subjects will be deployed.
  - Neutralizing antibody activity will be summarized based on the original randomization treatment group (Table 1A) from Day 0 to Day 35, and based on Day 189 re-randomization treatment group (i.e., A, B1, B2, C1, C2, D, and E) (Table 1B) from Day 189 (as the second reference time point) to Day 546 regardless of baseline immune status. GMFR and SCR (≥ 4-fold change) will be presented across study visit comparison for all timepoints, but treatment group comparison will only be included at Day 35.
  - For treatment groups B and C with 12-month booster dose (Table 1C), neutralizing antibody activity will be summarized based on the Day 189 re-randomization treatment group (i.e., B1, B2, C1, C2). GMFR and SCR (≥ 4-fold change) will only be presented across study visit comparison based on Day 357 (as the third reference time point) to Day 371 and Day 546 and regardless of baseline immune status.

- Serum IgG antibody levels specific for the SARS-CoV-2 rS protein antigen(s) as detected by ELISA using GMT or GMFR at Days 189, 217, and 357 for all treatment groups and additionally at Days 371 and 546 for Treatment Groups B and C for boosting assessment with either placebo or active boost. GMFR will only be presented across study visit comparison at Days 189 (as the second reference time point), 217, 357, 371 and 546 for boosting assessment. For treatment groups B and C with 12-month booster dose, GMFR will only be presented across study visit comparison based on Day 357 (as the third reference time point) to Day 371 and Day 546 and regardless of baseline immune status.

Due to Day 189 re-randomization, exploratory data collected on or after Day 189 will be presented as re-randomization split (Table 1B).

### 4. General Statistical Considerations

Data from subjects excluded from an analysis set will be included in the applicable data listings but not included in the calculation of summary statistics for that analysis set.

For categorical variables, frequencies, percentages, and 95% CIs (where applicable) will be presented. All percentages will be rounded to one decimal place. The number and percentage will be presented in the form *XX* (*XX.X*), where the percentage is displayed in parentheses.

Continuous variables will be summarized using descriptive statistics (i.e. number of subjects, mean, median, SD, minimum, maximum, and 95% CIs [where applicable]). All mean and median values will be formatted to one more decimal place than the actual value. Standard deviation values will be formatted to two more decimal places than the actual value. Minimum and maximum will be formatted to the same decimal place as the actual value.

When count data are presented, the percentage will be suppressed when the count is zero in order to draw attention to the non-zero counts. The denominator for all percentages will be the number of subjects in the given treatment group within the analysis set of interest and stratification level (if applicable) unless otherwise specified in a footnote.

All CIs will be 2-sided and performed using a 5% significance level. No multiplicity adjustment will be made for this early phase study where multiple treatment groups and endpoints are being evaluated.

Unless specified otherwise, baseline will be defined as the last non-missing assessment prior to the first study vaccine administration. Both scheduled and unscheduled visits and assessments will be used in determining baseline. The change from baseline is calculated as the post-baseline value minus the baseline value.

Unscheduled visits will not be summarized separately in by-visit summaries of data but may contribute to either initial baseline, 6-month booster baseline, 12-month booster baseline, or post-baseline Worst-Case (i.e. maximum value) values where applicable. All data collected at unscheduled visits will be included in the applicable data listings. Similarly, Early Termination visits will not be summarized separately in by-visit summaries of data but may contribute to post-baseline Worst-Case (i.e. maximum value) value where applicable.

All analyses adjusting for age stratification (18-59, 60-84 years) will use the stratification values as randomized in the interactive response technology (IRT).

Study day (also referred to as Analysis Day) will be calculated per CDISC ADaM standards relative to the first vaccination date as:

- If assessment/collection date is on or after the first dose of study vaccine administration, then Study Day = Assessment/Collection Date – First Vaccination Date + 1
- Otherwise, Study Day = Assessment/Collection Date – First Vaccination Date

Using the above formulae, baseline will correspond with Day 1 rather than Day 0 and the target study day for all post-baseline visits will be 1 day later than described in the schedule of events in Appendix 1.

For GMT, GMFR, and SCR calculations, immunogenicity values reported as below the lower limit of quantification (LLOQ) will be replaced by 0.5 × LLOQ as applicable. Values that are greater than the upper limit of quantification (ULOQ) will be replaced by the ULOQ as applicable. Missing results will not be imputed.

For immunogenicity data, if multiple non-missing values are available at the same visit then the later value will be used in the analysis. In cases where multiple non-missing values are on the same date and time, the value with the larger Study Data Tabulation Model (SDTM) record sequence number will be used in the analysis.

For subgroup analyses by baseline serostatus, any subjects with indeterminate PCR results will be grouped with subjects with negative results.

All statistical analyses detailed in this SAP will be conducted using SAS® 9.4 or higher.

#### 4.1. Sample Size

The decision on the choice of formulation and dosing regimen will be made based on the totality of the immunogenicity and safety data rather than any individual measurement. No multiplicity adjustment will be made for this early phase study where multiple treatment groups and endpoints are being evaluated.

With 150 subjects in each treatment group, there is a greater than 99.9% probability to observe at least 1 subject with an AE if the true incidence of the AE is 5% and 77.9% probability if the true incidence of the AE is 1%.

Table 4 presents the width of the 95% CI (based on the Clopper-Pearson method) for 150 subjects in each treatment group under seroconversion rate assumptions between 50% and 90%.

**Table 4 Width of the 95% Confidence Interval (based on the Clopper-Pearson method)**

| Seroconversion Rate (%) | 95% CI (%) | Width (%) of 95% CI |
|---|---|---|
| 50 | 41.7, 58.3 | 16.5 |
| 60 | 51.7, 67.9 | 16.2 |
| 70 | 62.0, 77.2 | 15.2 |
| 80 | 72.7, 86.1 | 13.4 |
| 90 | 84.0, 94.3 | 10.3 |

Notes: Width of 2-sided 95% confidence interval (based on the Clopper-Pearson method) for the specified seroconversion rate produced by a sample size of 150 subjects is presented. PASS 15.0.7 Confidence Intervals for One Proportion is used in the calculation.

Table 5 presents the power of detecting difference in SCRs between treatment groups for 150 subjects in each treatment group.

**Table 5 Power of Detecting Difference in Seroconversion Rates between Treatment Groups**

| Seroconversion Rate (%) in the Control Group | Seroconversion Rate (%) in the Comparison Group | Power of Detecting the Difference in Seroconversion Rate |
|---|---|---|
| 50 | 55 | 14.6% |
| | 60 | 42.9% |
| | 65 | 76.1% |
| | 70 | 94.7% |
| 60 | 65 | 14.3% |
| | 70 | 44.0% |
| | 75 | 79.6% |
| | 80 | 97.0% |
| 70 | 75 | 16.1% |
| | 80 | 51.9% |
| | 85 | 88.3% |
| | 90 | 99.4% |
| 80 | 85 | 20.6% |
| | 90 | 68.6% |
| | 95 | 98.5% |
| 90 | 95 | 36.9% |

Notes: Power of detecting the specified difference in SCRs between treatment groups, assuming a significance level of 0.05, produced by a sample size of 150 subjects in each treatment group is presented. PASS 15.0.7 Superiority by a Margin Tests for the Difference Between Two Proportions (Miettinen & Nurminen Likelihood Score Test) is used in the calculation.

Table 6 presents the power of detecting ratio of geometric means between treatment groups for 150 subjects in each treatment group.

**Table 6 Power of Detecting Ratio of Geometric Means between Treatment Groups**

D189 Re-randomization - Subjects initially randomized to treatment groups B and C through Day 21 will be re-randomized at a 1:1 ratio to receive either a booster dose of vaccine (B2 and C2) or placebo (B1 or C1) at Day 189 (Table 1B). To maintain the blind, every subject that remained in the study on or after Day 189 will be re-randomized, although groups A, D, and E will be assigned their same group.

D357 Additional Dose - Subjects in Treatment Groups B and C who agree to continue study participation for an additional 6 months will receive a booster dose of vaccine (B1, B2, and C1) or placebo (C2) at Day 357 (Table 1C).

Due to the nature of study design, it would be hard to maintain the blind post Day 357. Study subjects who agree to 12-month booster (i.e., after Day 357) can be inferred to be in treatment group B or C. Also, there's increased possibility that one could use subject-level to unblind themselves via a group-level summary using unique data point combinations. However, these are all known and acceptable risks. For more details regarding how to maintain the blind, please refer to Novavax Phase 2 Blinding Plan.

For subjects who do not wish to be re-randomized at Day 189 due to receiving approved EUA or early discontinuation or any other safety concerns, those in Treatment A, D, or E before Day 189 will be analyzed according to their original treatments and those in Treatment B or C before Day 189 will be analyzed according to B1 and C1, respectively. The coalescent header will be designed for this accommodation on unsolicited AEs, PCR-confirmed SARS-CoV-2 positivity by COVID-19, vaccine efficacy, and baseline tables where the table is not being analyzed in the fashion of pre-/post-Day 189. Solicited AEs and immunogenicity tables will be split between pre-/post-Day 189 with denominator of subjects who receive corresponding vaccine in each study period.

This is an observer-blinded study. To maintain the blind, placebo vaccination via intramuscular (IM) route will be included, and unblinded site personnel will manage vaccine logistic, preparation, and administration (if necessary) so as to maintain the blind from the remainder of the site personnel and subjects. The unblinded site personnel will not be involved in study-related assessments or have subject contact for data collection following study vaccine administration.

A subject's treatment assignment will not be broken until the end of the study for the clinical site study team unless medical treatment of the subject depends on knowing the study treatment the subject received. In the event that the blind needs to be broken because of a medical emergency or planned receipt of an approved/authorized vaccine, the investigator may unblind an individual subject's treatment allocation. A subject's treatment assignment may also be broken in the event that the Novavax study vaccine, or another vaccine from a different manufacturer, is demonstrated to be safe and efficacious and approved/authorized by the regulatory authority in Australia and the subject plans on receiving the approved/authorized vaccine. Personnel at the clinical study site including, investigators and study staff, research site, and study subjects will be unblinded at EOS after all assessments are completed. For subject in treatment group A, D, and E and for subjects in Treatment Groups B or C who disagree to the 12-month booster dose, EOS is Day 357. For subjects in Treatment Groups B or C who agree to the 12-month booster dose, the EOS is Day 546.

For a medical emergency, as soon as possible, the investigator should first contact the medical monitor to discuss the medical emergency and the reason for revealing the actual treatment received by that subject; in either case, the treatment assignment will be unblinded through IRT. Reasons for treatment unblinding must be clearly explained and justified in the electronic case report form (eCRF). The date on which the code was broken together with the identity of the person responsible must also be documented.

For unblinding associated with receipt of an approved/authorized vaccine, site personnel who become unblinded may continue to be involved in follow-up safety monitoring. It is expected that a significant number of study subjects will have the opportunity to receive an approved/authorized vaccine to prevent COVID-19 following the primary analysis at Day 35. It will be difficult for sites to maintain the blind for these subjects since the approved/authorized vaccine will be documented as a concomitant medication in the subject's medical records and in the study database which are readily available to blinded staff.

In addition to the aforementioned situations where the blind may be broken, the data will also be unblinded to a statistical team at specified time points for interim analyses, as outlined in Section 10.

##### 4.3. Analysis Sets

The All Screened Subjects Analysis Set will include all subjects who sign the ICF.

The Intent-to-Treat Analysis Set will include all subjects who are randomized regardless of protocol violations or missing data.

The Full Analysis Set (FAS) will include all ITT subjects who receive at least 1 dose of study vaccine (SARS-CoV-2 or placebo). The Full Analysis Set will be used for supportive analyses. Subjects will be analyzed according to the randomized treatment assignment.

The Safety Analysis Set will include all subjects who receive at least 1 dose of study vaccine (SARS-CoV-2 rS or placebo). In cases of treatment misallocation where the vaccine combination/sequence received differs from the randomized treatment arm but still corresponds with one of the 5 planned treatment arms, subjects will be analyzed according to the actual vaccine combination/sequence received; in cases of treatment misallocation where the vaccination combination/sequence received differs from the randomized treatment arm and does not correspond with any of the 5 planned treatment arms, subject will be analyzed by actual treatment received at first dose prior to Day 189 (Table 1A) and by actual treatment received following re-randomization (Table 1B) on or after Day 189.Actual treatment received will be assumed to correspond with randomized treatment assignment except in cases where a site reports having used incorrect investigational product (IP) during vaccine administration.

The Per-Protocol (PP) Analysis Set will be determined for each study visit and will include all subjects who are randomized, all subjects who receive the initial dose of study vaccine (SARS-CoV-2 rS or placebo) for all analyses through Day 21, all subjects who receive vaccine doses at both Day 0 and Day 21 for all analyses starting at Day 28, all subjects who receive vaccine doses at Days 0, 21, and 189 for all analyses starting at Day 217, all subjects who receive vaccine doses at Days, 0, 21, 189, and 357 for all analyses starting at Day 371, all subjects who have at least a baseline and 1 post-baseline serum sample IgG result available at the corresponding study visit, all subjects who have no major protocol violations (e.g. having randomized treatment assignment unblinded due to medical emergency, having taken prohibited medication on-study such as insulin, having received a vaccination containing IP that differs from randomized treatment assignment) that impact immunogenicity response at the corresponding study visit, and all subjects who do not have a positive Anti-Nucleocapsid Protein result on or after Day 371. Subjects who receive an approved/authorized COVID-19 vaccine but are unblinded or not unblinded by their site will be excluded on the date of vaccination.

Two analyses will be conducted with subjects who are unblinded to receive an approved/authorized COVID-19 vaccine. First, all subjects will be excluded from the Per Protocol Set the day following their date of unblinding. Second, subjects who receive vaccine doses at days 0, 21, 189 and 357 and meet all other PP Analysis Set criteria but are unblinded to receive an approved/authorized vaccine will be included in a separate Sensitivity Analysis. The Sensitivity analysis may be performed for immunogenicity analysis at Day 357 to Day 546 (EOS) by using the same definition of the Per-Protocol (PP) Analysis Set, except the unblinding restriction. Subjects who are unblinded to receive an approved/authorized COVID-19 vaccine will be included in the Sensitivity Analysis if there's no evidence to suggest they received the approved/authorized COVID-19 vaccine. As a general rule, data collected on the same date as the unblinding event will be included in analysis, e.g. if a blood draw occurred on the same date as unblinding for vaccine use. Subjects who are unblinded unexpectedly will be reviewed via protocol deviation review meeting with the Study Deviations Rules document to determine their analysis set membership.

The review and determination for exclusion from the Per Protocol Set will be carried out in a blinded fashion by a study clinician prior to unblinding for each analysis based on all available information from either the locked database or from a database freeze, depending on the SAP defined analysis. All subjects in the Per-Protocol Analysis Set will be analyzed according to the randomized treatment assignment. Any subject who is SARS-CoV-2 positive by qualitative PCR testing from screening and prior to the immunogenicity assessment for a particular time point will be excluded from the Per-Protocol Analysis Set for that time point onward. The protocol violations that impact the immunogenicity response and impact upon the Per-Protocol Analysis Set are detailed in the study deviation rules document.

The PP-Immunogenicity PBMC Subset will include all subjects in the Per-Protocol Analysis Set but further restricted to subjects who had blood samples taken and PBMCs harvested for analysis of cell-mediated immunity.

The PP-Immunogenicity Analysis Set will include all subjects who are randomized, received their full vaccination schedule at Day 35 (i.e. Day 0 and Day 21), have at least a baseline and a Day 35 serum sample IgG result available for analysis, have no major protocol violations up to and including Day 35 that impact immunogenicity response and no evidence of positive serology at baseline (ie, hepatitis B, hepatitis C, or HIV). Subjects who are unblinded – including those that chose to receive an approved/authorized COVID-19 vaccine – will be excluded from the PP-Immunogenicity Analysis Set the day following their date of unblinding. Subjects who received an approved/authorized COVID-19 vaccine but were not unblinded by their site will be excluded on the date of vaccination. Subjects who are unblinded unexpectedly will be reviewed via protocol deviation review meeting with the Study Deviations Rules document to determine their analysis set membership. The PP-Immunogenicity Analysis Set will be used for the primary immunogenicity analysis (i.e. Serum IgG antibody levels specific for the SARS-CoV-2 rS protein antigen[s] as detected by ELISA using GMT or SCR for the two-dose regimens at Day 35 regardless of baseline immune status). All subjects in the PP-Immunogenicity Analysis Set will be analyzed according to the randomized treatment assignment. If the number of subjects in the PP-Immunogenicity and PP Analysis Set differ (defined as the difference divided by the total number of subjects in the given PP analysis set) by more than 10%, supportive analyses of immunogenicity may be conducted using the PP analysis set.

The PP-Efficacy Analysis Set will be the PP Analysis Set restricted to subjects who received their full vaccination schedule (i.e. Day 0 and Day 21) except were not required to receive their booster dose(s). In addition, subjects will not be excluded from the analysis set due to their SARS-CoV-2 status except if positive SARS-CoV-2 status is observed at Visit 1 (Day 0). All subjects in the PP-Efficacy Analysis Set will be analyzed according to the randomized treatment assignment.

A listing of analysis sets will be provided.

### 5. Subject Disposition and Protocol Deviations

#### 5.1. Disposition

Subject disposition will be summarized for the All Screened Subjects Analysis Set. The number of subjects screened, number and percentage of subjects randomized, number and percentage of subjects who screen failed, and primary reason for screen failure will be summarized.

Subject disposition will also be summarized by treatment group and total for the Full Analysis Set. The number of subjects who are randomized in the study and the number and percentage of subjects who complete the study (i.e. study participation through the EOS Visit without early termination) will be presented. The frequency and percentage of subjects who withdraw or discontinue from the study and the reason for withdrawal or discontinuation will also be summarized. In addition, the frequency and percentage of subjects who are ongoing at the time of the data freeze will also be summarized for interim analyses. Lastly, the percent difference between the Per-Protocol Analysis Set and the PP-Immunogenicity Analysis Set will be presented (defined as the difference divided by the total number of subjects in the Per-Protocol Analysis Set). Subject disposition table will be presented by original planned treatment group assignment (Table 1A), across study time points within the re-randomized treatment groups (Table 1B) beginning at Day 189 (as the second reference time point), and for subjects in Treatment Groups B and C who agree to the 12-month booster dose (Table 1C) beginning at Day 357 (as the third reference time point).
If the day part is missing for subject withdrawal date, then impute the date with the below algorithm:

1. If last known contact date is in the same month and year as partial withdrawal date, then impute the partial date with last known contact date.
2. If last known contact date is in an earlier month and year than partial withdrawal date, then impute the partial date with the 1st day of the month.
3. If last known contact date is in a later month and year than partial withdrawal date, then impute the partial date with last known contact date.

Missing month and/or year part for subjects' withdrawal date will not be imputed.

Subject disposition data and randomization data will also be presented in data listings.

#### 5.2. Protocol Deviations

A protocol deviation is any change, divergence, or departure from the study design or procedures defined in the protocol. A major protocol deviation (sometimes referred to as an important or significant deviation) is a subset of protocol deviations that leads to a subject being discontinued from the study or significantly affects the subject's rights, safety, or well-being and/or the completeness, accuracy, and reliability of the study data, especially as it pertains to elicited immune response to planned study vaccination. A major deviation can include nonadherence to inclusion or exclusion criteria or nonadherence to regulatory authority including International Council for Harmonisation E6 (R2) guidelines.

Protocol deviations will be documented by the clinical monitor throughout the course of monitoring visits. The investigator will be notified in writing by the monitor of deviations. The human research ethics committee/IRB should be notified of all protocol deviations, if appropriate, in a timely manner.

Major protocol deviations will be summarized for the ITT analysis set.

All protocol deviations will be presented in a data listing, including the categorization of the deviation as major or not. Details of admission criteria deviations will be presented in a separate data listing. Protocol deviation table will be presented by original planned treatment group assignment (Table 1A) throughout the study.

### 6. Demographics and Baseline Characteristics

#### 6.1. Demographics

Demographic and baseline characteristics will be summarized by treatment group and total for all subjects in the Safety Analysis Set. This summary will be further delineated by age stratification (18-59 years, 60-84 years).

The demographic characteristics consist of age (years), age stratification (18-59 and 60-84 years), sex, child-bearing potential (percentage based on females), birth control method (percentage based on number of females), race, and ethnicity. The baseline characteristics consist of baseline weight (kg), baseline height (cm), baseline body mass index (BMI, $kg/m^2$), and baseline SARS-CoV-2 serostatus (positive/negative/indeterminate). Country and site will also be included in this summary.

Subject demographics table will be presented by original actual treatment group assignment (Table 1A) throughout the study. Subject demographic and baseline characteristics will also be presented in a data listing.

#### 6.2. Medical History

Medical history will be coded using the MedDRA (version to be delineated in the CSR).

Medical history will be summarized by SOC and PT by treatment group and total for the Safety Analysis Set. Medical history will also be presented in a data listing. Medical history table will be presented by original actual treatment group assignment (Table 1A) throughout the study.

### 7. Treatments, Medications, and Recent Vaccinations

#### 7.1. Recent Vaccinations and Concomitant Medications

Administration of medications, therapies, or vaccines will be recorded in the eCRF. Recent vaccinations will include vaccinations taken ≤ 90 days prior to taking study vaccination. Concomitant medications will include all medications (including vaccines) taken by the subject from first vaccination through EOS (or through the early termination visit if prior to that time).

Recent vaccinations and concomitant medications and therapies will be summarized for the Safety Analysis Set by treatment group, anatomical therapeutic chemical [ATC] class and preferred drug name as coded using the World Health Organization (WHO) Drug Dictionary (version to be delineated in the CSR). At each level of summarization (e.g. ATC class or preferred drug name), a subject is counted once if he/she reported one or more medications at that level.

For the purpose of inclusion in recent vaccination and concomitant medication tables, incomplete start and stop dates will be imputed according to the below rules (where UK, UKN, and UNKN indicate unknown or missing day, month, and year, respectively):

<u>Missing Start Dates</u>

- First, remove any non-missing date component following any missing date component (e.g. set day to UK if month equals UKN or set month and day to UK and UKN, respectively, if year equals UNKN)
- UK-MMM-YYYY: Assume 01-MMM-YYYY, but if month and year are the same as the first study vaccination month and year, then assume the date of first study vaccination
- UK-UKN-YYYY: Assume 01-JAN-YYYY, but if year is the same as the first study vaccination year, then assume the date and month of first study vaccination
- UK-UKN-UNKN: Assume date of first study vaccination

Should the imputed start date fall after a complete stop date provided, use the stop date instead of the date that would otherwise be imputed.

If the Concomitant medication pertains to EUA approved vaccines and with missing day part (i.e., UK-MMM-YYYY): Assume 01-MMM-YYYY.

<u>Missing Stop Dates</u>

- First, remove any non-missing date component following any missing date component (e.g. set day to UK if month equals UKN or set month and day to UK and UKN, respectively, if year equals UNKN)
- UK-MMM-YYYY: Assume the last day of the month
- UK-UKN-YYYY: Assume 31-DEC-YYYY
- UK-UKN-UNKN: Do not impute and assume ongoing

Should the imputed stop date fall before a complete start date provided, use the start date instead of the date that would otherwise be imputed.

Concomitant medication tables will be presented by the actual D189 treatment group (Table 1B). All recent vaccinations and concomitant medications will also be presented in a data listing.

#### 7.2. Medical and Surgical Treatment Procedures

The number and percentage of subjects with at least one medical and surgical treatment procedure during the study period will be summarized for the Safety Analysis Set Indication (i.e. Adverse Event vs. Other) and ongoing status for medical and surgical treatment procedures will also be summarized by frequency and percentage. Medical and Surgical tables will be presented by original actual treatment group (Table 1A) throughout the study. All medical and surgical treatment procedures will also be presented in a data listing.

#### 7.3. Study Vaccine Administration

Study vaccinations will comprise up to 3 IM injections (Day 0 and Day 21 [priming doses] and Day 189 [booster – active or placebo]), ideally in alternating deltoids for priming doses, with the study vaccine assigned in a full dose injection volume of approximately 0.5 mL (not to exceed 0.7 mL). Dose levels of SARS-CoV-2 rS (up to 2 doses) in Part 2 will be 5 µg and up to 25 µg, with 50 µg Matrix-M adjuvant.

Study vaccinations will be summarized including the number and percentage of subjects receiving each dose, whether administered per protocol or not, or not administered for the Full Analysis Set.

Study vaccinations will be presented by original planned treatment group (Table 1A) before Day 189. Planned D189 treatment group assignment will be presented on or after Day 189 (Table 1B).

All study drug administration data will also be presented in a data listing. Any treatment misallocation will be documented in a data listing.

### 8. Efficacy Analysis and Immunogenicity Analysis

#### 8.1. Efficacy Analysis

The efficacy analyses will be performed using the PP-Efficacy Analysis Set. All efficacy data will be listed for the Safety Analysis Set.

Numbers and percentages (with 95% CIs based on the exact Clopper-Pearson method) of subjects with occurrence of SARS-CoV-2 positivity following COVID-19 symptoms assessment from Day 28 through Day 217 will be summarized by overall, treatment group, COVID-19 disease severity (Table 3), and age strata (18-59, 60-84 years). Efficacy tables will be presented by original planned treatment assignment (Table 1A) before Day 189 and by D189 re-randomization treatment group assignment on or after Day 189 (Table 1B).

Assessment of SARS-CoV-2 by qualitative PCR testing based on routine screening without symptomatology will be done by nasal mid-turbinate sample self-collection from Day 28 through Day 217. Below classifications will be summarized by overall, treatment group, and age strata (18-59, 60-86 years). Asymptomatic infection, period of transmission, and other defined infection parameters will be assessed along with severity and progression of disease.

- **Asymptomatic infection** will be defined as a positive PCR-confirmed SARS-CoV-2 infection with no symptoms (regardless of past positivity) in the 7 days prior to self-collection.

- **Primary infection** will be defined as the first positive PCR-confirmed SARS-CoV-2 infection regardless of symptoms.

- **Primary symptomatic infection** will be defined as the first positive PCR-confirmed SARS-CoV-2 infection that is triggered by the symptomatic algorithm. A symptomatic infection will initiate further evaluation of physical status with severity scoring applied and monitoring of duration.

- **Period of transmission** for any subject (eg, transmission potential) is the period from a positive PCR test until the time of the first negative PCR test that follows a positive PCR test (if follow-up PCR test results are available for that subject). Period of transmission will only be calculated for illness episodes with a subsequent negative PCR test result at the time of the data cut for the given interim or final analysis.

- **Duration of resolved illness episodes** (i.e. number of days from the start of illness symptoms for illnesses followed up by FLU-PRO assessments until the day at which health returned to normal as reported in the electronic diary) classified by COVID-19 disease severity (Table 3). Duration will not be calculated for asymptomatic infections or illness episodes in which the subject's health has not returned to normal at the time of the data cut for the given interim or final analysis. Duration of resolved illness episodes will be omitted for Day 105.

Details related to sample collection (e.g. nasal mid-turbinate swab) for SARS-CoV-2 qualitative PCR test will be included in a data listing.

##### 8.1.1. Vaccine Efficacy

If frequent (i.e. 15% overall across all treatment groups) clinical PCR-confirmed SARS-CoV-2 infections occur during the study follow-up period (i.e. from Day 28 through Day 217), vaccine efficacy assessments for primary vaccination for active treatment groups compared to placebo may be generated. Vaccine efficacy (VE) is defined as follows:

$$VE\ (\%) = 100 \times (1 - RR)$$

where

RR = Relative Risk of incidence rates between the two treatment groups (SARS CoV-2 rs / Placebo)

A two-sided 95% CI for the VE will accompany the point estimate. The estimated RR and its associated 95% CI will be derived using Poisson regression with robust error variance (Zou, 2004). The explanatory variables in the model will include treatment group and age strata (18-59, 60-84 years). The dependent variable will be the incidence status as 0 (no infection) or 1 (one or more infections). The robust error variances will be estimated using the repeated statement and the subject identifier. The Poisson distribution will be used with a logarithmic link function.

#### 8.2. Immunogenicity Analysis

The primary immunogenicity analysis at Day 35 will be performed using the PP-Immunogenicity Analysis Set. All analyses involving cell-mediated response will be performed using the PBMC immunogenicity subset within the Per-Protocol Analysis Set. The remaining immunogenicity analyses will be performed using the Per-Protocol Analysis Set for the applicable visit. All immunogenicity data will be listed for the Full Analysis Set. The assessment for Wuhan strain will be analyzed per the SoE; however, the exploratory variants (e.g., South Africa variant B.1.351, Delta, and Omicron BA1/BA2/BA5) will be analyzed at various timepoints per available data import.

For the primary immunogenicity analysis (i.e. Serum IgG antibody levels specific for the SARS-CoV-2 rS protein antigen[s] as detected by ELISA using GMT or SCR for the two-dose regimens at Day 35 regardless of baseline immune status), the following treatment comparisons (consisting of differences of SCRs with 95% CI calculated using the method of Miettinen and Nurminen and ratios of geometric least-squares means and associated 95% CI from the ANCOVA model described in Section 8.2.1) will be performed:

- Two-dose regimens by dose compared to placebo:
  B vs A; D vs A; (B+D) vs A (Table 1A)

For analyses involving immunogenicity-related secondary endpoints, the following treatment comparisons (consisting of differences of SCRs with 95% CI calculated using the method of Miettinen and Nurminen and ratios of geometric least-squares means and associated 95% CI from the ANCOVA model described in Section 8.2.1) will be performed:

- Treatment comparisons will consist of the following including single-dose regimens compared to the two-dose regimens and to placebo:
  B vs A;
  C vs B; C vs D; C vs (B+D) (at Day 35 and following); C vs A;
  D vs A;
  E vs B; E vs D; E vs (B+D) (at Day 35 and following); E vs A;
  (B+D) vs A (at Day 35 and following); (C+E) vs A (at Day 35 and following);
  (B+C) vs A (at Day 21 only); (B+C) vs (D+E) (at Day 21 only);
  (B+D) vs (C+E) (at Day 35 and following);
  (D+E) vs A (at Day 21 only) (Table 1A). However, treatment comparisons for GMFR and SCR (≥ 4-fold change) will not be performed on or after Day 189 nor will they be performed for variants; instead, analyses will be presented across study time points within the re-randomized treatment groups (Table 1B) beginning at Day 189 (as the second reference time point) and for subjects in Treatment Groups B and C who agree to the 12-month booster dose (Table 1C) from Day 357 (included as the third reference timepoint) to Day 546 regardless of baseline immune status.

###### 8.2.1. Serum IgG Antibody Levels and Serum ACE-2 Receptor Binding Inhibition Specific for SARS-CoV-2 rS Protein Antigen

The following evaluations will be performed for the serum IgG antibody levels and serum ACE-2 receptor binding inhibition specific for the SARS-CoV-2 rS protein antigen(s), as detected by ELISA separately:

- GMT (reported in GMEUs) with 95% CI at each immunogenicity visit. The 95% CI will be calculated based on the t-distribution of the log-transformed values for GMTs, then back transformed to the original scale for presentation. An analysis of covariance

(ANCOVA) model will be constructed at each post-vaccination immunogenicity visit on the log transformed titer, including the treatment group, site, and age strata (18-59; 60-84 years) as fixed effects and the baseline log transformed titer as a covariate. If the model fails to converge for interim analyses involving approximately 150 subjects per treatment group (or more), it will be attempted without the age strata and/or site covariates. If the model fails to converge for preliminary database freezes involving 50 subjects per treatment group, no changes to the model will be made. Comparisons of selected treatment groups will be performed within each post-baseline visit and no adjustments for multiplicity will be carried out.

- For ANCOVA analyses involving immunogenicity-related secondary endpoints before Day 189 group comparison will be performed with below treatment comparison:
  B vs A;
  C vs B; C vs D; C vs (B+D) (at Day 35 and following); C vs A;
  D vs A;
  E vs B; E vs D; E vs (B+D) (at Day 35 and following); E vs A;
  (B+D) vs A (at Day 35 and following); (C+E) vs A (at Day 35 and following);
  (B+C) vs A (at Day 21 only); (B+C) vs (D+E) (at Day 21 only);
  (B+D) vs (C+E) (at Day 35 and following);
  (D+E) vs A (at Day 21 only) (Table 1A).

- For ANCOVA visit based analyses involving immunogenicity-related secondary endpoints on or after Day 189 (inclusive); re-randomization treatment group (i.e., A, B1, B2, C1, C2, D, and E) will be reported with Geometric Least Square Mean and 95% CI at each subsequent visit. No treatment comparison will be performed on or after Day 189 (inclusive).

- ANCOVA model will only be produced for Wuhan strain.

- ANCOVA model will be produced for subjects in Treatment Groups B and C who agree to the 12-month booster dose phase (i.e., after Day 357) for immunogenicity-related secondary endpoints.

- GMFR (compared to baseline [Day 0]) with 95% CI at each post-vaccination immunogenicity visit. The 95% CI will be calculated based on the t-distribution of the log-transformed fold-rise values for GMFRs, then back transformed to the original scale for presentation. Comparisons of GMFR between selected treatment groups will also be performed within each post-vaccination visit. Similar analyses will be performed with GMFR comparing post- (Day 217) to pre- (Day 189), and post- (Day 357) to pre- (Day 189); however, only across timepoints will be included based on the re-randomized treatment groups (Table 1B) on or after Day 189 (as second reference time point). For subjects in Treatment Groups B and C who agree to the 12-month booster dose; GMFR, GMFRs comparing post- (Day 371) to pre- (Day 357), and post- (Day 546) to pre- (Day 357) will be presented.

- SCRs with 95% CI based on the exact Clopper-Pearson method at each post-vaccination immunogenicity visit prior to Day 189. Seroconversion will be defined as ≥ 4-fold increases in titers at each post vaccination immunogenicity visit. Comparisons of SCR between selected treatment groups will also be performed within each visit. Differences between selected treatment groups along with 95% CIs within each visit will be calculated using the method of Miettinen and Nurminen (Miettinen and Nurminen, 1985). The SCRs with ≥ 4-fold increases in titers will only be performed across timepoints by utilizing Day 189 as second baseline. Similarly, Day 357 will be utilized as third reference time point for subjects in Treatment Groups B and C with 12-month booster dose.

The GMT will be calculated using the following formula:

$$\text{GMT} = 10^{\left\{\frac{\sum_{i=1}^{n} log_{10}(t_i)}{n}\right\}}$$

where $t_1$, $t_2$, … $t_n$ are observed immunogenicity titers for n subjects.

The GMFR measures the changes in immunogenicity titers within subjects. The GMFR will be calculated using the following formula:

$$\text{GMFR} = 10^{\left\{\frac{\sum_{i=1}^{n} \log_{10} {}^{v_{ij}}/_{v_{ik}}}{n}\right\}} = 10^{\frac{\{\sum_{i=1}^{n}(\log_{10} v_{ij} - \log_{10} v_{ik})\}}{n}}$$

where, for n subjects, $v_{jj}$ and $v_{ik}$ are observed immunogenicity titers for subject i at time points j and k=0 (Baseline, Day 0).

Subgroup analyses for serum IgG antibody levels at Day 35 by baseline serostatus (positive vs negative/indeterminate) will be performed where positive is defined as a serum IgG antibody result ≥ 1000 ELISA units. Subgroup analyses for serum IgG antibody levels and serum ACE-2 receptor binding inhibition by age stratification (18-59, 60-84 years) will also be performed separately; age strata will be removed from the ANCOVA model in these analyses.

Line plots (including 95% CIs) of GMT and GMFR for serum IgG antibody levels, serum ACE-2 receptor binding inhibition by treatment group and visit will be provided. Box plots of titer and fold rise for serum IgG antibody levels, serum ACE-2 receptor binding inhibition by treatment group and visit will be provided. In addition, scatter plots of serum IgG antibody levels and serum ACE-2 receptor binding inhibition versus Neutralization Assay Specific for SARS-CoV-2 Wildtype (or Variant) results will be provided. Lastly, plots of the reverse cumulative distribution curves also will be provided for serum IgG antibody levels and serum ACE-2 receptor binding inhibition by treatment group and visit. All plots will be presented with original planned treatment group assignment (Table 1A) before Day 189 re-randomization. Re-randomization treatment group assignment will be presented on or after Day 189 (Table 1B). For subjects in treatment groups B1, B2, C1, and C2, plots for only those who agree to the 12-month booster will be presented on or after Day 357 (Table 1C).

At Day 546, end of extension study, the exploratory variants (e.g., South Africa variant B.1.351, Delta, and Omicron BA1/BA2/BA5) will be analyzed per available data import.

###### 8.2.2. Neutralization Assay Specific for SARS-CoV-2 Wildtype (or Variant)

An analysis similar to the serum IgG antibody levels and serum ACE-2 receptor binding inhibition specific for the SARS-CoV-2 rS protein antigen will be performed based on a subset of subjects within the Per-Protocol Analysis Set. The average absorbance for the positive virus and negative cell only wells will be used to calculate the virus neutralizing serum endpoint titer.

A line plot (including 95% CIs) of GMT and GMFR for neutralization results by treatment group and visit will be provided. In addition, box plots of titer and fold rise by treatment group and visit will be provided. Lastly, a plot of the reverse cumulative distribution curves will also be provided by treatment group and visit. All plots will be presented with original planned treatment group assignment (Table 1A) before Day 189 re-randomization. Re-randomization treatment group assignment will be presented on or after Day 189 (Table 1B). For subjects in treatment groups B1, B2, C1, and C2, plots for only those who agree to the 12-month booster will be presented on or after Day 357 (Table 1C).

###### 8.2.3. Cell-Mediated Response for Type 1 T Helper (Th1) and Type 2 T Helper (Th2)

Cell-mediated response (immunity) for both Th1 and Th2 pathways will be assessed by cytokine profiling in harvested PBMCs and summarized by treatment group as mean, SD, median, min, max, and geometric mean of actual value and as mean, SD, median, min, and max of change from baseline as well as GMFR by visit based on subjects in the PBMC immunogenicity subset within the Per-Protocol Analysis Set. Summaries based on ELISpot assay results at Days 0, 7, and 28 (expressed as counts per 1 million cells) will include the following cytokines using separately peptide protein and spike protein: IFNg+, TNFa+, IFNg+TNFa+ (double positive), and IL5+. Summaries based on Intracellular Cytokine Staining (ICCS) assay results at Days 0 and 28 (expressed as counts per 1 million CD4 cells) will include the following cytokines using separately Nvx-2373 protein and a spike peptide pool: IFNg+, IFNg+IL2+TNFa-, IFNg+IL2-TNFa+, IFNg-IL2+TNFa+, IFNg+IL2+TNFa+, IL2+, IL5+, IL13+, IL5+IL13+, TNFa+, and Any 2 of Th1 (as defined below).

Any 2 = Sum(IFNg+IL2+TNFa-, IFNg+IL2-TNFa+, IFNg-IL2+TNFa+)
+ IFNg+IL2+TNFa+

Box plots of cell-mediated response for cytokines by treatment group, parameter, and visit will be provided.

All cytokine data will also be presented in a data listing for FAS.

###### 8.2.4. Anti-Nucleocapsid Protein

Anti-Nucleocapsid Protein will be collected at Days 371 and Day 546 (i.e., in the 12-month booster period). Subjects' Anti-Nucleocapsid Protein results will be based on their Per-Protocol (PP) Analysis Set membership for the two study visits mentioned above. No statistical analysis or figures will be performed for this assay; however, a data listing for FAS will be provided.

##### 8.3. Exploratory Analysis

Daily FLU-PRO scores (i.e. total score, domain subgroup scores, and severity) will be entered by the subject following self-reported SARS-CoV-2 positive illnesses, which will be summarized by D189 re-randomization treatment group, total, and age strata (18-59, 60-84 years) using descriptive statistics from day 1 to day 10 (if applicable) in cases where subjects have one or more qualifying illness episode for the Safety Analysis Set. The 32-item level responses for the FLU-PRO range from 0 (symptom free) to 4 (very severe symptoms) and will be reported daily for each episode. Then the mean domain subgroup scores and mean total score are calculated and available in the data extraction. The six domain subgroups scores include below aspects:

- Body/Systemic
- Chest/Respiratory
- Eyes
- Gastrointestinal
- Nose
- Throat

In addition, the severity responses include: 'No infection symptoms today', 'Mild', 'Moderate', 'Severe', and 'Very severe'. If a subject experiences multiple illness episodes during the study, data from each episode will be included in the descriptive statistics.

Six qualitative questions will also be listed for each episode on daily basis:

- Loss of Smell (Yes/No)?
- Loss of Taste (Yes/No)?
- How much did your infection symptoms interfere with your usual activities today (Not at all/A little bit/Somewhat/Quite a bit)?

- Have you returned to your usual activities today (Yes/No)?
- In general, how would you rate your physical health today (Excellent/Very Good/Good/Fair/Poor)?
- Have you returned to your usual health today (Yes/No)?

Maximum FLU-PRO total, domain subgroup scores, and severity reported during the daily follow-up for a COVID-19 illness episode (up to 10 days) will be summarized by treatment group, total, and age strata (18-59, 60-84 years) using descriptive statistics for the Safety Analysis Set. If a subject experiences multiple illness episodes during the study, data from each episode will be included in the descriptive statistics. Subjects enrolled at Australian sites will be excluded in this analysis following approval of Amendment 7 (Version 8.0) of the protocol.

An optional correlation analysis between the following FLU-PRO scores with COVID-19 disease severity (1=Asymptomatic case, 2=Virologically Confirmed, 3=Mild, 4=Moderate, 5=Severe) associated with qualitative PCR-confirmed COVID-19 illness episodes reported by subjects in the FLU-PRO may be performed by treatment group and total for the Safety Analysis Set:

- Maximum FLU-PRO daily total score per illness episode
- Maximum daily domain subgroup scores per illness episode
- Maximum daily severity per illness episode

If the COVID-19 disease episode overlapped with the FLU-PRO episode window, then those data will be considered paired for correlation analysis. The only overlapped scenarios are provided below with illustrations. If multiple COVID-19 disease episodes overlapped with the same FLU-PRO episode window, then the most severe will be used. Likewise, the maximum FLU-PRO daily score, domain subgroup score or severity will be used for each episode. If  COVID-19 disease episode end date is missing but start date happened on or before the end date of FLU-PRO end date and with ongoing status, these events are also considered overlapped and will be considered for correlation analysis (see illustration 3). If a subject experiences multiple overlapping illness episodes during the study, data from each FLU-PRO episode will be included in the analysis. The measure of correlation presented will be the Pearson product-moment correlation coefficient (range -1 to 1) and the non-parametric Spearman rank correlation.

Below are some illustrations for overlapping COVID-19 disease and FLU-PRO episodes in the optional correlation analysis:

Illustration 1: Flu-PRO episode started within the window of COVID-19 disease episode, or vice versa.

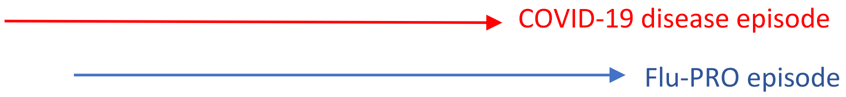

Illustration 2: Flu-PRO episode started and ended within a COVID-19 disease episode, or vice versa.

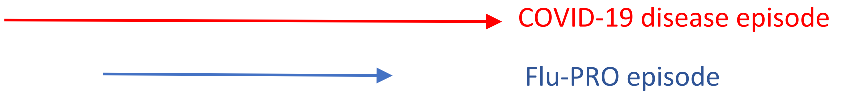

Illustration 3: COVID-19 disease episode started before the end date of Flu-PRO episode with ongoing status, but without end date.

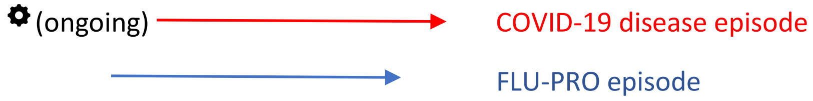

#### 9. Safety Analysis

All safety summaries and analyses will be conducted for the Safety Analysis Set.

All safety data collected will be included in the applicable summaries regardless of whether a subject has received incorrect treatment provided the subject is included in the Safety Analysis Set. In the safety tables where different vaccination periods are mentioned, subjects who miss one or more planned vaccinations will be adjusted via dynamic header; i.e., only subjects receive corresponding vaccine will be considered in the denominator within each vaccination period to avoid inflation. Safety data will be presented by actual treatment received at first dose prior to Day 189 (Table 1A) and by actual treatment received following re-randomization (Table 1B) on or after Day 189. Independent safety summaries will be produced for subjects in Treatment Groups B or C (Table 1C) who agree to the 12-month booster dose.

Due to many patients expected to receive an EUA approved non-study COVID-19 vaccine, Vaccine-Exposure Adjusted Event Rate (VEAER) analysis will be performed for the occurrence of adverse events within a specified period of time in order to minimize potential bias; in addition to standard frequency and percentage AE summaries. The numerator for the VEAER will be the number of Adverse Events that occurred during the specific period being summarized and before the subject is unblinded for any reason or takes an EUA approved vaccine (whichever is earlier). The denominator for the VEAER will be the summation of risk exposure in years for the subjects in specific treatment groups. For unsolicited AEs, the VEAER will be reported in the fashion of 100 person-years.

The VEAER per 100 person-years will be calculated as:

$$VEAER = \frac{Event}{sum(Days\ at\ risk)} \times 365.25\ \times 100$$

The overall exposure of days at risk will be calculated from the date of the *described* dose (i.e., first, second, or booster dose) to one day prior to the first dose date of an approved vaccine, date of unblinding, or truncation date (e.g. 21 days after first vaccine), whichever is earlier. For subjects who do not have an approved vaccine or date of unblinding the overall exposure period is from the date of the *described* dose to date of study discontinuation, data cut-off, or truncation date, whichever is earlier.

The details of subjects' days at risk are specified for the following periods:

- The entire study overall:
    - For subjects who receive EUA vaccine or are unblinded for any reason, the days at risk will be calculated as earliest date of EUA vaccine dose date or unblinded date minus the first dose date.
    - For subjects who do not receive EUA vaccine and are not unblinded for any reason, the days at risk will be calculated as date of study discontinuation, data cut-off, or truncation date, whichever is earlier minus date of the first dose + 1.

- Vaccination Dose 1 (i.e. the AE events are observed from first vaccination but prior to the second vaccination for subjects that receive the second vaccination and through 21 days *following* the first vaccination for subjects that do not receive the second vaccination)
    - For subjects who receive EUA vaccine or are unblinded for any reasons, the days at risk will be calculated as:
        - If subjects receive the second vaccination dose, the days at risk = Earliest date of (second vaccination dose date, EUA vaccine dose date or unblinded date) minus the first dose date.
        - If subjects do not receive the second vaccination dose, the days at risk = Earliest date of (first dose date + 21, EUA vaccine dose date or unblinded date) minus the first dose date.
    - For subjects who do not receive EUA vaccine and are not unblinded for any reason, the days at risk will be calculated as:

        - If subject receives the second vaccination dose, the days at risk = Earliest date of (second vaccination dose date-1, study discontinuation, data cut-off, or truncation date) minus the first dose date + 1.
        - If subjects do not receive the second vaccination dose, the days at risk = Earliest date of (first dose date + 20, study discontinuation, data cut-off, or truncation date) minus the first dose date + 1.

- Vaccination Dose 2 (i.e. from second vaccination but prior to the 6-month boost vaccination for subjects that received the 6-month boost vaccination, and through 167 days *following* the second vaccination for subjects that do not receive the third dose/6-month boost vaccination)
    - For subjects who receive EUA vaccine or are unblinded for any reason, the days at risk will be calculated as:
        - If subjects receive the third vaccination dose, the days at risk = Earliest date of (third vaccination dose date, EUA vaccine dose date or unblinded date) minus the second dose date.
        - If subjects do not receive the third vaccination dose, the days at risk = Earliest date of (second dose date + 167, EUA vaccine dose date or unblinded date) minus the second dose date.
    - For subjects who do not receive EUA vaccine and are not unblinded for any reason, the days at risk will be calculated as:
        - If subjects receive the third vaccination dose, the days at risk = Earliest date of (third vaccination dose date - 1, study discontinuation, data cut-off, or truncation date) minus the second dose date + 1.
        - If subjects do not receive the third vaccination dose, the days at risk = Earliest date of (second dose date + 166, study discontinuation, data cut-off, or truncation date) minus the second dose date + 1.

- Vaccination Dose 3 (i.e. from 6-month boost vaccination through the EOS visit or through the early termination visit if prior to that time for subjects in treatment group A, D, E or those who disagree to the 12-month boost; or from 6-month boost vaccination but prior to the 12-month boost vaccination for subjects in treatment groups B or C that received the 12-month boost vaccination and through 167 days *following* the 6-month boost vaccination for subjects in treatment groups B or C that do not receive the 12-month boost vaccination)
    - For subjects in treatment group A, D, E, or those who disagree to the 12-month booster:
        - For subjects who receive EUA vaccine or are unblinded for any reason, the days at risk will be calculated as earliest date of EUA vaccine dose date or unblinded date minus the third dose date.
        - For subjects who do not receive EUA vaccine and are not unblinded for any reason, the days at risk will be calculated as date of study

discontinuation, data cut-off, or truncation date, whichever is earlier minus date of the third dose date + 1 .

- For subjects in treatment group B and C who agree to the 12-month boost vaccination:
  - For subjects who receive EUA vaccine or are unblinded for any reason, the days at risk will be calculated as:
    - If subjects receive the fourth vaccination dose, the days at risk = Earliest date of (fourth vaccination dose date, EUA vaccine dose date or unblinded date) minus the third dose date.
    - If subjects do not receive the fourth vaccination dose, the days at risk = Earliest date of (third dose date + 167, EUA vaccine dose date or unblinded date) minus the third dose date.
  - For subjects who do not receive EUA vaccine and are not unblinded for any reason, the days at risk will be calculated as:
    - If subjects receive the fourth vaccination dose, the days at risk = Earliest date of (fourth vaccination dose date-1, study discontinuation, data cut-off, or truncation date) minus the third dose date + 1.
    - If subjects do not receive the fourth vaccination dose, the days at risk = Earliest date of (third dose date + 166, study discontinuation, data cut-off, or truncation date) minus the third dose date + 1.

- Vaccination Dose 4 (i.e. from 12-month boost vaccination through the EOS visit or through the early termination visit if prior to that time for subjects in treatment groups B or C)
  - For subjects who receive EUA vaccine or are unblinded for any reason, the days at risk will be calculated as earliest date of EUA vaccine dose date or unblinded date minus the fourth dose date.
  - For subjects who do not receive EUA vaccine and are not unblinded for any reason, the days at risk will be calculated as date of study discontinuation, data cut-off, or truncation date, whichever is earlier minus date of the fourth dose date + 1.

- 1$^{st}$ Boost vaccination through 28 days *after* 6-month boost vaccination
  - The days at risk will be taking the minimal values between days at risk for Vaccination Dose 3 and 28 days.

- 2$^{nd}$ Boost vaccination through 28 days *after* 12-month boost vaccination
  - The days at risk will be taking the minimal values between days at risk for Vaccination Dose 4 and 28 days.

- Treatment-emergent MAAEs through 217 days *after* first vaccination

  - The days at risk will be taking the minimal values between days at risk for Vaccination Dose 1 and 217 days.

- TEAEs through 35 days *after* first vaccination
  - The days at risk will be taking the minimal values between days at risk for Vaccination Dose 1 and 35 days.

For solicited AEs, the VEAER will not be reported as solicited AEs are observed in a short time period (7-days).

#### 9.1. Unsolicited Adverse Events

An AE is defined as any untoward medical occurrence in a subject enrolled into this study regardless of its causal relationship to study vaccination. Adverse events will be captured after first dose of study vaccination administered with the exception of an AE related to study procedure or one that causes a delay in study vaccination administration (e.g., acute illness). Subjects will be instructed to contact the investigator at any time after randomization if any symptoms develop.

A treatment-emergent AE (TEAE) is defined as any event not present before exposure to study vaccination or any event already present that worsens in intensity or frequency after exposure. Adverse events will also be evaluated by the investigator for the coexistence of MAAEs which are defined as AEs that lead to an unscheduled visit to a healthcare practitioner.

Subjects will be assessed for diagnosis of an AESI at all study visits. Adverse events of special interest include potential immune-mediated medical conditions (PIMMC), AESIs relevant to COVID-19 disease, or other potential AEs that may be determined at any time by regulatory authorities as additional information concerning COVID-19 is obtained. Given the concern for cytokine storm, an AESI of cytokine release syndrome will be included as an AESI relevant to COVID-19 disease. Listings of AESIs are provided in Protocol Appendix 4, Tables 16-5 and 16-6.

Adverse events will be coded by preferred term (PT) and system organ class (SOC) using MedDRA (version to be delineated in the CSR).

Solicited AEs continuing from data recorded in the ePRO (i.e. those extending >6 days *after* vaccination) and recorded on the AE CRF page will be excluded from all unsolicited AE summaries with the exception of those that are designated as serious.

Only TEAEs will be included in summary tables and will be summarized by treatment group and vaccination (first, second, boost, and overall). Percentages will be based upon the number of subjects in the Safety Analysis Set within each vaccination group. A subject with 2 or more AEs within the same level of summarization (e.g. SOC or PT) will be counted only once in that level.

For the purpose of inclusion in TEAE tables, incomplete AE onset and end dates will be imputed according to the below rules (where UK, UKN, and UNKN indicate unknown or missing day, month, and year, respectively):

Missing Onset Dates

- First, remove any non-missing date component following any missing date component (e.g. set day to UK if month equals UKN or set month and day to UK and UKN, respectively, if year equals UNKN)
- UK-MMM-YYYY: Assume 01-MMM-YYYY, but if month and year are the same as the first study vaccination month and year, then assume the date of first study vaccination
- UK-UKN-YYYY: Assume 01-JAN-YYYY, but if year is the same as the first study vaccination year, then assume the date and month of first study vaccination
- UK-UKN-UNKN: Assume date of first study vaccination

Should the imputed start date fall after a complete stop date provided, use the stop date instead of the date that would otherwise be imputed.

Missing End Dates

- First, remove any non-missing date component following any missing date component (e.g. set day to UK if month equals UKN or set month and day to UK and UKN, respectively, if year equals UNKN)
- UK-MMM-YYYY: Assume the last day of the month
- UK-UKN-YYYY: Assume 31-DEC-YYYY
- UK-UKN-UNKN: Do not impute and assume ongoing

Should the imputed stop date fall before a complete start date provided, use the start date instead of the date that would otherwise be imputed.

All AEs will also be presented in a data listing.

##### 9.1.1. Incidence of Adverse Events

An overview of TEAEs will be presented by vaccination period (Entire Study Overall, Vaccination Dose 1, Vaccination Dose 2, and Vaccination Dose 3), treatment group, total, and age strata (18-59, 60-84 years) including number and percentage of subjects with any of the following:

- TEAEs
- TEAEs through 35 days *after* first vaccination and from 6-month or 12-month boost vaccinations until 28 days *after* corresponding boost vaccinations
- Severe TEAEs

- Severe TEAEs through 35 days *after* first vaccination, and from 6-month or 12-month boost vaccinations until 28 days *after* corresponding boost vaccinations
- Treatment-related TEAEs
- Treatment-related TEAEs through 35 days *after* first vaccination and from 6-month or 12-month boost vaccinations until 28 days *after* corresponding boost vaccination
- Treatment-related severe TEAEs
- Treatment-emergent MAAEs attributed to vaccine
- Treatment-emergent MAAEs through 217 days *after* first vaccination
- Serious TEAEs
- TEAEs leading to vaccination discontinuation
- TEAEs leading to study discontinuation
- AESIs: PIMMC
- AESIs: Relevant to COVID-19
- Treatment-related MAAEs
- Treatment-related serious TEAEs
- Treatment-related TEAEs leading to vaccination discontinuation
- Treatment-related TEAEs leading to study discontinuation
- Treatment-related AESIs: PIMMC
- Treatment-related AESIs: Relevant to COVID-19
- Solicited Local TEAE
- Solicited Local TEAE Grade 3 or Higher
- Solicited Systemic TEAE
- Solicited Systemic TEAE Grade 3 or Higher

All TEAEs will also be presented in summary tables by age strata (18-59, 60-84 years), treatment group, total, SOC and PT separately for each of the following periods:

- The entire study overall
- Vaccination Dose 1 (i.e. from first vaccination but prior to the second vaccination for subjects that received the second vaccination and through 21 days *following* the first vaccination for subjects that did not receive the second vaccination)
- Vaccination Dose 2 (i.e. from second vaccination but prior to the boost vaccination for subjects that received the boost vaccination and through 167 days *following* the second vaccination for subjects that did not receive the boost vaccination)
- Vaccination Dose 3 (i.e. from 6-month boost vaccination through the EOS visit or through the early termination visit if prior to that time for subjects in treatment group A, D or E; or from 6-month boost vaccination but prior to the 12-month boost vaccination for subjects in treatment group B and C that received the 12-month boost vaccination and through 167 days *following* the 6-month booster vaccination for subjects in treatment groups B or C that do not receive the 12-month boost vaccination)

- Vaccination Dose 4 (i.e. from 12-month boost vaccination through the EOS visit or through the early termination visit if prior to that time for subjects in treatment groups B or C)
- Through 35 days *after* first vaccination
- From 6-month boost vaccination until 28 days *after* 6-month boost vaccination
- From 12-month boost vaccination until 28 days *after* 12-month boost vaccination for subjects that received 12-month boost vaccination

Summary tables in which the reporting period is through 35 days *after* first vaccination will include data from the first dose date to the second dose date (denoted as “Vaccination Dose 1”), from second dose date to 35 days *after* first vaccination (denoted as “Vaccination Dose 2”), and from first dose date to 35 days *after* first vaccination (denoted as “Overall”).

Summary tables in which the reporting period is from 6-month boost vaccination until 28 days *after* 6-month boost vaccination will include data from the third dose date to 28 days *after* third dose date.

For subjects in treatment groups who received 12-month boost vaccination, summary tables in which the reporting period is from 12-month boost vaccination until 28 days *after* boost vaccination will include data from the fourth dose date to 28 days *after* fourth dose date.

##### 9.1.2. Relationship of Adverse Events to Study Vaccine

The relationship or association of the study vaccine in causing or contributing to the AE will be characterized by the investigator as “Related” or “Not Related”.

All TEAEs will be presented in a summary table for each treatment group and total by SOC, PT, and relationship to study vaccine. Separate summaries will be provided for TEAEs in each vaccination period, through 35 days after first vaccination, and from boost vaccination until 28 days after boost vaccination (6-month and 12-month boost vaccination included). If a subject has 2 or more TEAEs in the same SOC (or within the same PT) with a different relationship to study vaccine, then the subject will be counted under maximum relatedness (i.e. “Related”). If the relationship information is missing, the AE will be considered related in the summary but will be presented as missing in the data listings.

A listing will also be presented for unsolicited treatment-related TEAEs.

##### 9.1.3. Severity of Adverse Event

The severity (or intensity) of an AE refers to the extent to which it affects the subject's daily activities and will be classified as mild, moderate, or severe using the following criteria:

- Mild: These events require minimal or no treatment and do not interfere with the subject's daily activities.
- Moderate: These events result in a low level of inconvenience or require minor therapeutic measures. Moderate events may cause some interference with normal functioning.
- Severe: These events interrupt a subject's usual daily activity and may require systemic drug therapy or other treatment. Severe events are usually incapacitating.

All TEAEs and treatment-related TEAEs will be summarized for each treatment group and total by maximum severity, SOC, and PT. Separate summaries will be provided for TEAEs in each vaccination period, through 35 days after first vaccination, and from boost vaccination until 28 days after boost vaccination; this includes the 6-month and 12-month boost vaccinations. If a subject has more than one AE on the same level (e.g. SOC or PT), the most severe AE will be summarized. If the severity information is missing, the AE will be considered severe in the summary but will be presented as missing in the data listings. If a solicited adverse event (e.g., reactogenicity) exceeds the duration of 7 days follow vaccination, then that event will be captured as an unsolicited adverse event and further classified by severity as above (not by toxicity grade) and followed to conclusion. The onset of that event will be noted to be associated with the date of onset during the period of solicited reactogenicity.

##### 9.1.4. Serious Adverse Events

An SAE is defined as any event that

- Results in death
- Is Immediately life-threatening
- Requires inpatient hospitalization or prolongation of existing hospitalization
- Results in persistent or significant disability/incapacity
- Is a congenital anomaly/birth defect

Important medical events that may not result in death, be life threatening, or require hospitalization may be considered SAEs when, based upon appropriate medical judgment, they may jeopardize the subject or may require medical or surgical intervention to prevent one of the outcomes listed in this definition. Examples of such medical events include allergic bronchospasm requiring intensive treatment in an emergency room or at home, blood dyscrasias or convulsions that do not result in inpatient hospitalization, or the development of drug dependency or drug abuse.

All Serious TEAEs will be presented in a summary table for each treatment group, total, and age strata (18-59, 60-84 years) by SOC, PT, and relationship to study vaccine. All SAEs will also be presented in a data listing.

###### 9.1.5. Medically-Attended Adverse Events

All MAAEs through 217 days after first vaccination and treatment-related MAAEs through the end of study will be presented in separate summary tables for each treatment group, total, and age strata (18-59, 60-84 years) by SOC, PT, and relationship to study vaccine. All MAAEs will also be presented in a data listing.

###### 9.1.6. Adverse Events of Special Interest

All AESIs associated with PIMMC through end of study will be presented in separate summary tables for each treatment group, total, and age strata (18-59, 60-84 years) by SOC, PT, and relationship to study vaccine. AESIs associated with PIMMC and COVID-19 disease potential exacerbation will be listed separately.

###### 9.1.7. Adverse Events Leading to Study Vaccine Discontinuation

All AEs leading to study vaccine discontinuation will be presented in a data listing.

###### 9.1.8. Adverse Events Leading to Discontinuation from Study

All AEs leading to discontinuation from study will be presented in a data listing.

##### 9.2. Solicited Adverse Events

Solicited local and general systemic reactogenicity will be assessed for occurrence and intensity (toxicity grade) of selected signs and symptoms from the subject during a specific post-vaccination follow-up period (day of vaccination and 6 subsequent days), using Immediate Reactogenicity – Local and Immediate Reactogenicity – Systemic CRFs and also a pre-defined checklist in their electronic diary. In general, toxicity grading will be provided by subjects in the ePRO diary according to the FDA toxicity grading scale (Protocol Appendix 3, Table 16-1). For solicited local symptoms of redness and swelling, toxicity grades 1 to 3 will be derived in the analysis database based on redness size and swelling size, respectively. For the solicited systemic symptom of temperature, toxicity grades 1 to 4 will be derived in the analysis database based on temperature. Should any reactogenicity event extend beyond 6 days after vaccination (toxicity grade ≥1), then it will be recorded as an AE with a start date corresponding to Day 7 following the most recent vaccination (i.e. the first day after completion of the ePRO diary) as the reactogenicity event and followed to resolution per FDA guidelines for dataset capture.

The following local AEs will be solicited at the injection site: pain, tenderness, erythema/redness, and induration/swelling.

The following systemic AEs will be solicited: fever, nausea/vomiting, headache, fatigue, malaise, myalgia (muscle pain), arthralgia (joint pain).

The number and percentage (with 95% CIs based on the exact Clopper-Pearson method) of subjects who report each solicited AE on the day of vaccination and through the 6 subsequent days (i.e. daily on Day 0 through Day 6, daily on Day 21 through Day 27, daily on Day 189 through Day 195, and overall for each of these 7 day periods) will be tabulated by maximum toxicity grade and treatment group for each vaccination. Subjects will be counted once for the most severe event in cases where the subject reported more than one event per solicited AE category (e.g. pain). If the toxicity grade is missing for a given solicited AE, then it will not be imputed and the solicited AE will be summarized in the “Grade Missing” category. Percentages will be based upon the number of subjects in the Safety Analysis Set within each vaccination group who reported data for the respective category. In addition, the duration (in days) of solicited AEs after each vaccination will be summarized by treatment group for each vaccination. For solicited AEs occurring <=6 days after vaccination, duration will correspond to the number of days reported by the subject with toxicity grade >=1 for a given solicited AE category in the ePRO diary. For solicited AEs extending >6 days after vaccination, duration will be calculated as (AE End Date as recorded on the Adverse Events CRF) – (AE Start Date as derived based on initial study day of occurrence relative to date of recent vaccination) + 1 day. If AE Start Date is a partial date or completely missing for AEs extending >6 days after vaccination, then the imputation rules outlined in Section 9.1 will be followed. If AE End Date is a partial date for AEs extending >6 days after vaccination, then the imputation rules outlined in Section 9.1 will be followed. If AE End Date is completely missing, then it will be imputed using the data cutoff date for the given analysis.

All solicited local and systemic AEs will be presented in data listings.

If more than 10% of total subjects are seropositive to SARS-CoV-2 infection at baseline, exploratory analyses by SARS-CoV-2 positivity at Day 0 may be performed.

#### 9.3. Clinical Laboratory Evaluations

The following clinical laboratory assessments will be recorded:

| | |
|---|---|
| Hematology | Hemoglobin, hematocrit, platelet count, and complete white blood cell count |
| Serum Chemistry | Alanine aminotransferase, aspartate aminotransferase, total bilirubin, creatinine |
| Serology | Hepatitis B surface antigen, hepatitis C virus antibody, and human immunodeficiency virus antibody types 1 and 2 |
| Other analyses | Female subjects: Urine pregnancy test at screening and prior to each vaccination (human chorionic gonadotropin) ;serum FSH may be requested to confirm postmenopausal status |

Note: A serum pregnancy test may be substituted for a urine pregnancy test at screening or at the discretion of the investigator.

Clinical laboratory assessments will not be summarized (since they are only performed at screening and applicable unscheduled visits) but will be presented by re-randomization treatment group assignment (Table 1B) in data listings by lab category (i.e. serum chemistry, hematology, other).

#### 9.4. Vital Sign Measurements

Actual values and changes from baseline for vital sign data (i.e. temperature, systolic & diastolic blood pressure, pulse rate) will be summarized by visit, and treatment group for subjects in the Safety Analysis Set. Changes from baseline to each scheduled post-baseline visit will be presented.

A summary of the toxicity grades (according to the FDA toxicity grading scale, Protocol Appendix 3, Table 16–1) will be summarized by visit, and treatment group using the frequency count and percentage of subjects in each category. Shift in toxicity grades from baseline will be summarized by visit and treatment group using the frequency count and percentage of subjects in each category. The shift in toxicity grades from baseline to the worst-case post-vaccination, which will include unscheduled assessments, will also be presented. All vital sign measurements will also be presented in a data listing. Vital sign data will be presented by re-randomization treatment group assignment (Table 1B).

#### 9.5. Physical Examination

Physical examination results will not be summarized but will be presented by re-randomization treatment group assignment (Table 1B) in a data listing.

#### 10. Interim Analyses

A primary database lock will occur following Day 35 data for all primary endpoints including:

- Serum IgG antibody levels specific for the SARS-CoV-2 rS protein antigen(s) as detected by ELISA using GMT or SCR for the two-dose regimens by dose at Day 35 regardless of baseline immune status
- Numbers and percentages (with 95% CIs) of subjects with solicited AEs (local, systemic) for 7 days following each primary vaccination (Days 0, 21) by toxicity grade (daily and maximum) and duration. Unsolicited AEs (eg, treatment-emergent, serious, suspected unexpected serious, those of special interest, MAAEs) through 35 days by MedDRA classification, severity score, and relatedness

This analysis will be performed in order to assess treatment group responses in the Part 2 study, inform decisions around recommended dose regimens by age strata, and facilitate the initiation of the Phase 3 efficacy trial.

Followed by the Day 35 database lock, there will be a Day 217 and a Day 385 database lock to support early submission if deemed necessary.

However, earlier database freezes may occur to potentially allow earlier initiation of Phase 3 efficacy trials in consultation with the SMC and regulatory agencies. These database freezes may be based on data accrued in either Australia or the United States or accrued across the 2 countries combined (depending on the timing of availability of results to allow assessments to occur).

Database freezes may occur when approximately 50 subjects for either age group (18-59, 60-84 years) of each of the 2 first dose formulations (5 µg + 50 µg and 25 µg + 50 µg) have: a) reached Day 21 for safety follow-up; and b) baseline (Day 0) and Day 21 serum IgG antibody levels are available for immunogenicity follow-up. Data for database freezes will be based on partially cleaned and verified data. Immunogenicity data at these reviews may contain microneutralization data or other immunogenicity assessments that are available at the time of the database freeze.

Database freezes may also occur to assess the Day 35 data when approximately 50 subjects for either age group (18-59, 60-84 years) in vaccine groups B and D have: a) reached Day 35 for safety follow-up; and b) baseline (Day 0) and Day 35 serum IgG antibody levels are available for immunogenicity follow-up. Data for database freezes will be based on partially cleaned and verified data. Immunogenicity data at these reviews may contain microneutralization data or other immunogenicity assessments that are available at the time of the database freeze.

A subsequent database freeze will occur at Day 105 for analysis of primary and/or secondary endpoints, with a final database lock at Day 546 (EOS).

In addition, planned analyses for COVID-19 endpoints will be included in all scheduled SMC reviews to allow review of potential vaccine efficacy (if sufficient COVID-19 events occur in the jurisdiction(s) where the study is conducted) or safety concerns.

The planned analyses will be performed by an unblinded biostatistics and programming team. Prior to the primary (Day 35) analysis, the sponsor biostatistics and programming team may be provided with the immunogenicity data set with a limited number of variables and dummy subject identifiers if sponsor analyses are planned, such that the sponsor biostatistics and programming team remain blinded at the subject level. The variables to be included in this blinded immunogenicity data transfer are the dummy subject identifiers, treatment group assignments, visit numbers, assay identifiers, and results only with the dataset subset by the unblinded team to include only those subjects meeting Per-Protocol population criteria for the applicable interim analysis. Only group level unblinded summaries will be generated at planned analyses and subject-level treatment assignment will not be released to the sponsor until the primary (Day 35) analysis. Investigator/site subject level blinding will be maintained until the study is completed. PPD Biostatistics will be unblinded after client signature of approval for SAP v2.1, Addendum v1.0.

#### 11. Changes from Analyses Planned in the Protocol

Contrary to Section 14.2 of the protocol,

- in cases of treatment misallocation subjects will not necessarily be analyzed according to the vaccine actually received (See Section 4.3 for details).
- the full vaccination schedule for PP-Immunogenicity Analysis Set will require subjects to receive study drug at Day 0 and Day 21 only.
- the PP-Efficacy Analysis Set will not exclude subjects from the analysis set due to their SARS-CoV-2 status except if positive SARS-CoV-2 status is observed at Visit 1 (Day 0).
- the Per-Protocol Analysis Set excludes subjects who have a positive Anti-Nucleocapsid Protein result on or after Day 371.

Since some subjects are expected to receive authorized COVID-19 vaccines, the below analysis sets were updated to minimize potential bias/confounding incurred by unblinding. Subjects will be censored (i.e. excluded one day after the date of unblinding or on the date of COVID-19 vaccination, whichever is earlier) for the below analysis sets:

- Per-Protocol Analysis Set
- PP-Immunogenicity PBMC Subset
- PP-Immunogenicity Analysis Set
- PP-Efficacy Analysis Set

A Sensitivity Analysis will be conducted with subjects who are unblinded to receive an approved/authorized COVID-19 vaccine and otherwise meet all Per-Protocol Analysis Set criteria.

Contrary to protocol Section 9.2, Neutralizing antibody activity will also be collected at Day 189.

Contrary to Section 14.5 of the protocol,

- the timing of sponsor unblinding will occur at the time of the primary (Day 35) analysis rather than at the end of the study.
- PPD Biostatistics will be unblinded after client signature of approval for SAP v2.1, Addendum v1.0.
- there will be a Day 217 and a Day 385 database lock to support early submission if deemed necessary. There will be no Day 371 database freeze.

Not only will the Australian sites no longer be required to monitor symptoms associated with SARS-CoV-2 infection via the ePRO system, but their data captured prior to protocol amendment v7.0 will be excluded from analysis in Section 8.3.

[i] IgG antibody to SARS-CoV-2 protein, Neutralization testing, and ACE2 receptor binding inhibition assays (Wuhan and B.1.351 strain or other variants) will be performed at each of the time points indicated.

[j] If applicable, based on reason for unscheduled visit. Subjects may also be evaluated by other methods (eg, telemedicine, hospital/COVID-19 intensive care ward records, home visits).

[k] To include PIMMC (Appendix 4, Table 16 5), AESIs relevant to COVID-19 disease (Appendix 4, Table 16 6), or any newly identified potential AESI followed through EOS.

[l] EOS form will be completed for all participants, including those who terminate early.

[m] Telephone call. Should subjects decide to terminate early, an EOS visit will occur to collect the maximum safety data possible.

[n] Concomitant medications associated with any MAAE attributed to vaccine, potential AESI, or SAE will be recorded. Receipt of any authorized or approved COVID-19 vaccine will also be recorded as a concomitant medication.

DocuSign

#### Certificate Of Completion

Envelope Id: 17076162D91B4C2A858DFE87428F04D3 Status: Completed
Subject: Novavax Phase 2 SAP v2.1 Addendum 4.0
Source Envelope:

| | | |
|---|---|---|
| Document Pages: 66 | Signatures: 3 | Envelope Originator: |
| Certificate Pages: 5 | Initials: 0 | Wei-Ti Huang |
| AutoNav: Enabled | | 929 N Front St |
| EnvelopeId Stamping: Disabled | | Wilmington, NC 28401 |
| Time Zone: (UTC) Monrovia, Reykjavik | | |
| | | IP Address: 134.238.171.24 |

#### Record Tracking

| | | |
|---|---|---|
| Status: Original<br>11 October 2022 \| 13:38 | Holder: Wei-Ti Huang<br> | Location: DocuSign |

#### Signer Events

| Signer Events | Signature | Timestamp |
|---|---|---|
| Jay Pottala<br><br>PPD 21 CFR Part 11<br>Security Level: Email, Account Authentication (Required) | Jay Pottala<br><br>Signature Adoption: Pre-selected Style<br>Signature ID:<br>CC5EF4DE-359C-46C0-A9F2-B4123D2D40FE<br>Using IP Address: 165.1.178.246<br><br>With Signing Authentication via DocuSign password<br>With Signing Reasons (on each tab):<br>I approve this document | Sent: 11 October 2022 \| 13:43<br>Viewed: 11 October 2022 \| 13:51<br>Signed: 11 October 2022 \| 13:53 |

**Electronic Record and Signature Disclosure:**
Not Offered via DocuSign

| Signer Events | Signature | Timestamp |
|---|---|---|
| Shirley Galbiati<br><br>NOVAVAX<br>Security Level: Email, Account Authentication (Required) | Shirley Galbiati<br><br>Signature Adoption: Pre-selected Style<br>Signature ID:<br>B290CF83-7761-47A6-B0D6-A2867BC2FBAA<br>Using IP Address: 68.237.65.104<br><br>With Signing Authentication via DocuSign password<br>With Signing Reasons (on each tab):<br>I approve this document | Sent: 11 October 2022 \| 13:43<br>Viewed: 11 October 2022 \| 13:45<br>Signed: 11 October 2022 \| 13:46 |

**Electronic Record and Signature Disclosure:**
Accepted: 08 August 2022 | 17:19
ID: 74f86e87-1448-4863-8fb5-7cdb77438b97

| Signer Events | Signature | Timestamp |
|---|---|---|
| Wei-Ti Huang<br><br>PPD 21 CFR Part 11<br>Security Level: Email, Account Authentication (Required) | Wei-Ti Huang<br><br>Signature Adoption: Pre-selected Style<br>Signature ID:<br>DC6BED28-310F-47C7-A207-CAEA10F2DD91<br>Using IP Address: 134.238.171.24<br><br>With Signing Authentication via DocuSign password<br>With Signing Reasons (on each tab):<br>I approve this document | Sent: 11 October 2022 \| 13:43<br>Viewed: 11 October 2022 \| 13:44<br>Signed: 11 October 2022 \| 13:45 |

**Electronic Record and Signature Disclosure:**
Not Offered via DocuSign

| In Person Signer Events | Signature | Timestamp |
|---|---|---|

| Editor Delivery Events | Status | Timestamp |
|---|---|---|

| Agent Delivery Events | Status | Timestamp |
|---|---|---|

| Intermediary Delivery Events | Status | Timestamp |
|---|---|---|

| Certified Delivery Events | Status | Timestamp |
|---|---|---|

| Carbon Copy Events | Status | Timestamp |
|---|---|---|

| Witness Events | Signature | Timestamp |
|---|---|---|

| Notary Events | Signature | Timestamp |
|---|---|---|

| Envelope Summary Events | Status | Timestamps |
|---|---|---|
| Envelope Sent | Hashed/Encrypted | 11 October 2022 \| 13:43 |
| Certified Delivered | Security Checked | 11 October 2022 \| 13:44 |
| Signing Complete | Security Checked | 11 October 2022 \| 13:45 |
| Completed | Security Checked | 11 October 2022 \| 13:53 |

| Payment Events | Status | Timestamps |
|---|---|---|

#### Electronic Record and Signature Disclosure

#### ELECTRONIC RECORD AND SIGNATURE DISCLOSURE

From time to time, PPD 21 CFR Part 11 (we, us or Company) may be required by law to provide to you certain written notices or disclosures. Described below are the terms and conditions for providing to you such notices and disclosures electronically through the DocuSign system. Please read the information below carefully and thoroughly, and if you can access this information electronically to your satisfaction and agree to this Electronic Record and Signature Disclosure (ERSD), please confirm your agreement by selecting the check-box next to 'I agree to use electronic records and signatures' before clicking 'CONTINUE' within the DocuSign system.

##### Getting paper copies

At any time, you may request from us a paper copy of any record provided or made available electronically to you by us. You will have the ability to download and print documents we send to you through the DocuSign system during and immediately after the signing session and, if you elect to create a DocuSign account, you may access the documents for a limited period of time (usually 30 days) after such documents are first sent to you. After such time, if you wish for us to send you paper copies of any such documents from our office to you, you will be charged a $0.00 per-page fee. You may request delivery of such paper copies from us by following the procedure described below.

##### Withdrawing your consent

If you decide to receive notices and disclosures from us electronically, you may at any time change your mind and tell us that thereafter you want to receive required notices and disclosures only in paper format. How you must inform us of your decision to receive future notices and disclosure in paper format and withdraw your consent to receive notices and disclosures electronically is described below.

##### Consequences of changing your mind

If you elect to receive required notices and disclosures only in paper format, it will slow the speed at which we can complete certain steps in transactions with you and delivering services to you because we will need first to send the required notices or disclosures to you in paper format, and then wait until we receive back from you your acknowledgment of your receipt of such paper notices or disclosures. Further, you will no longer be able to use the DocuSign system to receive required notices and consents electronically from us or to sign electronically documents from us.

##### All notices and disclosures will be sent to you electronically

Unless you tell us otherwise in accordance with the procedures described herein, we will provide electronically to you through the DocuSign system all required notices, disclosures, authorizations, acknowledgements, and other documents that are required to be provided or made available to you during the course of our relationship with you. To reduce the chance of you inadvertently not receiving any notice or disclosure, we prefer to provide all of the required notices and disclosures to you by the same method and to the same address that you have given us. Thus, you can receive all the disclosures and notices electronically or in paper format through the paper mail delivery system. If you do not agree with this process, please let us know as described below. Please also see the paragraph immediately above that describes the consequences of your electing not to receive delivery of the notices and disclosures electronically from us.

**How to contact PPD 21 CFR Part 11:**

You may contact us to let us know of your changes as to how we may contact you electronically, to request paper copies of certain information from us, and to withdraw your prior consent to receive notices and disclosures electronically as follows:

**To advise PPD 21 CFR Part 11 of your new email address**

To let us know of a change in your email address where we should send notices and disclosures electronically to you, you must send an email message to us at and in the body of such request you must state: your previous email address, your new email address. We do not require any other information from you to change your email address.

If you created a DocuSign account, you may update it with your new email address through your account preferences.

**To request paper copies from PPD 21 CFR Part 11**

To request delivery from us of paper copies of the notices and disclosures previously provided by us to you electronically, you must send us an email to and in the body of such request you must state your email address, full name, mailing address, and telephone number. We will bill you for any fees at that time, if any.

**To withdraw your consent with PPD 21 CFR Part 11**

To inform us that you no longer wish to receive future notices and disclosures in electronic format you may:

i. decline to sign a document from within your signing session, and on the subsequent page, select the check-box indicating you wish to withdraw your consent, or you may;

ii. send us an email to and in the body of such request you must state your email, full name, mailing address, and telephone number. We do not need any other information from you to withdraw consent.. The consequences of your withdrawing consent for online documents will be that transactions may take a longer time to process..

**Required hardware and software**

The minimum system requirements for using the DocuSign system may change over time. The current system requirements are found here: https://support.docusign.com/guides/signer-guide-signing-system-requirements.

**Acknowledging your access and consent to receive and sign documents electronically**

To confirm to us that you can access this information electronically, which will be similar to other electronic notices and disclosures that we will provide to you, please confirm that you have read this ERSD, and (i) that you are able to print on paper or electronically save this ERSD for your future reference and access; or (ii) that you are able to email this ERSD to an email address where you will be able to print on paper or save it for your future reference and access. Further, if you consent to receiving notices and disclosures exclusively in electronic format as described herein, then select the check-box next to 'I agree to use electronic records and signatures' before clicking 'CONTINUE' within the DocuSign system.

By selecting the check-box next to 'I agree to use electronic records and signatures', you confirm that:

- You can access and read this Electronic Record and Signature Disclosure; and
- You can print on paper this Electronic Record and Signature Disclosure, or save or send this Electronic Record and Disclosure to a location where you can print it, for future reference and access; and
- Until or unless you notify PPD 21 CFR Part 11 as described above, you consent to receive exclusively through electronic means all notices, disclosures, authorizations, acknowledgements, and other documents that are required to be provided or made available to you by PPD 21 CFR Part 11 during the course of your relationship with PPD 21 CFR Part 11.
